# Contingency-based flexibility mechanisms through a reinforcement learning model in adults with Attention-Deficit/Hyperactivity Disorder and Obsessive-Compulsive Disorder

**DOI:** 10.1101/2024.01.17.24301365

**Authors:** R. Rodríguez-Herrera, J.J. León, P. Fernández-Martína, A. Sánchez Kuhn, M. Soto-Ontoso, L. Amaya-Pascasio, P. Martínez-Sánchez, P. Flores

**Affiliations:** Department of Psychology, Faculty of Psychology, University of Almeria, Carretera de Sacramento S/N, 04120, La Cañada de San Urbano, Almeria, Spain; Health Research Centre (CEINSA), University of Almeria, Carretera de Sacramento S/N, 04120, La Cañada de San Urbano, Almeria, Spain; Department of Neurology and Stroke Centre. Torrecárdenas University Hospital, Spain; Mental Health Departament. Torrecárdenas University Hospital, Spain

**Keywords:** Attention-Deficit/Hyperactivity Disorder, Obsessive-Compulsive Disorder, adults, probabilistic reversal learning, resting-state functional connectivity, frontoparietal networks, reinforcement learning model

## Abstract

The probabilistic reversal learning paradigm is one of the most used to assess cognitive flexibility during a contingency-based learning process. Lack of cognitive flexibility is related to the symptomatology that characterizes disorders such as Attention-Deficit/Hyperactivity Disorder (ADHD) and Obsessive-Compulsive Disorder (OCD). Resting-state functional connectivity (rsFC) could be a specific predictor of performance in contingency-based cognitive flexibility. However, the mechanisms underlying learning and flexibility in an environment of uncertainty with OCD or ADHD adults have not been widely explored. Computational modelling may be helpful to separate components of the behavioural processes and identify the source of the disorders. In the present study, we aimed to identify the mechanisms underlying contingency-based cognitive flexibility in a sample of the impulsive-compulsive spectrum and healthy controls, and explore the rsFC between the frontoparietal networks (FPN) regions as a possible neuromarker. 148 Spanish-speaking participants (43 patients with OCD, 53 with ADHD, and 52 healthy controls) completed a probabilistic reversal learning task (PRLT). Previously, we obtained a record of FNP rsFC using functional near-infrared spectroscopy (fNIRS). For this purpose, we applied the reinforcement learning model in combination with Bayesian GLM. We found that groups showed an optimal performance in the acquisition phase of the PRLT and higher performance of HC compared to the diagnostic groups in the reversal block, although performance is still optimal in all groups. Likewise, we found that the parameters studied (reinforcement learning rate, punishment learning rate and inverse temperature) predict task performance differently by phase and group. Regarding FNP rsFC, we found that rsFC between left posterior parietal cortex (lpPC) and right posterior parietal cortex (rpPC) seems to credibly predict performance in the acquisition block in the healthy controls. These findings suggest that reducing the uncertainty between action-outcome may help to improve the adaptation of ADHD and OCD patients to changing environments. Thus, understanding sensitivity to punishment or reinforcement and its influence on the decision-making may be important for designing case-specific interventions. According to our data, rsFC between lpPC and rpPC could be important for optimal learning of our task.

## Introduction

Attention-Deficit/Hyperactivity Disorder (ADHD) and Obsessive-Compulsive Disorder (OCD), archetypal phenotypes of impulsive-compulsive spectrum disorders, are characterised by deficits in contingency-based cognitive flexibility (Fineberg et al., 2014). Cognitive flexibility during a contingency-based learning process is the key to adapting behaviour by balancing our experiences with constant updates in the environment to maximise rewards and avoid punishments (Fineberg et al., 2014; Kanen et al., 2019). Lack of flexibility in learning leads to inappropriate persistence, rigid strategies, or maladaptive behaviour (Robbins et al., 2019). It is associated with the clinical symptomatology of OCD such as ruminations, self-doubt, compulsions, or formation of nonadaptive habits (Chamberlain et al. 2007, 2008; Tezcan et al., 2017). A possible underlying mechanism is sensitivity to punishment, which could affect decision-making in the OCD population (Morein-Zamir et al., 2013). Indeed, patients with a diagnosis of OCD show resistance to change in both beliefs and repetitive behaviours despite long-term negative consequences (Allen et al., 2003; Morein-Zamir et al., 2013). Deficits in contingency-based cognitive flexibility are also related to impulsive, quick and unrewarded decisions in the long-term that characterises ADHD (Ziegler et al., 2016). It is well-known the reinforcement sensitivity hypothesis in ADHD to explain deficits in decision-making (Barkley, 1997; Bari and Robbins, 2013; Sonuga-Barke, 2003). Hence, OCD and ADHD could be the extremes of the continuum that is characterized by shared pathophysiological processes, being different from the neuropsychological mechanisms involved (compulsions vs. impulsivity, persistence vs. “inflated flexibility”, exploitation vs. exploration) (Adiccot et al., 2017; Allen et al., 2003; Colzato et al., 2022; Fienberg et al., 2014).

The probabilistic reversal learning paradigm is one of the most widely used to assess contingency-based cognitive flexibility (Bari and Robbins, 2013; Keller and Robbins, 2011). This paradigm explores the ability to change the previously rewarded response when it becomes disadvantageous (Perandrés-Gómez et al., 2021). Patients with a diagnosis of OCD (Remijnse et al., 2006; Tezcan et al., 2017) and ADHD (McCarthy et al., 2016; Chantiluke et al., 2015; Portengen et al., 2021) showed underperformance compared to a matched healthy control group, in terms of lower rewarded correct responses and higher mean errors per reversal. However, the results are inconsistent since other studies were unable to report differences in the performance of the OCD (Chamberlain et al., 2007) and ADHD (Hauser et al., 2014) population compared to healthy controls.

Measures such as the observable error rate do not seem to provide evidence to identify the underlying neuropsychological mechanisms of cognitive processes (Den Ouden et al., 2013; Weiss et al., 2021). Computational modelling, such as reinforcement learning models, may be helpful to separate components of the behavioural processes and identify the source of the disorders (Busemeyer and Stout, 2002; Ahn, 2011). Several studies have developed reinforcement learning models to investigate the mechanisms of decisions making in OCD patients (Apergis-Schoute et al., 2023; Hauser et al., 2017; Kanen et al., 2019; Marzuki et al., 2021) and ADHD patients (Addicott et al., 2021; Hauser et al., 2014; Sethi et al., 2018). Nevertheless, the continuum “persistence vs. inflated flexibility” (Colzato et al., 2022) is not widely explored and may be particularly important to understand contingency-based cognitive flexibility deficits. The metacontrol model (Hommel, 2015, Hommel and Colzato, 2017), theories that the meta-control is the ability to make a balance between persistence and cognitive flexibility in decision-making. An imbalance between these processes could characterise a cognitive processing dimension with two opposite poles: persistence (related to OCD) and inflated flexibility (related to ADHD). Following this model, while the OCD is depicted by repetitive behaviour and flexibility deficits, ADHD is defined by variability in behaviour and impaired goal-directed behaviours (Colzato et al., 2022; Hommel and Colzato, 2017).

The reward processing has been investigated in decision-making paradigms using different resting-state functional connectivity (rsFC) networks (Haynos et al., 2021; Hobkirk et al., 2019). rsFC allows us to explore abnormalities in the temporal synchronisation of brain circuits, regardless of stimuli presentations that may involve shared neural mechanisms underpinning impulsive-compulsive disorders (Haynos et al., 2021). Neuroimaging studies showed evidence of abnormal rsFC in ADHD: hypoconnectivity of the cingulofrontal-parietal (CPF) network and elevated connectivity in the dorsomedial prefrontal cortex (dmPFC) system (Sutcubasi et al., 2020). Concerning OCD, prior studies reported dysconnectivity between frontoparietal networks (FPN) and striatal circuits (Gürsel et al., 2018; Liu et al., 2022). The study of different rsFC networks may contribute to the identification of specific biomarkers in impulsive-compulsive spectrum disorders (Bush et al., 2011). Abnormalities within the FPN seem to be associated with cognitive deficits, especially those related to the ability to coordinate goal-directed adaptive response (Marek and Dosenbach, 2018). Despite FPN integrity seems to be related to rapid, coordinated, flexible, and goal-driven response and its dysfunction affects mental disorders such as ADHD, schizophrenia, or OCD (González-Madruga et al., 2022; Gürsel et al., 2018), its role of FPN rsFC as a specific predictor of performance in contingency-based cognitive flexibility has not been widely investigated.

In the present study, our objective was to identify the mechanisms underlying contingency-based cognitive flexibility in a sample of the impulsive-compulsive spectrum and healthy controls. We used a probabilistic reversal learning task since participants must learn in an uncertain environment, which is more representative of daily decision making (Ahn, 2016). Decision-making tasks engage different parameters such as reward and punishment learning rates, reinforcement sensitivity or stickiness (Ahn, 2016; Marzuki et al., 2021). Thus, we have applied reinforcement learning models to estimate variables that do not depend on the structure of the task. From this framework, computational models can be used to relate these behaviour components to neuroimaging outcomes. Therefore, the second objective was to explore the rsFC between the FPN regions as a possible neuromarker of contingency-based cognitive flexibility.

Based on previous research, we expected that (i) patients with a diagnosis of ADHD or OCD will perform significantly worse on the reversal block of the task than healthy controls, (ii) reward and punishment learning rates will predict the performance of OCD and ADHD groups differently, that is, the performance of OCD patients will be negatively related to the punishment learning rate while (iii) ADHD patients will show an opposite profile, so their performance will be negatively related to reward learning rate, and (iv) FPN rsFC will distinguish and predict contingency-based cognitive flexibility behaviour of different clinical profiles.

## Method

### Participants

In this study, 148 Spanish-speaking participants (43 patients with OCD, 53 with ADHD, and 52 healthy controls) aged 18 to 55 years were evaluated (Table 1). The clinical sample was recruited through the Mental Health and Neurology Units of the Torrecardenas University Hospital. Exclusion criteria were (i) neurological disease, genetic disorder, or traumatic brain injury; (ii) diagnosis of intellectual disability or psychosis; or (iii) motor or sensory disabilities that prevent them from participating in the study. They were contacted by phone to invite them to participate in the project. Then, the participants completed a semi-structured clinical interview to confirm the diagnosis by a psychiatrist with 11 years of experience in the public health system [MSO], a neurologist [LAP], or a predoctoral-level health psychologist [RRH]. Part of the sample was not officially diagnosed prior to the study, although they showed symptomatology in line with the clinical presentation sought. Therefore, 43.4% (*n*=23) of ADHD and 4.6% (n = 2) of cases of OCD were evaluated for possible diagnosis, following the criteria of the International Classification of Diseases 11th Revision (ICD-11) (World Health Organization, 2019). All completed clinical questionnaires to check or rule out whether they met the diagnostic criteria. We administered the ADHD Rating Scale-5 (Richarte et al., 2017), Obsessive-Compulsive Inventory-Revised (Fullana et al., 2005) and Adult Self-Report (Rescorla et al., 2003). Healthy controls (HC) did not have to meet criteria for any psychiatric disorder. Before assessment, participants signed a written informed consent. The local Institutional Ethics Committees (University of Almeria and Torrecardenas University Hospital) approved the project.

**Table 1.**
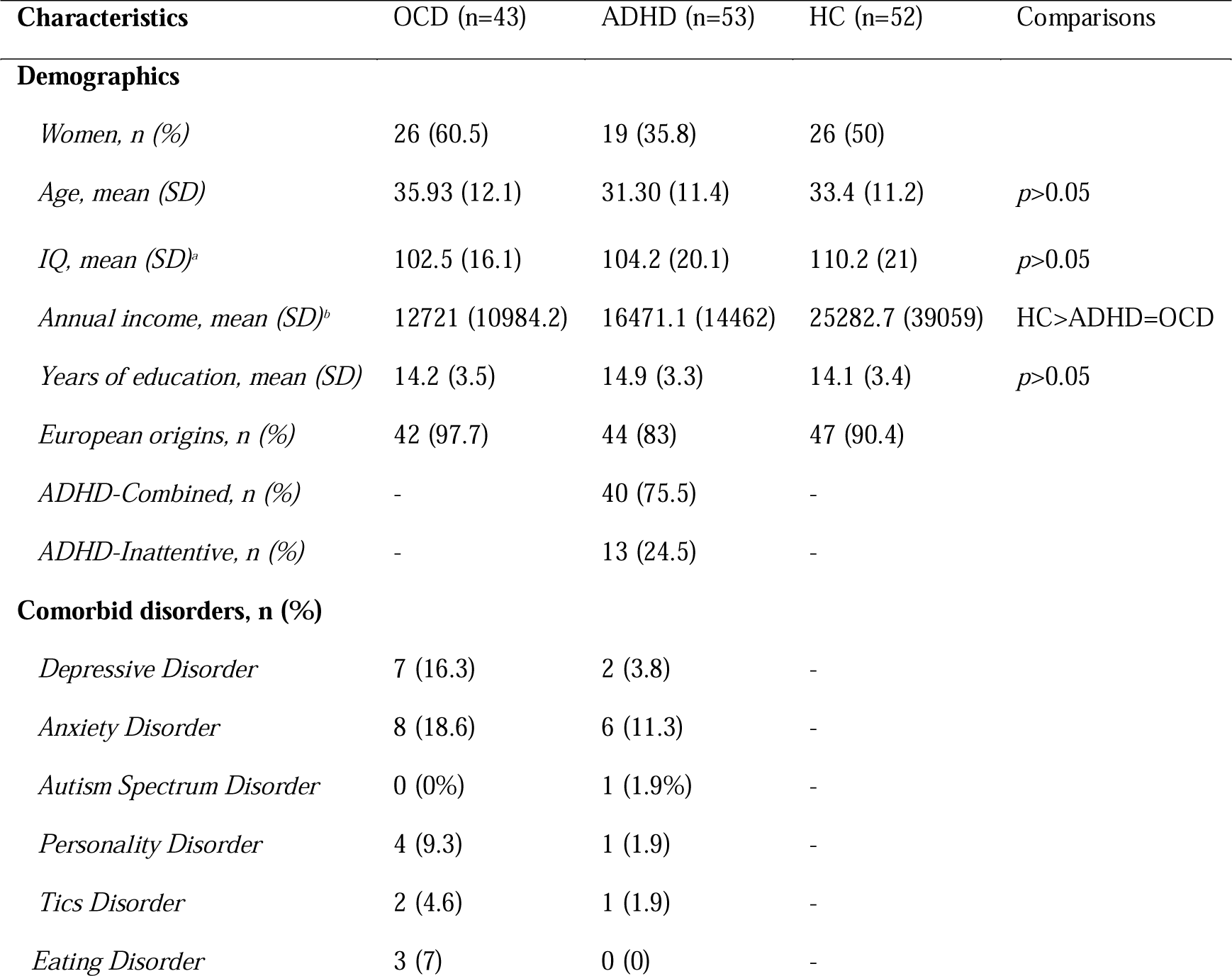
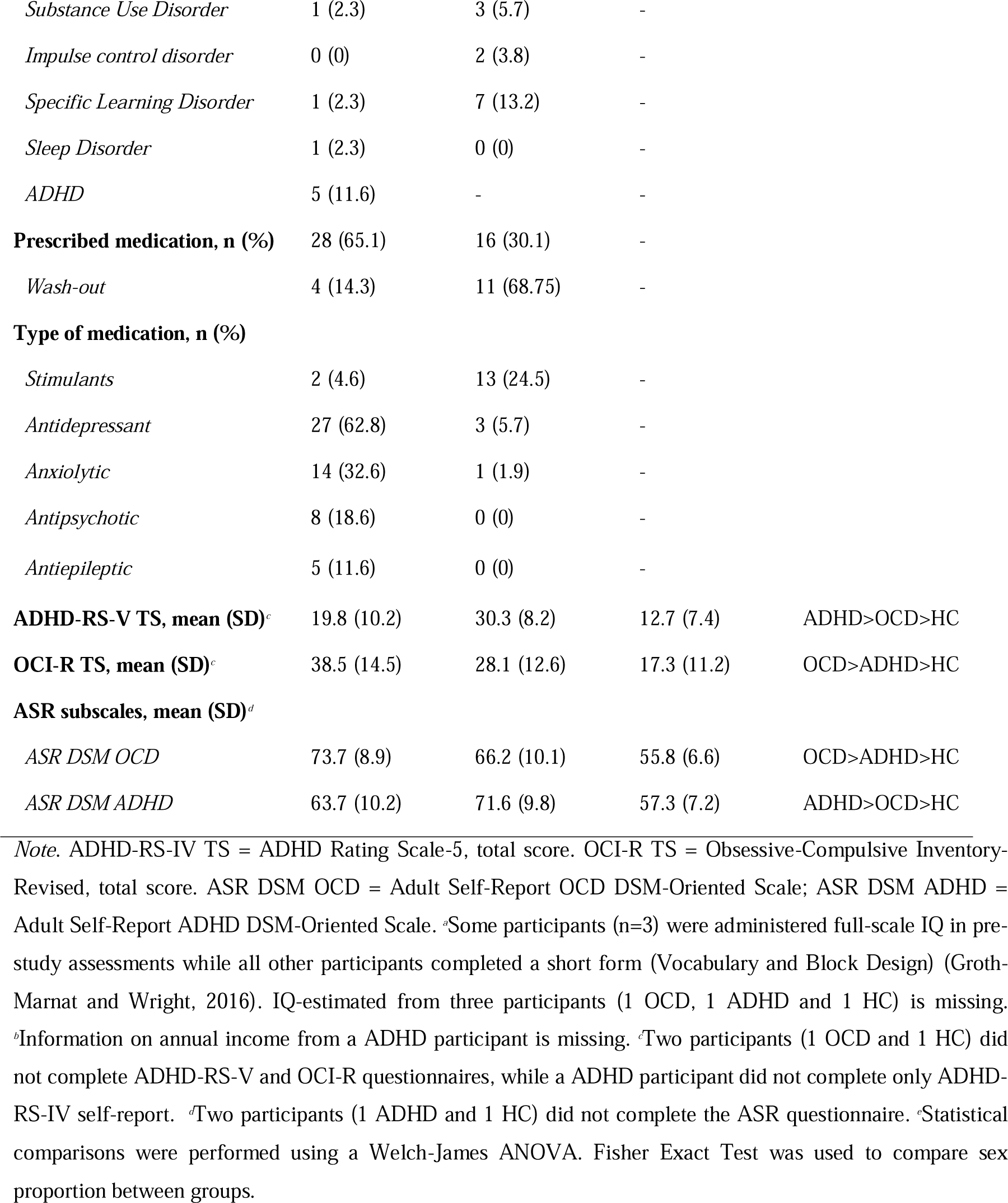
Sample characteristics.

| Characteristics | OCD (n=43) | ADHD (n=53) | HC (n=52) | Comparisons |
| --- | --- | --- | --- | --- |
| **Demographics** |  |  |  |  |
| *Women, n (%)* | 26 (60.5) | 19 (35.8) | 26 (50) |  |
| *Age, mean (SD)* | 35.93 (12.1) | 31.30 (11.4) | 33.4 (11.2) | $p>0.05$ |
| *IQ, mean (SD)<sup>a</sup>* | 102.5 (16.1) | 104.2 (20.1) | 110.2 (21) | $p>0.05$ |
| *Annual income, mean (SD)<sup>b</sup>* | 12721 (10984.2) | 16471.1 (14462) | 25282.7 (39059) | HC>ADHD=OCD |
| *Years of education, mean (SD)* | 14.2 (3.5) | 14.9 (3.3) | 14.1 (3.4) | $p>0.05$ |
| *European origins, n (%)* | 42 (97.7) | 44 (83) | 47 (90.4) |  |
| *ADHD-Combined, n (%)* | - | 40 (75.5) | - |  |
| *ADHD-Inattentive, n (%)* | - | 13 (24.5) | - |  |
| **Comorbid disorders, n (%)** |  |  |  |  |
| *Depressive Disorder* | 7 (16.3) | 2 (3.8) | - |  |
| *Anxiety Disorder* | 8 (18.6) | 6 (11.3) | - |  |
| *Autism Spectrum Disorder* | 0 (0%) | 1 (1.9%) | - |  |
| *Personality Disorder* | 4 (9.3) | 1 (1.9) | - |  |
| *Tics Disorder* | 2 (4.6) | 1 (1.9) | - |  |
| *Eating Disorder* | 3 (7) | 0 (0) | - |  |
| *Substance Use Disorder* | 1 (2.3) | 3 (5.7) | - |  |
| *Impulse control disorder* | 0 (0) | 2 (3.8) | - |  |
| *Specific Learning Disorder* | 1 (2.3) | 7 (13.2) | - |  |
| *Sleep Disorder* | 1 (2.3) | 0 (0) | - |  |
| *ADHD* | 5 (11.6) | - | - |  |
| **Prescribed medication, n (%)** | 28 (65.1) | 16 (30.1) | - |  |
| *Wash-out* | 4 (14.3) | 11 (68.75) | - |  |
| **Type of medication, n (%)** |  |  |  |  |
| *Stimulants* | 2 (4.6) | 13 (24.5) | - |  |
| *Antidepressant* | 27 (62.8) | 3 (5.7) | - |  |
| *Anxiolytic* | 14 (32.6) | 1 (1.9) | - |  |
| *Antipsychotic* | 8 (18.6) | 0 (0) | - |  |
| *Antiepileptic* | 5 (11.6) | 0 (0) | - |  |
| **ADHD-RS-V TS, mean (SD)<sup>c</sup>** | 19.8 (10.2) | 30.3 (8.2) | 12.7 (7.4) | ADHD>OCD>HC |
| **OCI-R TS, mean (SD)<sup>c</sup>** | 38.5 (14.5) | 28.1 (12.6) | 17.3 (11.2) | OCD>ADHD>HC |
| **ASR subscales, mean (SD)<sup>d</sup>** |  |  |  |  |
| *ASR DSM OCD* | 73.7 (8.9) | 66.2 (10.1) | 55.8 (6.6) | OCD>ADHD>HC |
| *ASR DSM ADHD* | 63.7 (10.2) | 71.6 (9.8) | 57.3 (7.2) | ADHD>OCD>HC |

*Note.* ADHD-RS-IV TS = ADHD Rating Scale-5, total score. OCI-R TS = Obsessive-Compulsive Inventory-Revised, total score. ASR DSM OCD = Adult Self-Report OCD DSM-Oriented Scale; ASR DSM ADHD = Adult Self-Report ADHD DSM-Oriented Scale. <sup>a</sup>Some participants (n=3) were administered full-scale IQ in pre-study assessments while all other participants completed a short form (Vocabulary and Block Design) (Groth-Marnat and Wright, 2016). IQ-estimated from three participants (1 OCD, 1 ADHD and 1 HC) is missing. <sup>b</sup>Information on annual income from a ADHD participant is missing. <sup>c</sup>Two participants (1 OCD and 1 HC) did not complete ADHD-RS-V and OCI-R questionnaires, while a ADHD participant did not complete only ADHD-RS-IV self-report. <sup>d</sup>Two participants (1 ADHD and 1 HC) did not complete the ASR questionnaire. <sup>e</sup>Statistical comparisons were performed using a Welch-James ANOVA. Fisher Exact Test was used to compare sex proportion between groups.

## Materials

### Probability Reversal Learning Task *(PRLT)*

We used a computerised version of the PRLT (Jara-Rizzo et al., 2020; Verdejo-García et al., 2010). The task is divided into 2 blocks of 40 trials each. After reading the instructions and before starting the task, the participants completed 10 practice trials. In each trial, the participant must choose between two squares of different colours by clicking with the mouse on it. These squares are the same size and could be blue or orange in the practice trials, and yellow or violet in the experimental trials. The squares always appeared to the left or right of the centre of the screen; the position of the colour was randomised from trial to trial. In each block, one of the stimuli is programmed to be correct and is probabilistically rewarded with 5 points. When they choose the wrong square, 5 points are probabilistically subtracted from the total score. In block 2 the correct stimulus is reversed. Contingencies are the same in both blocks: the correct option is rewarded in 80% of the trials and punished in 20%.

**Figure 1.**
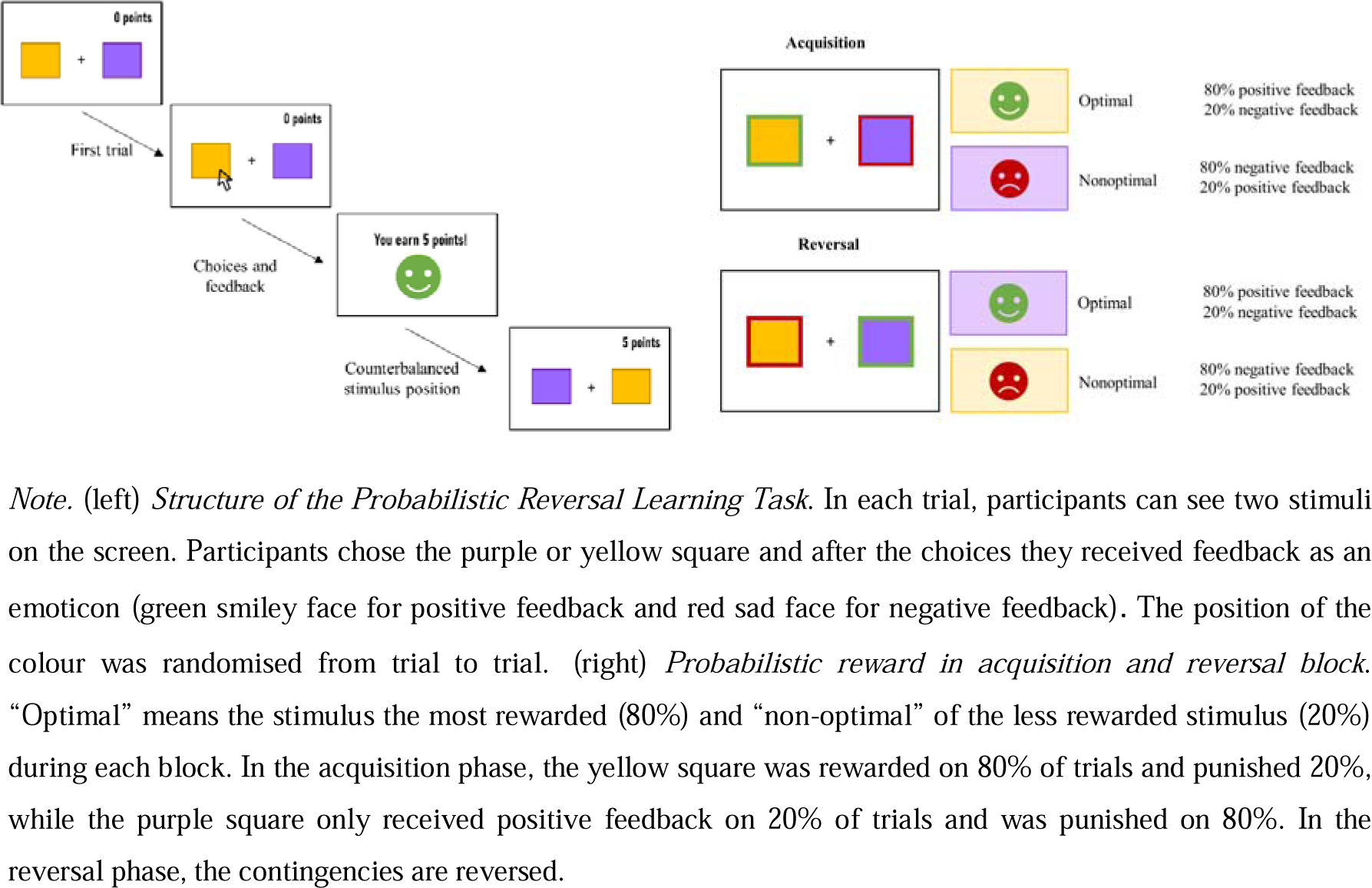
Probabilistic Reversal Learning Task.

### rsFC recording and analyses

We acquired FNP rsFC recording using continuous-wave functional near-infrared spectroscopy (fNIRS) systems (NIRSport device, NIRx Medical Technologies LLC, Berlin, Germany) at wavelengths of 760 and 850 nm. We used two portable fNIRS systems in tandem modem to measure changes in the concentration of oxyhemoglobin (HbO2) and deoxygenated hemoglobin (HbR) with a sampling rate of 3.41 Hz. fNIRS data were collected by NIRStar Software version 15.0 (NIRx Medical Technologies LLC, Berlin, Germany). The cap contained 16 source and 16 detector optodes (54 channels) with an inter-optodes distance of approximately 30 mm compatible with the International 10-10 system. In this study, hemodynamic measurements were recorded from the cortical areas of the FNP. We selected 18 channels related to regions of interest (ROIs). OFC, DLPFC, and posterior parietal cortex (pPC), in both left and right hemispheres (León et al., 2023).

**Figure 2.**
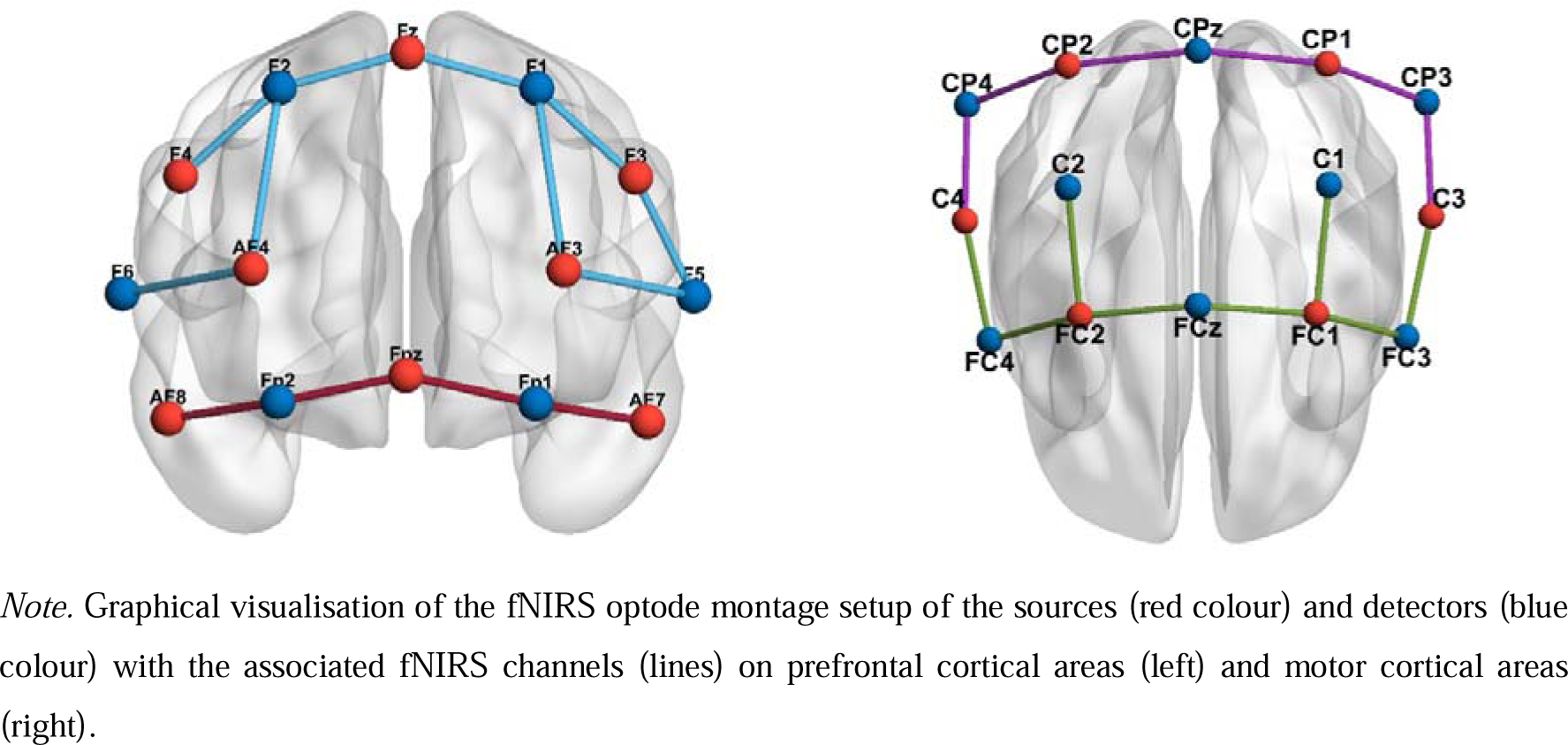
Graphical visualisation of the fNIRS optode montage setup.

All rsFC pre-processing and analyses were performed with MATLAB, using the package NIRS Brain AnalyzIR toolbox (Santosa et al., 2018). We reduced raw NIRS light intensity at 1 Hz and next transformed it into optical density signal. We acquired changes relating to oxyhemoglobin (HbO2) and deoxyhemoglobin (HbR) concentrations by applying the modified Beer-Lambert law (Delpy et al., 1988). HbO2 is the most commonly used measure to correlate with the blood oxygen level-dependent (Duan et al., 2012), then we opted for HbO2 signals to perform the analysis. We filtered optical density data using pre-whitening and pre-weighting methods to assure the correction of band signals such as large intervals of motion artifacts or physiological fluctuations. Both methods are used in combination for filtration process as necessary sept to control type-I errors (Santosa et al., 2017).

After fNIRS signals were pre-processed, we employed Pearson correlations between the time series of each ROIs pair to calculate the functional connectivity at the time domain between brain areas. We used a whole-brain temporal correlation method by estimating the strength of the temporal correlation of the hemodynamic activity of each pairwise measurement channel (Xu et al., 2015).

### Procedure

First, before registering the rsFC, we explained to the participants the procedure for using fNIRS. The recording of the rsFC data lasted approximately 9 minutes. The participants were seated in an experimental room adapted for recording. All subjects were instructed to be relaxed and in silence, as quietly as possible, stay awake with their eyes open and look at the white wall in front of them. Data from four participants could not be recorded due to technical issues.

Then, the participants completed the PRLT which takes about 6 minutes. Before starting, participants were required to read the instructions (see Supplementary Material) and perform the practice tests. After that, the evaluator asked them to explain the task and what the correct stimulus was in that case to make sure they understood it.

The self-reports and WAIS-IV subtests (Vocabulary and Block Design) were administered in a previous clinical session.

## Statistical analysis

### Reinforcement learning model

A Hierarchical Bayesian Analysis (HBA; Ahn et al., 2011) approach was followed to estimate the computational reinforcement learning parameters that models the behaviour of participants. HBA is particularly effective for examining individual differences in parameters, as it calculates individual and group-level parameters concurrently. HBA operates within the realm of Bayesian statistics, where prior knowledge is refined into posterior distributions of various parameters based on evidence, following Bayes’ rule (Kruschke, 2014). In our study, this evidence comprises trial-by-trial data on participant choices and outcomes. For the mathematical details of the model and simulated choice probabilities from estimated RL parameters, see Supplementary Material.

### Generalised Bayesian Logistic Model

To analyse behavioural performance, we generated a Bayesian Generalised Logistic Model tailored to accurately gauge optimal or suboptimal task performance. This model enables us to derive distinct parameters, clarifying interpretation of participant performance and aiding inferences drawn from this performance. Optimal performance is identified as effective learning, indicated by a greater than 50% likelihood of making a correct decision in the final trial of each task block. Suboptimal performance, on the other hand, is marked by insufficient learning or a sub-50% chance of making a correct decision at the end of a block. We focus on three primary parameters to measure performance: “final accuracy” (α), indicating the probability of a correct choice in the task’s last trial, considering all preceding trials; “learning” (βTrial), representing a significant shift in the likelihood of making a beneficial choice between the first and last trials, with a larger difference suggesting more learning; and “learning speed” (γ), reflecting the rate at which a participant moves from their initial to optimal state. Detailed mathematical explanations of the model are available in the Supplementary Material.

## Results

To ease the comprehension of the results, only credible differences and information that are essentially relevant to the hypotheses will be commented on. Mean of posterior distribution (and 95% HDIs) of all estimated parameters, as well as the mean of the differences can be consulted in Tables 1 to (4 in the Supplementary Material.

**Table 2.** Reinforcement learning model parameters predicting performance in each group and block.

| Block | Group |  |  |
| --- | --- | --- | --- |
|  | HC | OCD | ADHD |
| Acquisition | •Lower values of reward learning rate<br>•Higher values of punishment learning rate and inverse temperature | •Higher values of inverse temperature | •Higher values of inverse temperature |
| Reversal | •Higher values of punishment learning rate and inverse temperature | •Higher values of punishment learning rate<br>•Lower values of reward learning rate | •Negative relationship between reward learning rate and performance |

*Note.* High learning rate: quicker update of the expected value after positive (reward learning rate) or negative (punishment learning rate) prediction error. Low learning rate: slower update of the expected value after positive (reward learning rate) or negative (punishment learning rate) prediction error. High temperature inverse: deterministic behaviour driven by the expected value. Low temperature inverse: behaviour driven by the expected value.

### Behavioural performance and learning in each group and block

All groups showed an optimal performance in the first block of the task, since all of them showed a probability to make a correct response in the last trial of the block credibly above 0.5 (HC α_acc: mean = 1.441, 95% HDI from 1.231 to 1.655; OCD α_acc: mean = 1.337, 95% HDI from 1.119 to 1.557; ADHD: α_acc: mean = 1.374, 95% HDI from 1.184 to 1.564). Regarding the reversal block, performance is still considered as optimal for all groups (HC α_acc: mean = 1.322, 95% HDI from 1.115 to 1.509; OCD α_acc: mean = .913, 95% HDI from .702 to 1.117; ADHD: α_acc: mean = .909, 95% HDI from .740 to 1.085). However, in the reversal block, we found credible difference between HC and OCD (HC – OCD α_acc: mean of the differences = .407, 95% HDI from .122 to .686) and HC and ADHD (HC – ADHD α_acc: mean of the differences = .406, 95% HDI from .144 to .663) All groups showed a credible difference in the probability to make a correct choice between the first and the last trial of the reversal block (HC β_trial: Mean = 1.813, 95% HDI from 1.516 to 2.120; OCD β_trial: Mean = 1.472, 95% HDI from 1.149 to 1.811; ADHD β_trial: Mean = 1.376, 95% HDI from 1.086 to 1.668). Note that values of this parameter in the first block are not estimated because the probability of making a correct response in the first trial is fixed at 0.5 for theoretical reasons. Credible greater differences were found between HC and ADHD groups (β_trial: mean of the differences = .438, 95% HDI from .031 to .865).

**Figure 3.**
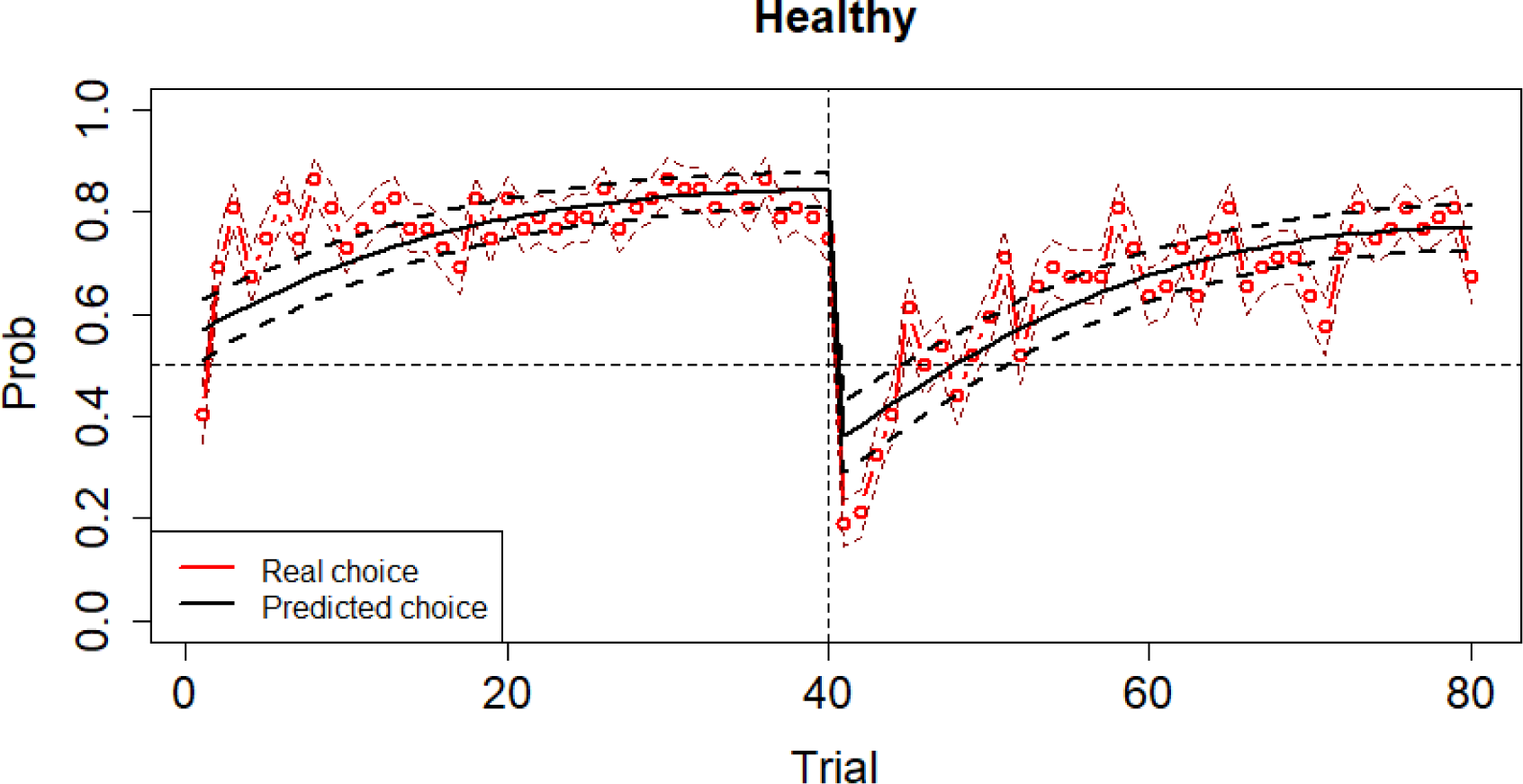
Behavioural performance and learning in HC group.

**Figure 4.**
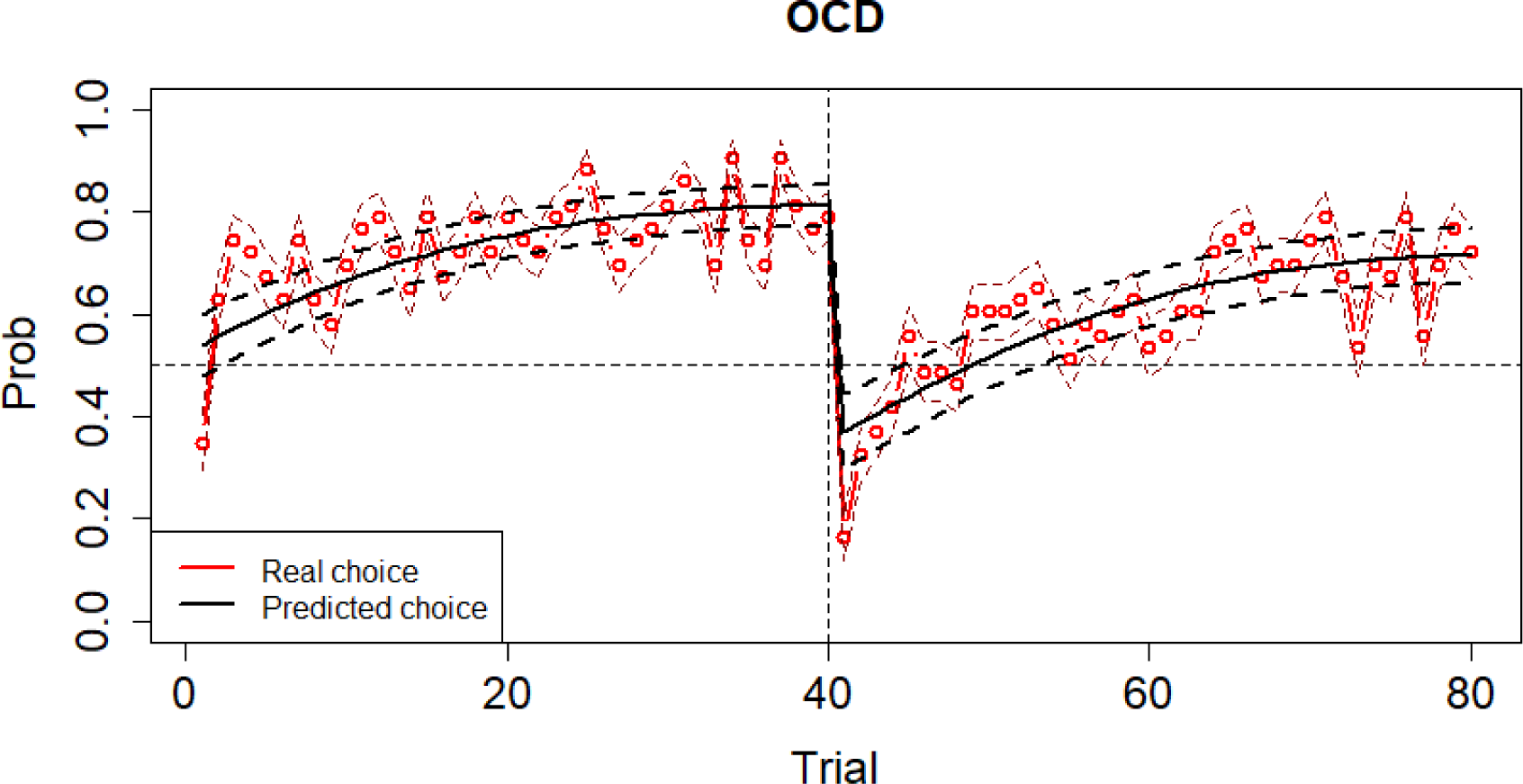
Behavioural performance and learning in OCD group.

**Figure 5.**
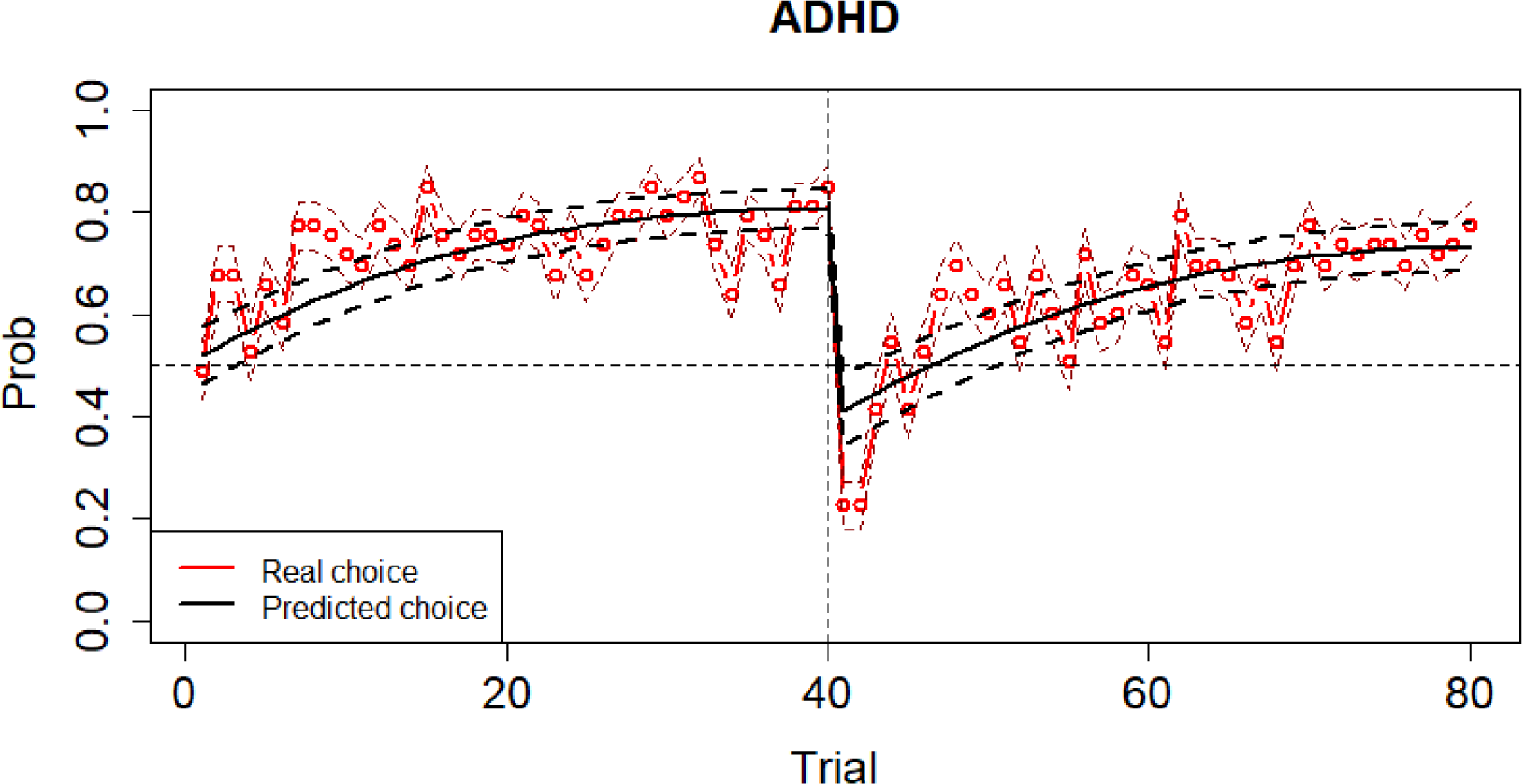
Behavioural performance and learning in ADHD group.

### Reinforcement learning model parameters predicting performance (**α**_acc parameter) in each group and block

#### Acquisition block

An optimal performance of HC group was credibly predicted by lower values of reward learning rate (β_arew: Mean = -.182, 95% HDI from -.292 to -.069) and higher values of punishment learning rate (β_apun: Mean = .211, 95% HDI from .096 to .335) and inverse temperature (β_tau: Mean = .168, 95% HDI from .049 to .285). Hence, good performance in the HC group in this block seems to be related to a slower update of the expected value after recent positive prediction error and a quicker update after negative prediction error together with a more deterministic than stochastic behaviour driven by that expected value.

Performance of the OCD group was credibly related to higher values of inverse temperature (β_tau: Mean = .267, 95% HDI from .134 to .401). Regarding the ADHD group, performance was credibly related to higher values of inverse temperature (β_tau: Mean = .102, 95% HDI from .002 to .210). Then, performance in clinical groups in the acquisition block seems to be only related to a more deterministic than stochastic behaviour driven by the expected value.

When comparing the regression coefficients of the parameters between groups, we found credible different regression coefficient values between HC and OCD (HC - OCD: Mean of the differences = .202, 95% HDI from .038 to .377) in punishment learning rate.

#### Reversal block

In the HC group performance was credibly predicted by higher values of punishment learning rate (β_apun: Mean = .251, 95% HDI from .142 to .359) and inverse temperature (β_tau: Mean = .196, 95% HDI from .093 to .299). Performance in this block of the HC group is then related to quicker update after negative prediction error and a more deterministic behaviour driven by the expected value.

Performance in the OCD group was credibly predicted by higher values of punishment learning rate (β_apun: Mean = .196, 95% HDI from .077 to .321) and lower values of reward learning rate (β_arew: Mean = -.165, 95% HDI from -.281 to -.055). This would mean that a more optimal performance in the OCD group seems to be related to a quicker update of the expected value after negative but a slower update after positive prediction error.

Regarding the ADHD group, we found a credible negative relationship between reward learning rate and performance (β_arew: Mean = -.157, 95% HDI from -.286 to -.027). Thus, in the ADHD group, performance in the reversal block would be related to a slower update of the expected value after positive prediction error.

When comparing the value of the regression coefficient of the punishment learning rate we found credibly different values between HC and ADHD (HC - ADHD: β_apun: mean of the differences = .206, 95% HDI from .058 to .348). A credible difference in the value of the regression coefficient for the inverse temperature parameter was found between HC and OCD (HC - OCD: β_tau: Mean of the differences = .203, 95% HDI from .043 to .366).

### Resting-state functional connectivity predicting performance in each group and block

RSFC between lpPC and rpPC seems to credibly predict performance in the acquisition block in the HC (Mean = .218, 95% HDI from .028 to .396).

## Discussion

This study explores the mechanisms underlying contingency-based cognitive flexibility and decision-making in 158 Spanish-speaking adults (18-55 years) with OCD or ADHD diagnosis and HC. First, we aimed to analyse possible deficits in cognitive flexibility contingency-based learning process of patients with impulsive-compulsive disorder compared to HC. We sought to understand the decision-making strategies of OCD and ADHD in an uncertain environment. We applied the reinforcement learning model in combination with Bayesian GLM to determine how reward learning rate, punishment learning rate and inverse temperature predicted performance on the task based on the impulsive-compulsive patients and healthy controls. Second, we explored whether the rsFC between the FPN regions could predict performance on the task as a possible neuromarker of contingency-based cognitive flexibility.

All groups showed an optimal performance in the acquisition phase of the PRLT, and no credible differences in performance were found between impulsive-compulsive patients and HC. Despite this, we found higher performance of HC compared to the diagnostic groups in the reversal block (greater probability to make a correct choice in the last trial, represented by the α_acc parameter in the Bayesian GLM) although performance is still optimal in all groups. Therefore, our first hypothesis has been partially supported by data. By contrast, prior studies supported the traditional theory of cognitive inflexibility in patients with diagnosis of OCD (Abramovitch et al., 2013; Gruner and Pittenger, 2017; Shin et al., 2014) and ADHD (Willcutt et al. 2005), which is inconsistent with our findings. It has been widely reported that ADHD (McCarthy et al., 2018; Portengen et al., 2021) or OCD (Remijnse et al., 2006; Tezcan et al., 2017) patients show underperformance in PRLT. However, our results showed that all groups perform optimally on the task, in spite of finding credible differences between the clinical groups and the healthy controls. So, our results are in line with others researches which supported that there might not be such difficulties in contingency-based cognitive flexibility, as they found no differences in performance between ADHD (Hauser et al., 2014) or OCD (Chamberlain et al., 2007) to healthy controls when PRLT is applied. This conclusion is also congruent with some studies that obtained when applied classical cognitive flexibility tests, such as the Wisconsin Card Sorting Test (WCST), in OCD (Henry, 2006; Kuelz et al. 2004; Marzurki et al., 2021) and ADHD children (Miranda et al., 2013). A possible explanation for this evidence would be that different executive functions support different kinds of cognitive flexibility: working memory or control inhibition (Blakey et al., 2016). Following this hypothesis, classical paradigms in cognitive and behavioural neuroscience could measure components of cognitive flexibility with diverse neuronal correlates (Uddin, 2021). Moreover, decision-making could be affected by multiple factors, not dependent on executive functions, such as the degree of uncertainty or motivation (Addicot et al., 2017; Sonuga-Barke, 2003).

Theoretically, the learning rate indicates the speed with which the expected value is updated, depending on the contingencies of the environment (Adiccot et al., 2021; Daw et al., 2006). We study how quickly participants update the values associated with choices after positive (reward learning rate) or negative (punishment learning rate) prediction error and whether that could be a predictor of behavioural performance. In our case, we found different parameters predicting the behaviour of each group. We also explore the inverse temperature parameter. This parameter, also known as “coherence” or “consistency” in decision-making, explains how much value is estimated to influence the choice (Nussenbaum and Hartley, 2019). In other words, it is related to the pattern to respond based on the expected value (Marzuki et al., 2021).

We found that HC performance was credibly predicted by higher values of inverse temperature in both acquisition and reversal blocks. Moreover, an optimal performance of HC group was credibly predicted by higher values of punishment learning rate also in both blocks, while the reward learning rate only predicted performance in the acquisition block. The task simulates an uncertain environment, especially with the unexpected changes in the reversal block, so a proper exploration-exploitation balance could be at the basis of optimal performance. Thus, a rapid update of the expected value based on negative prediction error, together with a response driven by that expected value seems to characterise an optimal performance in HC and, therefore, their exploration-exploitation balance in both blocks of the taskThis strategy could be very useful for obtaining information from the environment and adjusting our response to an adaptive choice (the most valuable) in an unstable environment (Daw et al., 2006). Interestingly, the updating process after positive prediction error seems to be not predictive in the reversal block, which could point towards that the adaptation to relatively new environments could not be driven by positive but negative feedback.

As well as the HC group, we found that the performance of the OCD and ADHD groups was credibly related to higher values of inverse temperature in the acquisition block. In this sense, we found no credible differences in the regression coefficients of the learning rates between groups, and no credible predictions in the acquisition block. This result is in line with those obtained for performance in the acquisition phase. We found no credible differences between the groups in the learning process. Learning for all groups is predicted by the tendency to respond based on the expected value. Previous studies have found no difference in the probabilistic feedback-based learning process in OCD and ADHD compared to controls (Endrass et al., 2011; Hauser et al., 2014). They found that OCD and ADHD patients learned to choose the better of the two stimuli based on negative or positive feedback (Endrass et al., 2011; Hauser et al., 2014). This is consequently congruent with the fact that for correct acquisition, it may be necessary to drive our responses by the expected value and not to show deterministic behaviour. The reported evidence may indicate the need for further parameters to clarify the learning process of ADHD and OCD in PRLT paradigm.

Regarding the reversal block, our results indicate that the OCD performance was credibly related to higher value of punishment learning rate, which may suggest a quick update of the expected value of their choices after negative feedback in environments of high uncertainty (Apergis-Schoute et al., 2023). Thus, our second hypothesis, which was that reward and punishment learning rates will predict the performance of OCD and ADHD groups differently and the performance of OCD patients will be negatively related to the punishment learning rate, has been partially supported by data. However, high punishment learning rate predicting performance in the reversal block is congruent with previous findings reporting that elevated sensitivity to negative feedback in OCD (Endrass et al., 2011; Morein-Zamir et al., 2013). Sensitivity to punishment is related to OCD symptomatology, as they show resistance to changing beliefs and associated rituals (compulsions), to avoid negative consequences (Allen et al., 2003; Morein-Zamir et al., 2013). Kanen et al., 2019 found that OCD participants had increased learning from negative feedback in terms of non-rewarded stimulus. Endrass et al., (2011) showed that OCD patients tend to avoid stimuli that have previously been punished more than to select reward stimuli after learning processes. Here, we could explain why the punishment learning rate predicted performance only in the reversal block in the OCD group. In fact, punishment learning rate predicted the performance differently of the OCD group compared to the HC in the acquisition block, with a stronger effect on the performance of the controls. Likewise, reduced learning from punishment is associated with perseverance behaviour, where punishing the previously reinforced stimulus has no effect on the response, so the stimulus is not devalued (Den Ouden et al., 2013). In this sense, following Marzuki et al., 2021, reduced punishment learning rate is compatible with the other clinical presentation of OCD: these patients engage in perseverative behaviours despite the long-term negative consequences (Chamberlain et al., 2021). Morein-Zamir et al. (2013) reported that OCD patients showed impulsive responses under impending punitive situations. Hommel and Cozalto., (2017) supported this proposal in the metacontrol model, which OCD is situated at the “persistence” pole. An explanation for our findings is that OCD only manifests ritual behaviours in familiar environments (Fradkin et al., 2020). Following this line, additional studies reported that, in deterministic tasks (low uncertain environment), OCD patients tended to establish ritualistic behaviours based on a history of learning in which a rule has been consistently rewarded (Apergis-Schoute et al., 2023; Marzuki et al., 2021). Indeed, Cozalto et al. (2022) proposed based-situation intra-individual variability to clarify that OCD patients may not necessarily exhibit this pattern. Therefore, they could show characteristic behaviours of both extremes depending on the environment (Cozalto et al., 2022). Alternatively, this dissociation might suggest that OCD could manifest itself through distinct behavioural endophenotypes (Robbins et al., 2019). As well, we found that the OCD performance of the reversal block is also predicted by a lower value of reward learning rate. Studies on motivational variables in OCD have reported poor motivation and apathy in goal-directed tasks (Moritz et al., 2012, Raffard et al., 2020). This conclusion fits well into the theory that OCD patients are more motivated by loss-avoidance than reinforcement (Kaufmann et al., 2013). So, taking together, results about reinforcement learning parameters in OCD could suggest a behavioural mechanism driving an optimal adaptation to changing environmental rules: a needed balance between how much negative/positive prediction weights the need to explore/exploit. This is in consonance with emerging literature showing that “choice volatility” (related to lower inverse temperature) seems to characterise OCD (Benzina et al., 2021; Hauser et al., 2017; Kanen et al., 2019; Marzuki et al., 2021). This could be in line with no credible relationship between inverse temperature and performance in the reversal block in OCD and its less influence compared to HC. The “exploration” strategy, based on the dilemma exploration-exploitation (EE) dilemma, may be due to greater uncertainty in decision-making, as OCD patients generally report more subjective uncertainty than HC (Addicot et al., 2017; Stern et al., 2013). Previous extensive literature supports this conclusion, finding that OCDs have seen increased exploration behaviours when the results are probabilistic (Banca et al., 2015; Hauser et al., 2017; Marzuki et al., 2021). Therefore, we can conclude that this parameter similarly predicts OCD and HC behaviour in the acquisition, but not in the reversal block, because people diagnosed with OCD may show reduced tolerance to uncertainty (Hauser et al., (2017).

Concerning ADHD, this group showed a credible negative relationship between reward learning rate and performance in the reversal block. Therefore, our third hypothesis has been supported by data. A potential reason for this behaviour might be a lack of motivation to heed signals that suggest forthcoming rewards (Cubillo et al., 2012; Stoy et al., 2011). This profile is also consistent with previous studies reporting reward sensitivity in ADHD, which could be understood as a preference for immediate rewards over delayed rewards with more long-term gains (Barkley, 1997; Bari y Robbins, 2013; Sonuga-Barke, 2003). Reward sensitivity in ADHD has been measured through paradigms such as reversal learning task (Alsop et al., 2016; Hauser et al., 2014; Portengen et al., 2021) and delay discounting task (Barkley et al., 2001; Marx et al., 2021). As the meta-control model proposes, the “inflated flexibility” in ADHD (giving more weight to immediate contingency over the past experiences) leads to difficulties in focusing on a goal (Hommel and Cozalto, 2017). So, the adaptability to changing environments in ADHDs seems to be mainly driven by the power that positive prediction error has to reinforce actions that lead to a reward. In ADHD research, Sethi et al., (2018) supported this and reported that ADHD adults choose an unfamiliar option with uncertain reinforcement over a non-risky option. This response tendency is also related to reward sensitivity, which could explain the decision-making strategy in the ADHD group (Ziegler et al., 2016). Prior studies have reported that adolescents and adults with ADHD had lower learning rates than controls because of a non-balance between both strategies (Hauser et al., 2014; Ziegler et al., 2016).

This mechanism could be shared by both clinical groups since in the OCD group we also found, as we have previously reported, that a lower value of the reward learning rate predicted their performance in this block. Thus, our results may provide evidence of shared neuropsychological mechanisms by ADHD and OCD groups (Colzato et al., 2022; Fienberg et al., 2014). On the other hand, we found no relationship between the ADHD performance and the punishment learning rate. Additionally, punishment learning rate predicted the performance differently of the ADHD group compared to the HC in the reversal block, with a stronger effect on the performance of the controls. As has been above-mentioned, rewards seem to moderate behaviour more than punishment in ADHD children and adults, reporting insensitivity to negative feedback or punishment in risky decision-making tasks (DeVito et al. 2008; Humphreys & Lee, 2011; Masunami et al., 2009; Oguchi et al., 2023). Therefore, reward insensitivity during the reversal block appears to produce poorer performance.

Regarding FNP rsFC as a predictor of performance, our fourth hypothesis is partially supported since we found significant results in relation to bilateral pPC rsFC. Concretely, we found that rsFC between lpPC and rpPC seems to credibly predict performance in the acquisition block in the HC. Also, we found that the rsFC between lpPC and rpPC predicted the performance differently of the ADHD group compared to the HC in the acquisition block, with a stronger effect on the performance of the controls. Previous literature relates the activation of the inferior and posterior parietal cortex (pPC) to decision-making under uncertainty in healthy subjects, especially in “quick” decisions without previously established rules (Krug et al., 2014; Volz et al., 2005). Volz et al., (2005) reported a positive correlation between uncertainty and bilateral activation of the posterior parietal cortex. This brain area seems to play a key role in the “exploration” strategy when information is not sufficient for decision making, as well as connecting with the orbitofrontal and dorsolateral cortex, which form a neural network involved in decision making in uncertain environments (Krug et al., 2014). So, according to our data, rsFC between lpPC and rpPC could be important for optimal learning of our task. Accordingly, other authors showed that the parietal cortex is involved in the relation between responses and its feedback, as well as the flexibility of this brain region to adapt these action-outcome associations according to the demands (Sugrue et al., 2004; Wisniewski et al., 2015). Wisniewski et al. (2015) supported the hypothesis that the parietal cortex may be important in driving contingency-based decisions. Instead, OFC and DLPFC rsFC has not shown any relationship with the performance of the participants. In this line, a recent exploratory study concluded that rsFC FPN was not a neuromarker of decision-making profiles in impulsive-compulsive populations and healthy controls (León et al., 2023). In contrast, prior studies typically report these areas are associated with the processing feedback (Marek and Dosenbach, 2018; O’Doherty et al., 2001). A possible explanation is the high variability found in the rsFC across participants for multiple variables such as personality traits or pharmacological treatment (Marek and Dosenbach, 2018). Although the studies reporting OFC, DLPFC and mPFC anomalies in OCD and ADHD patients during contingency-based cognitive flexibility and decision-making tasks employed functional magnetic resonance imaging (fMRI) (Chamberlain et al., 2008; Hauser et al., 2014; Gu et al., 2008), the rsFC measure has shown that dysfunctions also affect interneuronal networks such as FPN or DMN (Cubillo et al., 2012; Liu et al., 2022), whose functional connectivity is related to cognitive flexibility (Thomas et al., 2023). Variability in neuroimaging results might be caused by the different methodologies and conceptualisations for interpreting the findings. For future research a homogeneous conceptual and methodological framework is needed to obtain conclusive evidence between brain and behaviour (Marek and Dosenbach, 2018).

Our study provides evidence for neurobehavioral mechanisms of cognitive flexibility and decision-making, which may have clinical implications. In daily life, contingencies may change unexpectedly and adapting to these changes is key for successful response in several environments (Daw et al., 2006). Although they showed normative execution in our task, they performed credibly worse than the controls. This might be interpreted as poorer behavioural adaptation on the part of the OCD and ADHD patients when the reinforcement-punishment contingencies changed. It could be reflected in the symptomatology that characterises these disorders, especially when there is uncertainty about the behaviour-outcome association (Abramovitch et al., 2013; Alsop et al., 2016; Tripp & Wickens, 2008). These findings suggest that reducing the uncertainty between action-outcome may help to improve the adaptation of ADHD and OCD patients to changing environments. For example, providing detailed and explicit information on contingencies or structuring the tasks could be helpful to learn an adaptive response (Alsop et al., 2016; Moritz et al., 2004). Difficulties adapting their response to change could be explained by different underlying mechanisms in ADHD and OCD, both in learning and switching processes. Thus, understanding sensitivity to punishment or reinforcement and its influence on the decision-making may be important for designing case-specific interventions.

Finally, some limitations of our findings should be mentioned. First, most of the OCD patients (55,8%) and some ADHD patients (9,4%) were medicated without a wash out-time, so we could not control whether it influenced performance on PRLT and the rsFC recording. Second, relevant variables such as motivation (not real monetary reward) (Sonuga-Barke, 2003, Xu et al., 2018) or the degree of uncertainty of the environment (Addicot et al., 2017; Cozalto et al., 2022) were not controlled and may have affected the results. Third, subcortical areas of brain impaired contingency-based cognitive flexibility such as dorsal and ventral striatum (Yaple et al., 2019) seem affected in OCD (Piras et al., 2015) and ADHD (van Hulst et al., 2017). Regardless, we could not measure these areas due to the limitations of the fNIRS technique. Another limitation of the present study was the relatively small number of participants to obtain reliable brain conclusions (Turner et al., 2018). Fourth, emerging clustering analysis studies reported various profiles that cut across categorical nosologies in ADHD (Bergwerff et al., 2019; De Nadai et al., 2023; Fernández-Martín et al., 2023). We cannot rule out the existence of OCD and ADHD patterns that might explain these results and, perhaps, the heterogeneous decision-making strategies found in the previous literature (Apergis-Schoute et al., 2023; Hauser et al., 2014; Mazurki et al., 2021; Sethi et al., 2018). In this sense, further research could investigate the possible contingency-based cognitive flexibility profiles within the disorders, applying a design that includes a behavioural and neural-functional analysis of parameters involved in reward-punishment processing. It might also help to study whether prescription drugs for ADHD and OCD enhance processes such as sensitivity to reward or punishment.

## Data Availability

All data produced in the present study are available upon reasonable request to the authors

