## Supplementary material for "Contingency-based flexibility mechanisms through a reinforcement learning model in adults with Attention-Deficit/Hyperactivity Disorder and Obsessive-Compulsive Disorder": suplementary material: GLM_betas_group_graphs.pdf

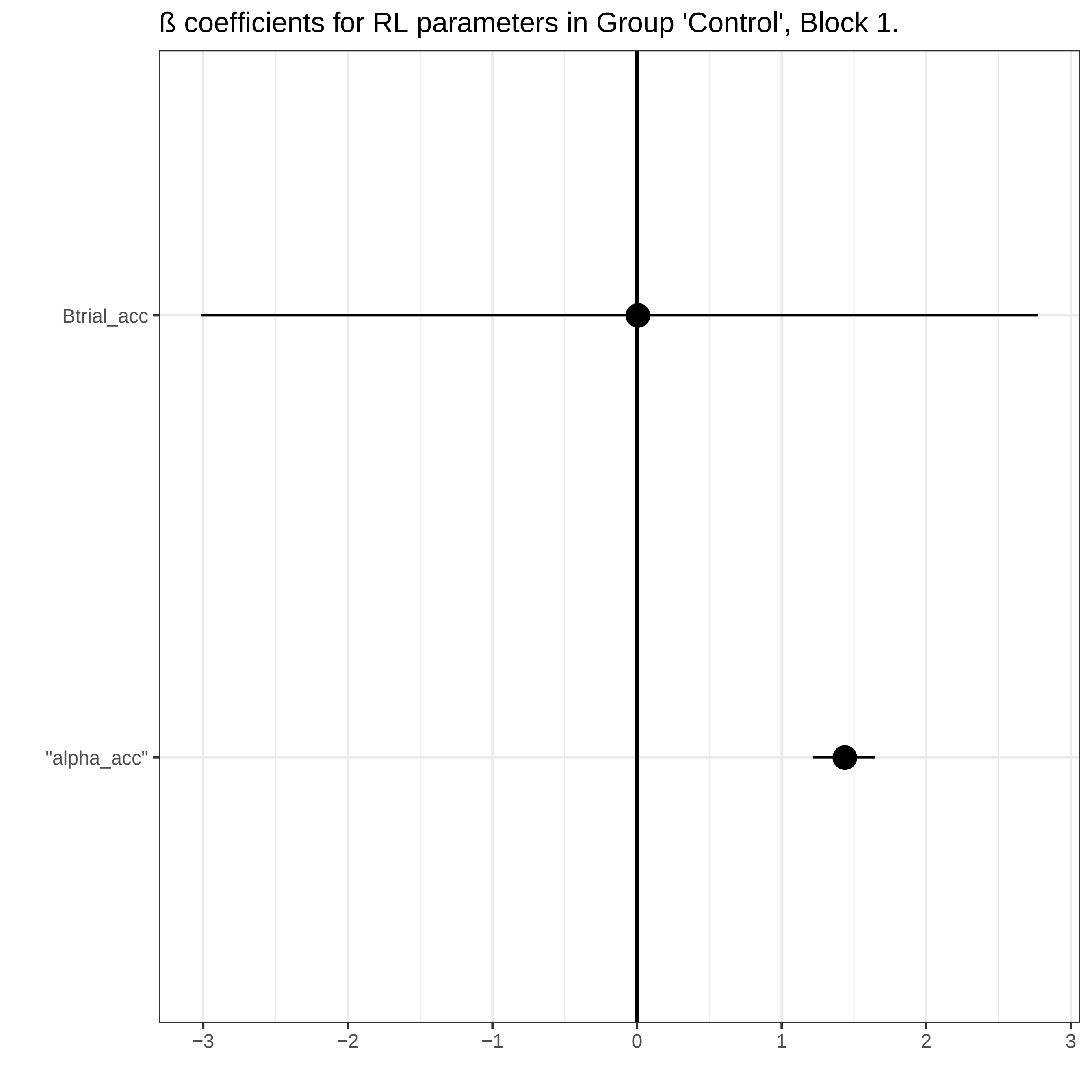
ß coefficients for RL parameters in Group 'Control', Block 1.
Btrial_acc
"alpha_acc"
−3
−2
−1
0
1
2
3

ß coefficients for RL parameters in Group 'Control', Block 2.

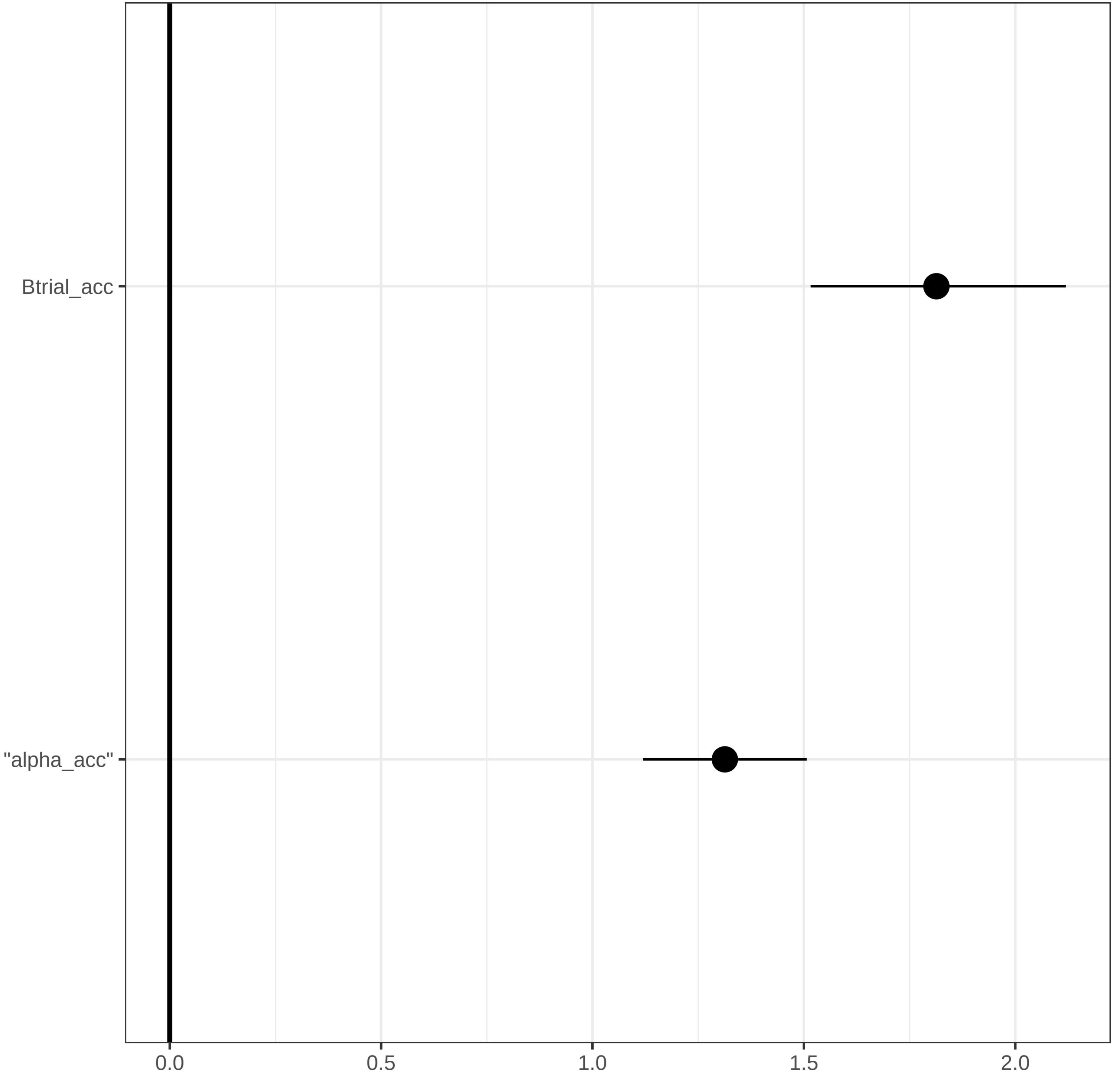

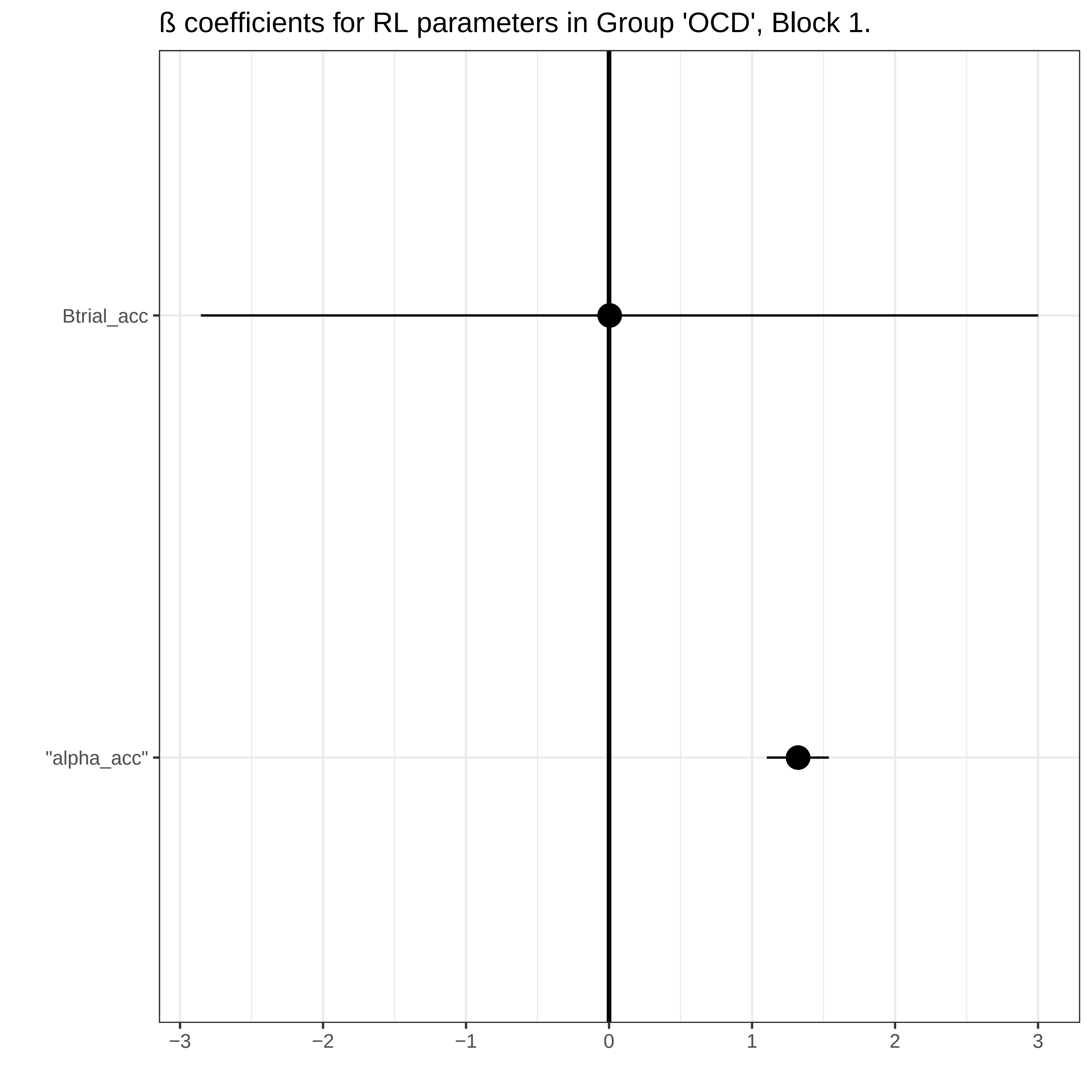
ß coefficients for RL parameters in Group 'OCD', Block 1.
Btrial_acc
"alpha_acc"
−3
−2
−1
0
1
2
3

ß coefficients for RL parameters in Group 'OCD', Block 2.

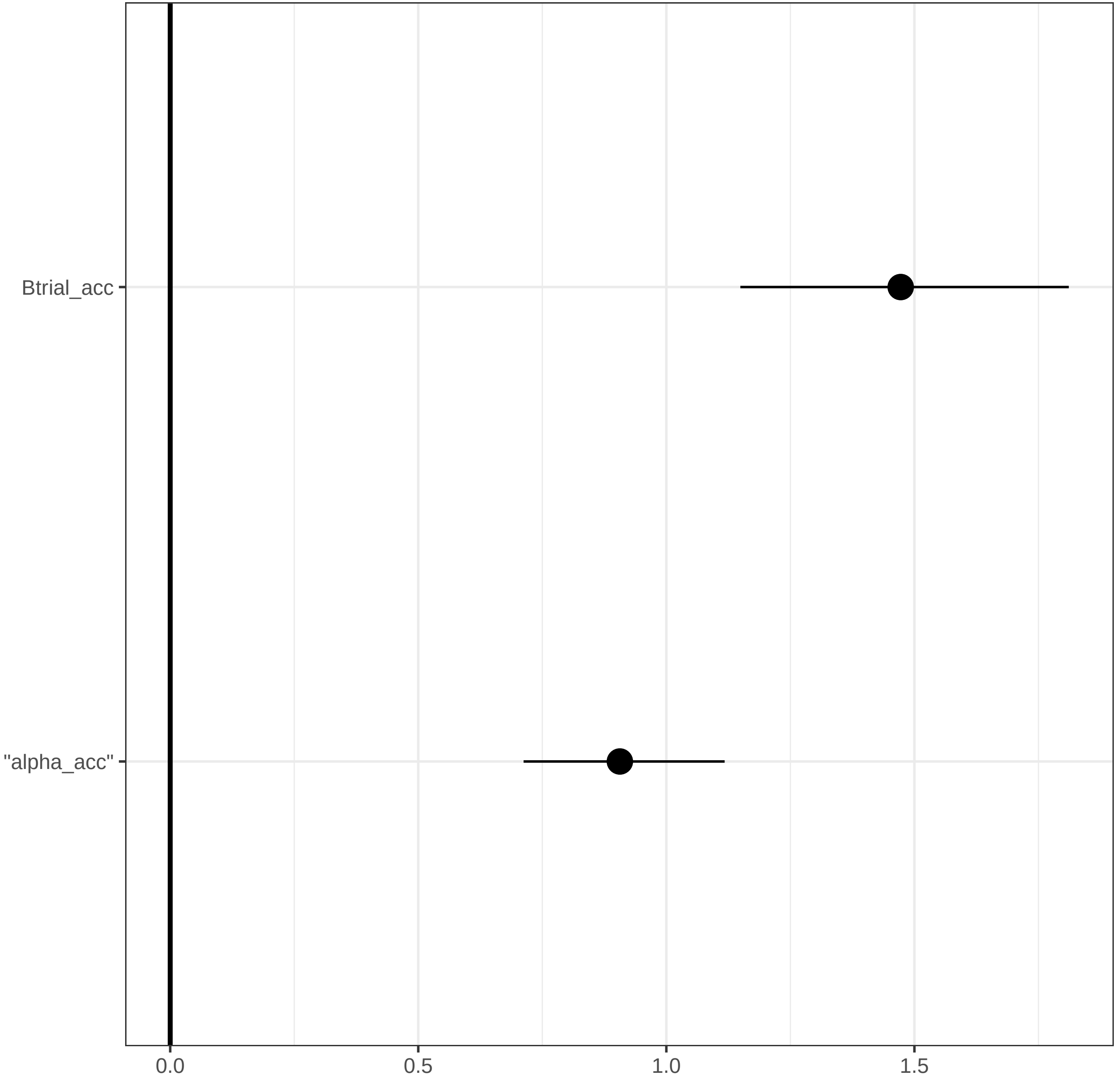

ß coefficients for RL parameters in Group 'ADHD', Block 1.

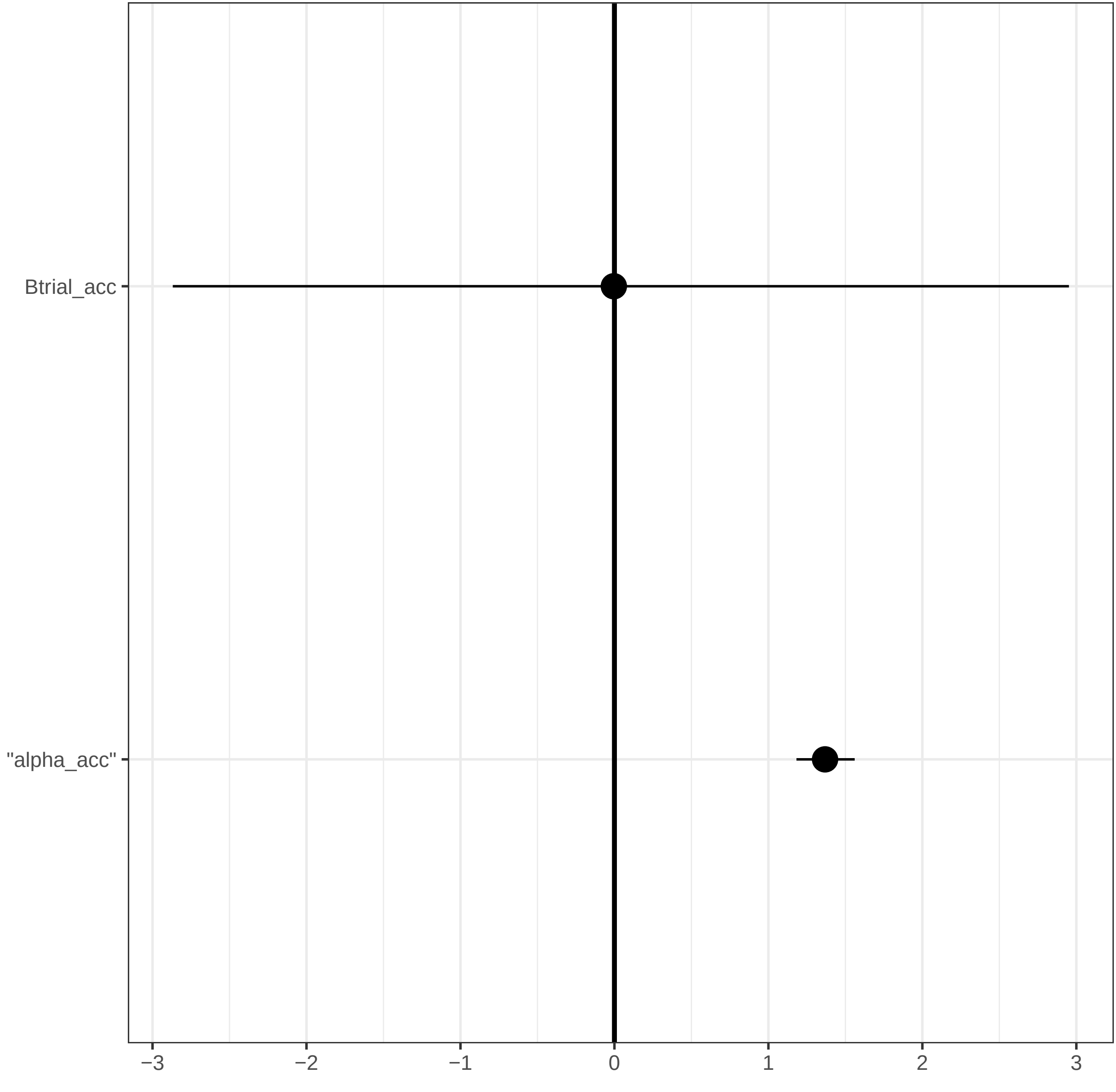

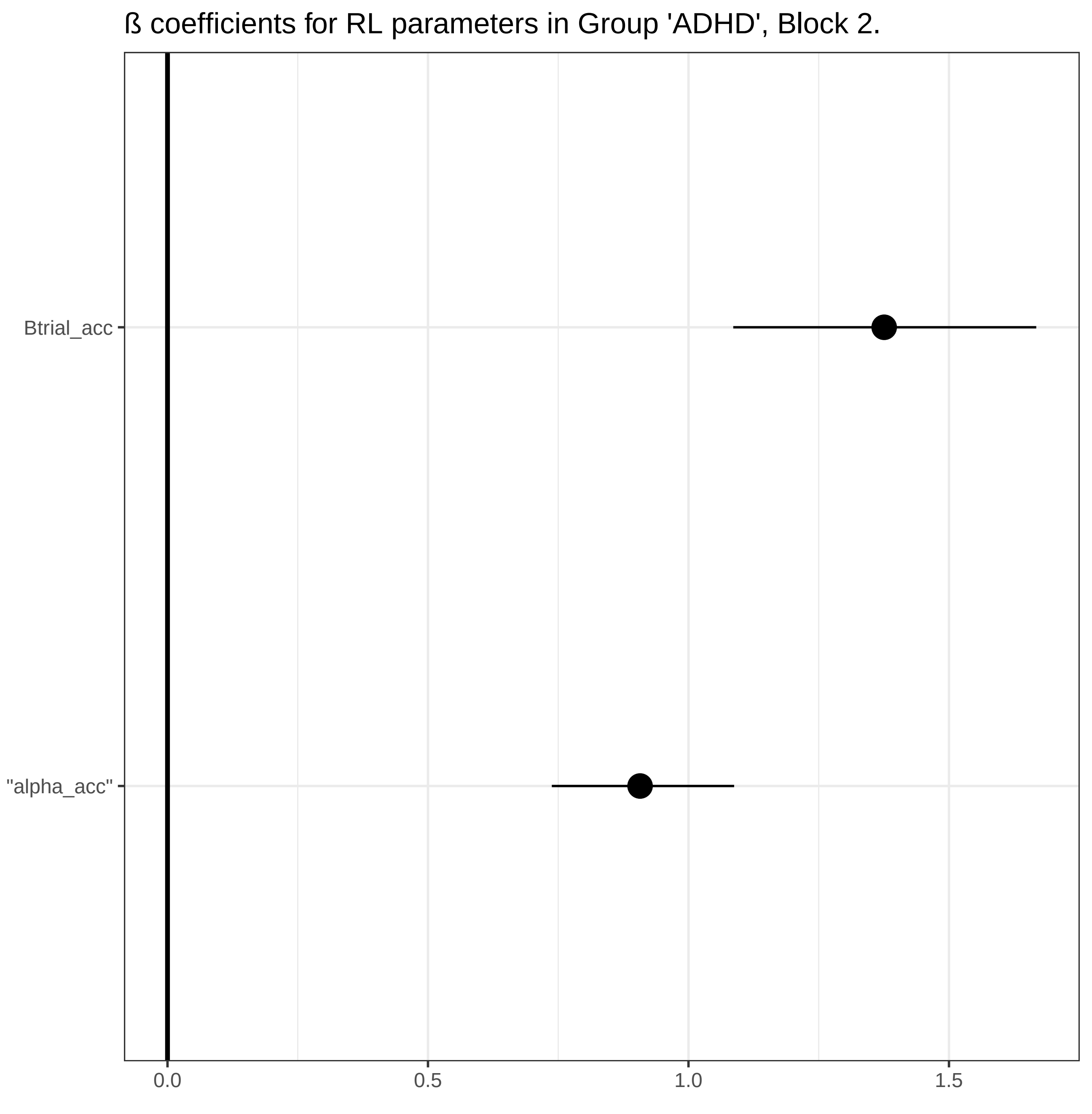
ß coefficients for RL parameters in Group 'ADHD', Block 2.
Btrial_acc
"alpha_acc"
0.0
0.5
1.0
1.5
