## Supplementary material for "Contingency-based flexibility mechanisms through a reinforcement learning model in adults with Attention-Deficit/Hyperactivity Disorder and Obsessive-Compulsive Disorder": suplementary material: Supplementary Material.docx

### **Instructions of Probabilistic Reversal Learning Task**

“In this task two squares of different colours will appear on the screen.

One square is considered "correct", most of the time you will get it right and earn 5 points.

The other square is considered "wrong", most of the time you will be warned of the mistake and you will lose 5 points.

Neither square is correct or incorrect 100% of the time. One is simply more likely to be correct than the other, and they may be interchanged at some point.

Pay attention to the scoreboard because your main task is to accumulate as many points as possible.

Press SPACEBAR to practice”

### **RL model definition**

Two learning rate parameters are defined as follows (Ahn et al., 2011; Ouden et al., 2013):

${EV}_{k, t+1}= {EV}_{k, t}+ \alpha_{rew}\left( R_{t}- {EV}_{k,t} \right) if R=1$	**Equation 1.**

${EV}_{k, t+1}= {EV}_{k, t}+ \alpha_{pun}\left( R_{t}- {EV}_{k,t} \right) if R=0$	**Equation 2.**

Following Equations 1 and 2, α_rew_ and α_pun_ are free parameters that inform about the weight that positive and negative prediction errors have on the expected value of a chosen option. Higher parameter values indicate a faster update of the values based on positive and negative outcomes. Just as in the simple Rescorla-Wagner model, the choice consistency parameter is extracted from a Softmax function (Equation 3).

$p_{t}\left( A \right)= \frac{e^{\tau\times{EV}_{t}(A)}}{e^{\tau\times{EV}_{t}(A)}+ e^{\tau\times{EV}_{t}(B)}}$	**Equation 3.**

This process will be performed by applying R software (R Core Team, 2019) and the “hBayesDM” package (Ahn et al., 2011). The model was applied in each group in an independent way, extracting 4 chains with 8000 iterations and saving 2000 iterations for the warm up. Simulations of real behavioural data based on the values of each RL parameter of each group will be performed in order to check the predictive feasibility of the model to make inferences about the behavioural mechanism that are assessed.

### **RL simulations**

All simulations were performed in R Software. 1000 simulations for each diagnostic group were obtained from the estimated values of each parameter obtained from the model. Using the same mathematical formulation for each parameter of the model, 1000 random combinations of parameters were given from each subject and 1000 probabilities of choices were simulated. In the following graphs, dotted lines represent the real probability of choice while dashed lines represent the simulated probabilities. Choice A and B are represented by red and black, respectively. The model showed an appropriate simulation of real data for each diagnostic group. Red and black color represent A and B choice probability, respectively. In supplementary figures 1 to 3, dashed lines represent simulated probability and dots represent real probability. Dotted lines represent the HDIs of the simulated data.

**Supplementary figure 1.**

*Real and simulated choice probability of in OCD group. Behavioural performance and learning in OCD group.*

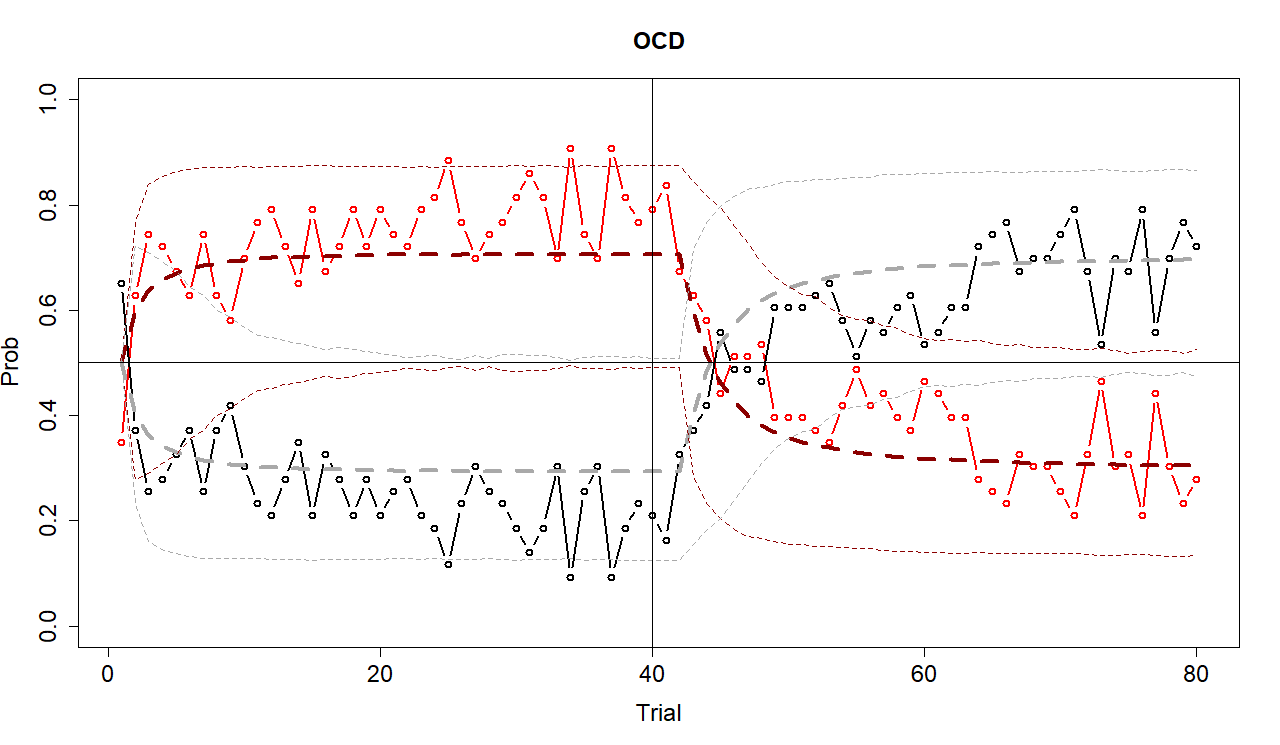

**Supplementary figure 2.**

*Real and simulated choice probability of in healthy group.*
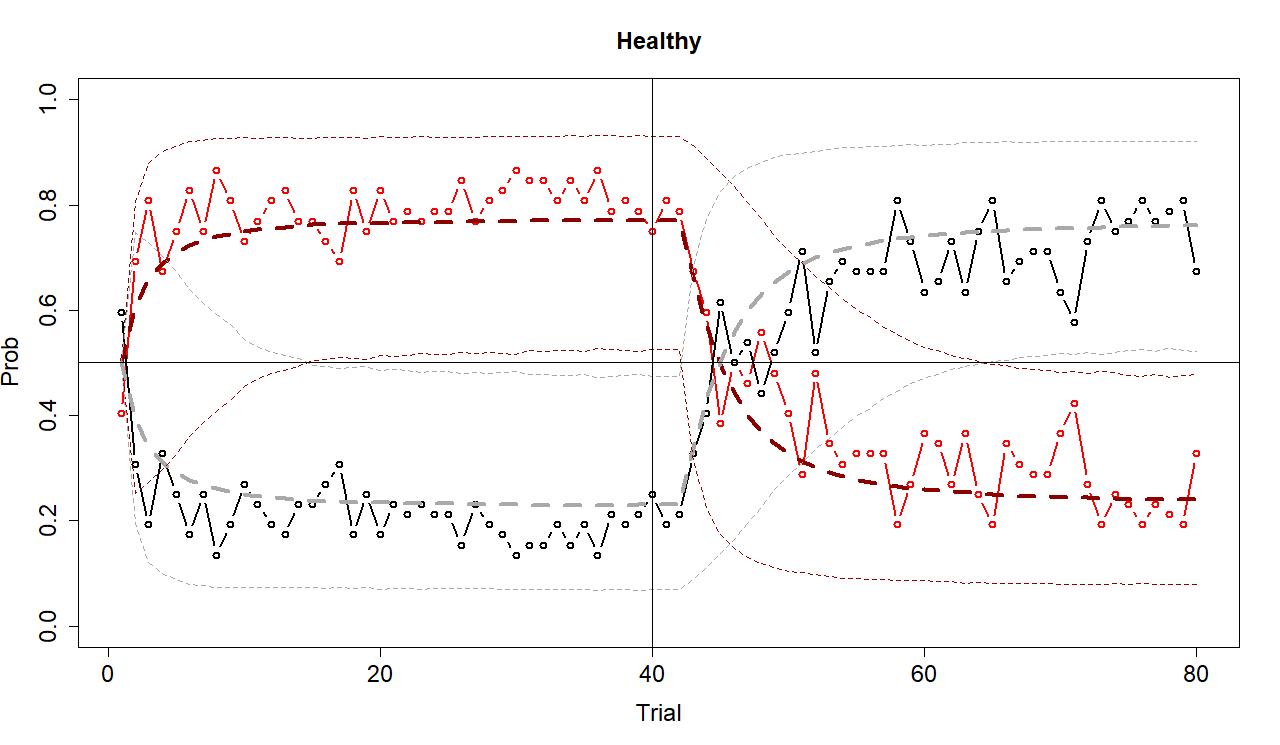

**Supplementary figure 3.**

*Real and simulated choice probability of in ADHD group.*

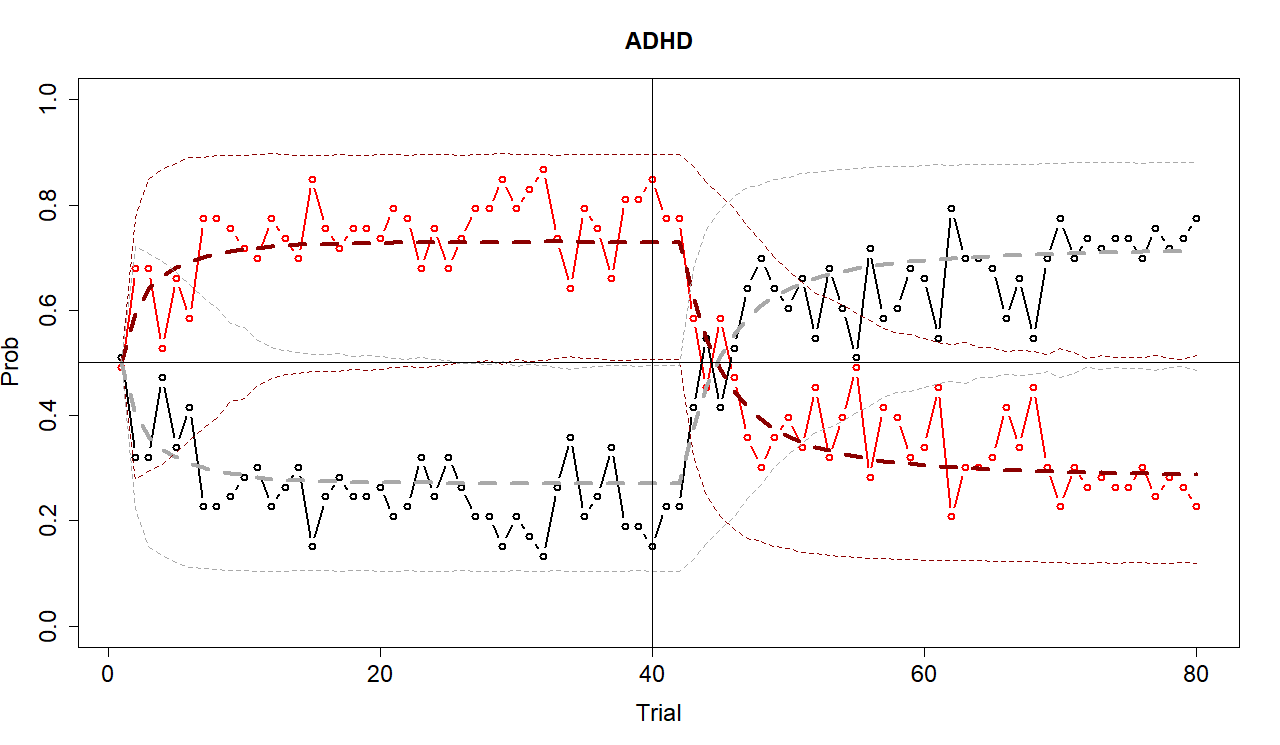

### **Bayesian Gerenalised Logistic Model**

The model is mathematically defined as follows:

$p_{\left[ i \right]}= {\alpha\_acc}_{\left[ Group\left[ i \right], Block\left[ i \right] \right]}+(\beta_{{Trial}_{\left[ Group\left[ i \right], Block\left[ i \right] \right]}}\times\left( \left( \left( \frac{60- n_{trial\left[ i \right]}}{59} \right)^{{(\gamma}_{\left[ Group\left[ i \right], Block\left[ i \right] \right]}\times\left( 1+\left( {mod}_{\gamma} \right) \right)}) \right) \right)+\beta_{{\alpha_{rew}}_{\left[ Group\left[ i \right], Block\left[ i \right] \right]}}\times\alpha_{rew\left[ i \right]}+ \beta_{{\alpha_{pun}}_{\left[ Group\left[ i \right], Block\left[ i \right] \right]}}\times\alpha_{pun\left[ i \right]}+ \beta_{\tau_{\left[ Group\left[ i \right], Block\left[ i \right] \right]}}\times\tau_{\left[ i \right]}+ \beta_{{rsFC1}_{\left[ Group\left[ i \right], Block\left[ i \right] \right]}}\times{rsFC1}_{\left[ i \right]}(\ldots) \beta_{{rsFC15}_{\left[ Group\left[ i \right], Block\left[ i \right] \right]}}\times{rsFC15}_{\left[ i \right]}$	**Equation 4.**

${mod}_{\gamma}= \beta_{\alpha_{rew\_\gamma[Group\left[ i \right], Block\left[ i \right]]}}\times\alpha_{rew\left[ i \right]}+ \beta_{\alpha_{pun\_\gamma[Group\left[ i \right], Block\left[ i \right]]}}\times\alpha_{pun\left[ i \right]}+\beta_{{\tau\_\gamma}_{[Group\left[ i \right], Block\left[ i \right]]}}\times\tau_{\left[ i \right]}$	**Equation 5.**

Equation 4 is designed to estimate the logit probability p_[i]_ of making a correct decision in each case [i]. The equation features an intercept, α_acc, which varies according to the group and block. This intercept indicates the likelihood of making a correct decision in the final trial for each group and block. The coefficients β_αrew_, β_αpun_, βτ, and β_rsFC1_ to β_rsFC15_ in the regression indicate the effects of normalized computational reinforcement learning parameters and resting state functional connectivity between Regions of Interest (ROIs) on p_[i]_ for each group and block. β_Trial_ is a coefficient measuring the variation in the probability of a correct answer from the first to the last trial for each group and block, adjusted by the exponential of γ divided by a value signifying the trial number [i]. In this task consisting of 40 trials per block, the trial number n_trial[i]_ is subtracted from 40 and then divided by 39, creating a scale from 1 (first trial) to 0 (last trial) for each trial number [i]. This formula evaluates the impact of individual trials on future decisions. The exponential γ, which also varies by group and block, is influenced by mod_γ_. As outlined in Equation 5, modγ is a specifically defined variable that moderates the effect of each reinforcement learning parameter on γ. The statistical analysis will utilize 95% Highest Density Intervals (HDIs). The RStan package (Stan Development Team, 2022) was used for model implementation, with data gathered through Markov Chain Monte Carlo sampling. Four chains were run, each with 4000 iterations, including 1000 for warm-up. To verify chain convergence, Traceplots and the Gelman-Rubin test were reviewed for all chains and parameters. These evaluations, including the Gelman-Rubin test (Gelman and Rubin, 1992), confirmed adequate convergence with all 𝑅̂ values being under 1.05. Additionally, Posterior predictive checks (PPCs) were conducted.

Prior distributions for all parameters were set according to Equations 6 to 30. In all the equations, the symbol “~” means “distributed as”, and N(x, y) indicates a normal distribution with mean = x and SD = y.

$\alpha$_acc_[Group[i],Block[i]]_ ~ *N*(0, 1)	**Equation 6.**

β_Trial[Group[i],Block[i]]_ ~ *N*(0, 1.5);	**Equation 7.**

γ_[Group[i],Block[i]]_ ~ *N*(2, 0.1);	**Equation 8.**

β_α_pun_γ[Group[i],Block[i]]_ ~ *N*(0, 0.1);	**Equation 9.**

β_α_rew_γ[Group[i],Block[i]]_ ~ *N*(0, 0.1);	**Equation 10.**

βτ_γ_[Group[i],Block[i]]_ ~ *N*(0, 0.1);	**Equation 11.**

β_α_pun[Group[i],Block[i]]_ ~ *N*(0, 0.2);	**Equation 12.**

β_α_rew[Group[i],Block[i]]_ ~ *N*(0, 0.2);	**Equation 13.**

βτ_[Group[i],Block[i]]_ ~ *N*(0, 0.2);	**Equation 14.**

β_rsFC1[Group[i]_ ~ *N*(0, 0.1);	**Equation 15.**

**…**

β_rsFC15[Group[i]_ ~ *N*(0, 0.1);	**Equation 30.**

### **Detailed statistical information about posterior distribution differences.**

**Supplementary table 1.**

Mean of the differences (and 95% HDIs) between diagnostic groups of the estimated parameters and regression coefficients in each block.

|  | **Group** | **Block 1** | **Block 2** |
| --- | --- | --- | --- |
| **α_acc** | HC vs. OCD | .115  (-.176, .433) | **.407**  **(.122, .686)** |
|  | HC vs. ADHD | .069  (-.230, .343) | **.406**  **(.144, .663)** |
|  | OCD vs. ADHD | -.046  (-.331, .254) | -.001  (-.262, .272) |
| **β_Trial_** | HC vs. OCD | - | .341  (-.113, .787) |
|  | HC vs. ADHD | - | **.438**  **(.031, .865)** |
|  | OCD vs. ADHD | - | .097  (-.347, .528) |
| **β_α_rew_** | HC vs. OCD | -.147  (-.305, .013) | .114  (-.031, .269) |
|  | HC vs. ADHD | -.083  (-.255, .101) | .106  (-.058, .273) |
|  | OCD vs. ADHD | .065  (-.123, .242) | -.008  (-.184, .160) |
| **β_α_pun_** | HC vs. OCD | **.218**  **(.038, .394)** | -.055  (-.112, .216) |
|  | HC vs. ADHD | **.263**  **(.108, .425)** | **.206**  **(.058, .348)** |
|  | OCD vs. ADHD | .045  (-.122, .216) | .151  (-.004, .306) |
| **βτ** | HC vs. OCD | -.099  (-.281, .076) | **.204**  **(.043, .366)** |
|  | HC vs. ADHD | .066  (-.100, .219) | .114  (-.028, .261) |
|  | OCD vs. ADHD | .165  (-.006, .336) | -.090  (-.249, .065) |

*Note*. Pairwise comparisons that showed credible differences (95% HDI does not include zero) are boldfaced.

**Supplementary table 2.**

Means (and 95% HDIs) of the estimated parameters and regression coefficients in each block for each group.

|  | **Group** | **Block 1** | **Block 2** |
| --- | --- | --- | --- |
| **α_acc** | HC | **1.437**  **(1.216, 1.646)** | **1.313**  **(1.119, 1.507)** |
|  | OCD | **1.322**  **(1.103, 1.537).** | **.906**  **(.712, 1.118)** |
|  | ADHD | **1.368**  **(1.182, 1.560)** | **.907**  **(.738, .1.088)** |
| **β_Trial_** | HC | - | **1.813**  **(1.516, 2.120)** |
|  | OCD | - | **1.472**  **(1.149, 1.811)** |
|  | ADHD | - | **1.376**  **(1.086, 1.667)** |
| **β_α_rew_** | HC | **-.182**  **(-.292, -.069)** | -.051  (-.150, .053) |
|  | OCD | -.035  (-.150, .079) | **-.165**  **(-.281, -.055)** |
|  | ADHD | -.100  (-.248, .032) | **-.157**  **(-.286, -.027)** |
| **β_α_pun_** | HC | **.211**  **(.096, .335)** | **.251**  **(.142, .359)** |
|  | OCD | -.007  (-.139, .128) | **.196**  **(.077, .321)** |
|  | ADHD | -.052  (-.152, .058) | .044  (-.053, .141) |
| **βτ** | HC | **.168**  **(.049, .285)** | **.209**  **(.107, .317)** |
|  | OCD | **.267**  **(.134, .401)** | .005  (-.121, .125) |
|  | ADHD | **.102**  **(.002, .210)** | .095  (-.003, .194) |

*Note*. Means of the regression coefficients and the estimated parameters that showed 95% HDI not including zero value are boldfaced.

**Supplementary table 3.**

Means and means of the differences (and 95% HDIs) of the regression coefficients for each ROI for each group in acquisition block.

|  | **HC** | **OCD** | **ADHD** | **HC – OCD** | **HC – ADHD** | **OCD - ADHD** |
| --- | --- | --- | --- | --- | --- | --- |
| **lOFC-rOFC** | .143  (-.030, 0.309) | .007  (-.163, .175) | .060  (-.102, .242) | .136  (-.117, .373) | .082  (-.167, .321) | -.054  (-.294, .188) |
| **lOFC-lDLPFC** | .004  (-.176, .183) | .002  (-.182, .181) | .088  (-.093, .265) | .001  (-.261, .256) | -.085  (-.326, .181) | -.086  (-.331, .189) |
| **lOFC-rDLPFC** | .031  (-.153, .206) | .087  (-.110, .261) | .020  (-.166, .203) | -.056  (-.311, .204) | .011  (-.250, .261) | .068  (-.187, .331) |
| **lOFC-lpPC** | .065  (-.122, .260) | .033  (-.157, .222) | .090  (-.103, .277) | .033  (-.224, .297) | -.025  (-.295, .239) | -.057  (-.324, .204) |
| **lOFC-rpPC** | .087  (-.103, .276) | -.019  (-.214, .173) | -.010  (-.189, .187) | .107  (-.168, .373) | .097  (-.166, .363) | -.009  (-.291, .252) |
| **rOFC-lDLPFC** | -.014  (-.201, .167) | .059  (-.130, .243) | -.004  (-.184, .171) | -.072  (-.339, .178) | -.010  (-.262, .250) | .062  (-.203, .319) |
| **rOFC-rDLPFC** | .026  (-.154, .200) | .111  (-.065, .300) | -.032  (-.214, .146) | -.085  (-.341, .168) | .058  (-.199, .305) | .143  (-.110, .400) |
| **rOFC-lpPC** | .019  (-.169, .216) | .067  (-.124, .249) | .014  (-.178, .197) | -.048  (-.323, .219) | .005  (-.251, .282) | .053  (-.210, .320) |
| **rOFC-rpPC** | .071  (-.121, .262) | -.012  (-.206, .167) | -.070  (-.256, .111) | .083  (-.177, .351) | .138  (-.137, .404) | .055  (-.204, .325) |
| **lDLPFC-rDLPFC** | .016  (-.163, .191) | .100  (-.093, .273) | .097  (-.079, .277) | -.084  (-.327, .181) | -.080  (-.334, .171) | .004  (-.247, .262) |
| **lDLPFC-lpPC** | .030  (-.159, .222) | .081  (-.104, .271) | .126  (-.072, .310) | -.052  (-.319, .218) | -.098  (-.363, .177) | -.045  (-.319, .216) |
| **lDLPFC-rpPC** | .045  (-.146, .233) | -.033  (-.223, .153) | .022  (-.171, .212) | .077  (-.190, .343) | .022  (-.242, .295) | -.053  (-.319, .221) |
| **rDLFPC-lpPC** | .023  (-.155, .206) | .092  (-.099, .280) | .100  (-.090, .282) | -.070  (-.331, .190) | -.078  (-.344, .183) | -.008  (-.277, .265) |
| **rDLPFC-rpPC** | .072  (-.114, .256) | -.007  (-.194, .188) | -.019  (-.203, .167) | .080  (-.180, .350) | .092  (-.175, .348) | .012  (-.257, 278) |
| **lpPC-rpPC** | **.234**  **(.061, .418)** | .035  (-.134, .226) | -.057  (-.233, .125) | .199  (-.057, .456) | **.292**  **(.035, .540)** | .092  (-.700, .347) |

*Note*. Means and pairwise comparisons that showed credible differences (95% HDI does not include zero) are boldfaced.

**Supplementary table 4.**

Means and means of the differences (and 95% HDIs) of the regression coefficients for each ROI for each group in reversal block.

|  | **HC** | **OCD** | **ADHD** | **HC – OCD** | **HC – ADHD** | **OCD - ADHD** |
| --- | --- | --- | --- | --- | --- | --- |
| **lOFC-rOFC** | .015  (-.153, .186) | .001  (-.178, .167) | .063  (-.108, .229) | .014  (-.231, .250) | -.049  (-.285, .184) | -.063  (-.304, .180) |
| **lOFC-lDLPFC** | -.053  (-.235, .126) | -.048  (-.232, .129) | .071  (-.106, .245) | -.006  (-.256, .256) | -.125  (-.371, .131) | -.119  (-.361, .145) |
| **lOFC-rDLPFC** | .013  (-.171, .192) | .006  (-.181, .180) | .130  (-.045, .318) | .007  (-.241, .274) | -.117  (-.368, .141) | -.124  (-.370, .144) |
| **lOFC-lpPC** | .045  (-.151, .192) | -.013  (-.197, .183) | .046  (-.138, .242) | .058  (-.205, .340) | -.001  (-.275, .264) | -.059  (-.343, .209) |
| **lOFC-rpPC** | .008  (-.185, .194) | .016  (-.184, .208) | .071  (-.112, .264) | -.008  (-.288, .247) | -.063  (-.323, .204) | -.055  (-.329, .205) |
| **rOFC-lDLPFC** | -.072  (-.254, .110) | .078  (-.112, .266) | -.078  (-.253, .102) | -.151  (-.411, .115) | .005  (-.250, .259) | .156  (-.102, .414) |
| **rOFC-rDLPFC** | -.078  (-.255, .106) | .021  (-.156, .198) | .033  (-.146, .208) | -.099  (-.354, .163) | -.111  (-.366, .141) | -.012  (-.261, .236) |
| **rOFC-lpPC** | -.014  (-.208, .169) | .036  (-.157, .225) | -.031  (-.223, .161) | -.050  (-.317, .224) | .017  (-.245, .290) | .067  (-.202, .340) |
| **rOFC-rpPC** | -.123  (-.316, .061) | .003  (-.181, .190) | .027  (-.159, .210) | -.126  (-.384, .139) | -.151  (-.415, .109) | -.024  (-.283, .236) |
| **lDLPFC-rDLPFC** | -.025  (-.199, .156) | -.031  (-.212, .153) | .045  (-.185, .189) | .006  (-.252, .263) | -.070  (-.324, .181) | -.076  (-.331, .181) |
| **lDLPFC-lpPC** | -.005  (-.183, .192) | .018  (-.176, .208) | .004  (-.141, .232) | -.022  (-.295, .238) | -.009  (-.273, .253) | .013  (-.265, .273) |
| **lDLPFC-rpPC** | -.051  (-.244, .131) | .024  (-.161, .213) | .040  (-.141, .176) | -.075  (-.338, .193) | -.090  (-.348, .195) | -.015  (-.287, .246) |
| **rDLFPC-lpPC** | -.004  (-.188, .187) | .015  (-.168, .208) | -.006  (-.198, .176) | -.019  (-.283, .225) | .002  (-.261, .266) | .021  (-.254, .273) |
| **rDLPFC-rpPC** | -.093  (-.280, .091) | .041  (-.151, .225) | .058  (-.131, .241) | -.135  (-.382, .145) | -.151  (-.414, .118) | -.016  (-.286, .242) |
| **lpPC-rpPC** | -.038  (-.215, .142) | .072  (-.108, .256) | .035  (-.145, .215) | -.109  (-.356, .146) | -.072  (-.323, .187) | .037  (-.218, .286) |

*Note*. Means and pairwise comparisons that showed credible differences (95% HDI does not include zero) are boldfaced.
