## Supplementary material for "Contingency-based flexibility mechanisms through a reinforcement learning model in adults with Attention-Deficit/Hyperactivity Disorder and Obsessive-Compulsive Disorder": suplementary material: NOTE.txt

The tittle of each graph follows the following structure:

- parameter_name[(group: 1, 2, 3), (block: 1, 2)] where group: 1, 2 or 3 corresponds to Healthy control, OCD and ADHD groups
  and block: 1 or 2 corresponds to acquisition or reversal block respectively.
