## Supplementary material for "Contingency-based flexibility mechanisms through a reinforcement learning model in adults with Attention-Deficit/Hyperactivity Disorder and Obsessive-Compulsive Disorder": suplementary material: Traceplots.pdf

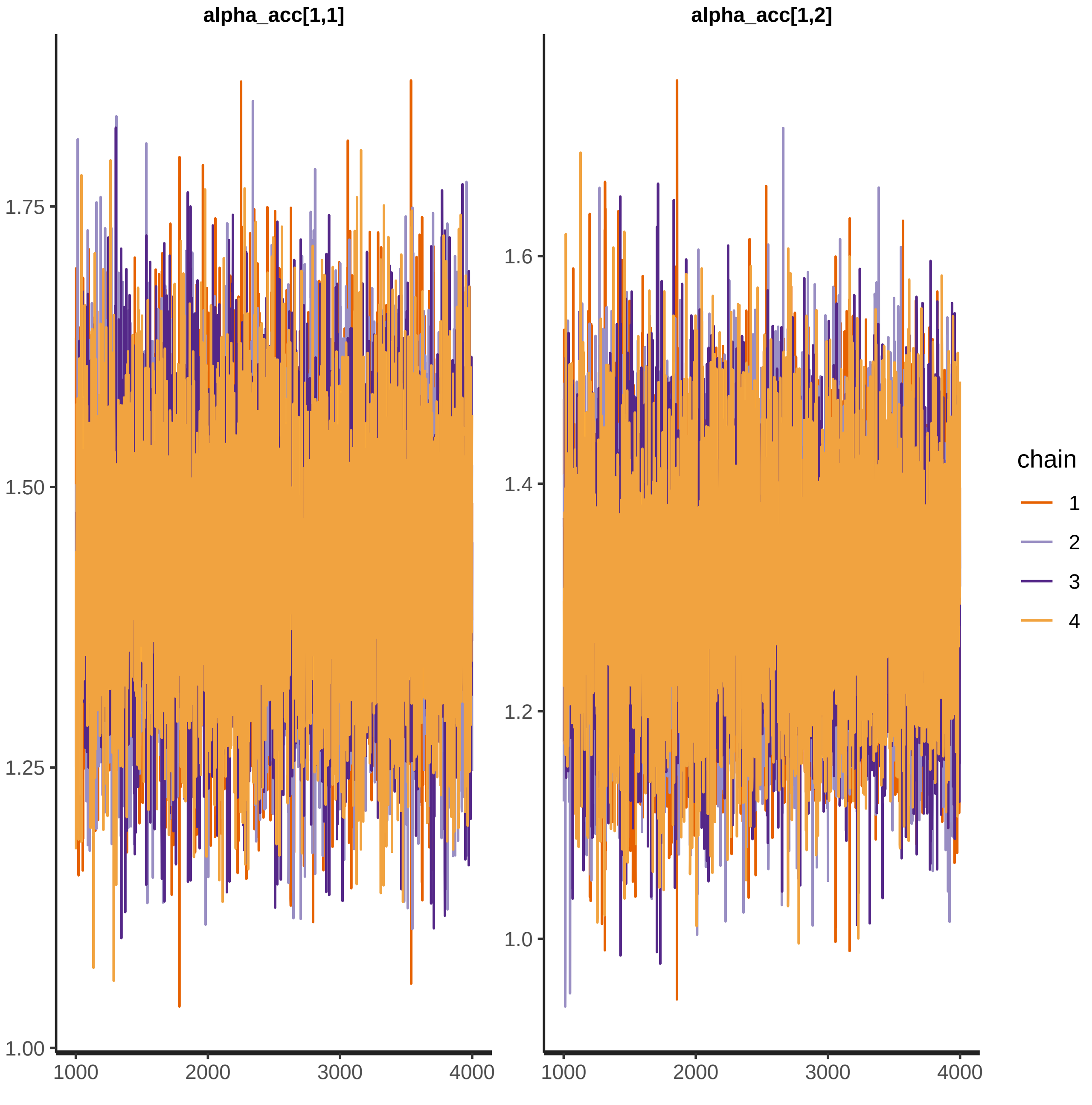

alpha_acc[1,1]
alpha_acc[1,2]
1.75
1.50
1.25
1.00
1.6
1.4
1.2
1.0
1000
2000
3000
4000
chain
1
2
3
4

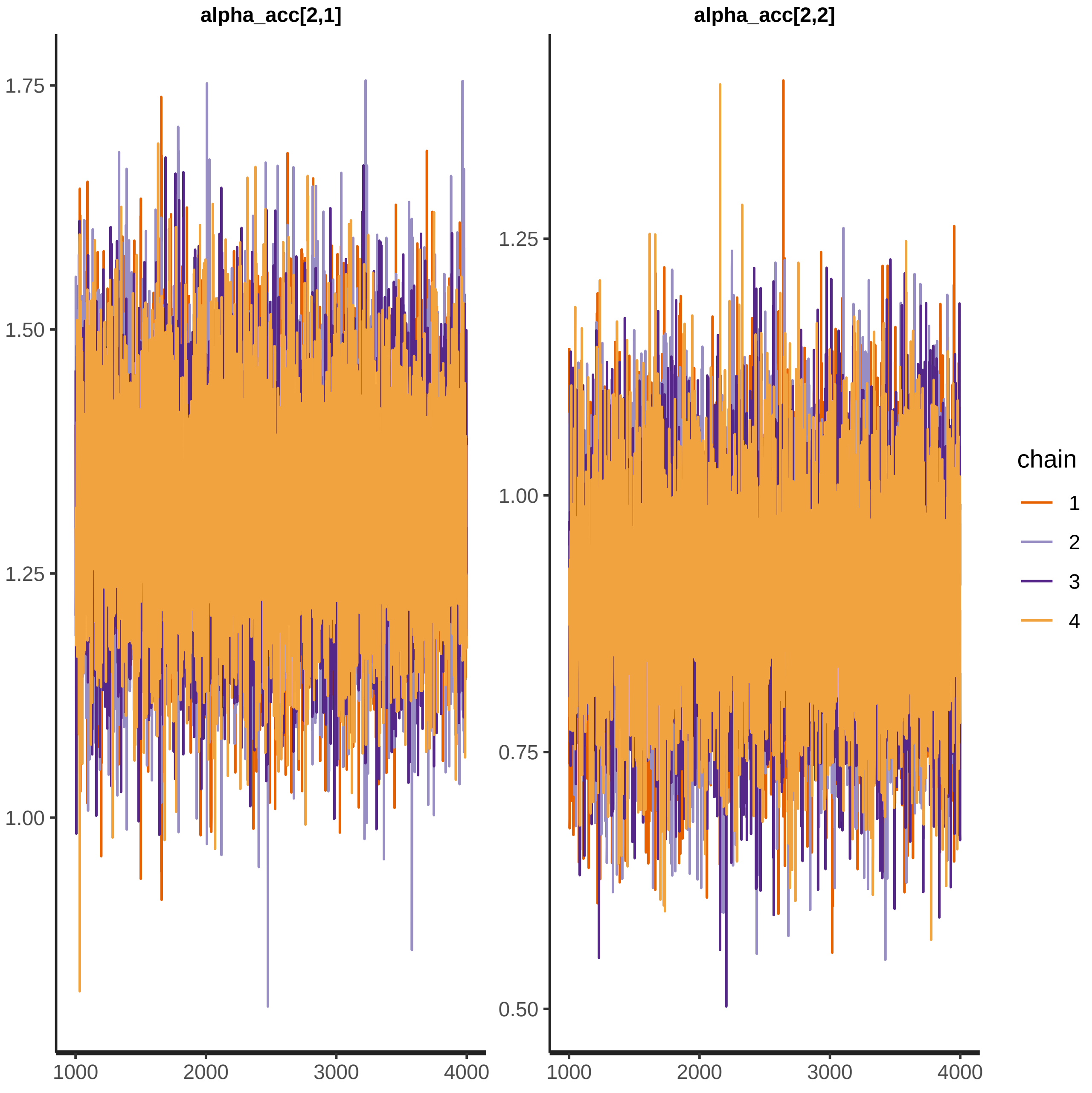

alpha_acc[2,1]
alpha_acc[2,2]
1.75
1.50
1.25
1.00
0.75
0.50
1000
2000
3000
4000
chain
1
2
3
4

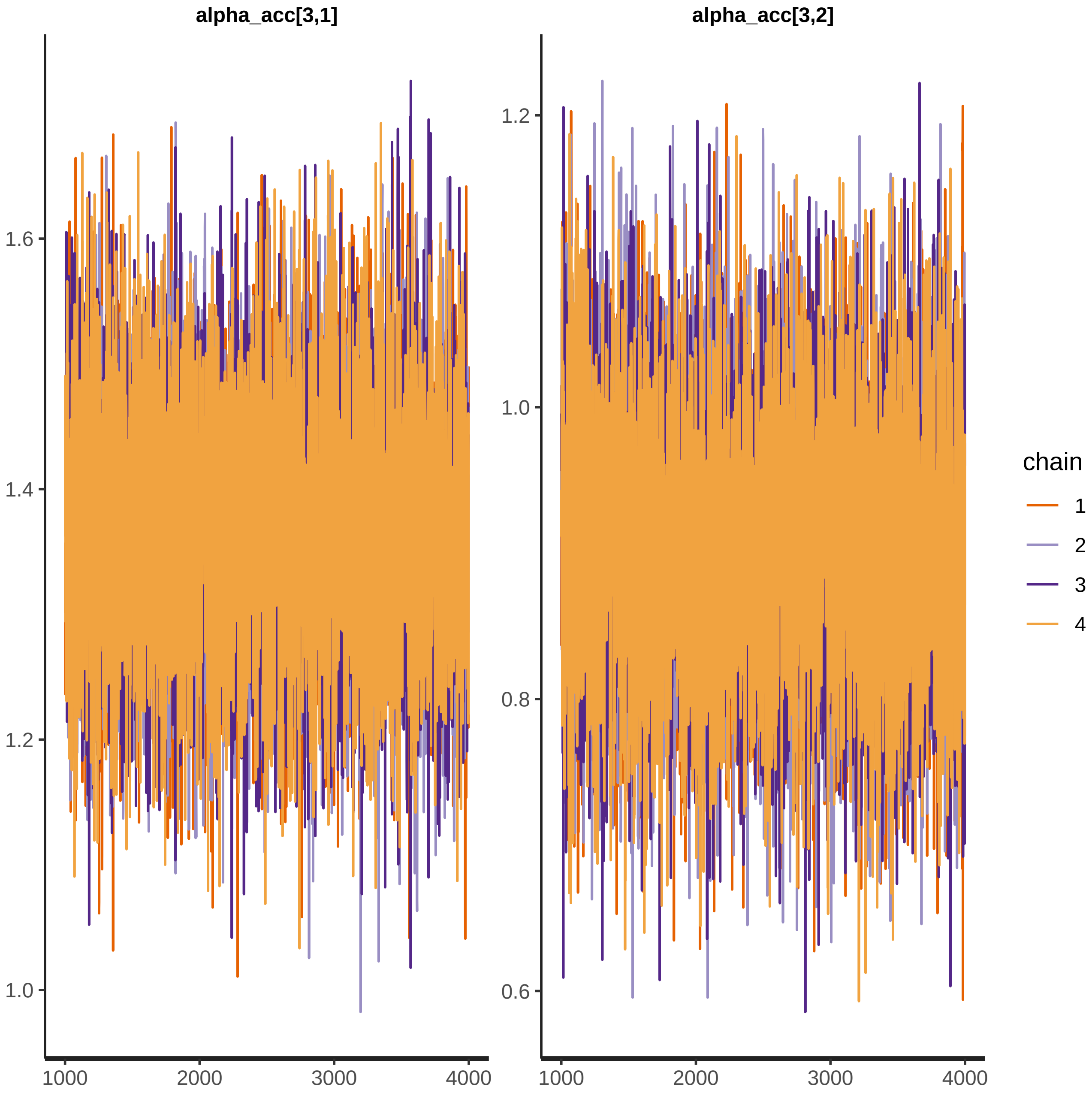

alpha_acc[3,1]
alpha_acc[3,2]
1.6
1.4
1.2
1.0
1.2
1.0
0.8
0.6
1000
2000
3000
4000
1000
2000
3000
4000
chain
1
2
3
4

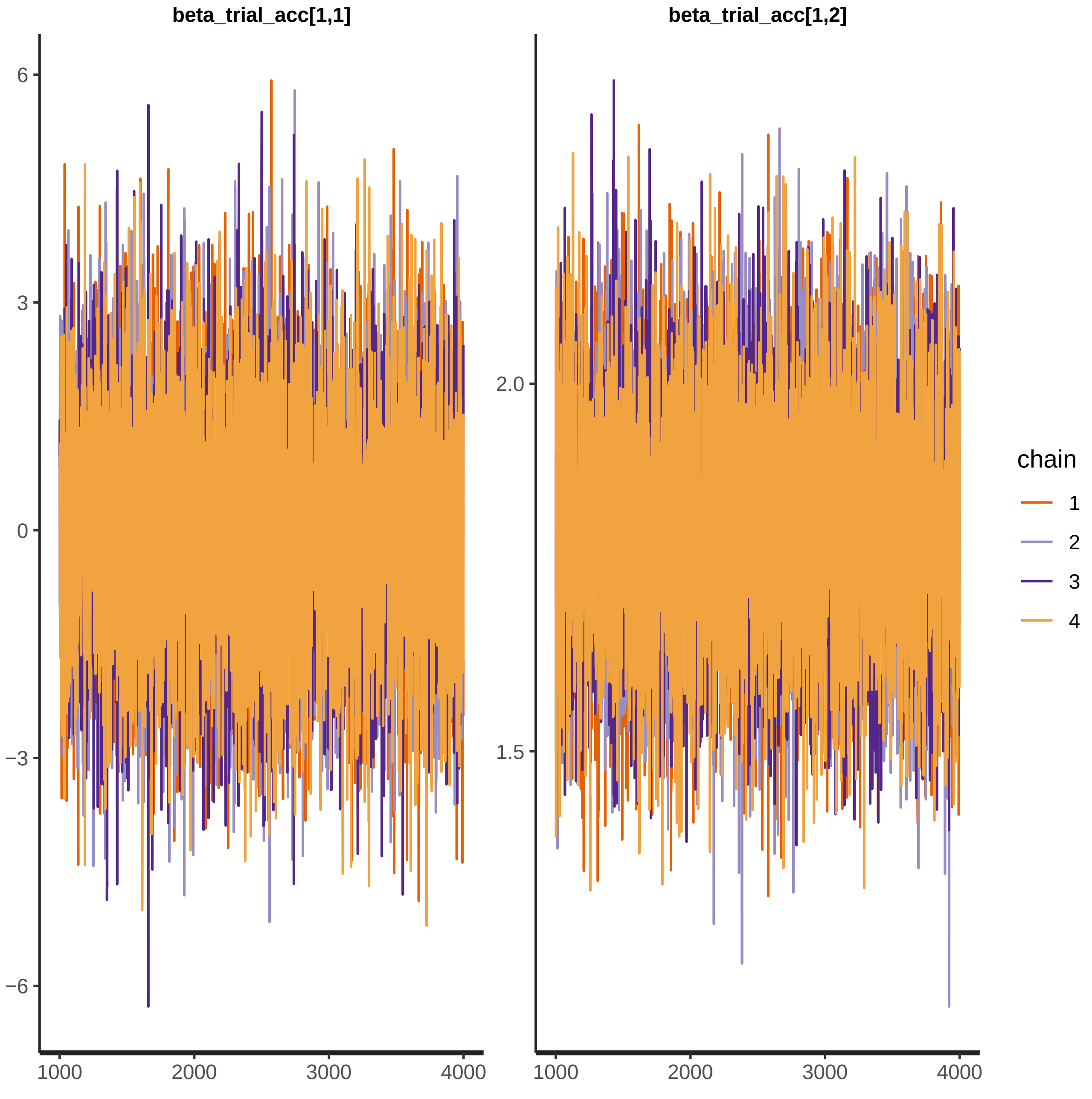

beta_trial_acc[1,1]
beta_trial_acc[1,2]
6
3
0
−3
−6
2.0
1.5
1000
2000
3000
4000
chain
1
2
3
4

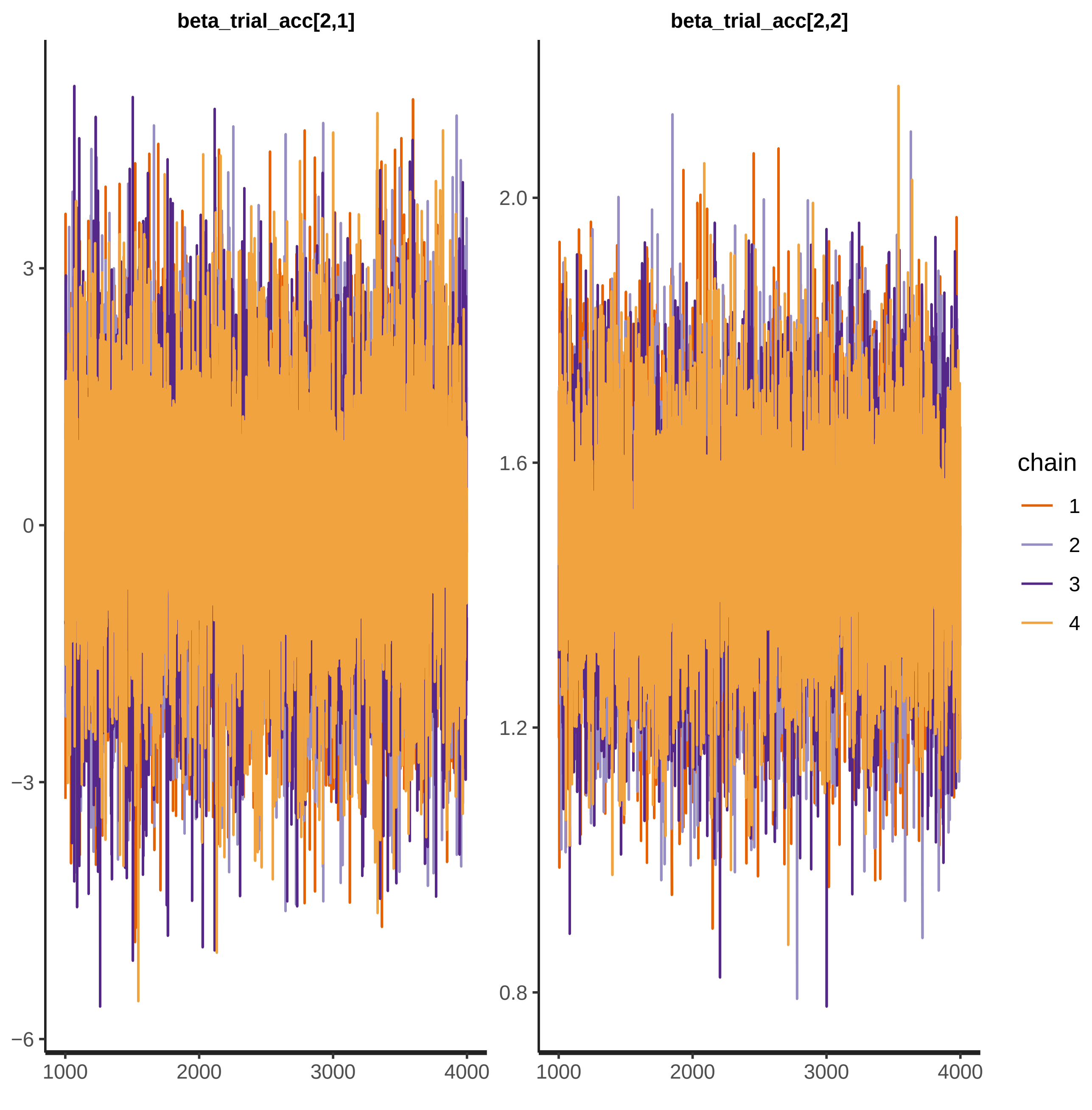
beta_trial_acc[2,1]
beta_trial_acc[2,2]
3
0
−3
−6
2.0
1.6
1.2
0.8
1000
2000
3000
4000
chain
1
2
3
4

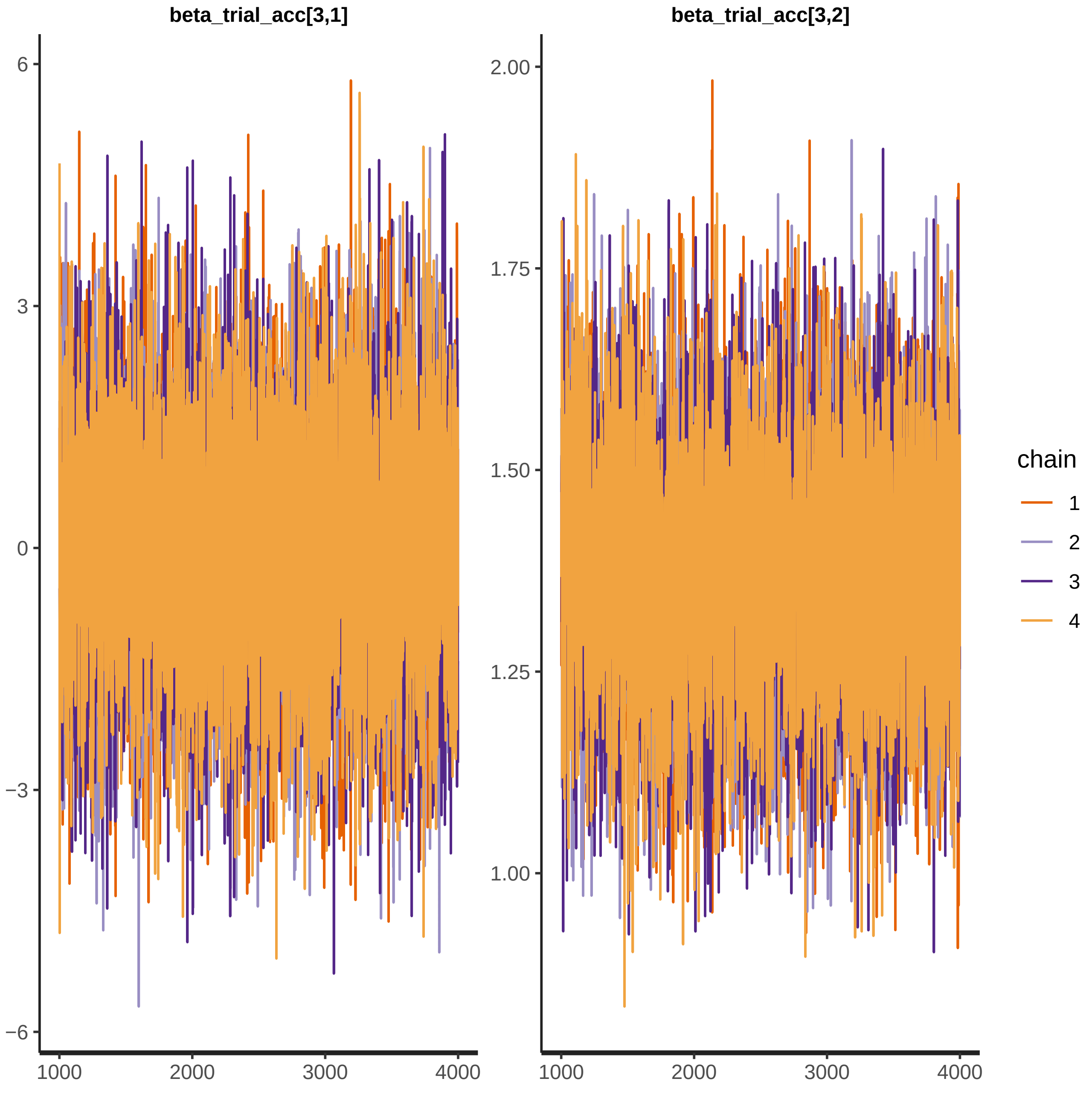
beta_trial_acc[3,1]
beta_trial_acc[3,2]
6
3
0
−3
−6
2.00
1.75
1.50
1.25
1.00
1000
2000
3000
4000
chain
1
2
3
4

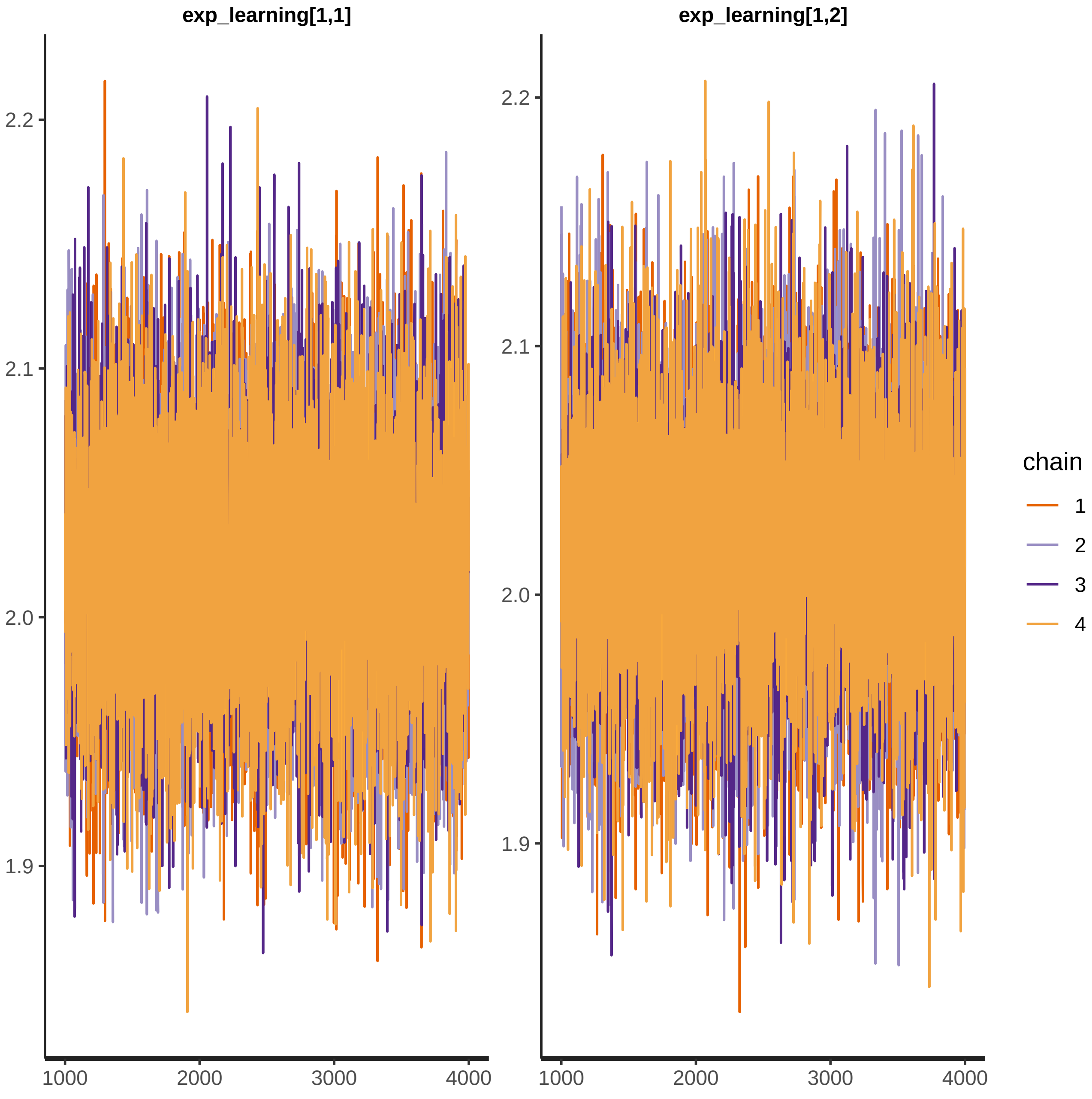
exp_learning[1,1]
exp_learning[1,2]
2.2
2.1
2.0
1.9
1000
2000
3000
4000
chain
1
2
3
4

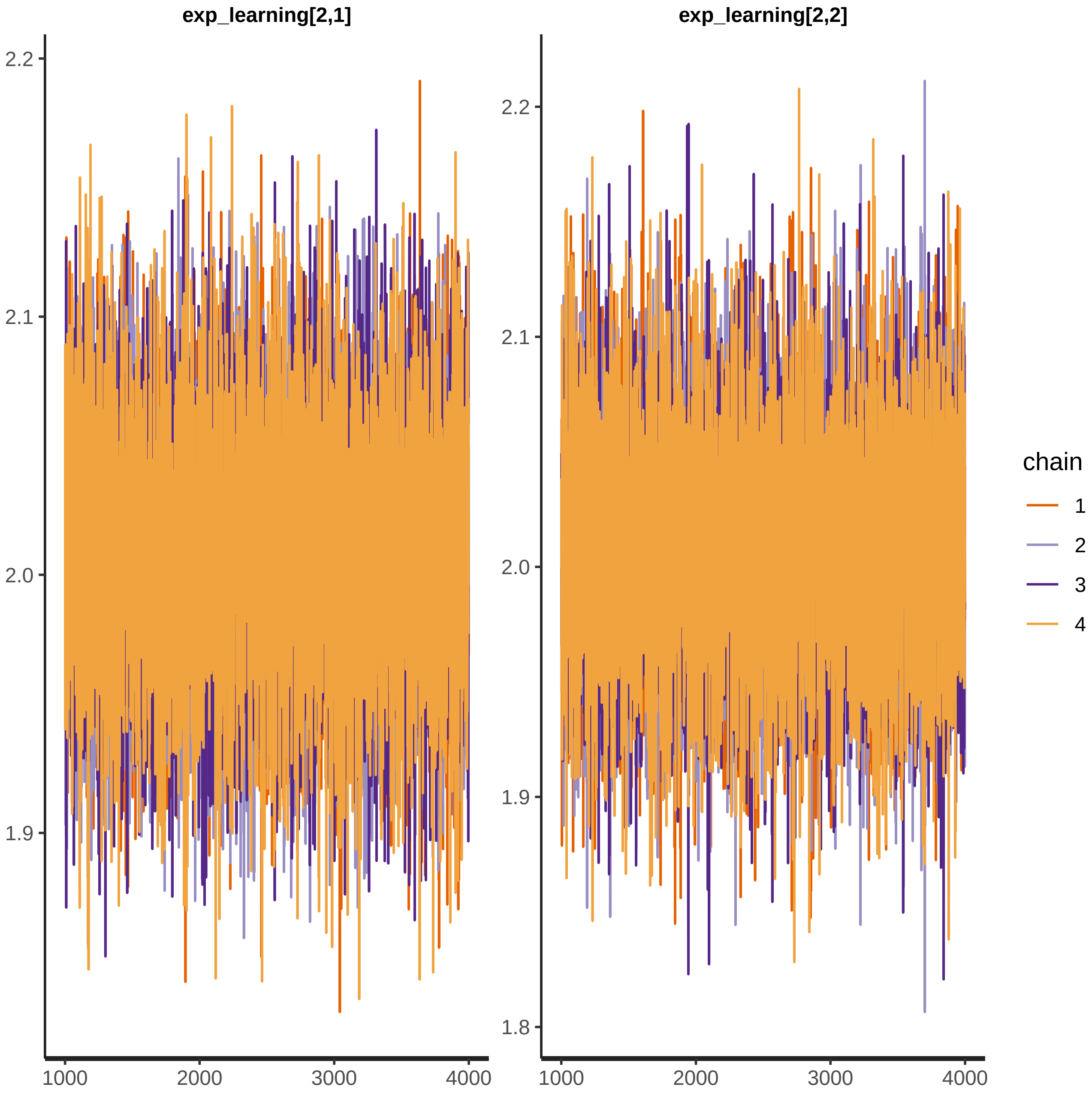

exp_learning[2,1]
exp_learning[2,2]
2.2
2.1
2.0
1.9
1.8
1000
2000
3000
4000
chain
1
2
3
4

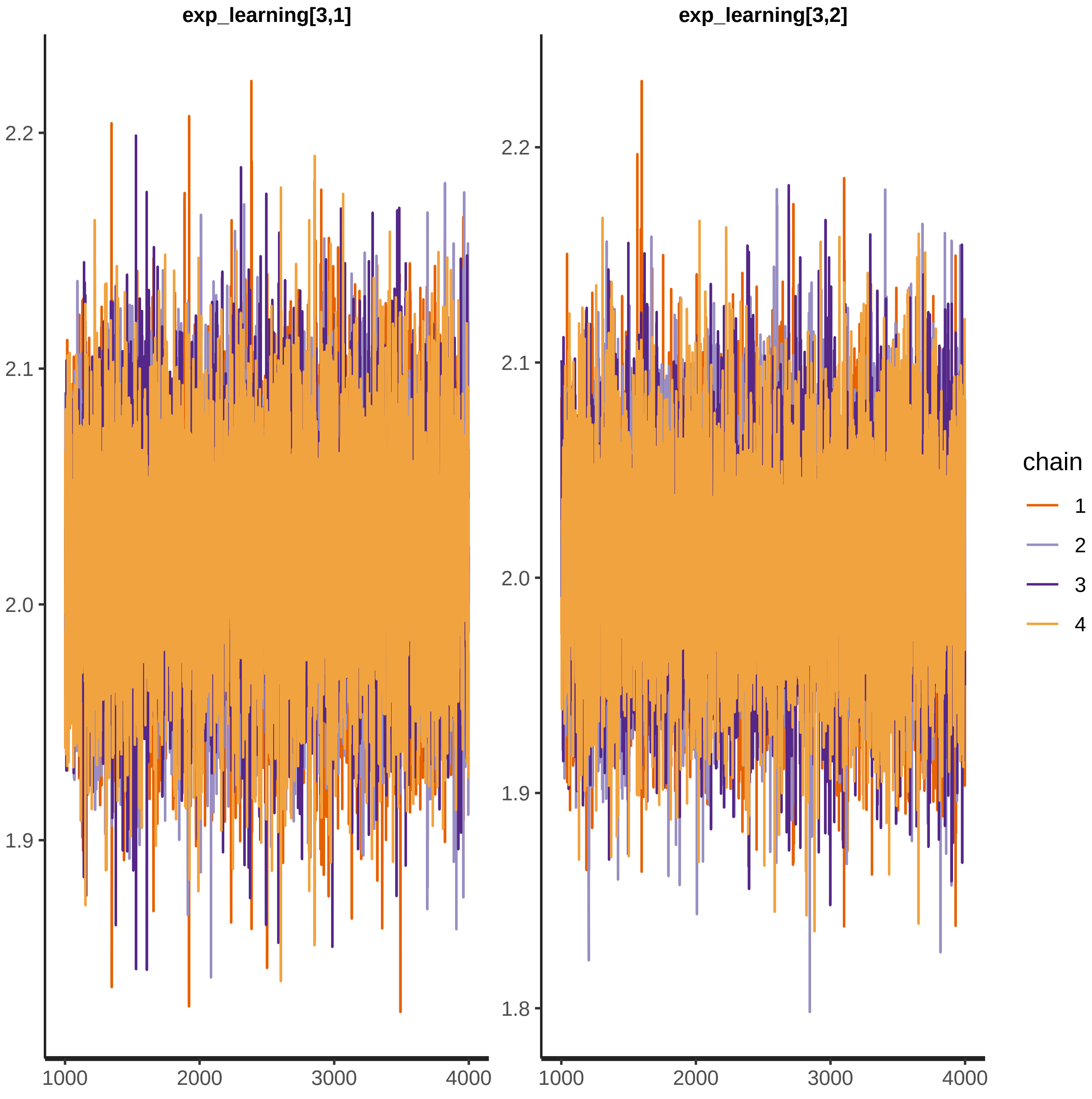

exp_learning[3,1]
exp_learning[3,2]
2.2
2.1
2.0
1.9
1.8
1000
2000
3000
4000
chain
1
2
3
4

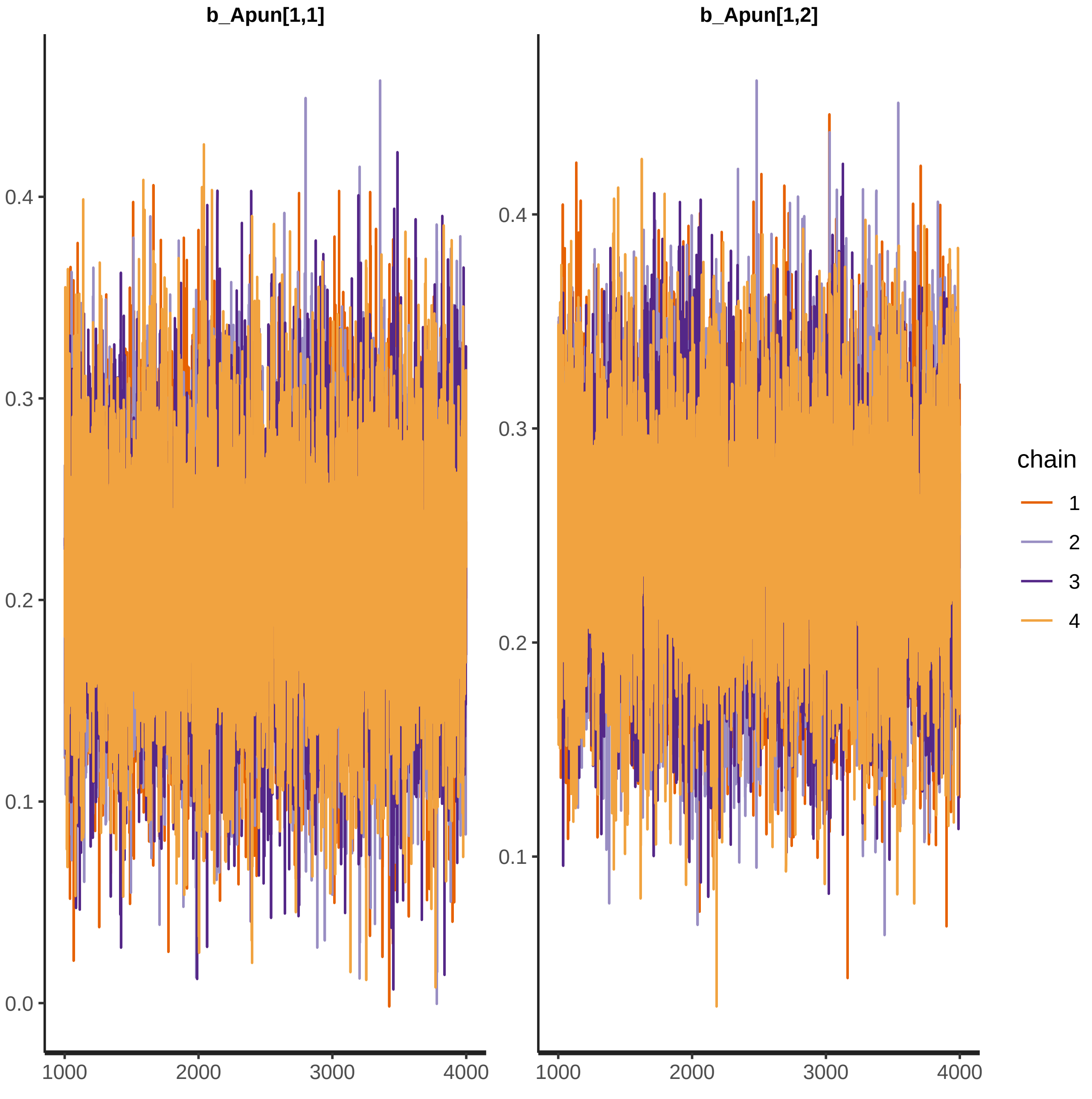

b_Apun[1,1]
b_Apun[1,2]
0.4
0.3
0.2
0.1
0.0
1000
2000
3000
4000
chain
1
2
3
4

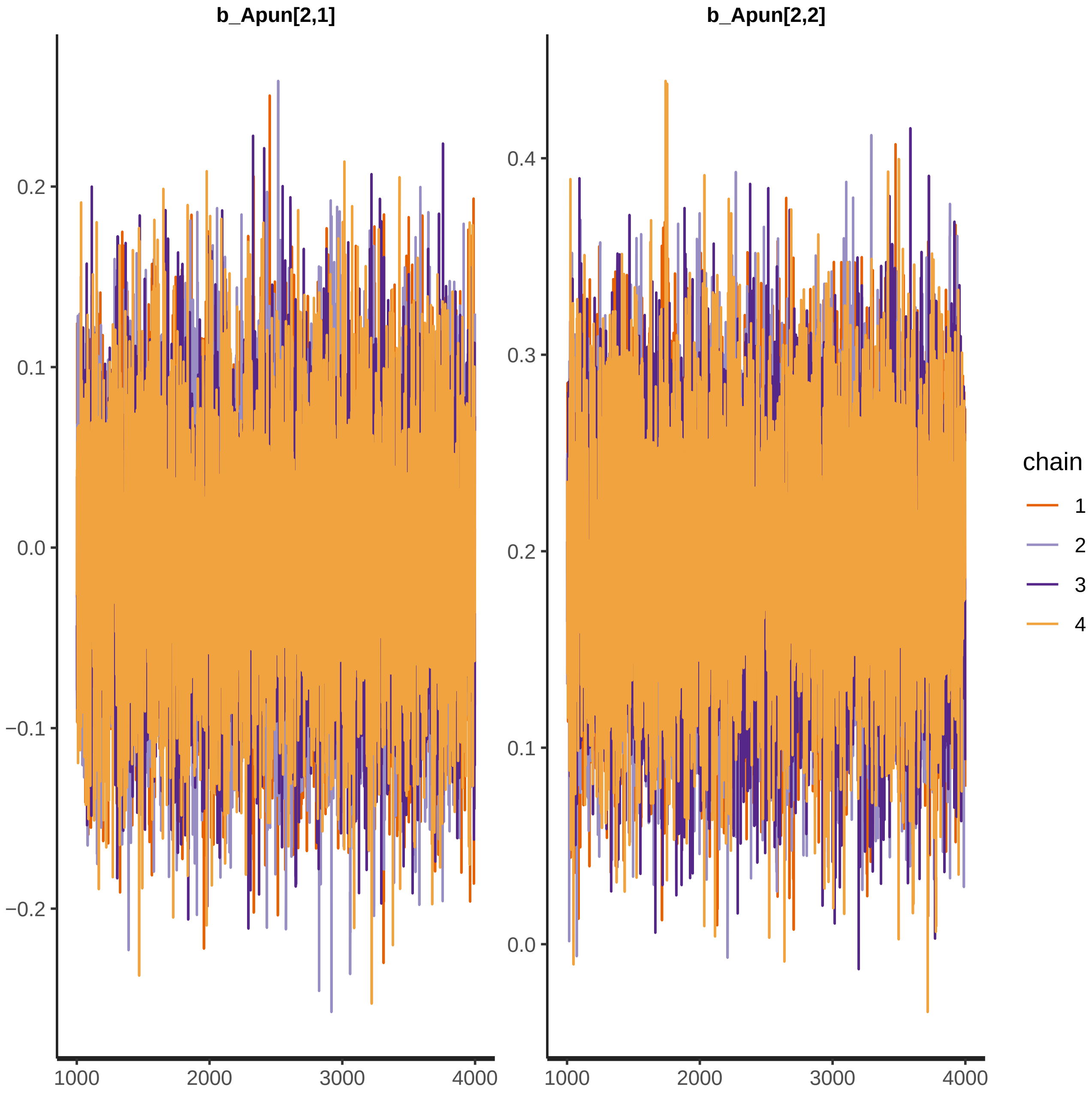
b_Apun[2,1]
b_Apun[2,2]
0.2
0.1
0.0
−0.1
−0.2
0.4
0.3
0.2
0.1
0.0
1000
2000
3000
4000
chain
1
2
3
4

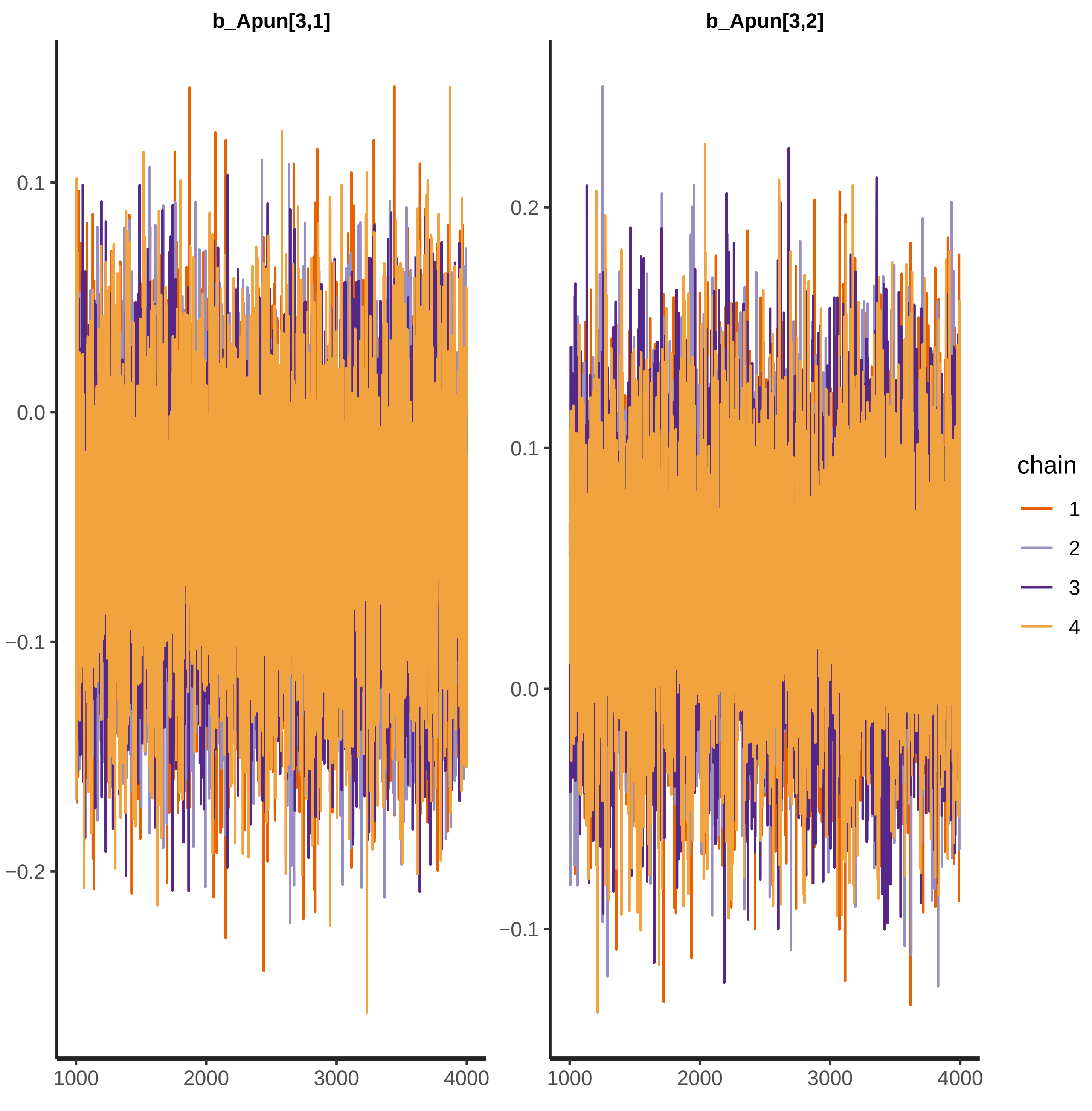
b_Apun[3,1]
b_Apun[3,2]
0.1
0.0
−0.1
−0.2
0.2
0.1
0.0
−0.1
1000
2000
3000
4000
1000
2000
3000
4000
chain
1
2
3
4

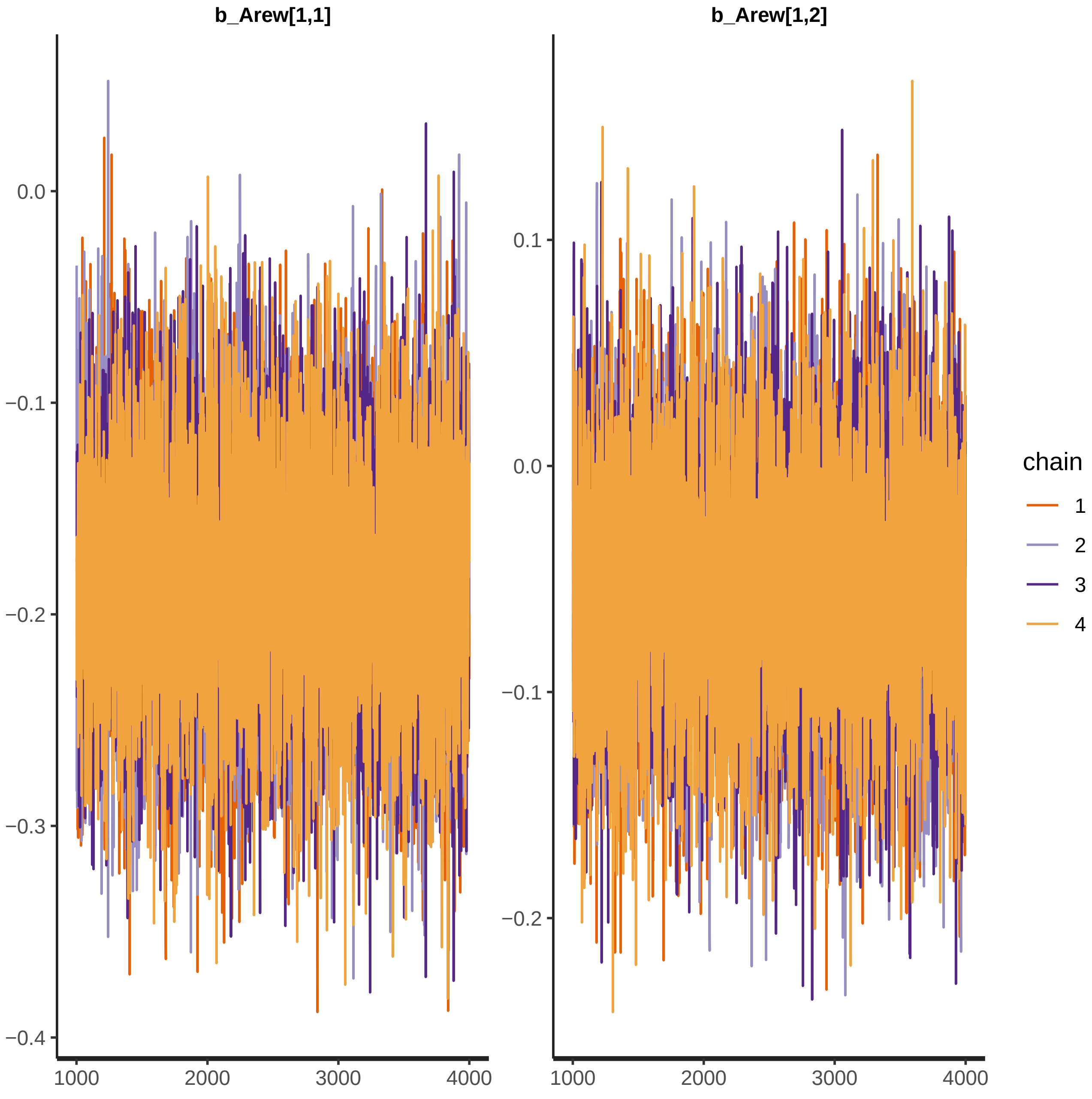

b_Arew[1,1]
b_Arew[1,2]
0.0
−0.1
−0.2
−0.3
−0.4
0.1
1000
2000
3000
4000
chain
1
2
3
4

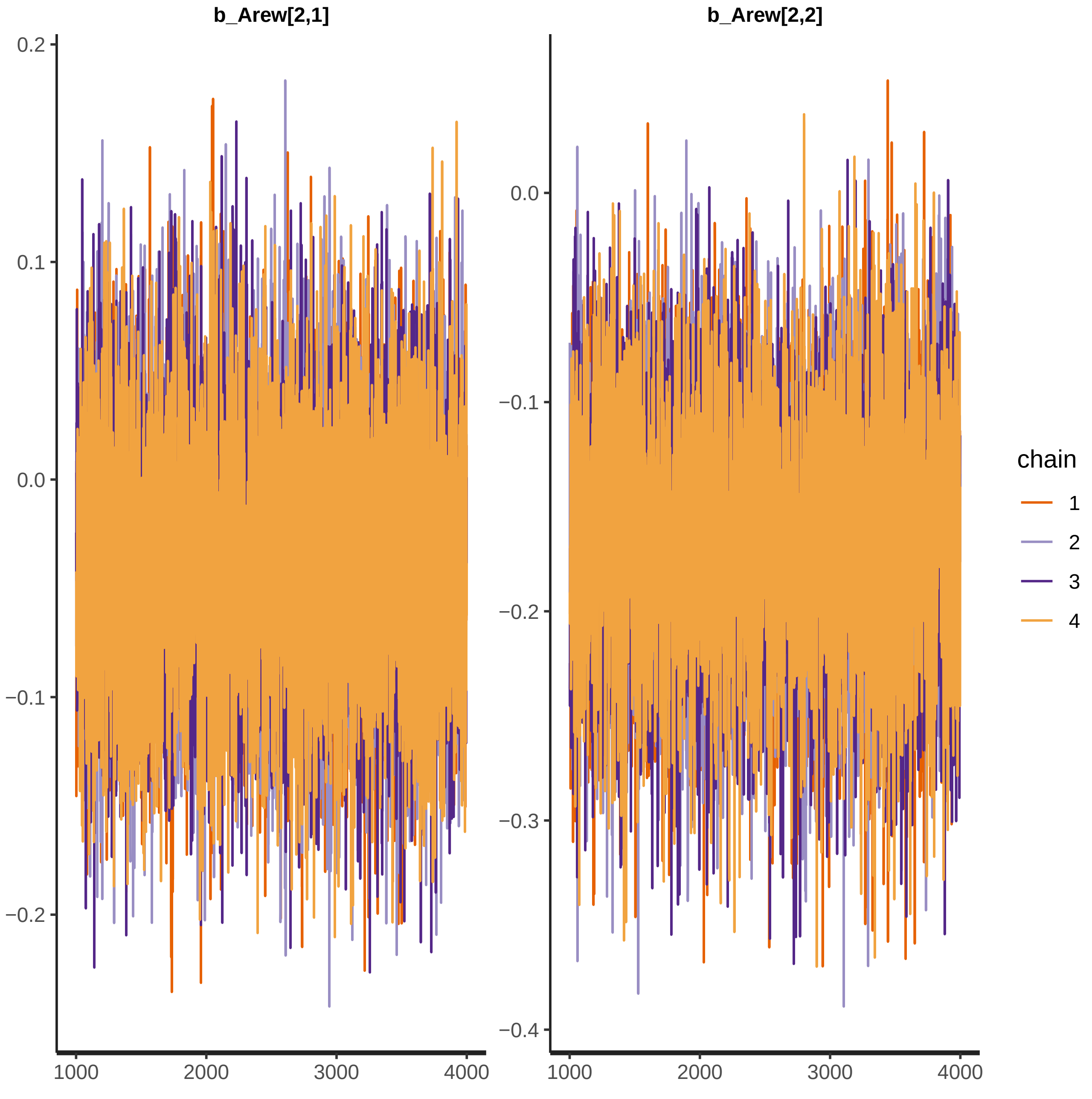

b_Arew[2,1]
b_Arew[2,2]
0.2
0.1
0.0
−0.1
−0.2
−0.3
−0.4
1000
2000
3000
4000
chain
1
2
3
4

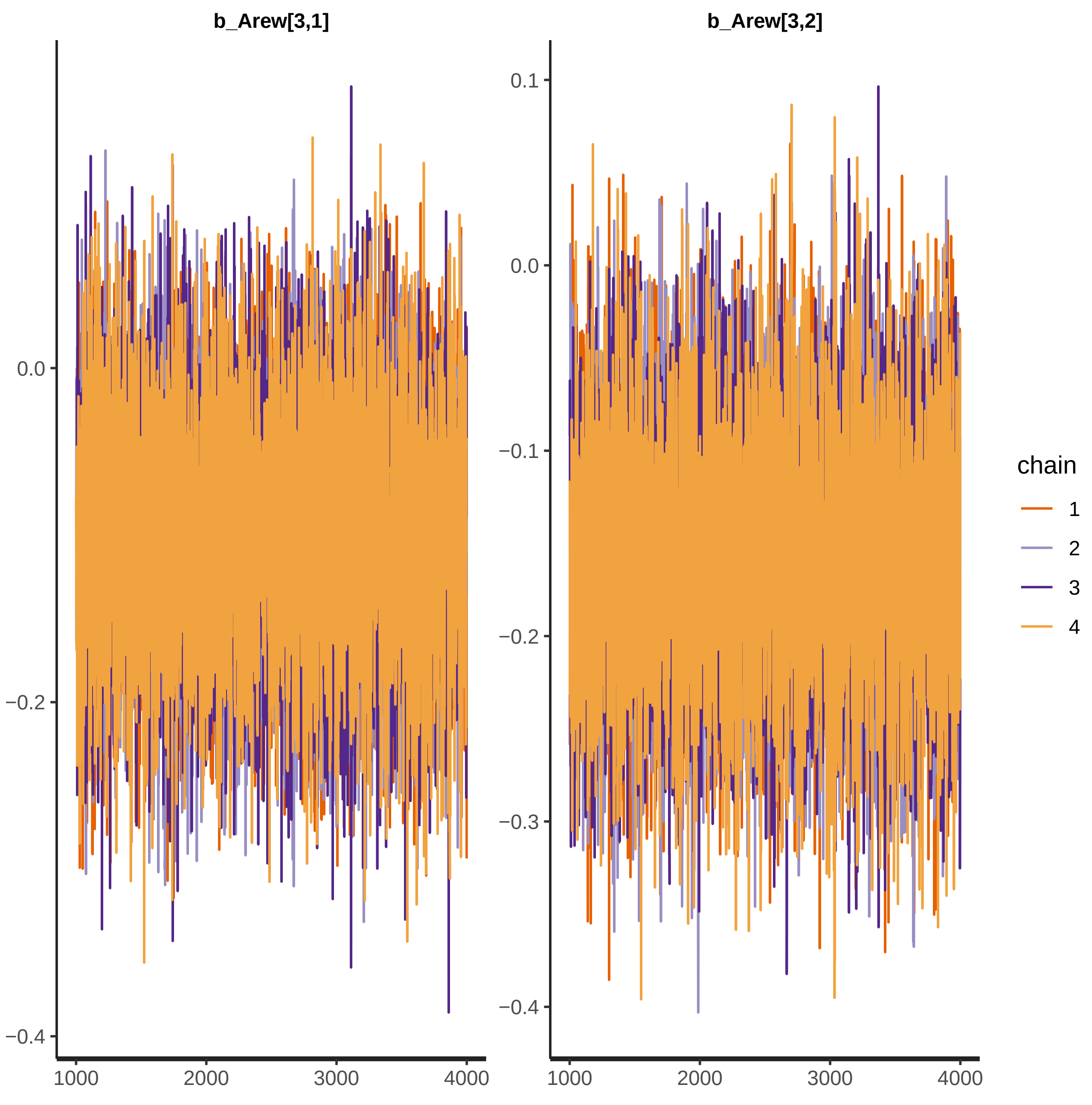

b_Arew[3,1]
b_Arew[3,2]
0.0
−0.2
−0.4
0.1
0.0
−0.1
−0.2
−0.3
−0.4
1000
2000
3000
4000
chain
1
2
3
4

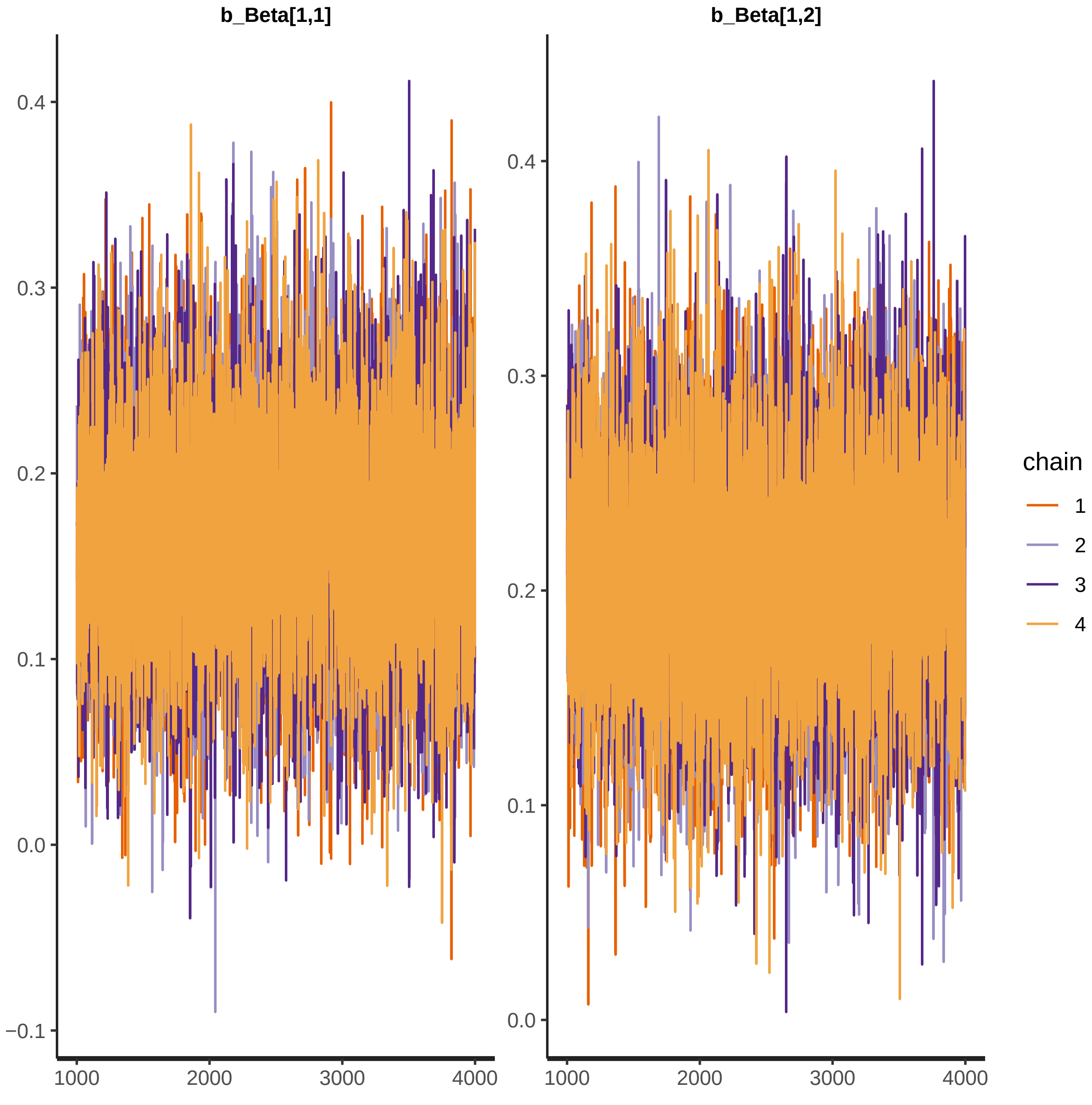
b_Beta[1,1]
b_Beta[1,2]
0.4
0.3
0.2
0.1
0.0
−0.1
1000
2000
3000
4000
chain
1
2
3
4

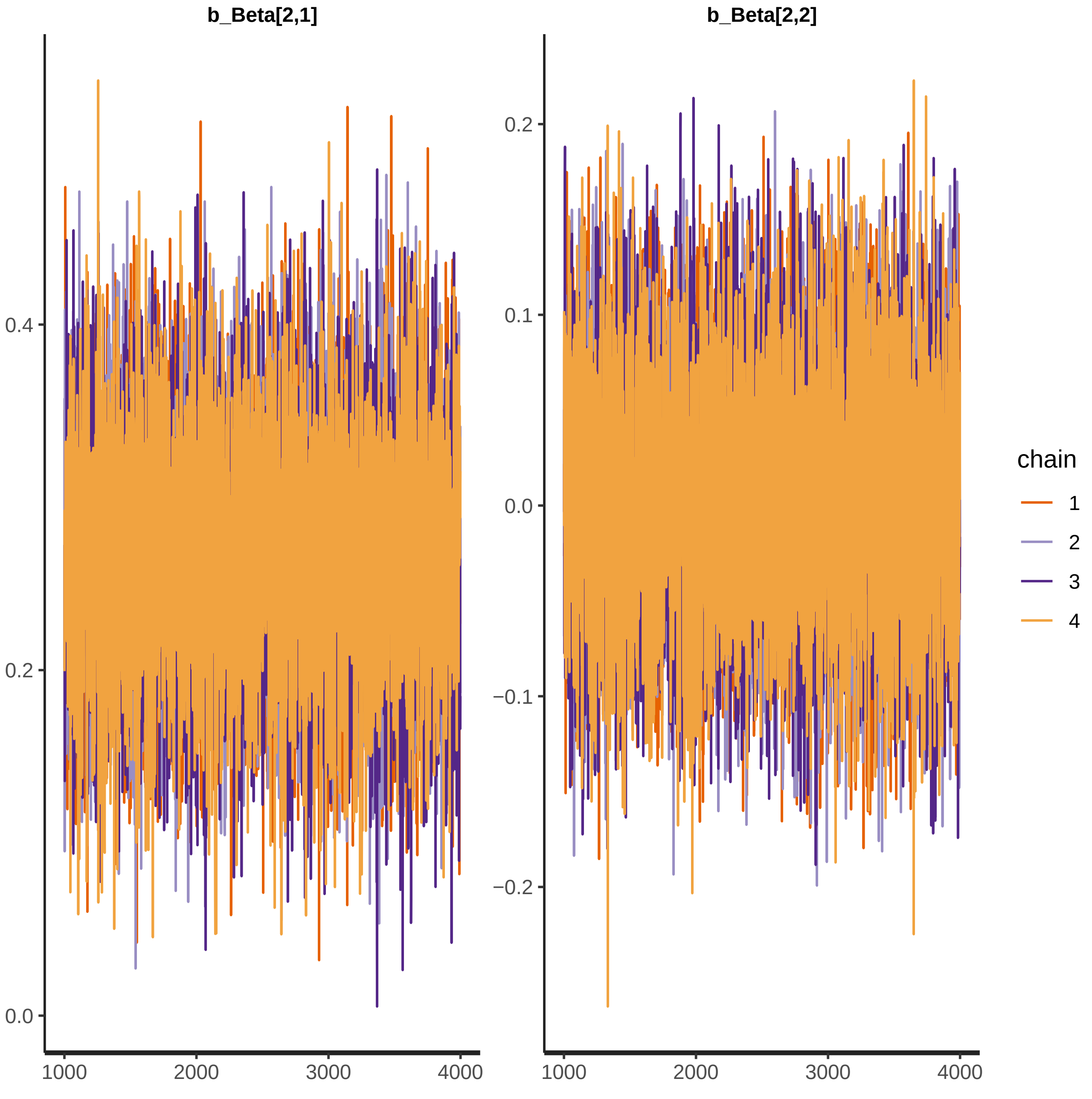
b_Beta[2,1]
b_Beta[2,2]
0.4
0.2
0.0
0.2
0.1
0.0
−0.1
−0.2
1000
2000
3000
4000
chain
1
2
3
4

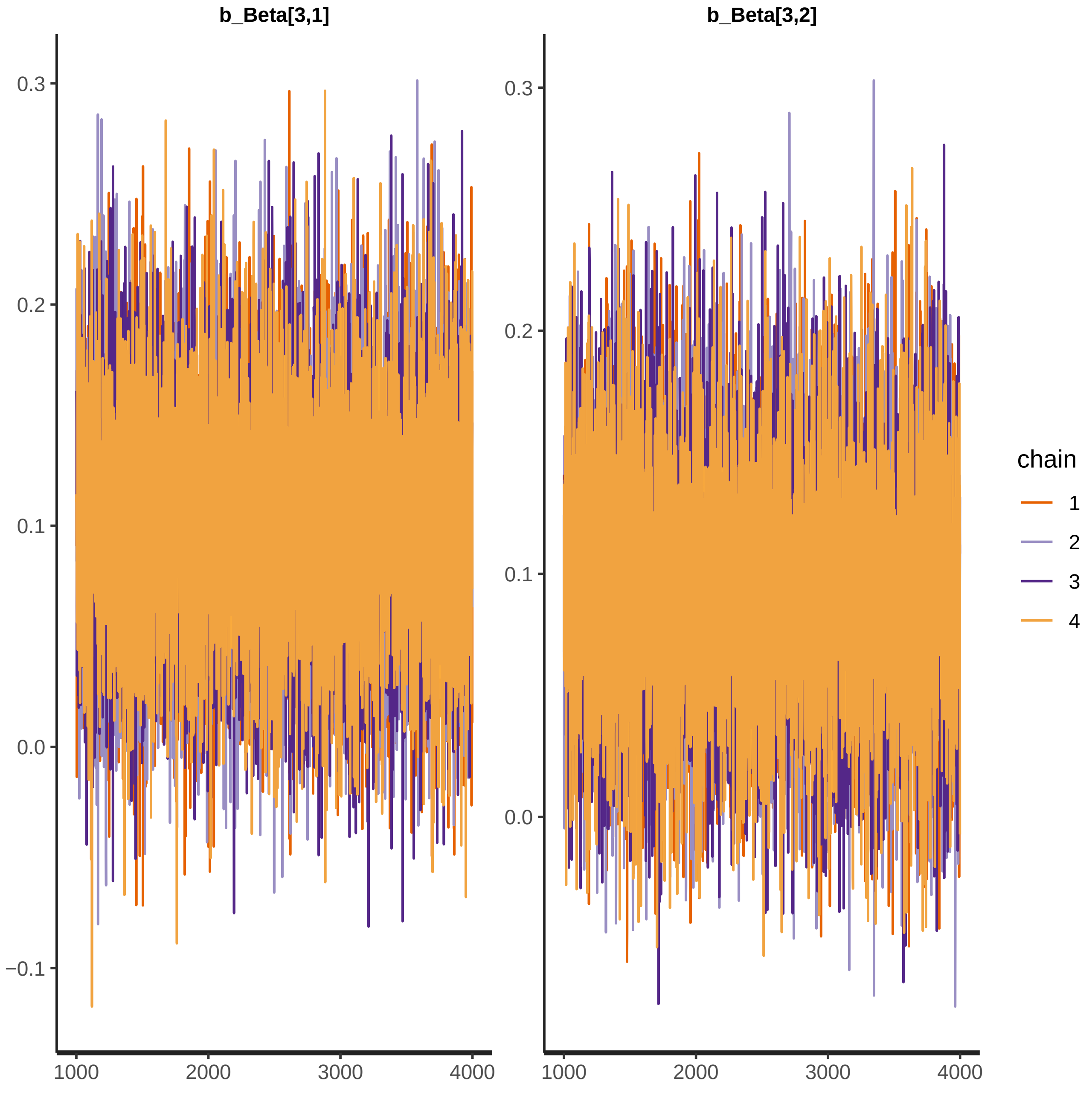
b_Beta[3,1]
b_Beta[3,2]
0.3
0.2
0.1
0.0
−0.1
1000
2000
3000
4000
chain
1
2
3
4

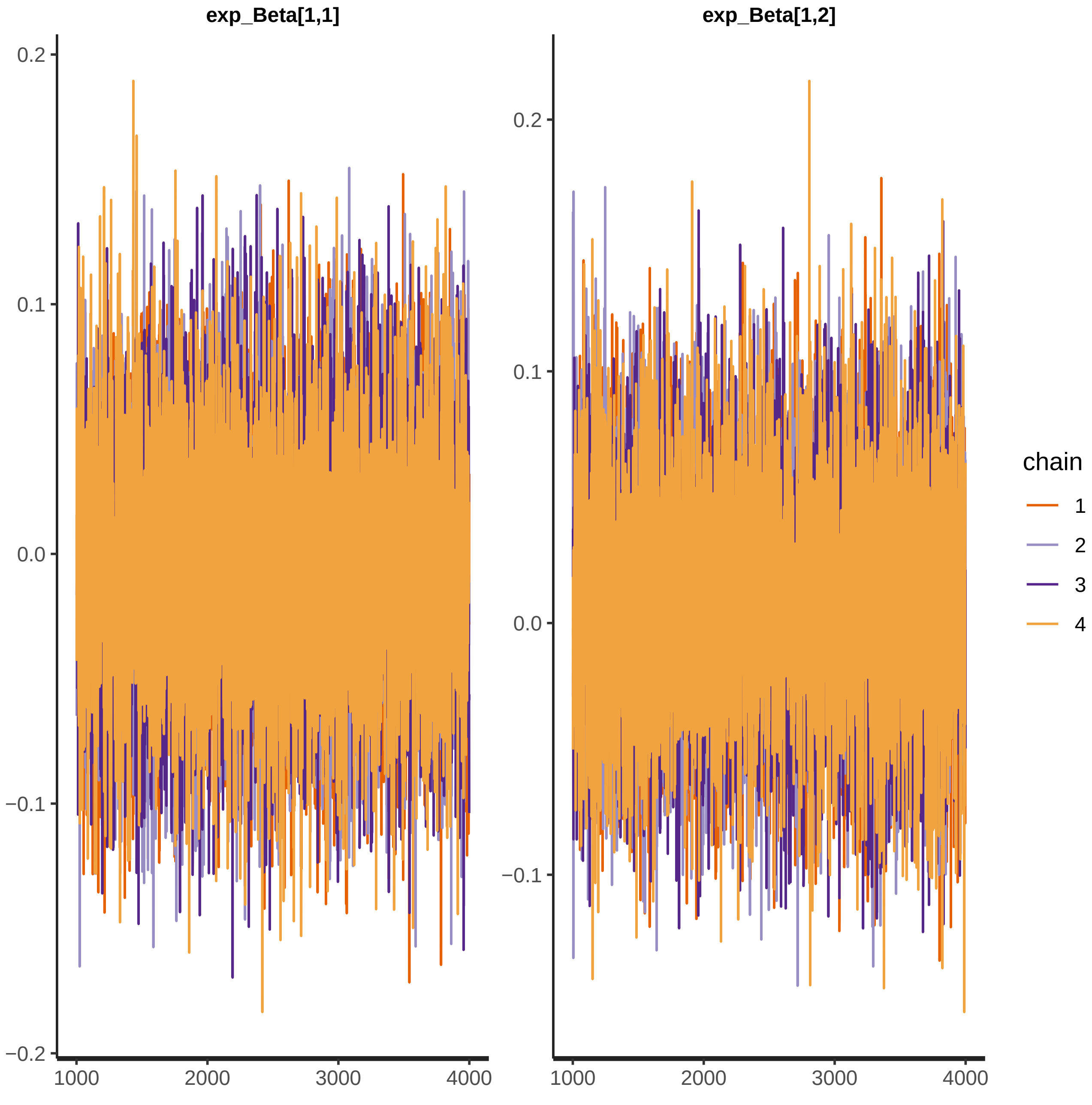

exp_Beta[1,1]
exp_Beta[1,2]
0.2
0.1
0.0
−0.1
−0.2
1000
2000
3000
4000
chain
1
2
3
4

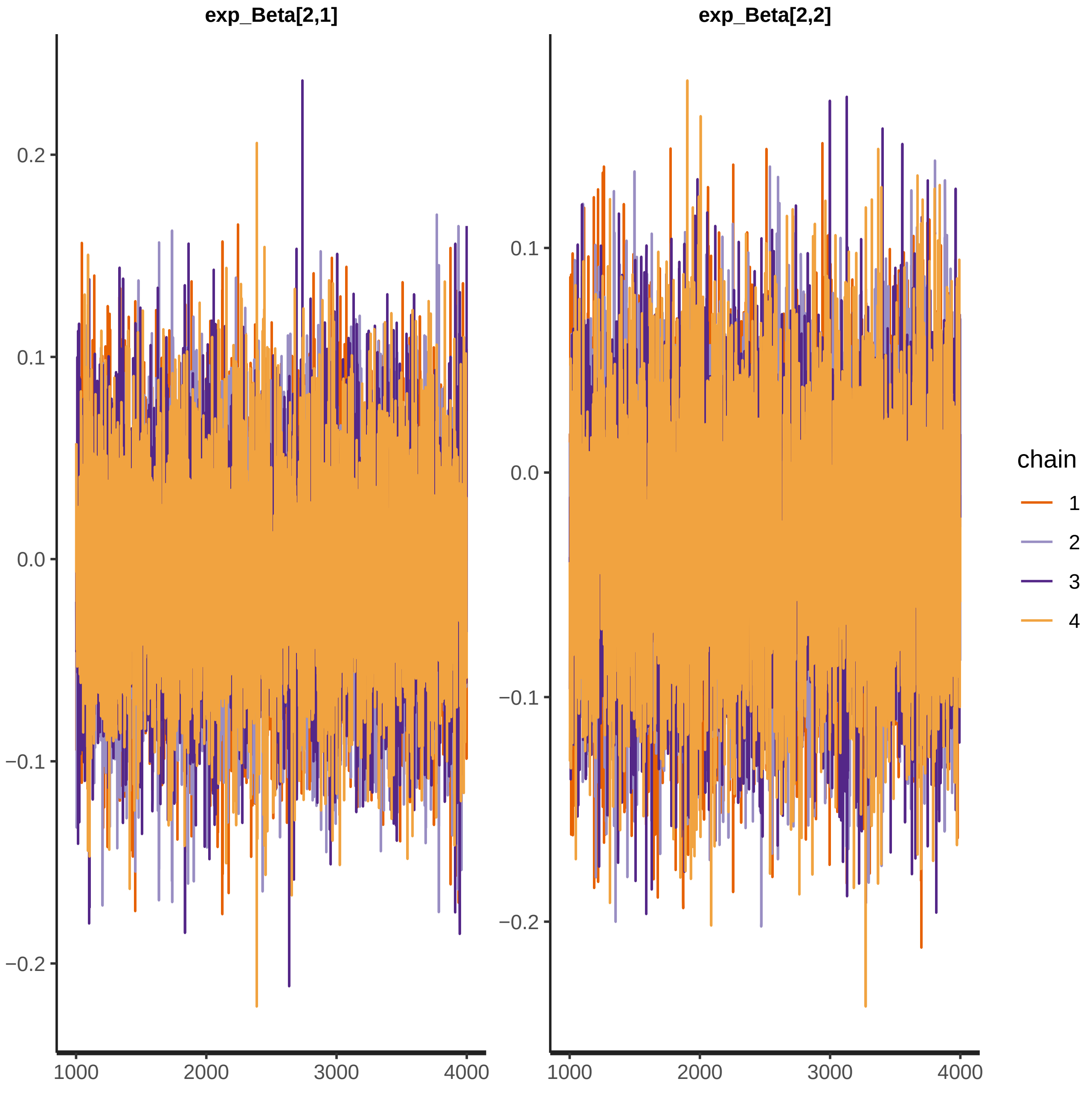
exp_Beta[2,1]
exp_Beta[2,2]
0.2
0.1
0.0
−0.1
−0.2
1000
2000
3000
4000
chain
1
2
3
4

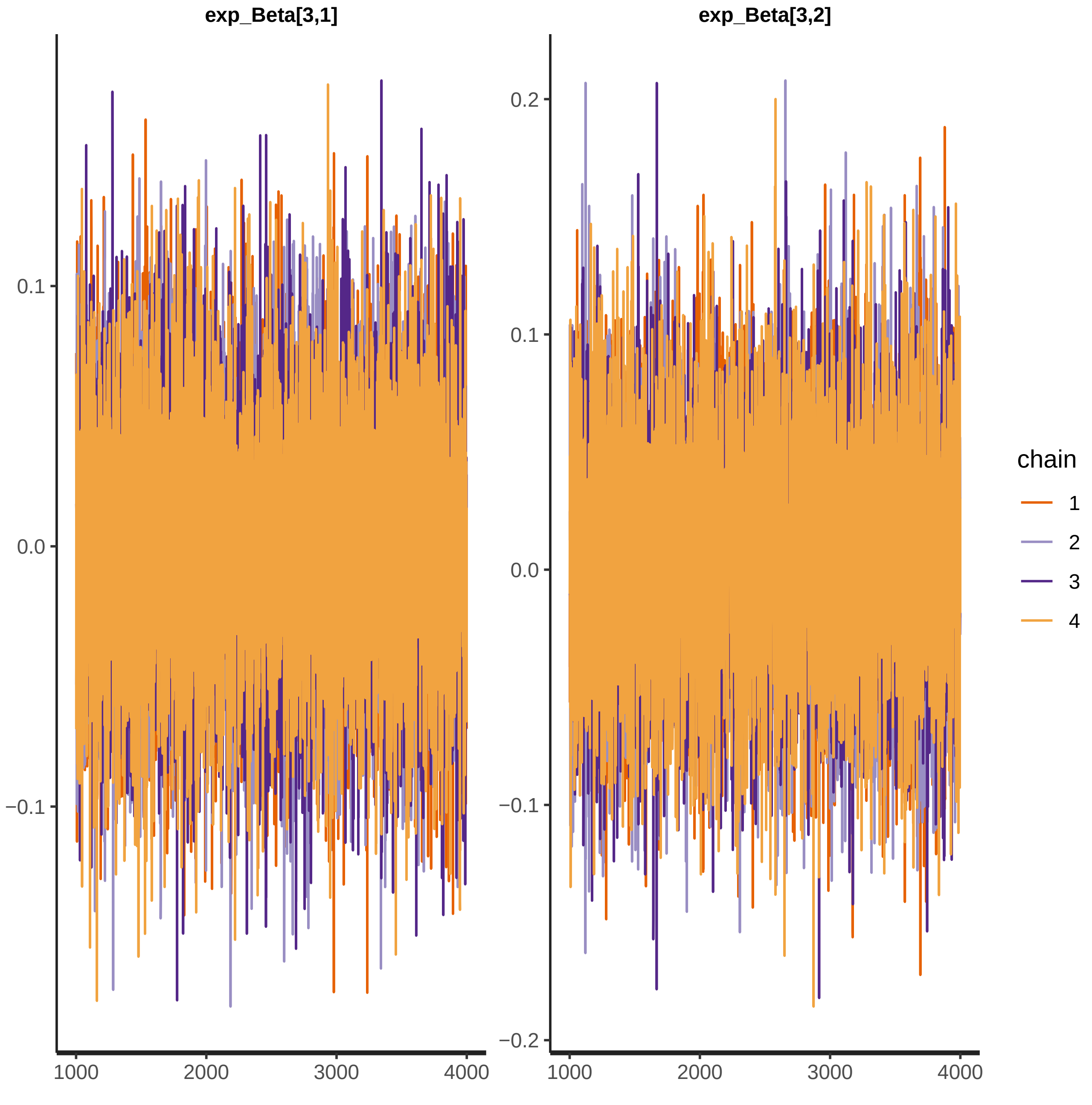

exp_Beta[3,1]
exp_Beta[3,2]
0.1
0.0
−0.1
0.2
0.1
0.0
−0.1
−0.2
1000
2000
3000
4000
1000
2000
3000
4000
chain
1
2
3
4

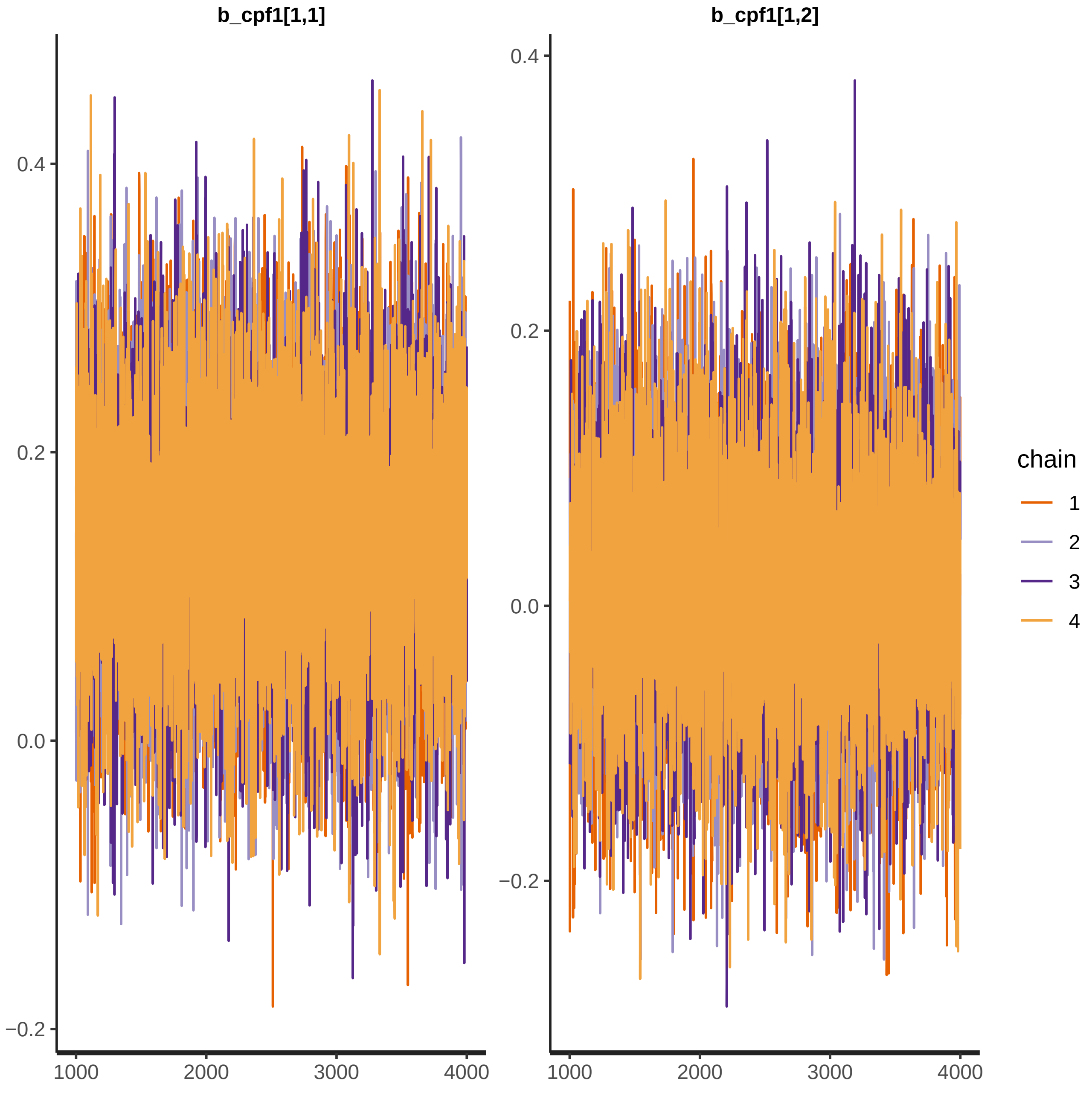

b_cpf1[1,1]
b_cpf1[1,2]
0.4
0.2
0.0
−0.2
1000
2000
3000
4000
chain
1
2
3
4

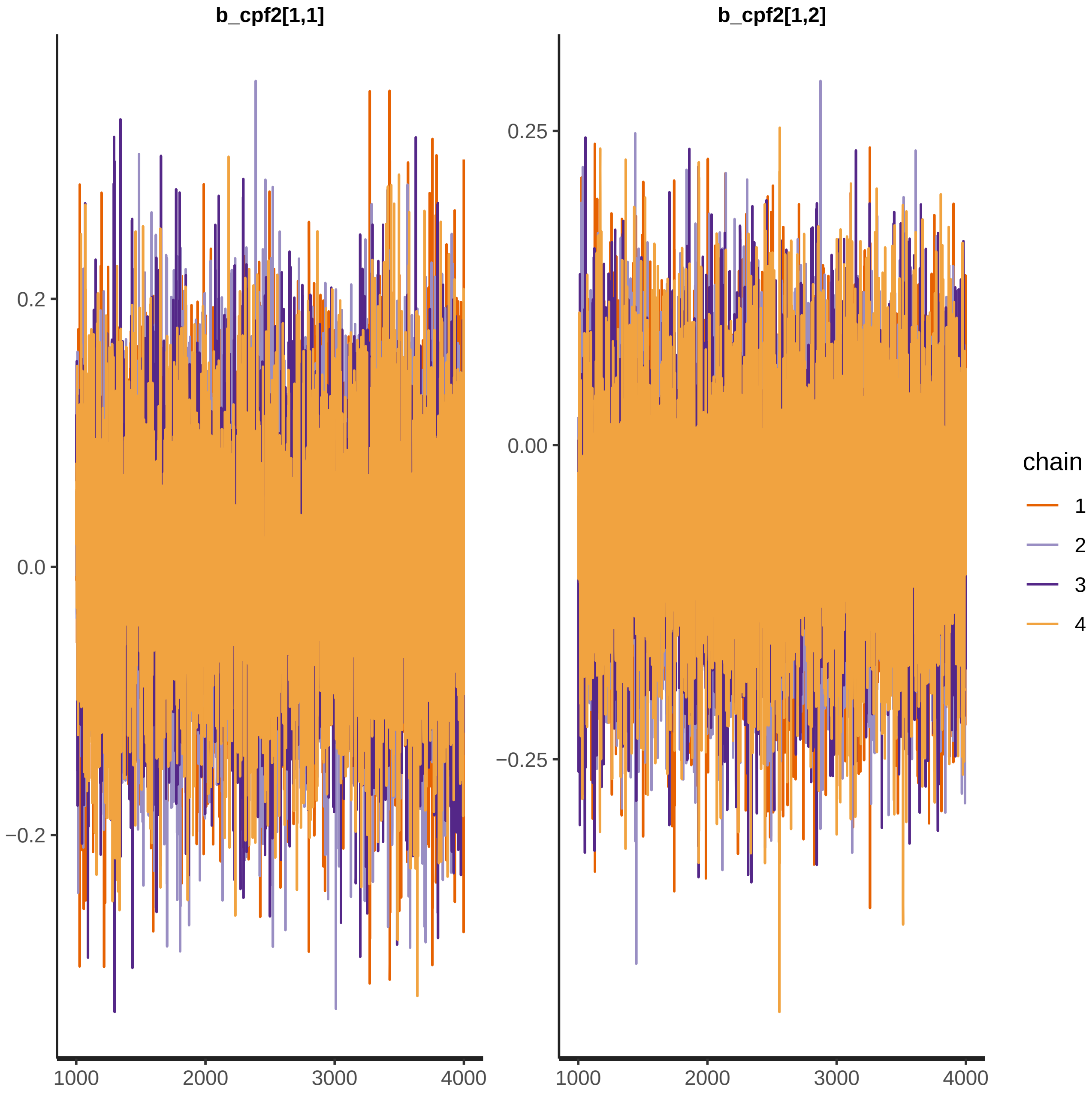
b_cpf2[1,1]
b_cpf2[1,2]
0.2
0.0
−0.2
0.25
0.00
−0.25
1000
2000
3000
4000
chain
1
2
3
4

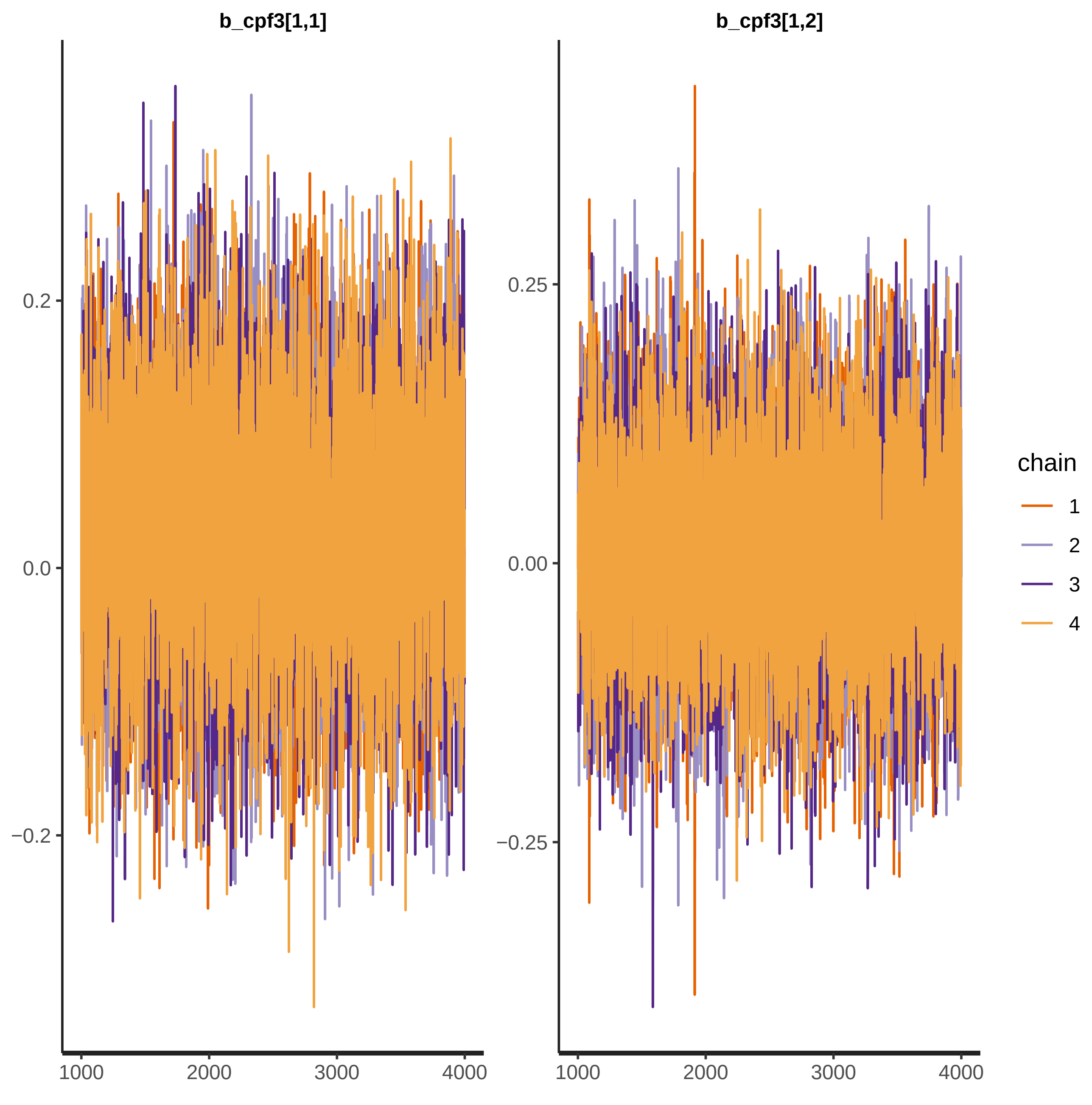
b_cpf3[1,1]
b_cpf3[1,2]
0.2
0.0
−0.2
0.25
0.00
−0.25
1000
2000
3000
4000
chain
1
2
3
4

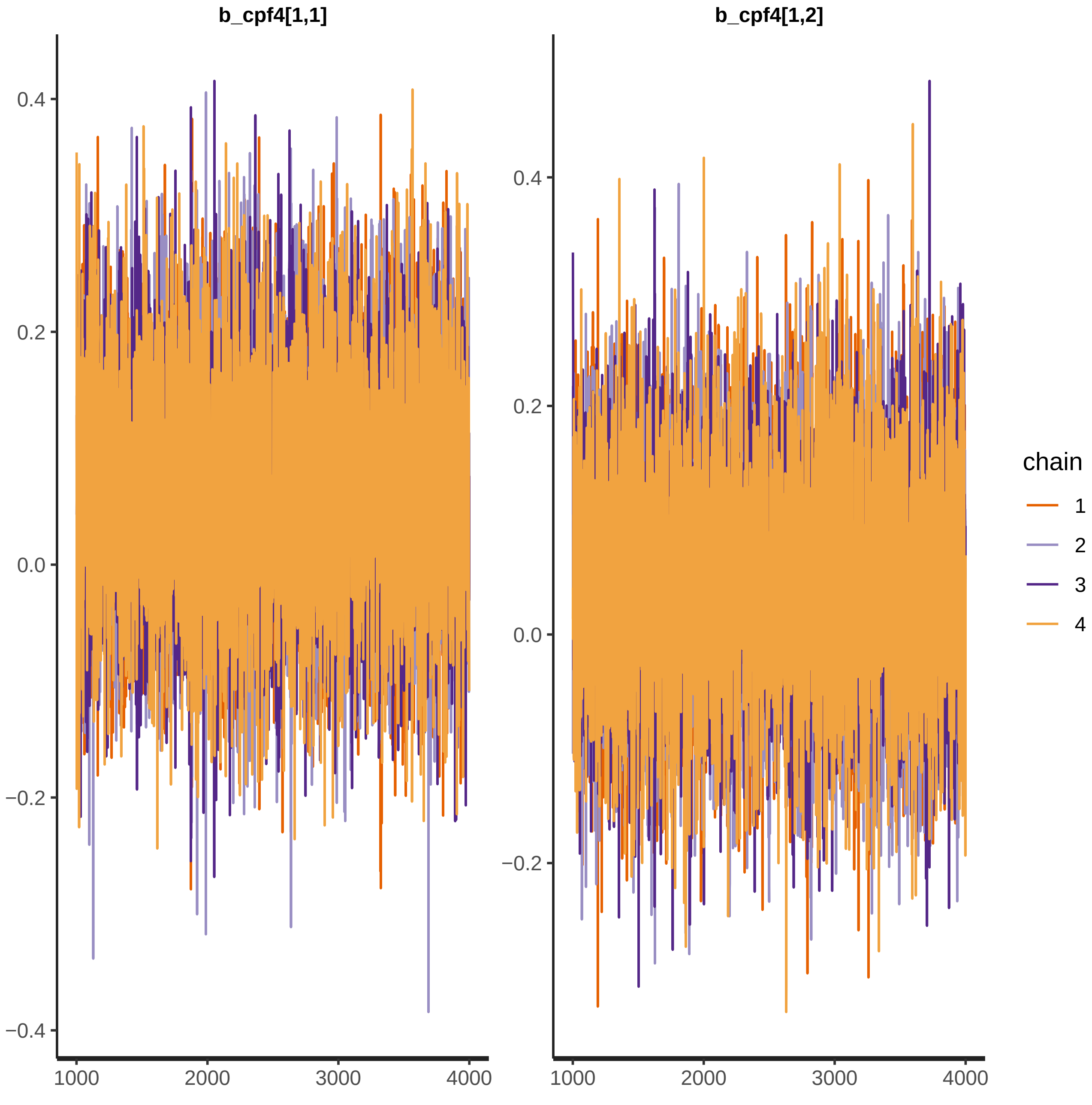

b_cpf4[1,1]
b_cpf4[1,2]
0.4
0.2
0.0
−0.2
−0.4
0.4
0.2
0.0
−0.2
1000
2000
3000
4000
1000
2000
3000
4000
chain
1
2
3
4

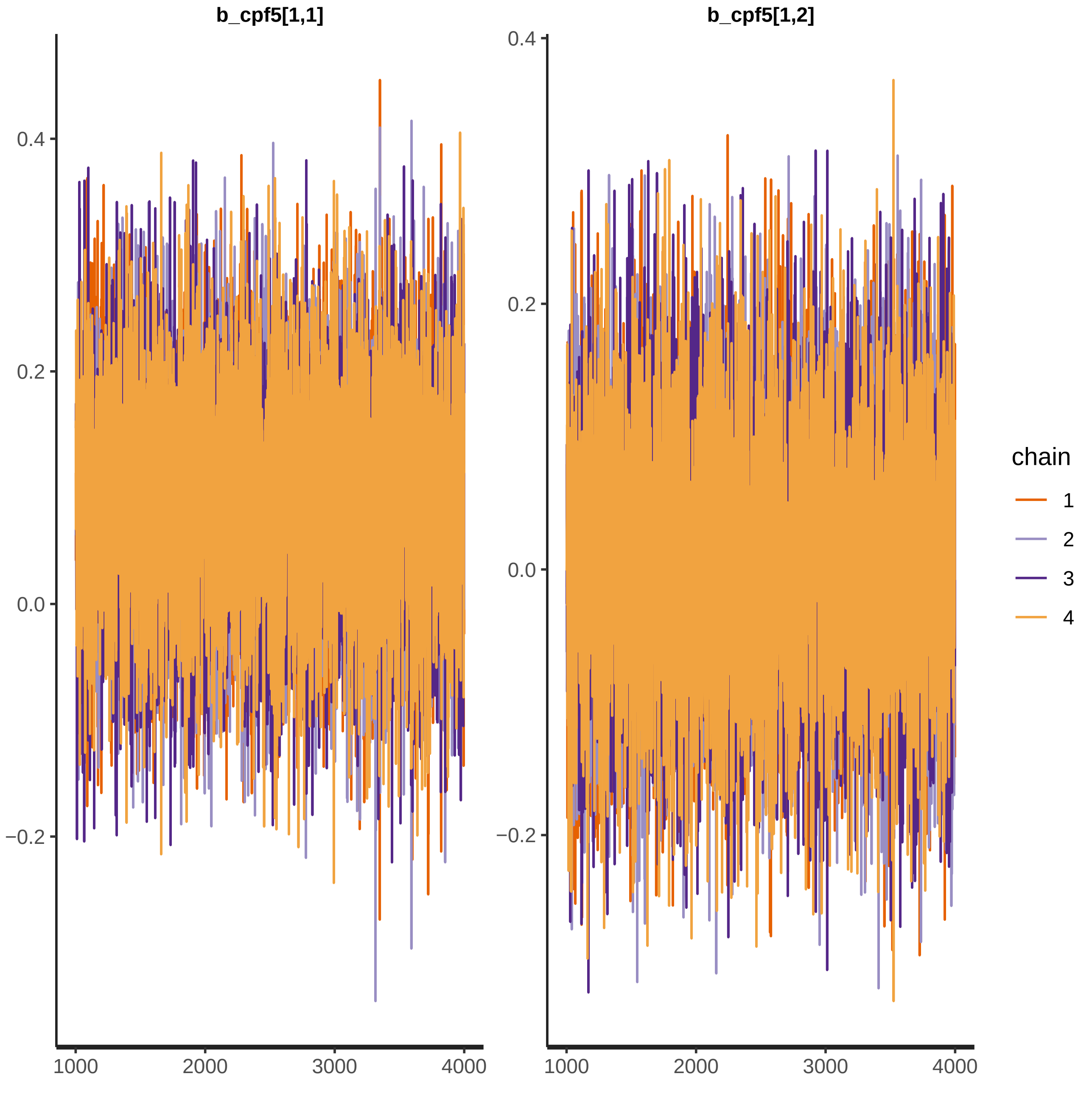
b_cpf5[1,1]
b_cpf5[1,2]
0.4
0.2
0.0
−0.2
1000
2000
3000
4000
chain
1
2
3
4

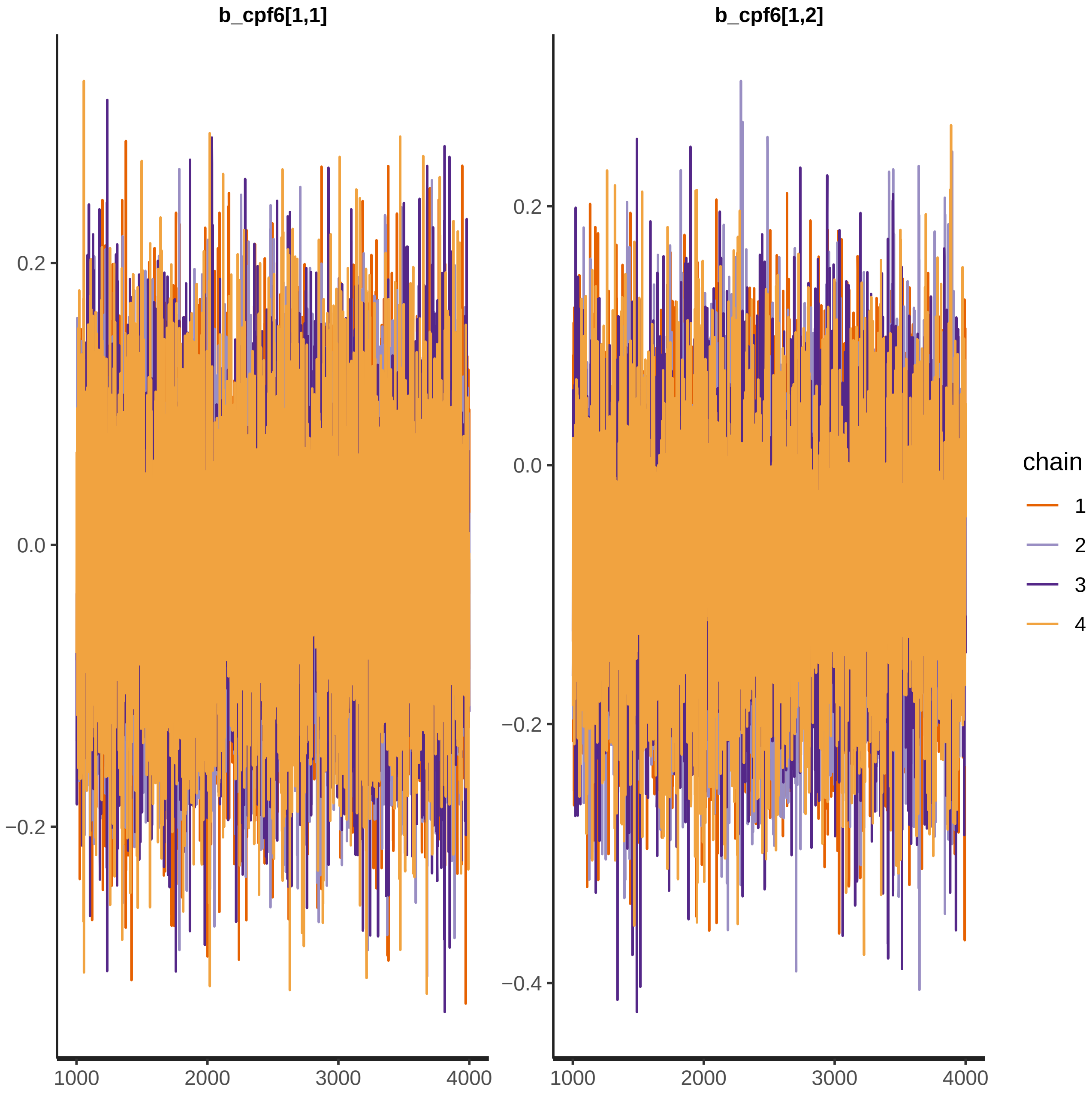
b_cpf6[1,1]
b_cpf6[1,2]
0.2
0.0
−0.2
−0.4
1000
2000
3000
4000
chain
1
2
3
4

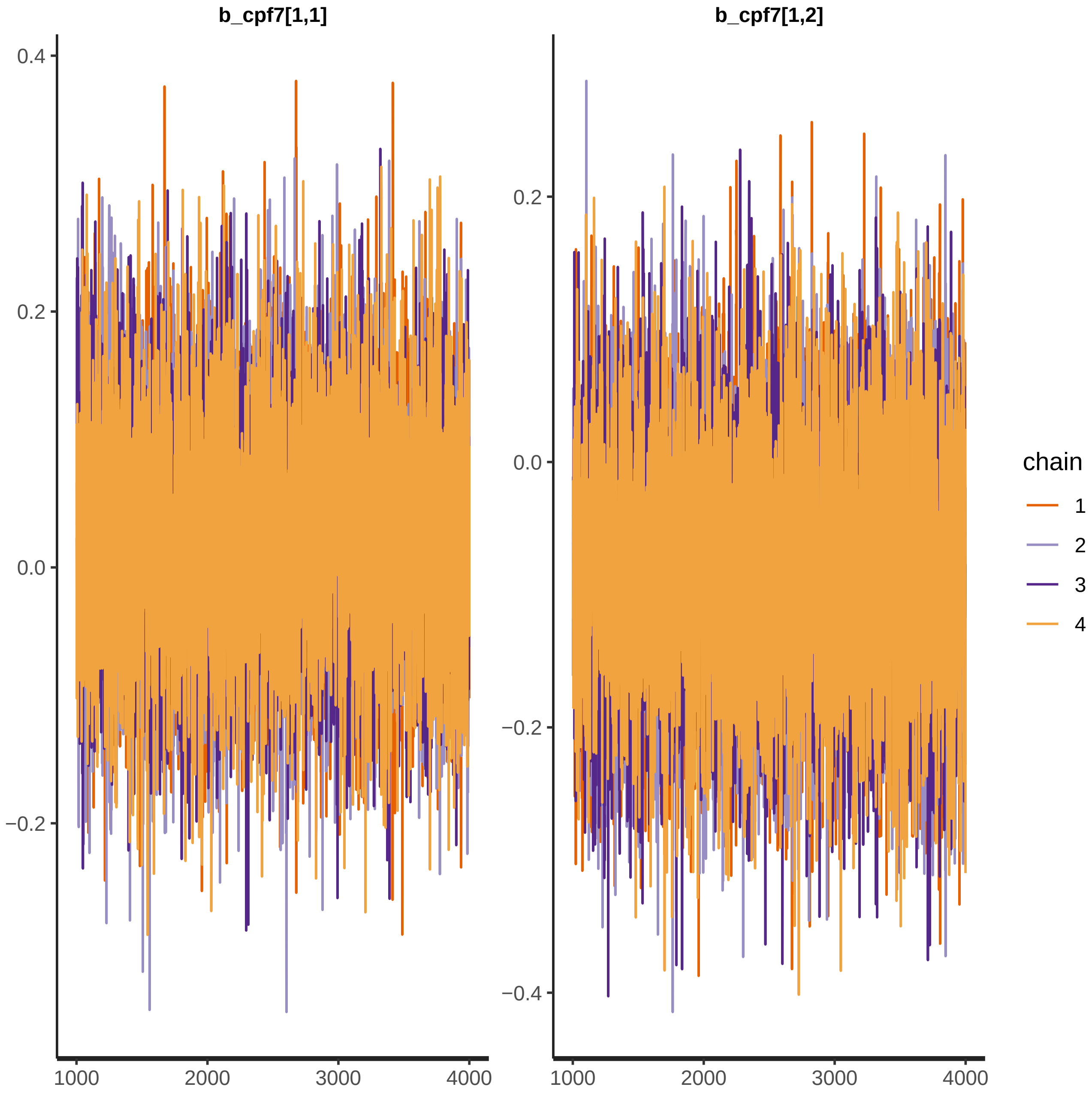

b_cpf7[1,1]
b_cpf7[1,2]
0.4
0.2
0.0
−0.2
−0.4
1000
2000
3000
4000
chain
1
2
3
4

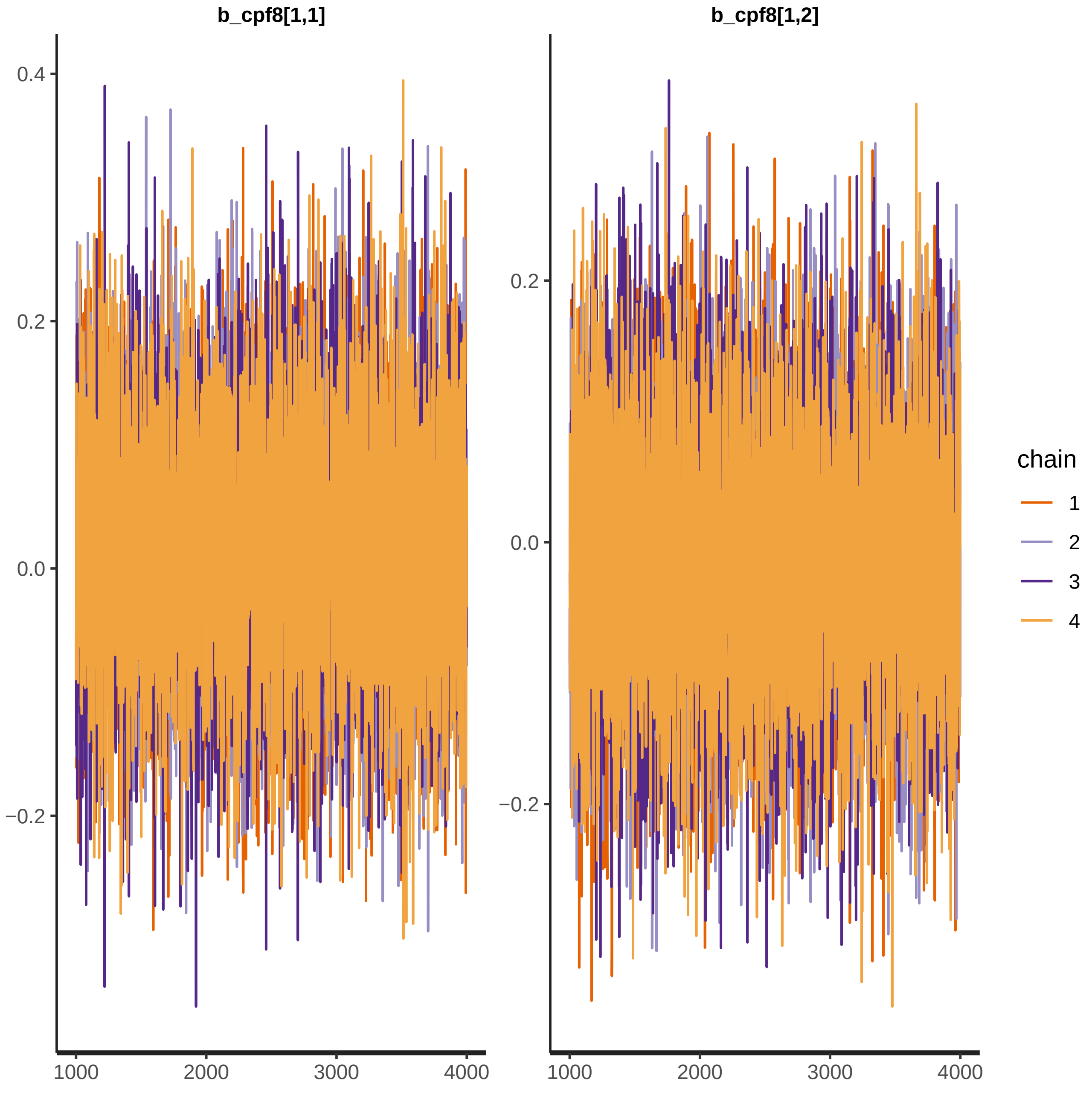
b_cpf8[1,1]
b_cpf8[1,2]
0.4
0.2
0.0
−0.2
1000
2000
3000
4000
chain
1
2
3
4

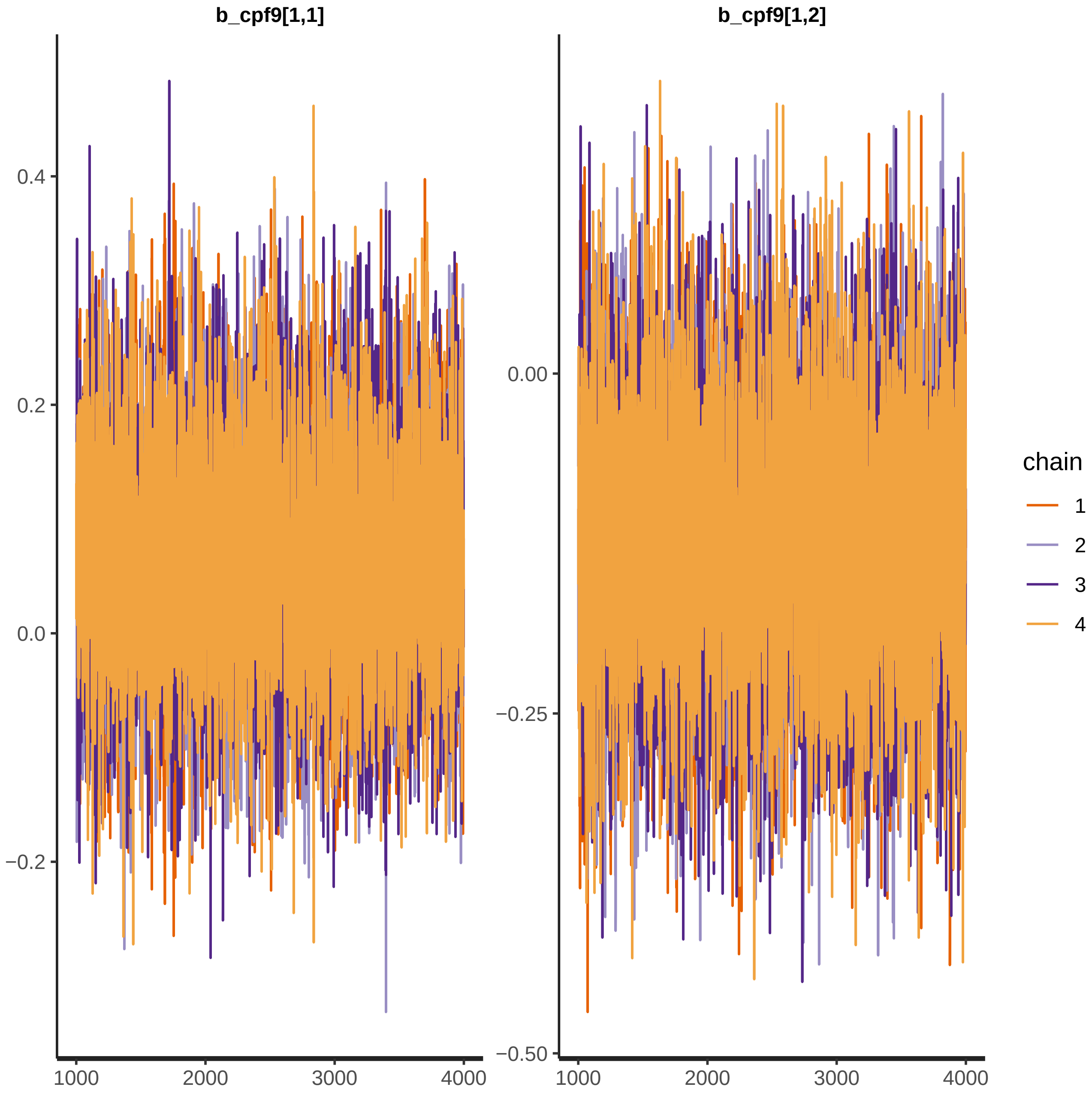

b_cpf9[1,1]
b_cpf9[1,2]
0.4
0.2
0.0
−0.2
0.00
−0.25
−0.50
1000
2000
3000
4000
chain
1
2
3
4

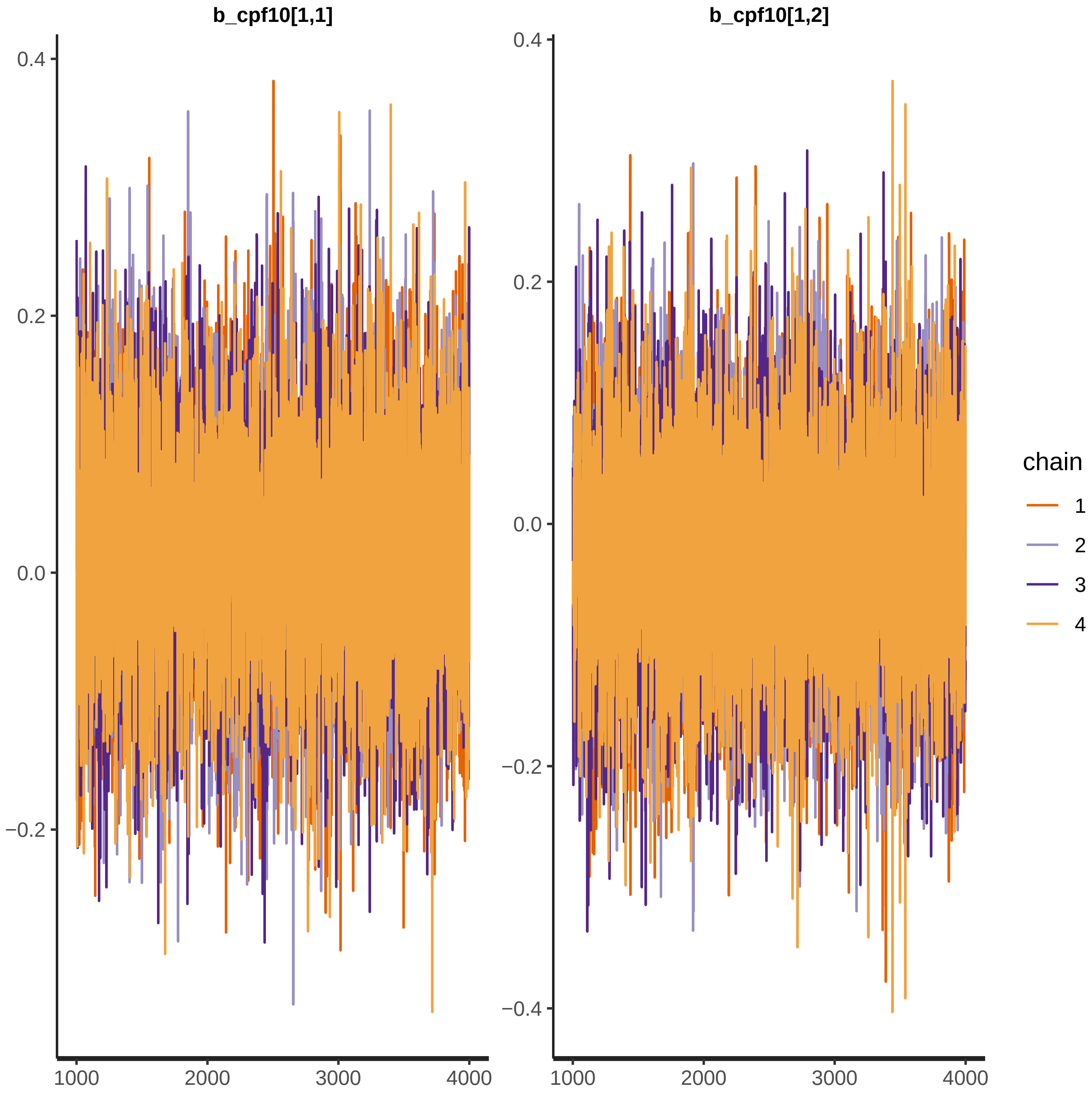

b_cpf10[1,1]
b_cpf10[1,2]
0.4
0.2
0.0
−0.2
−0.4
1000
2000
3000
4000
chain
1
2
3
4

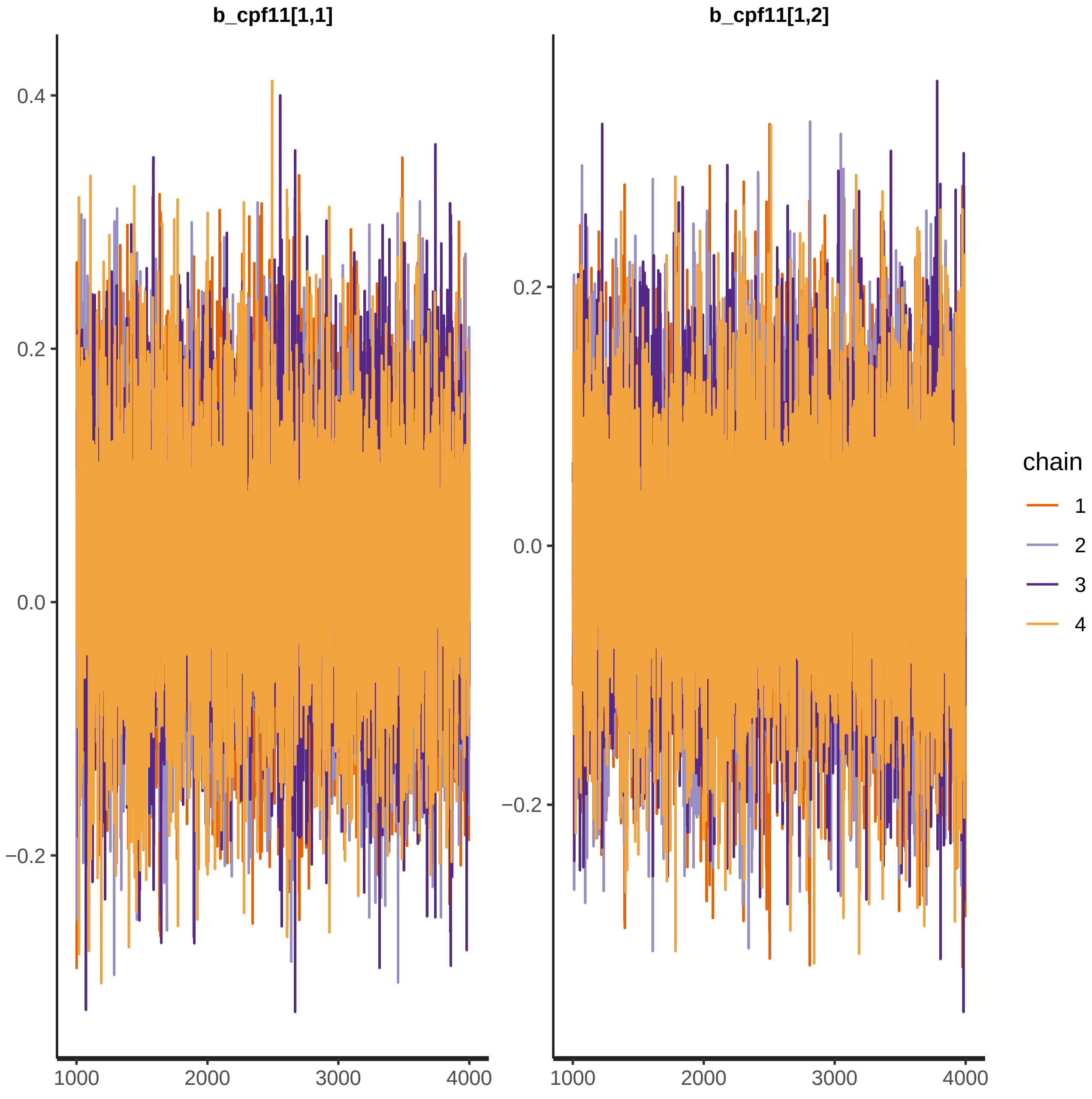
b_cpf11[1,1]
b_cpf11[1,2]
0.4
0.2
0.0
−0.2
1000
2000
3000
4000
chain
1
2
3
4

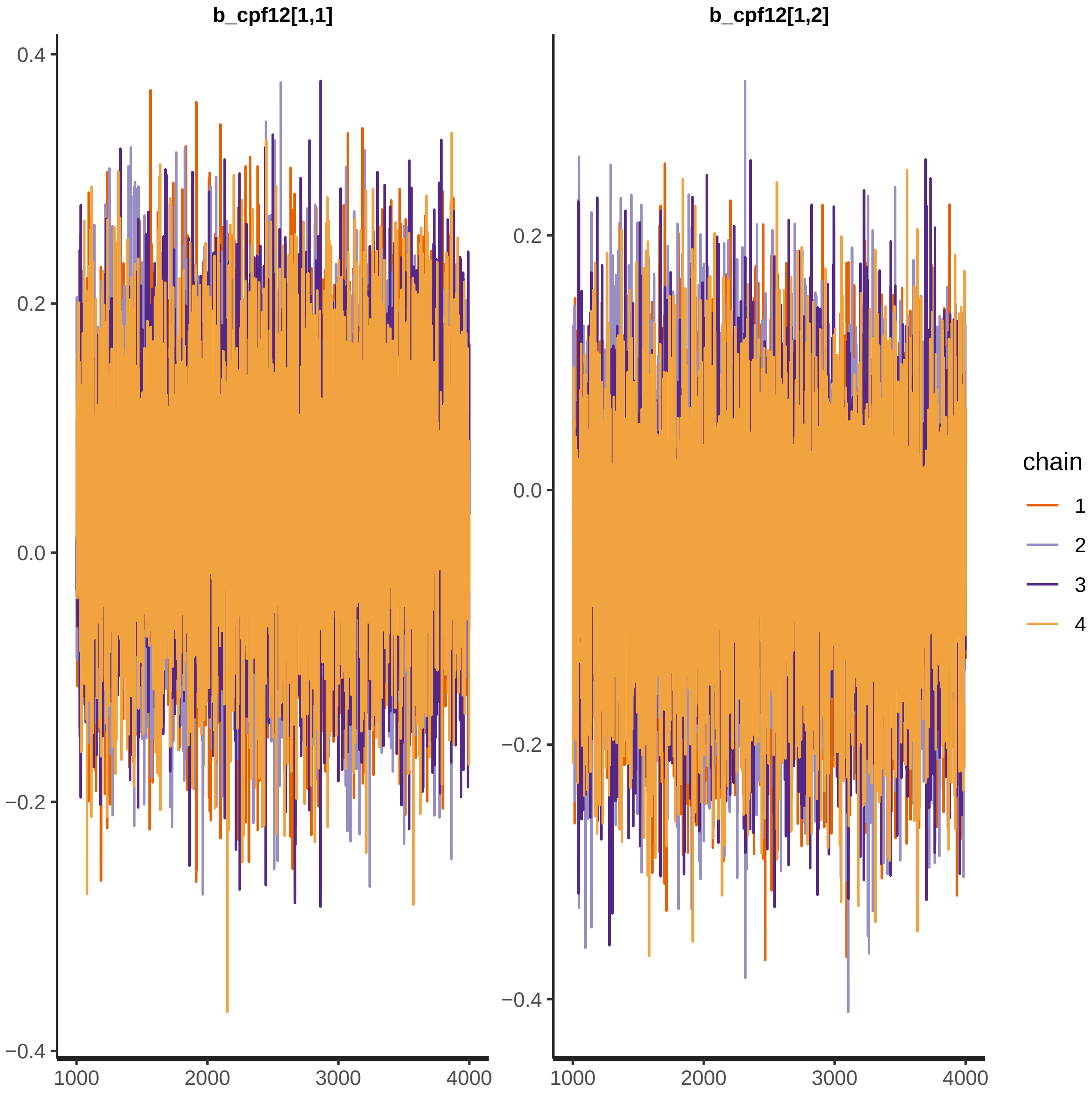
b_cpf12[1,1]
b_cpf12[1,2]
0.4
0.2
0.0
−0.2
−0.4
1000
2000
3000
4000
chain
1
2
3
4

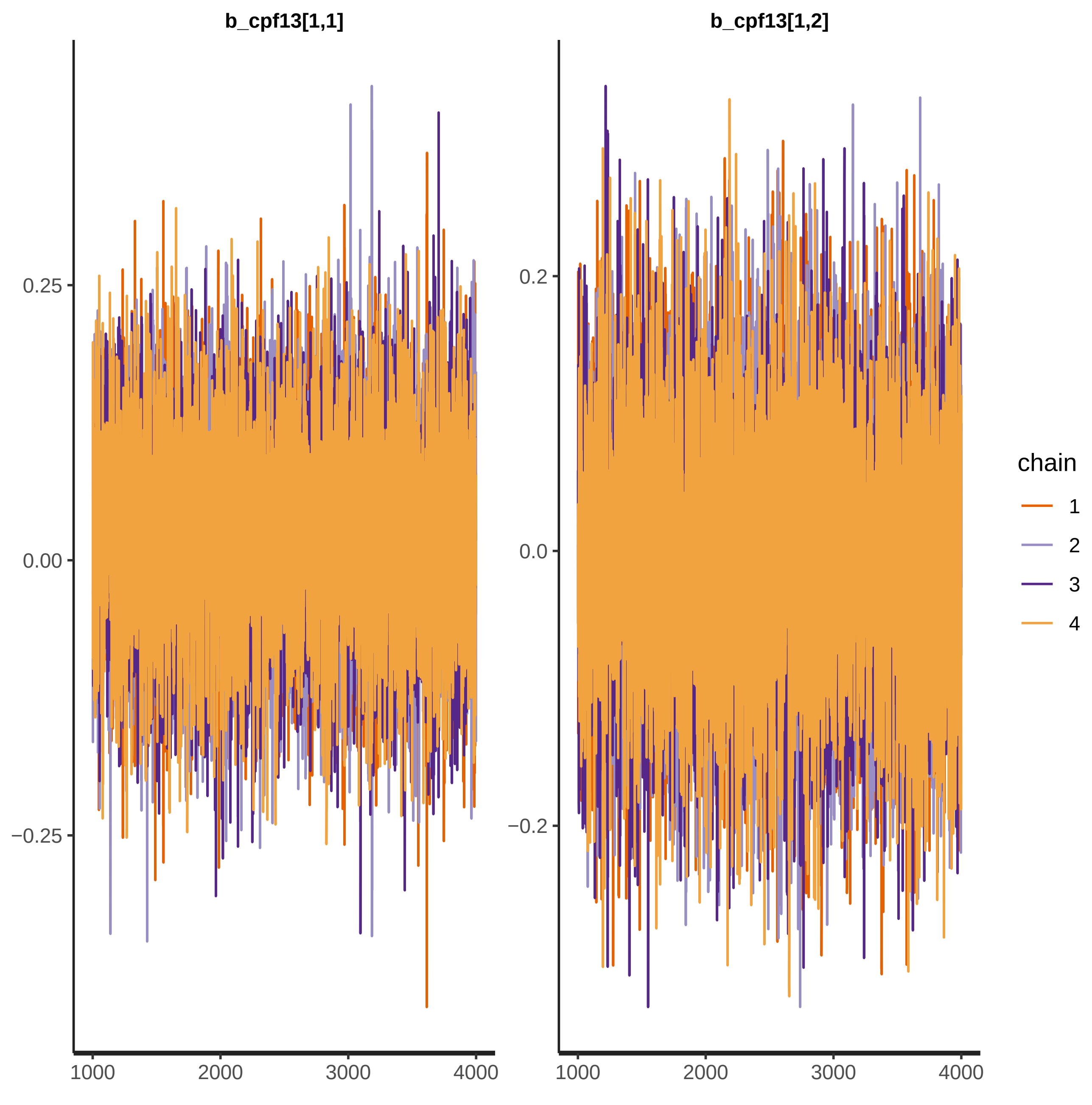

b_cpf13[1,1]
b_cpf13[1,2]
0.25
0.00
−0.25
0.2
0.0
−0.2
1000
2000
3000
4000
chain
1
2
3
4

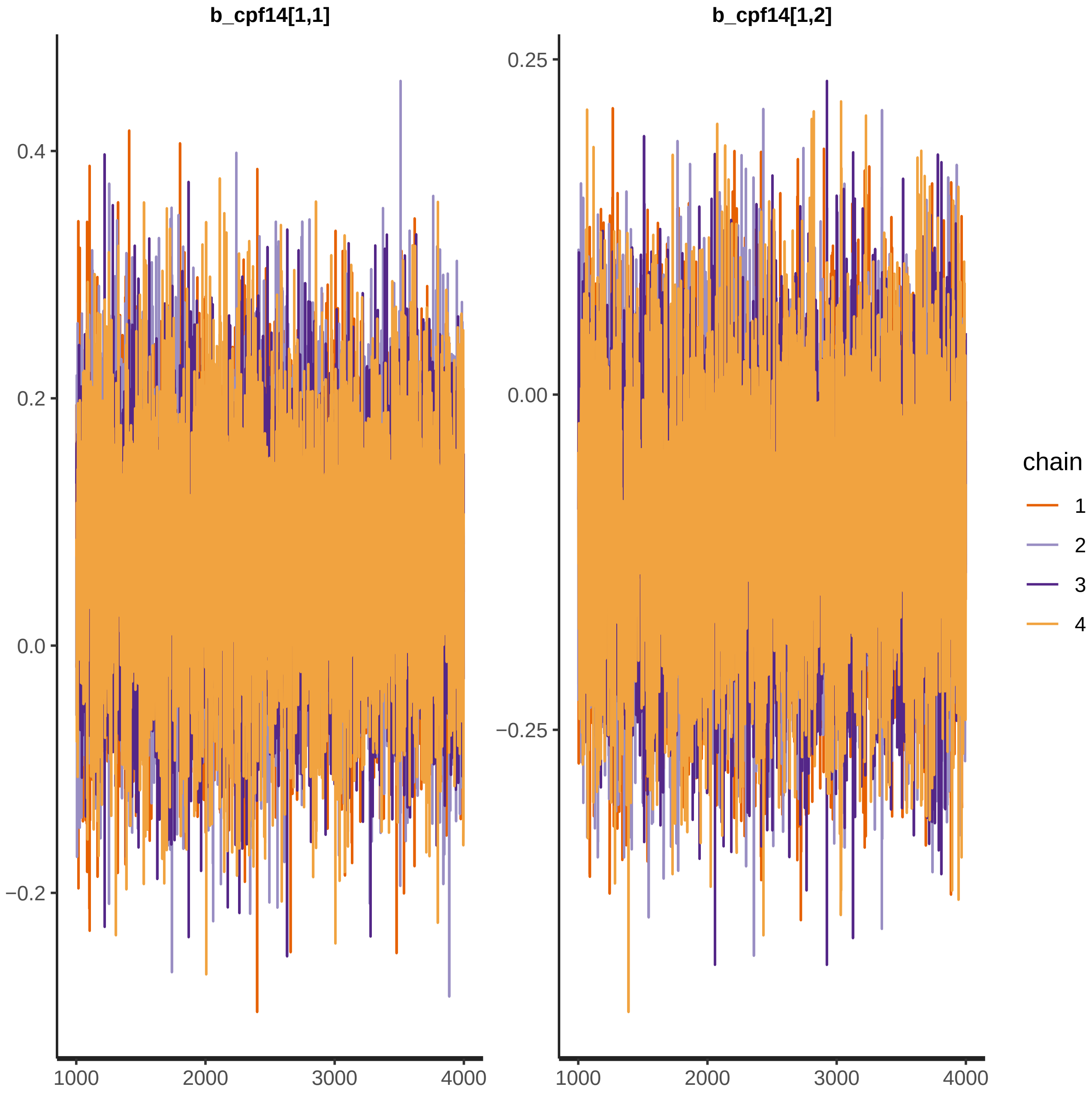
b_cpf14[1,1]
b_cpf14[1,2]
0.4
0.2
0.0
−0.2
0.25
0.00
−0.25
1000
2000
3000
4000
chain
1
2
3
4

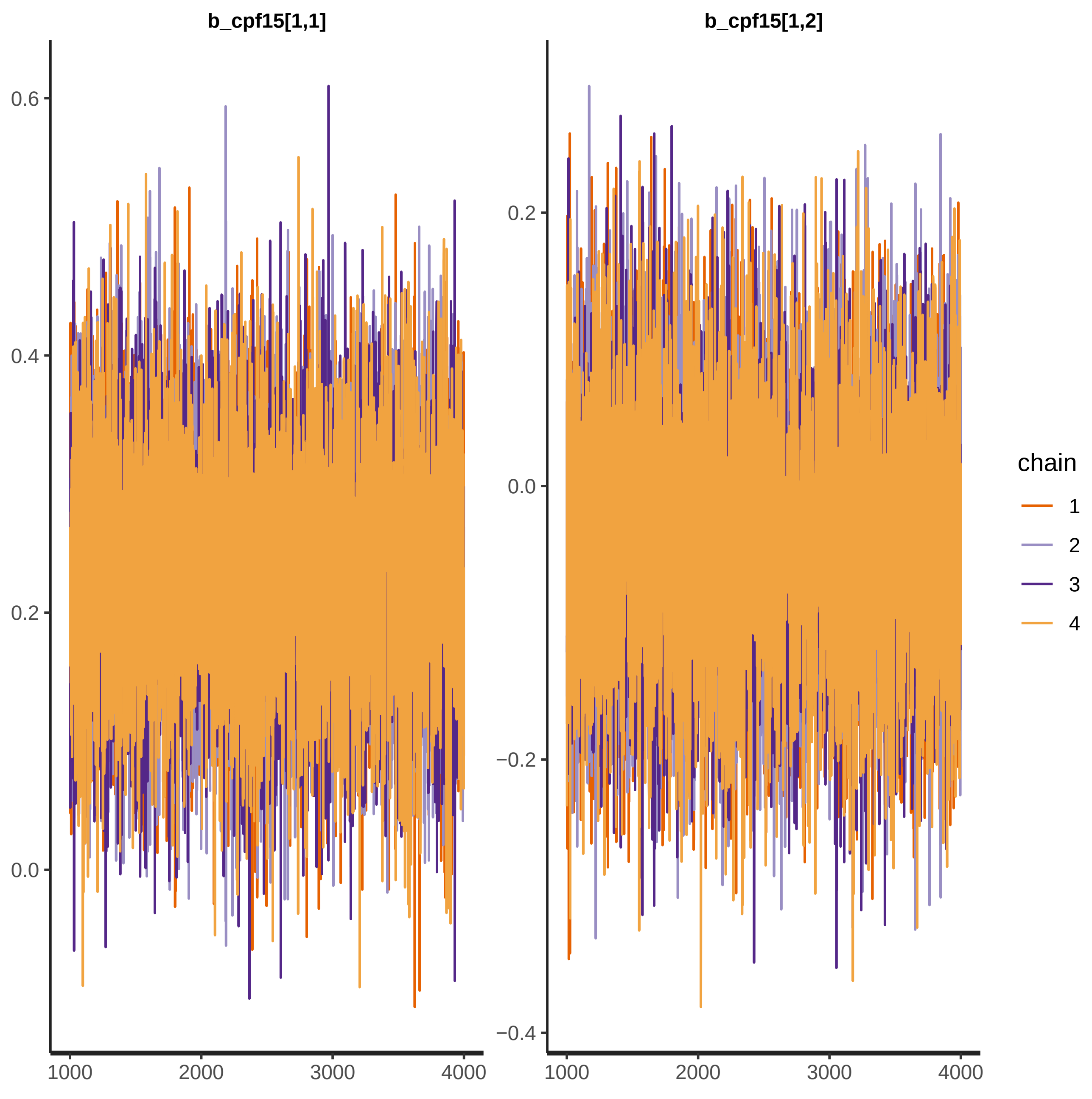

b_cpf15[1,1]
b_cpf15[1,2]
0.6
0.4
0.2
0.0
0.2
0.0
−0.2
−0.4
1000
2000
3000
4000
chain
1
2
3
4

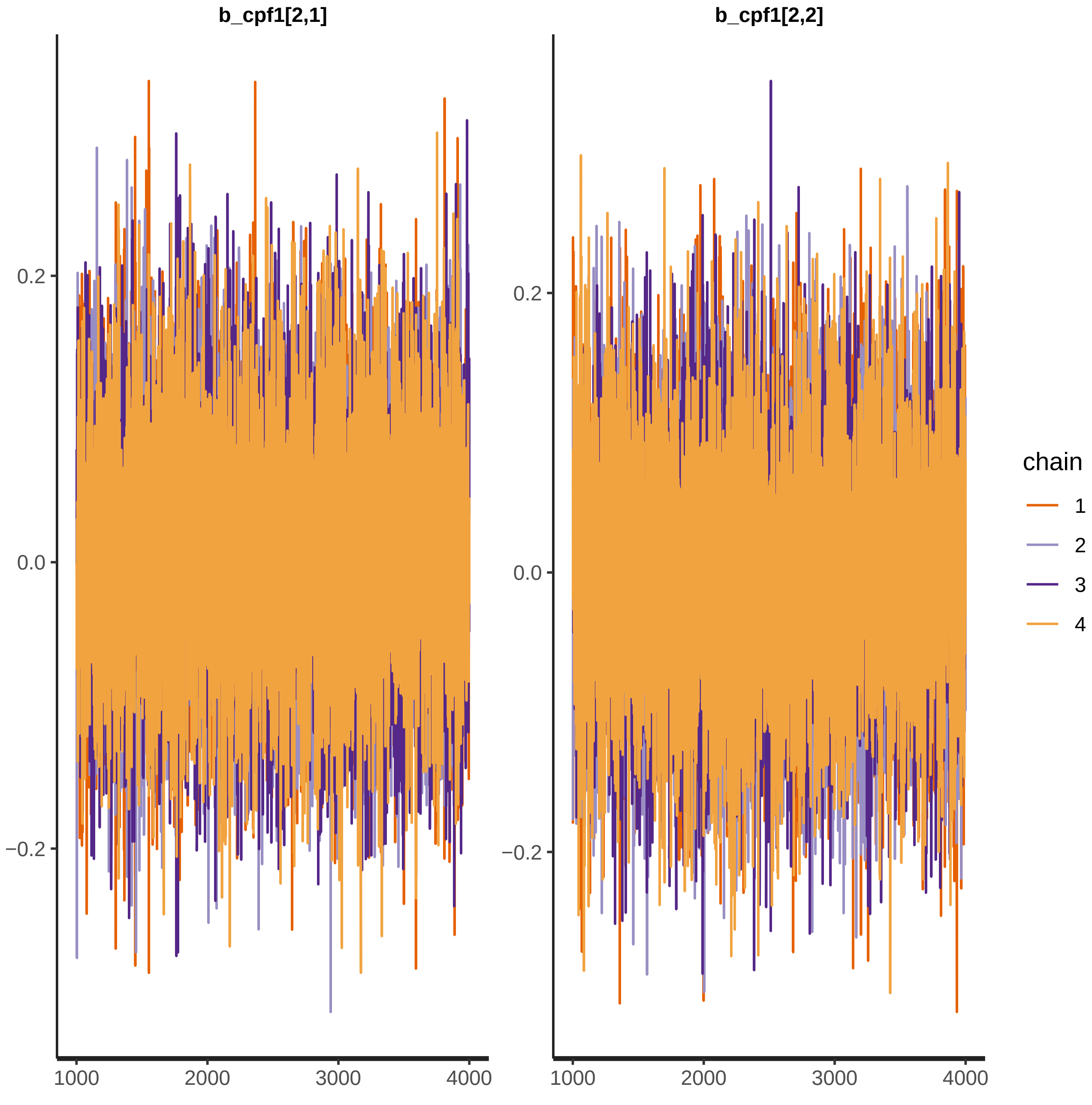

b_cpf1[2,1]
b_cpf1[2,2]
0.2
0.0
−0.2
1000
2000
3000
4000
chain
1
2
3
4

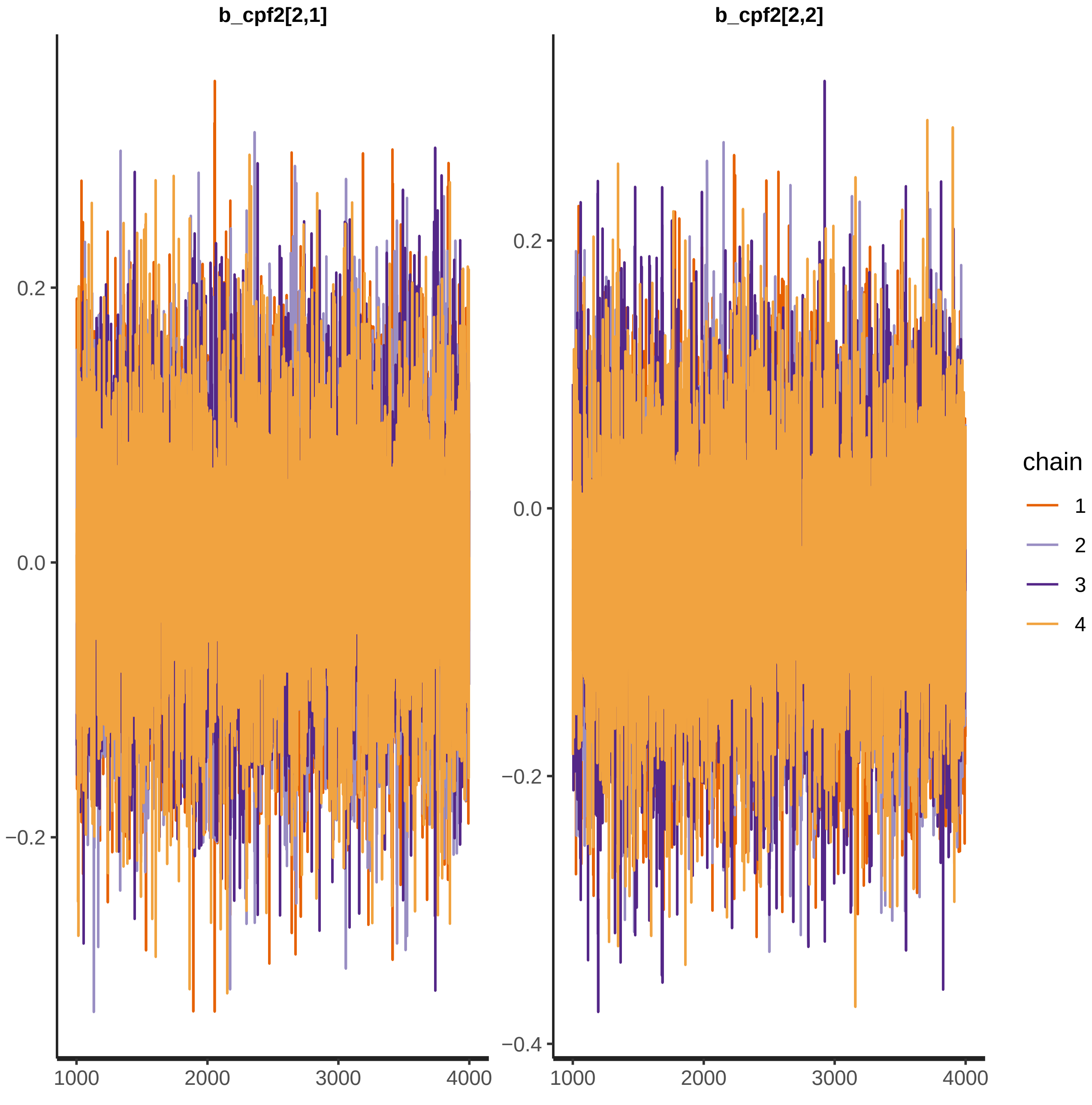

b_cpf2[2,1]
b_cpf2[2,2]
0.2
0.0
−0.2
−0.4
1000
2000
3000
4000
chain
1
2
3
4

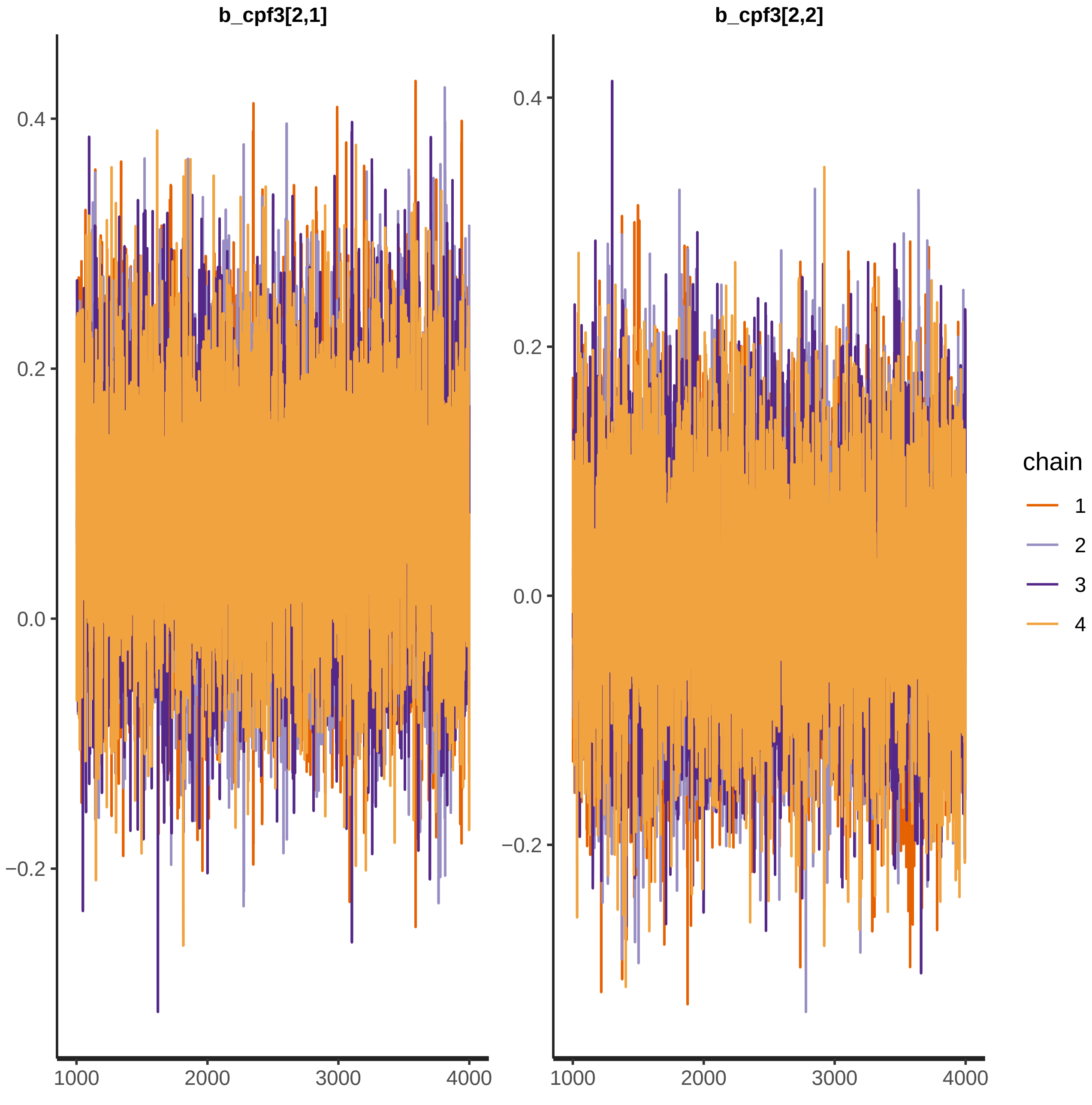
b_cpf3[2,1]
b_cpf3[2,2]
0.4
0.2
0.0
−0.2
1000
2000
3000
4000
chain
1
2
3
4

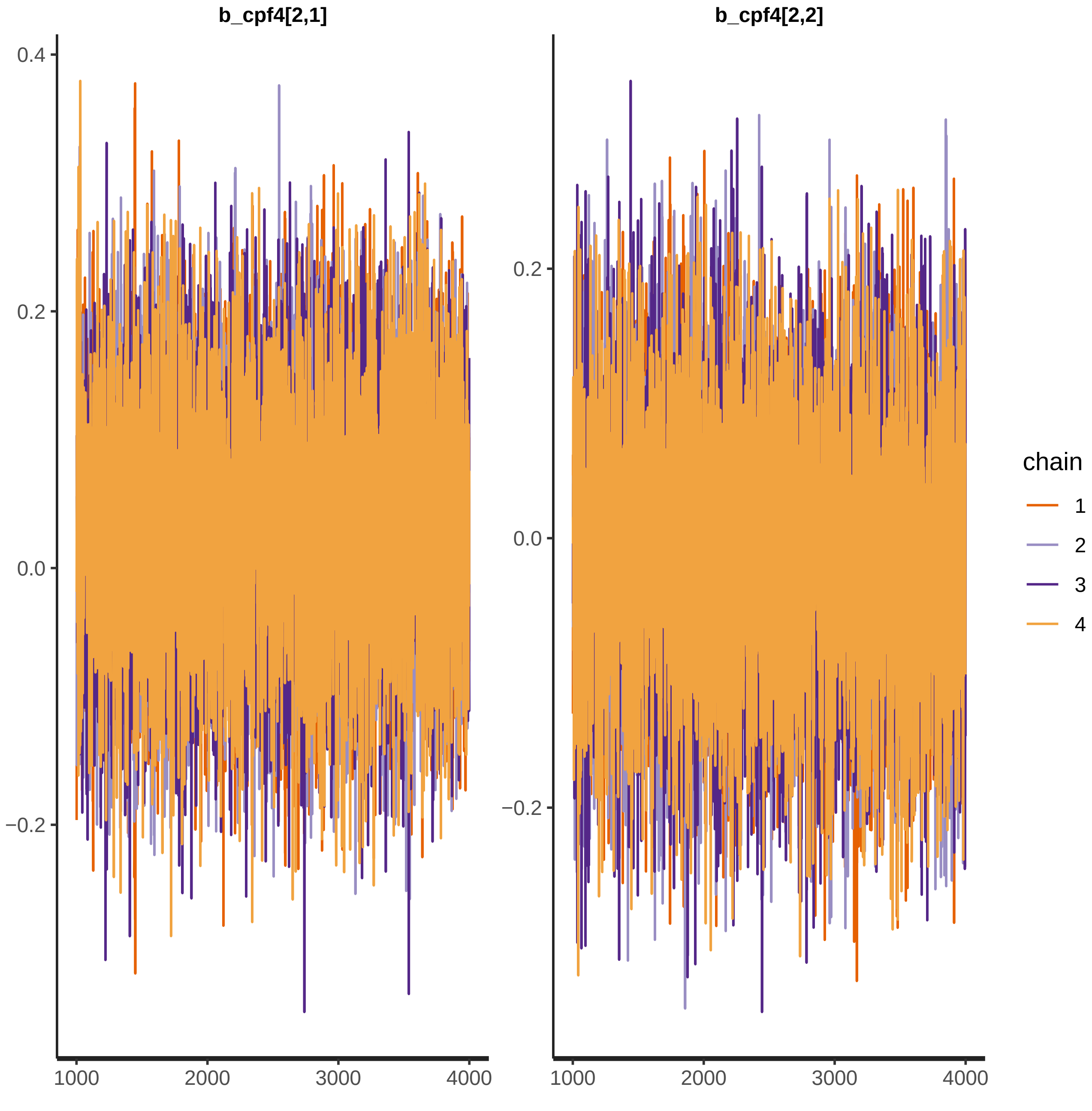
b_cpf4[2,1]
b_cpf4[2,2]
0.4
0.2
0.0
−0.2
1000
2000
3000
4000
chain
1
2
3
4

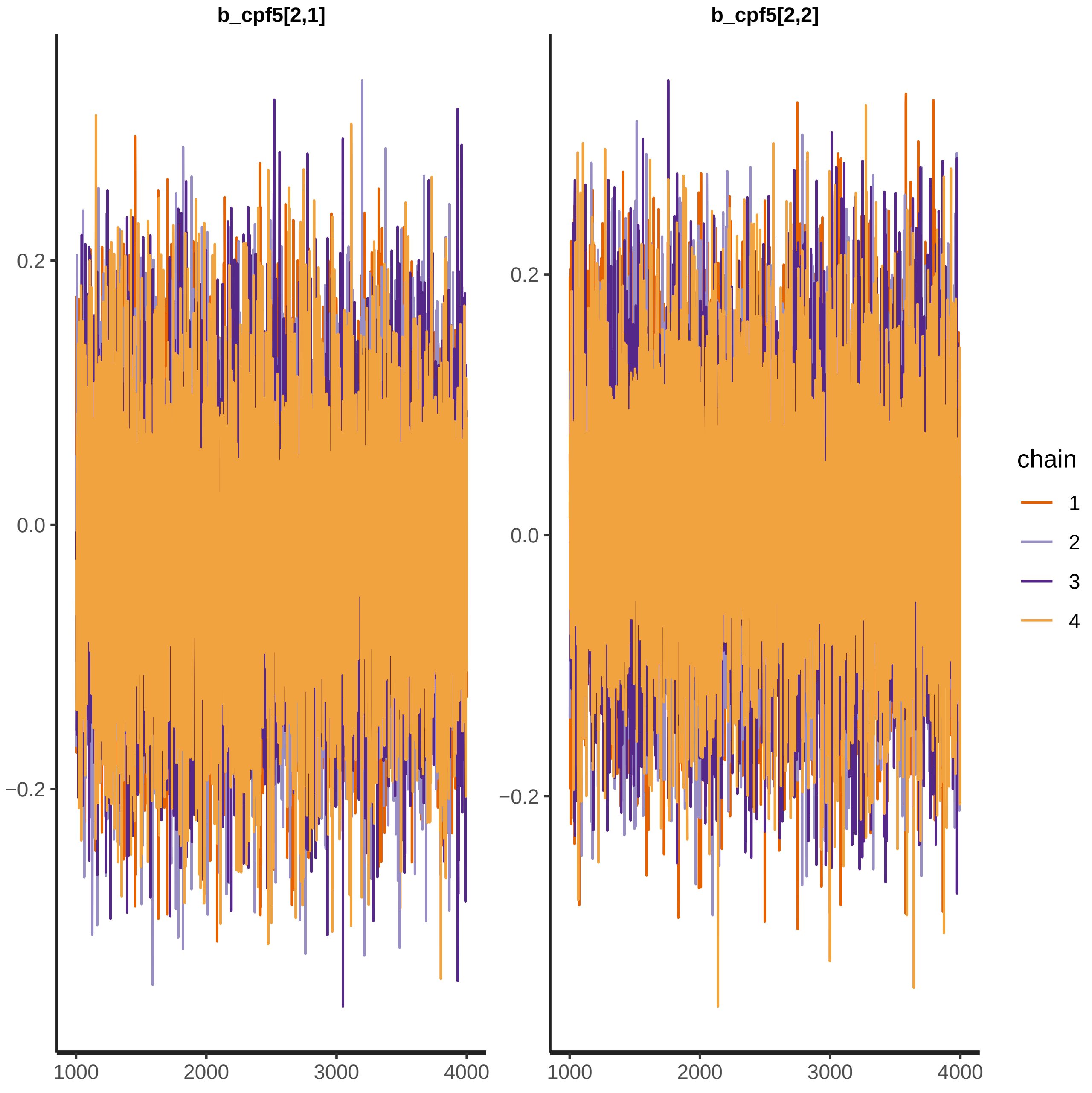

b_cpf5[2,1]
b_cpf5[2,2]
0.2
0.0
−0.2
1000
2000
3000
4000
chain
1
2
3
4

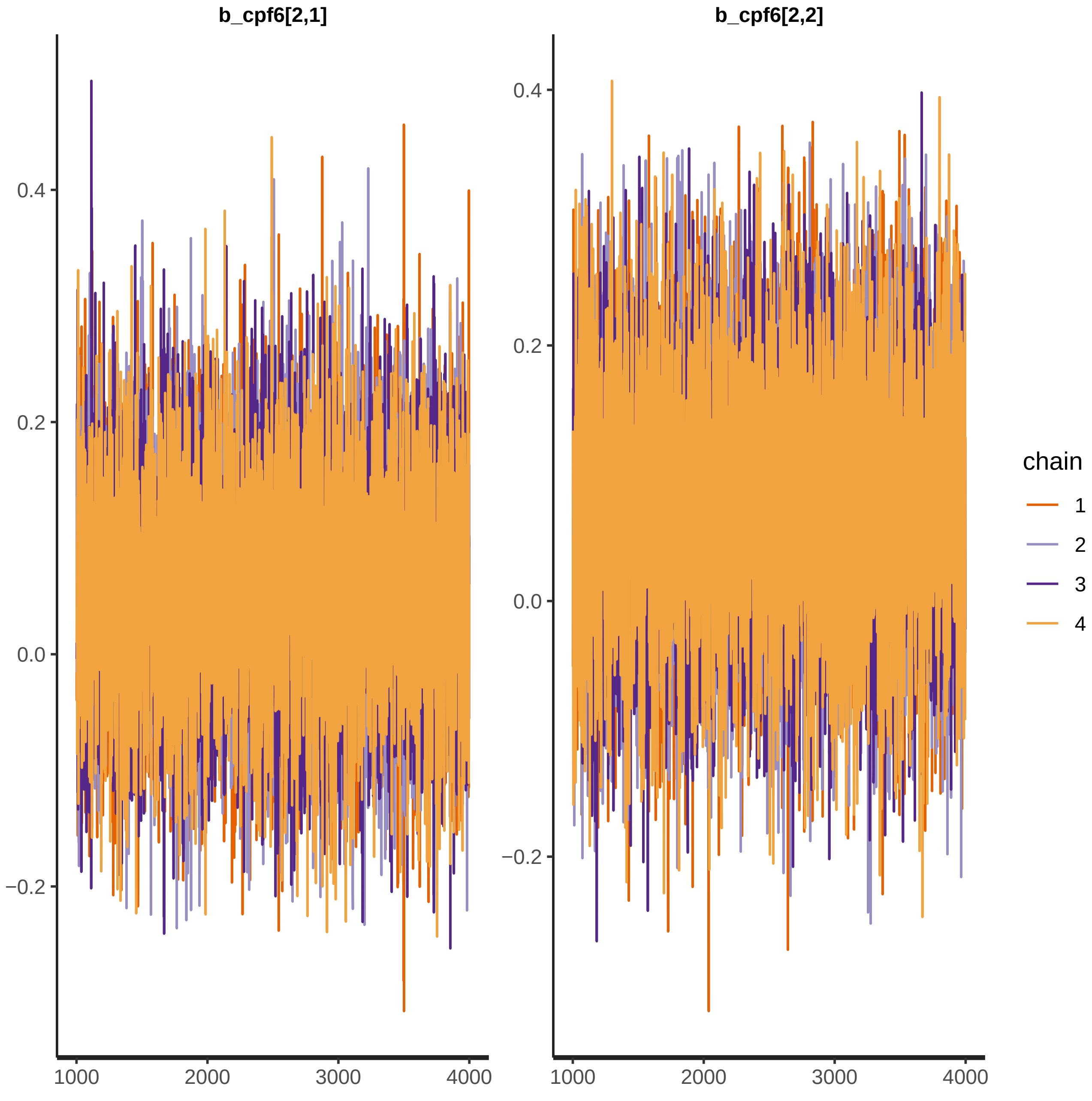
b_cpf6[2,1]
b_cpf6[2,2]
0.4
0.2
0.0
−0.2
1000
2000
3000
4000
chain
1
2
3
4

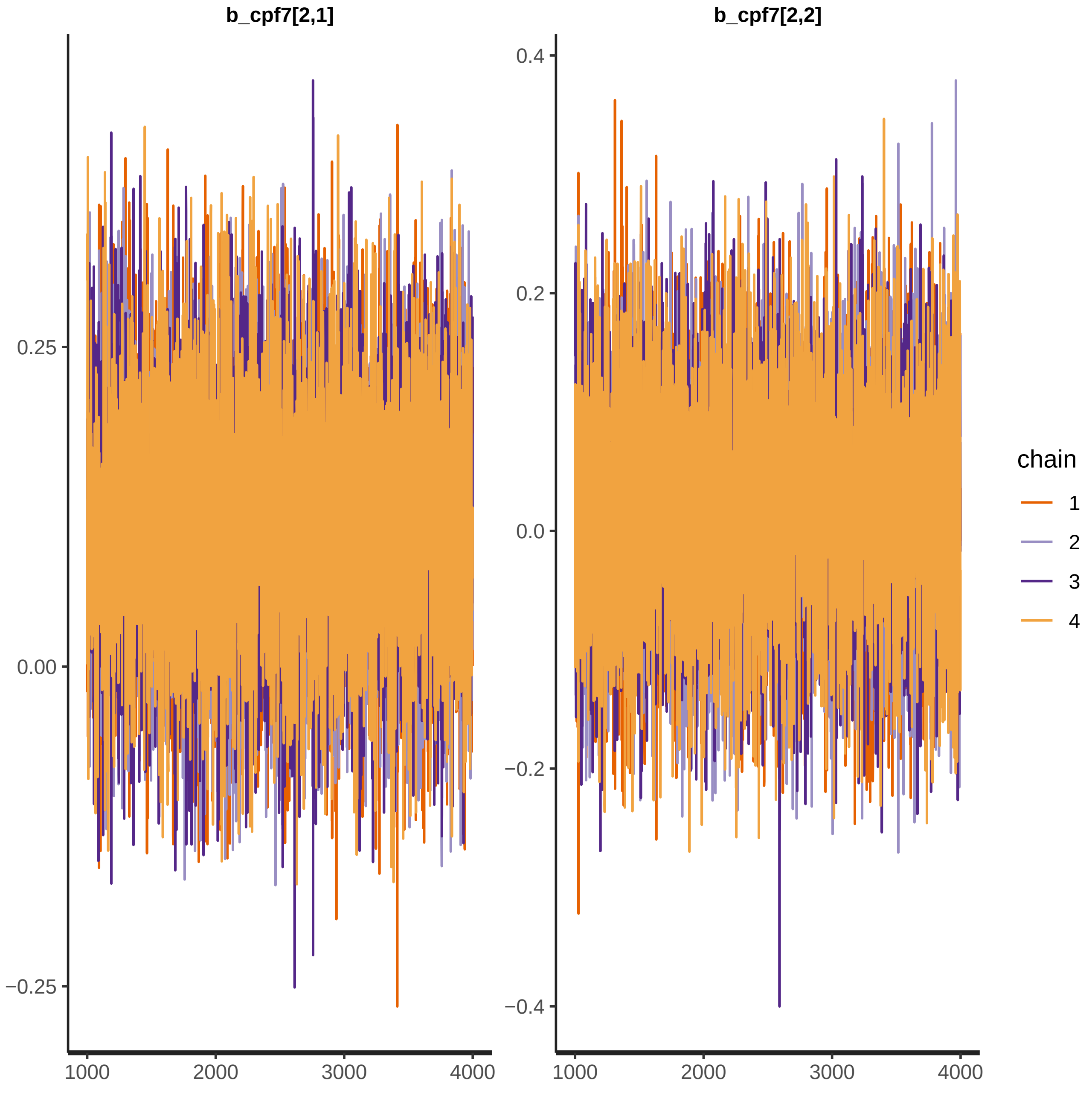

b_cpf7[2,1]
b_cpf7[2,2]
0.25
0.00
−0.25
0.4
0.2
0.0
−0.2
−0.4
1000
2000
3000
4000
chain
1
2
3
4

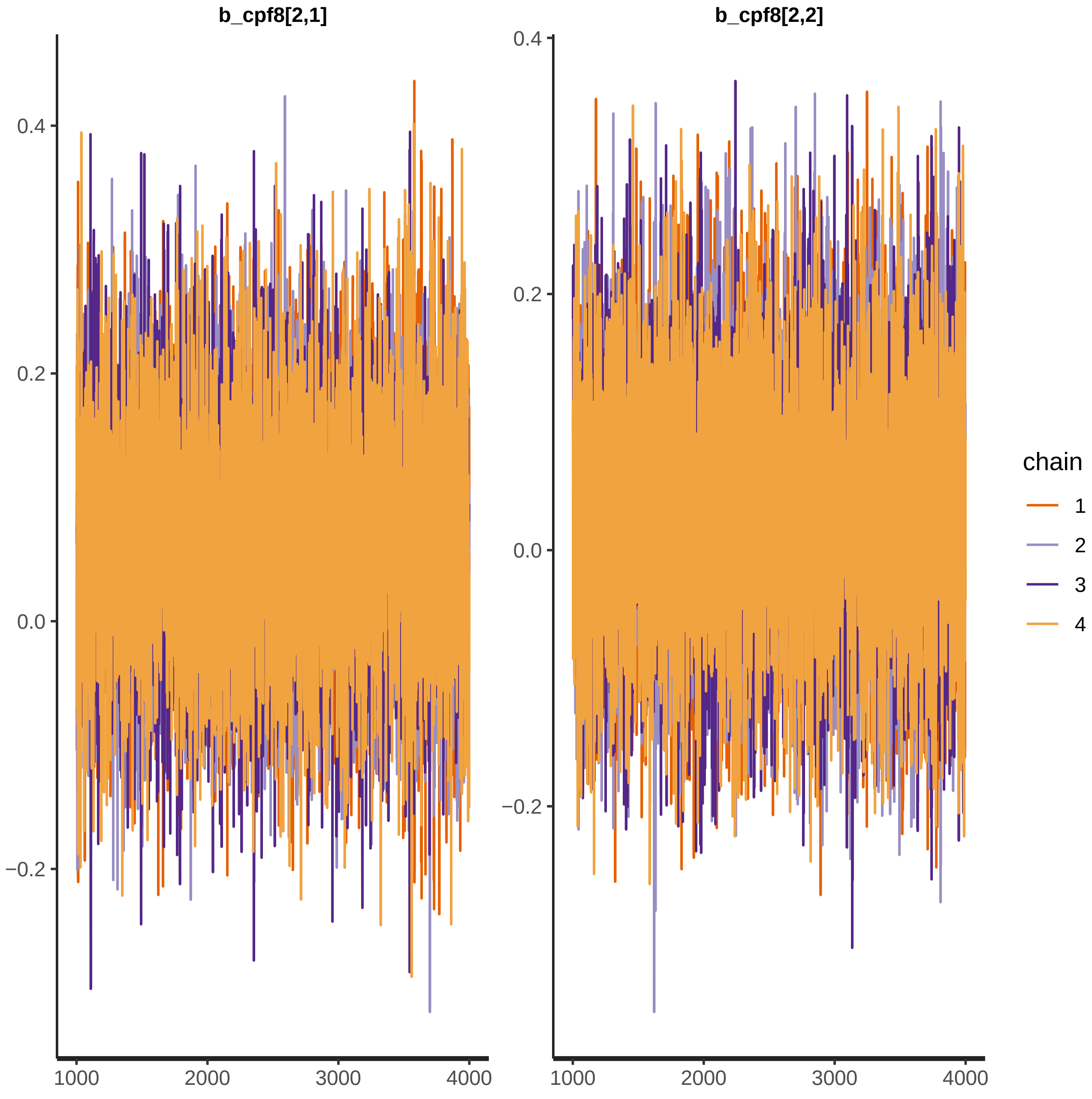
b_cpf8[2,1]
b_cpf8[2,2]
0.4
0.2
0.0
−0.2
1000
2000
3000
4000
chain
1
2
3
4

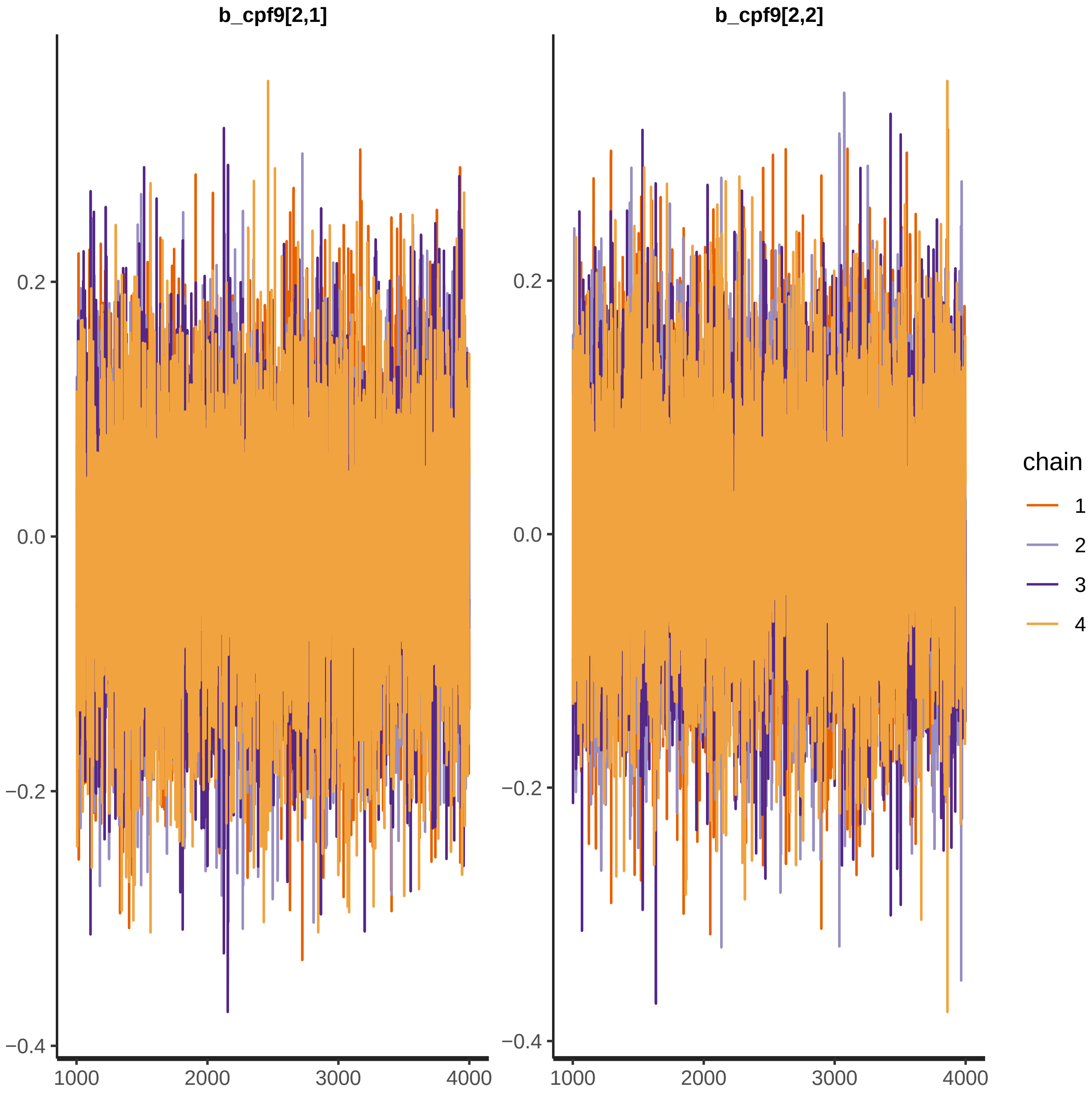

b_cpf9[2,1]
b_cpf9[2,2]
0.2
0.0
−0.2
−0.4
1000
2000
3000
4000
chain
1
2
3
4

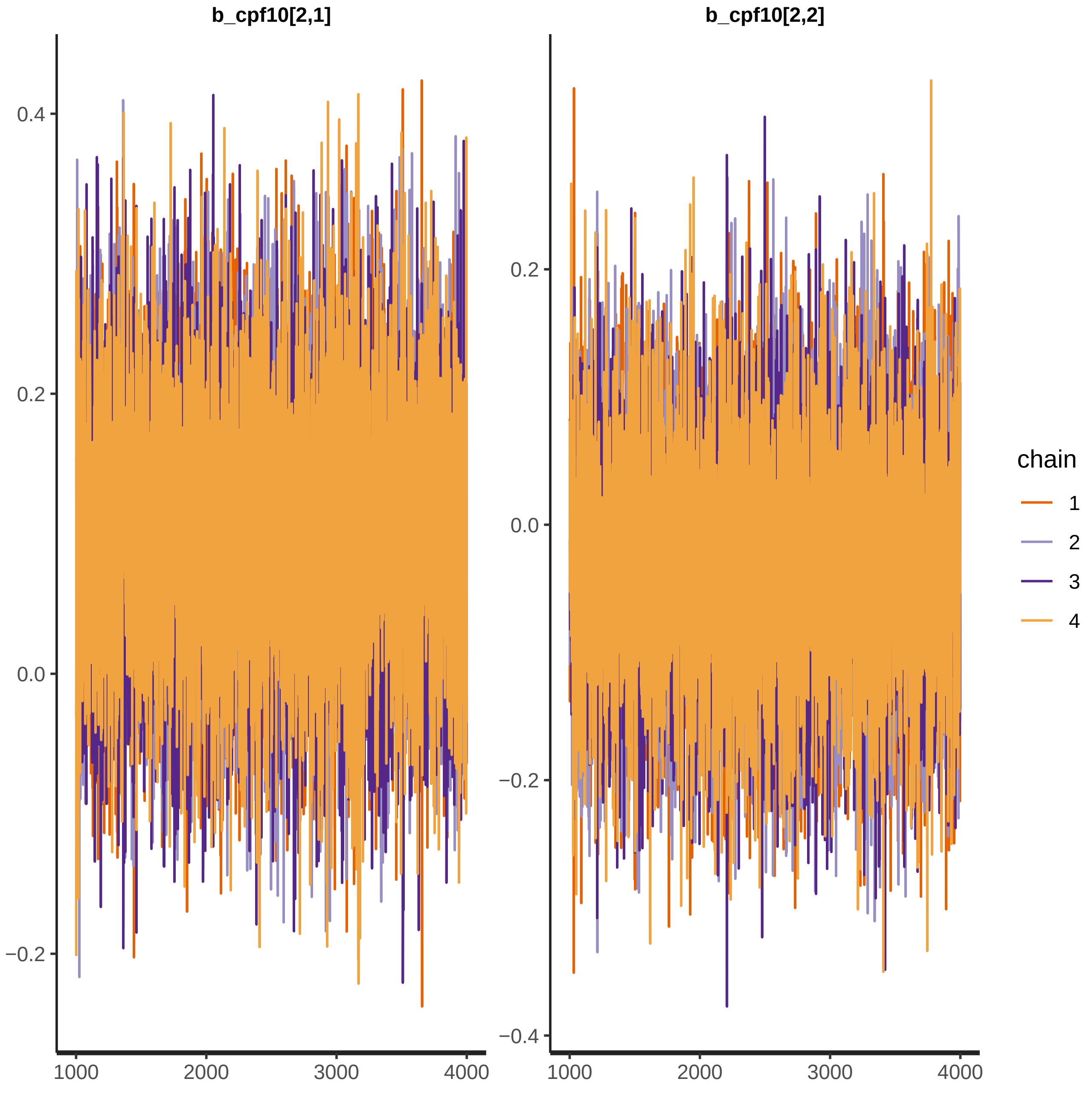
b_cpf10[2,1]
b_cpf10[2,2]
0.4
0.2
0.0
−0.2
−0.4
1000
2000
3000
4000
chain
1
2
3
4

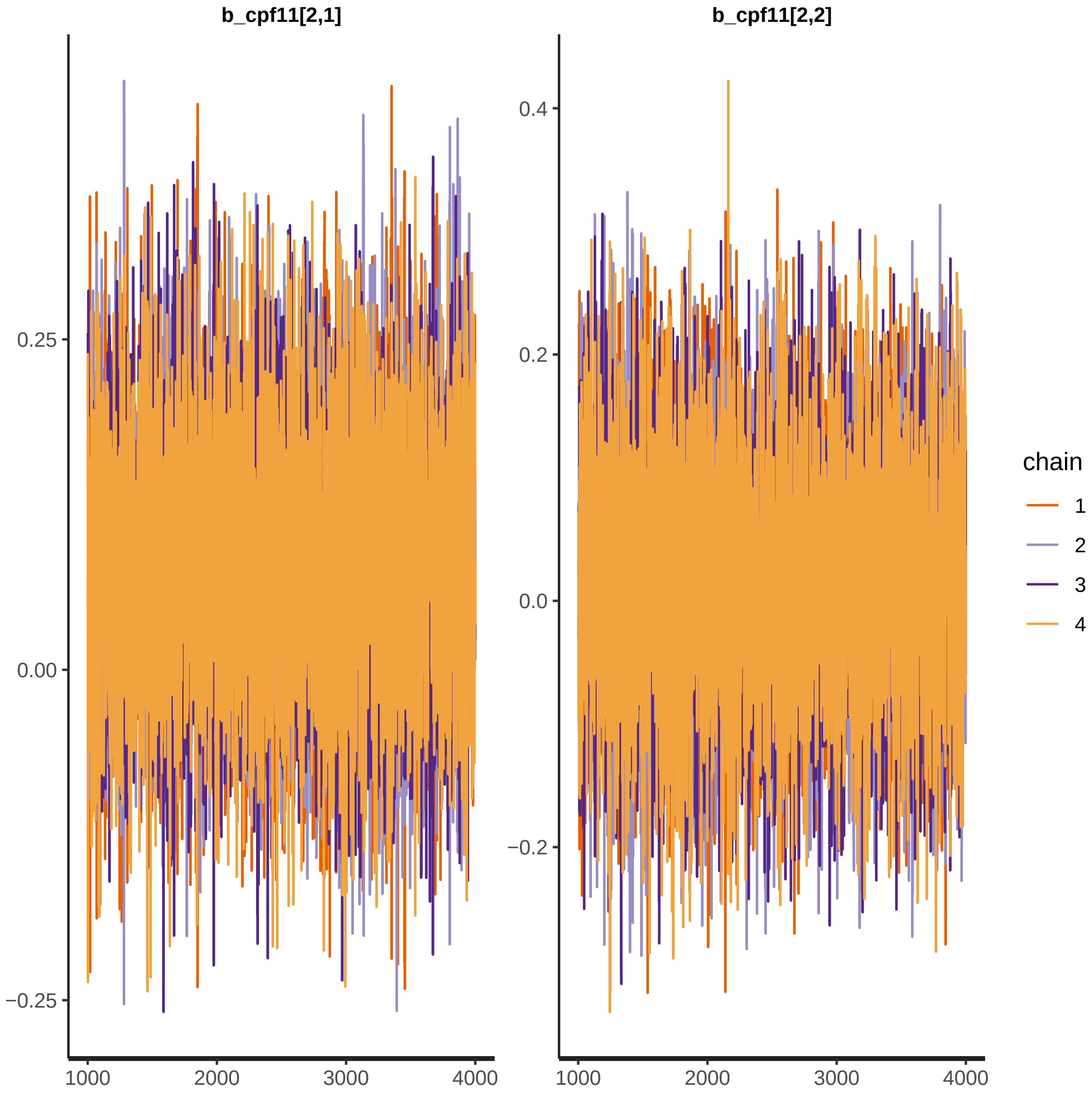

b_cpf11[2,1]
b_cpf11[2,2]
0.25
0.00
−0.25
0.4
0.2
0.0
−0.2
1000
2000
3000
4000
chain
1
2
3
4

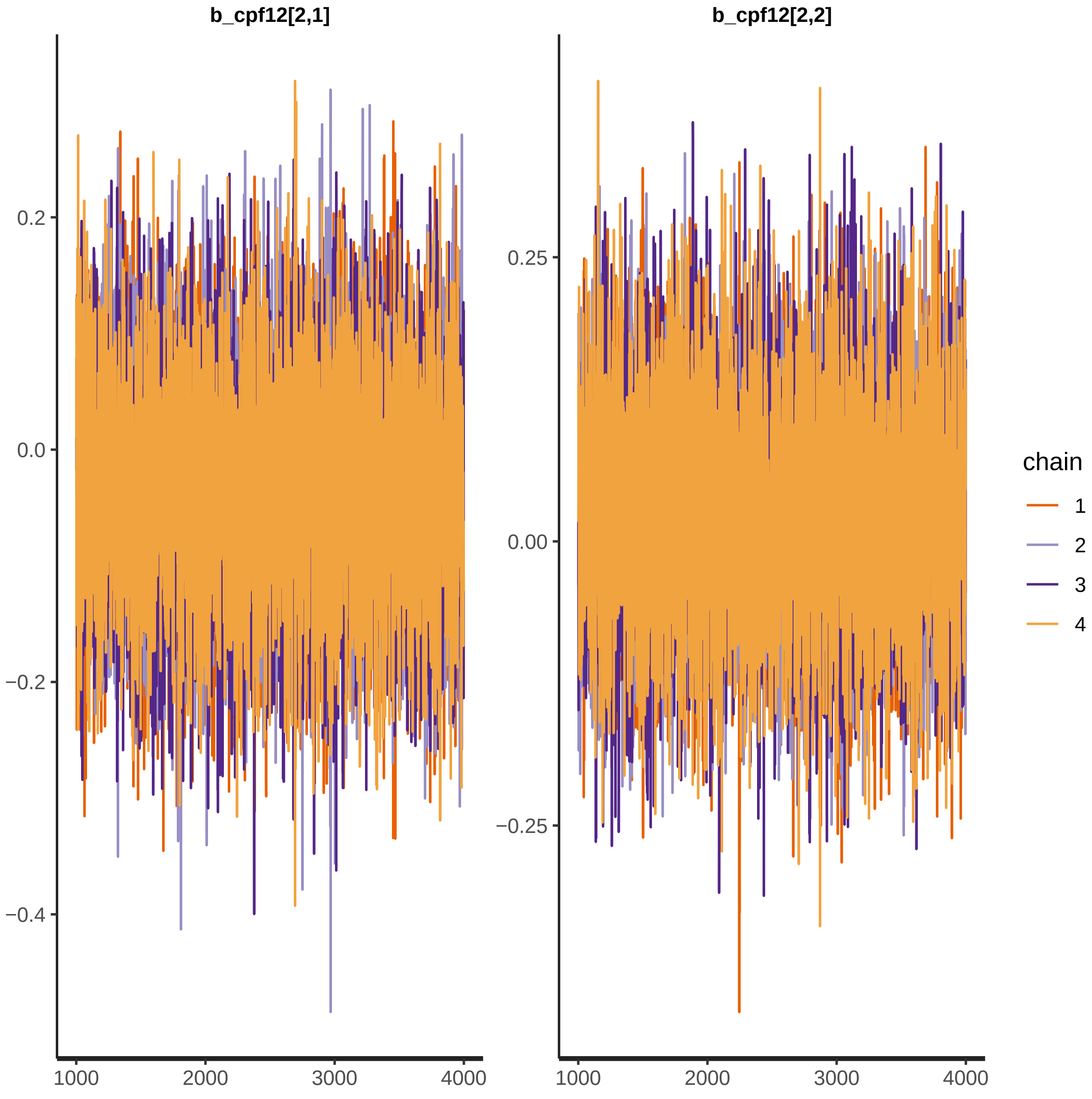

b_cpf12[2,1]
b_cpf12[2,2]
0.2
0.0
−0.2
−0.4
0.25
0.00
−0.25
1000
2000
3000
4000
chain
1
2
3
4

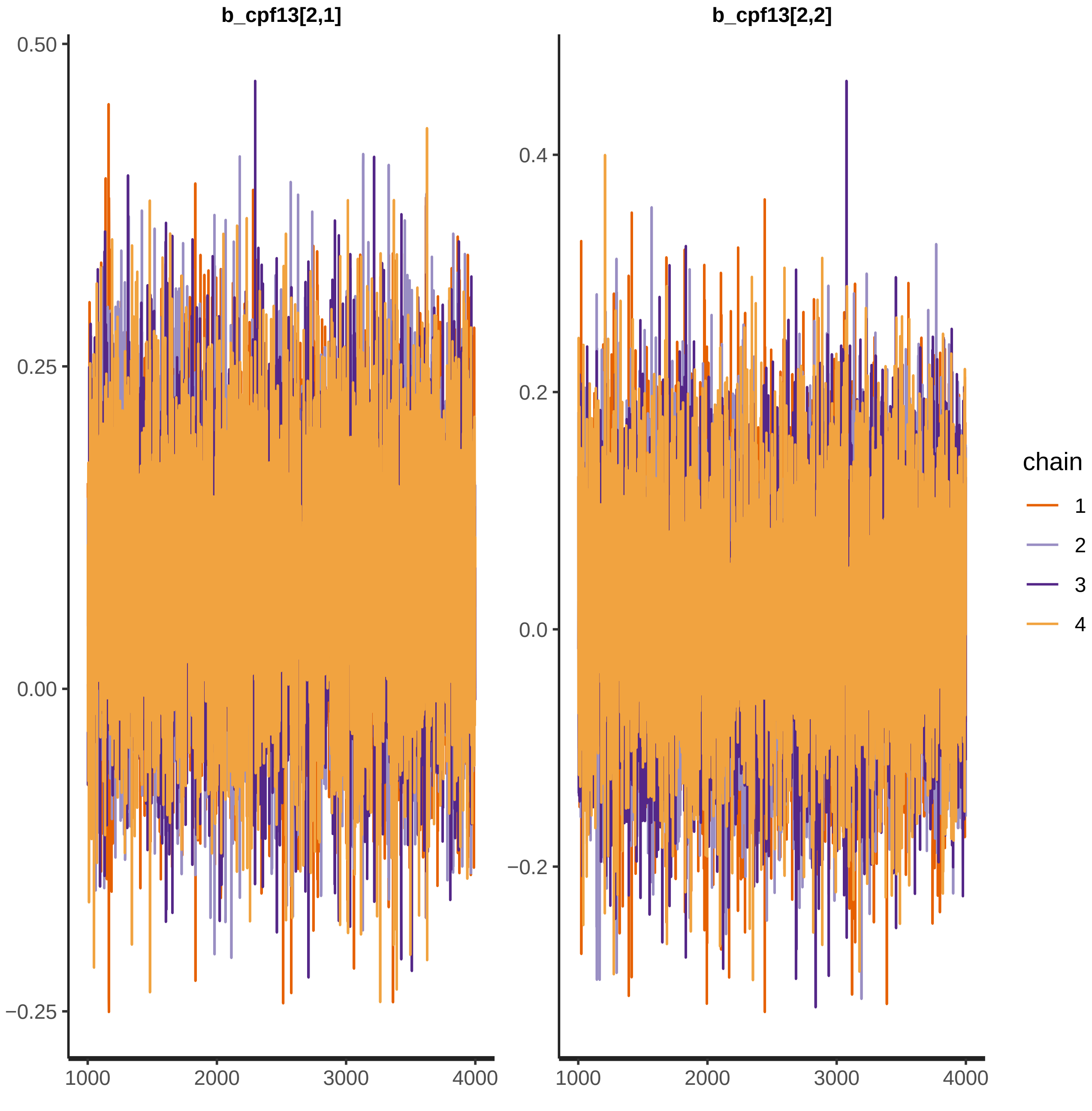
b_cpf13[2,1]
b_cpf13[2,2]
0.50
0.25
0.00
−0.25
0.4
0.2
0.0
−0.2
1000
2000
3000
4000
chain
1
2
3
4

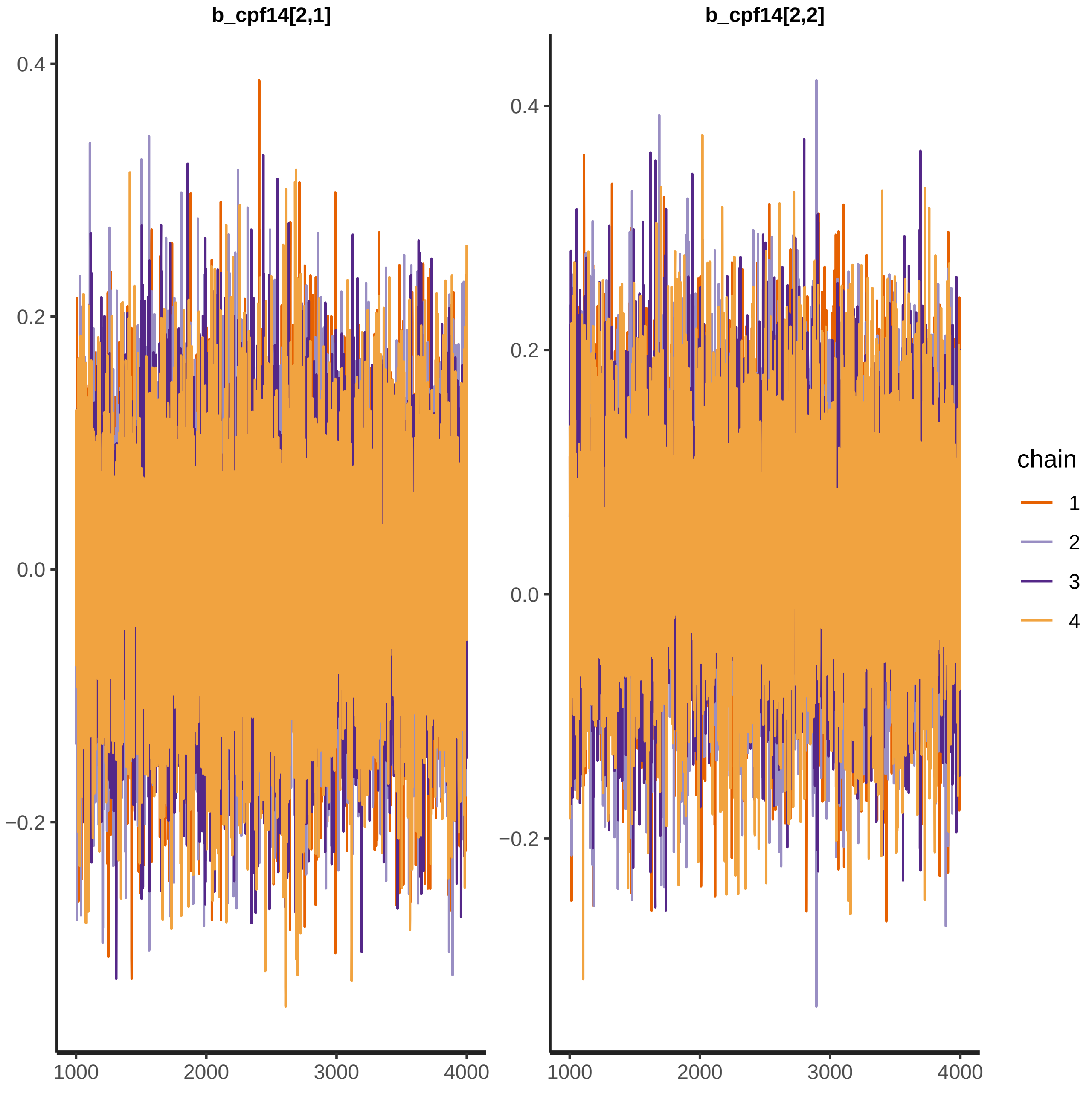

b_cpf14[2,1]
b_cpf14[2,2]
0.4
0.2
0.0
−0.2
1000
2000
3000
4000
chain
1
2
3
4

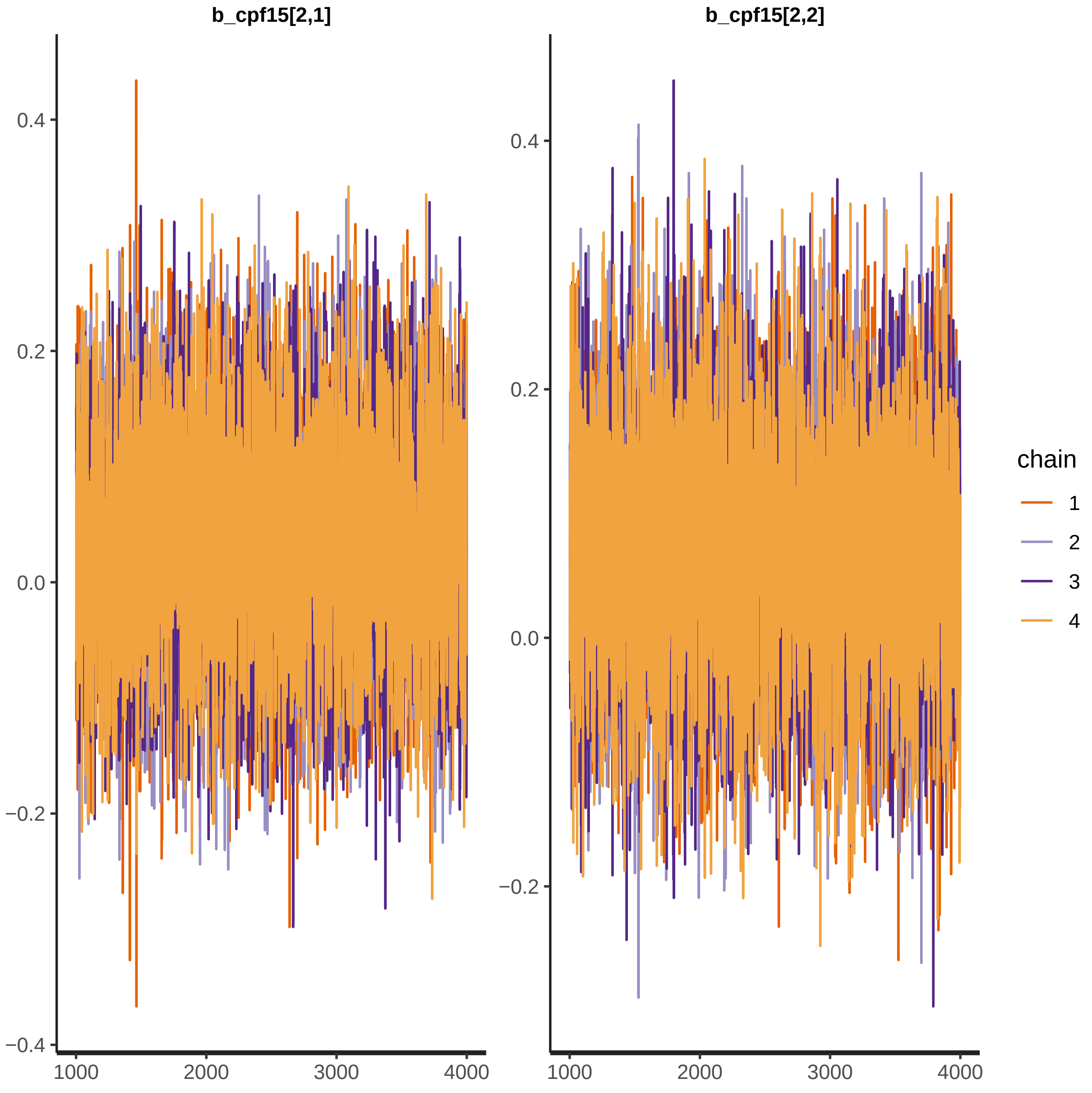
b_cpf15[2,1]
b_cpf15[2,2]
0.4
0.2
0.0
−0.2
−0.4
1000
2000
3000
4000
chain
1
2
3
4

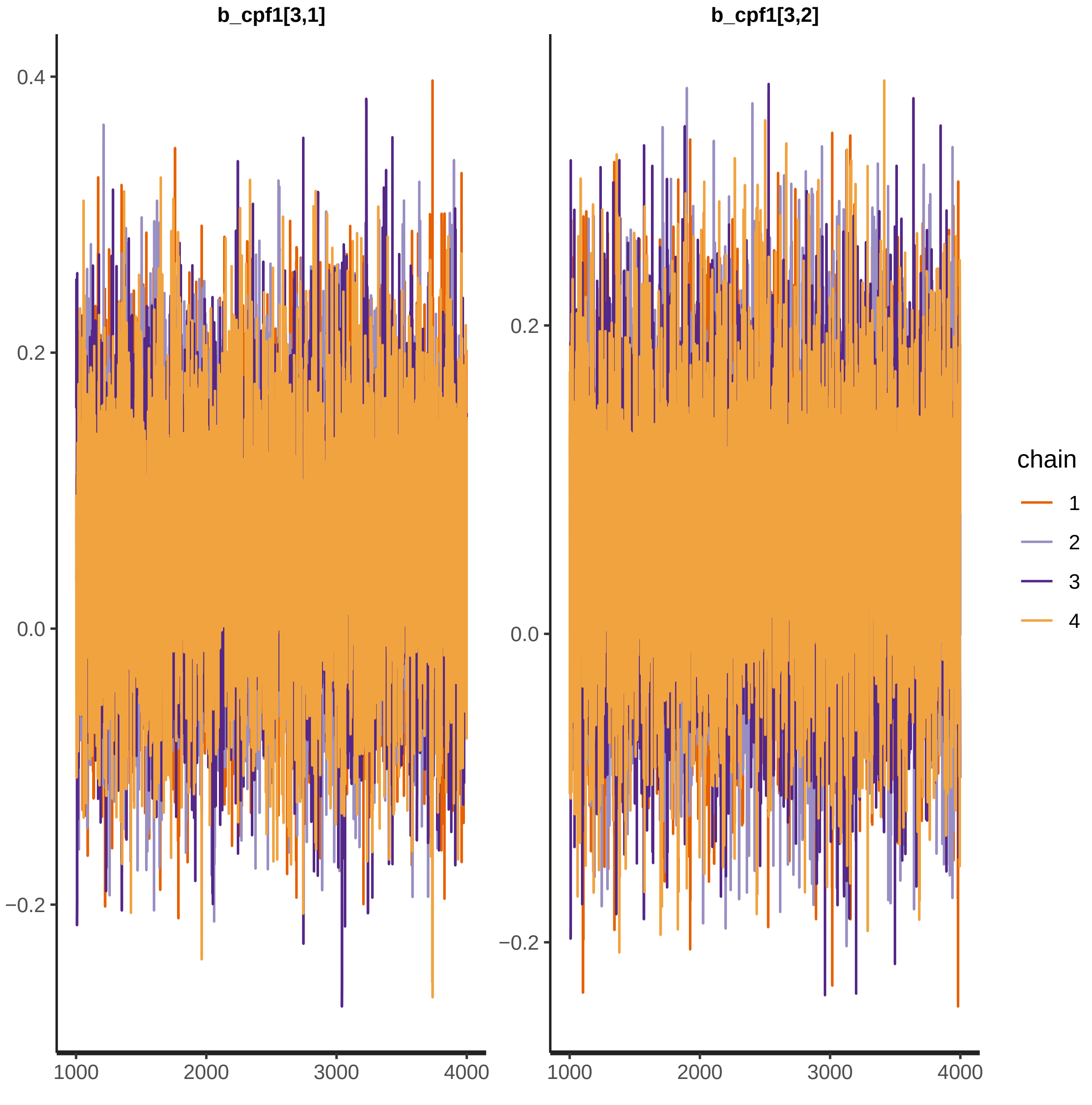

b_cpf1[3,1]
b_cpf1[3,2]
0.4
0.2
0.0
−0.2
1000
2000
3000
4000
chain
1
2
3
4

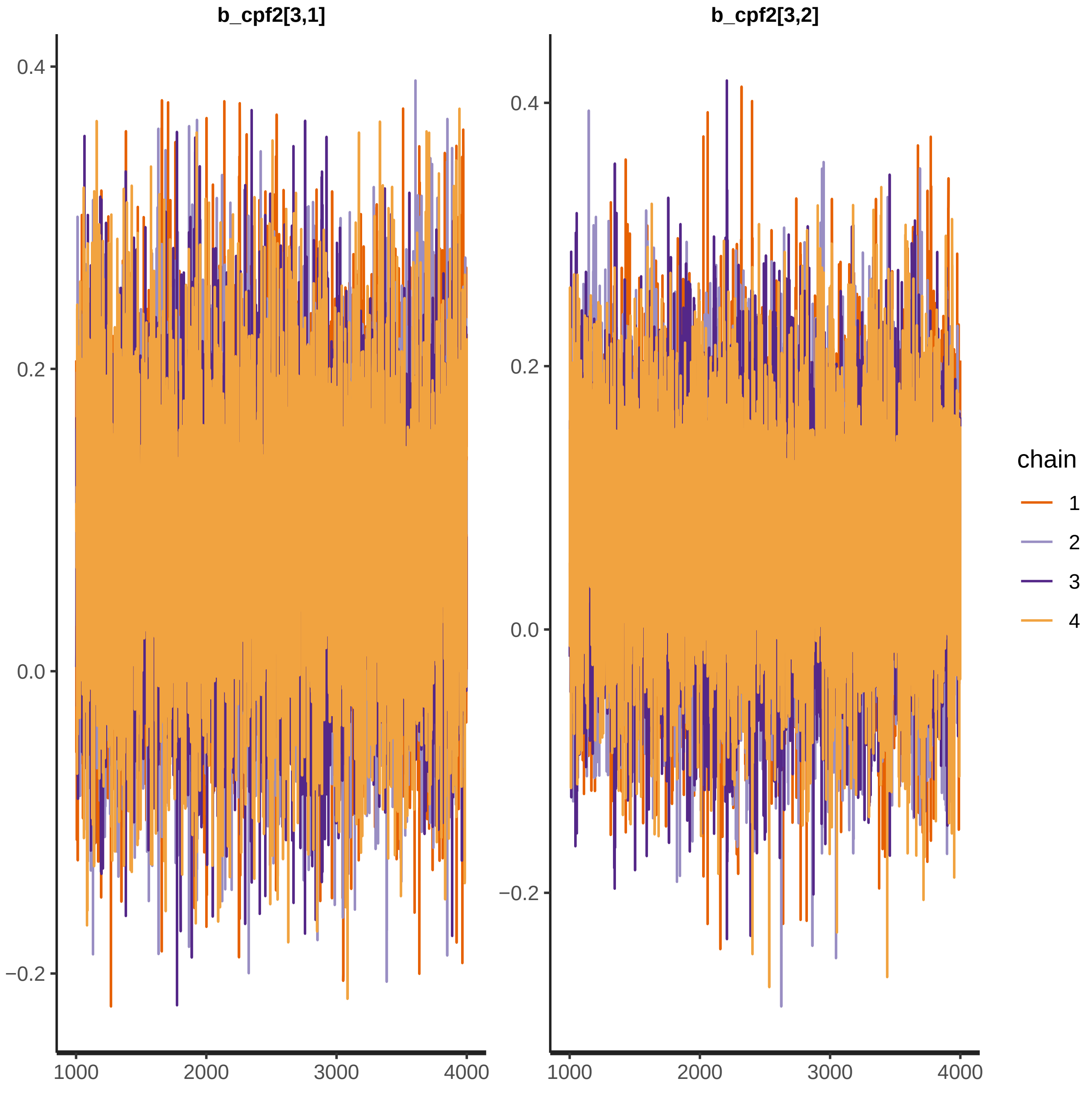

b_cpf2[3,1]
b_cpf2[3,2]
0.4
0.2
0.0
−0.2
1000
2000
3000
4000
chain
1
2
3
4

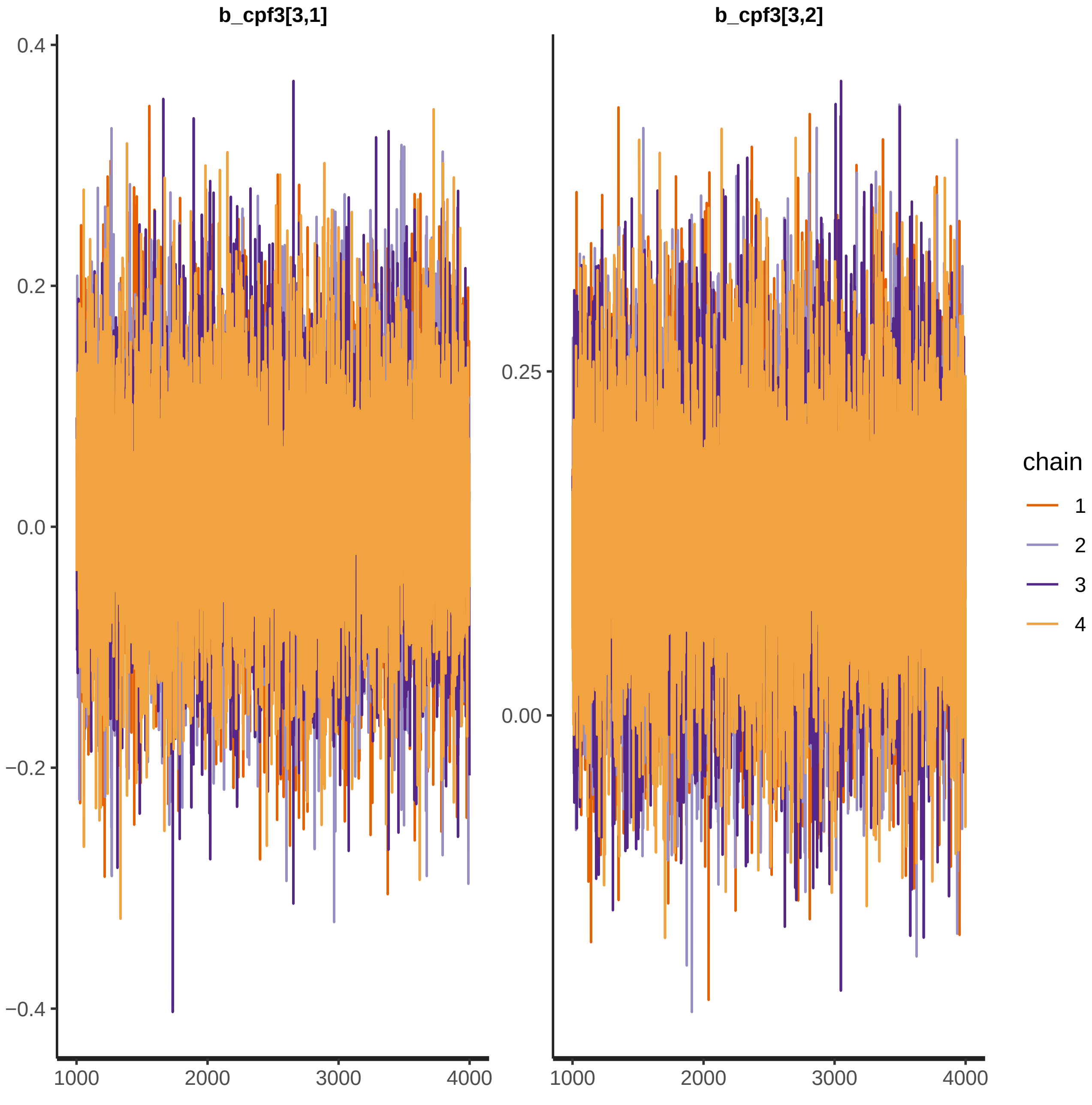

b_cpf3[3,1]
b_cpf3[3,2]
0.4
0.2
0.0
−0.2
−0.4
0.25
0.00
1000
2000
3000
4000
chain
1
2
3
4

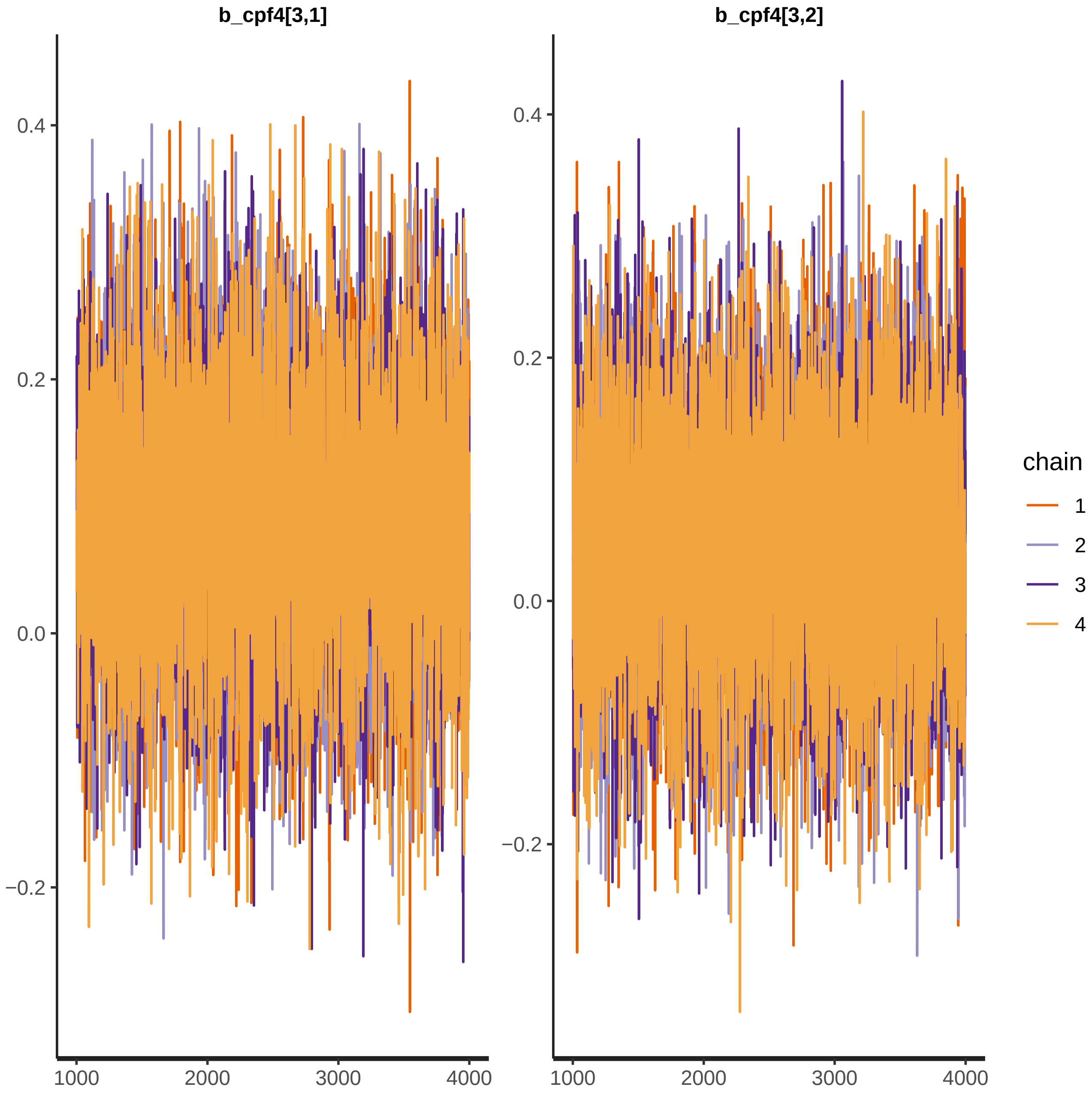

b_cpf4[3,1]
b_cpf4[3,2]
0.4
0.2
0.0
−0.2
1000
2000
3000
4000
chain
1
2
3
4

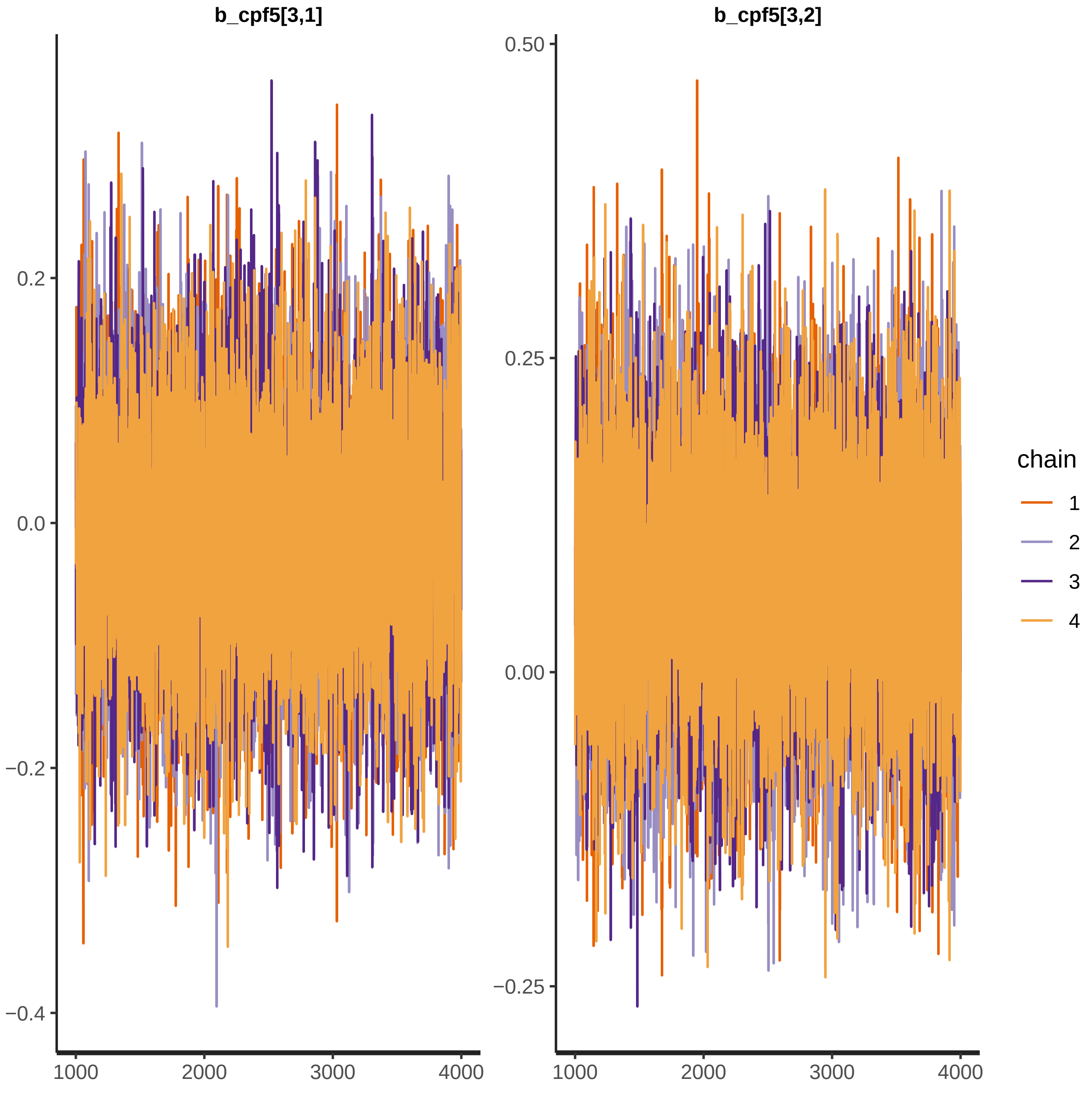

b_cpf5[3,1]
b_cpf5[3,2]
0.2
0.0
−0.2
−0.4
0.50
0.25
0.00
−0.25
1000
2000
3000
4000
chain
1
2
3
4

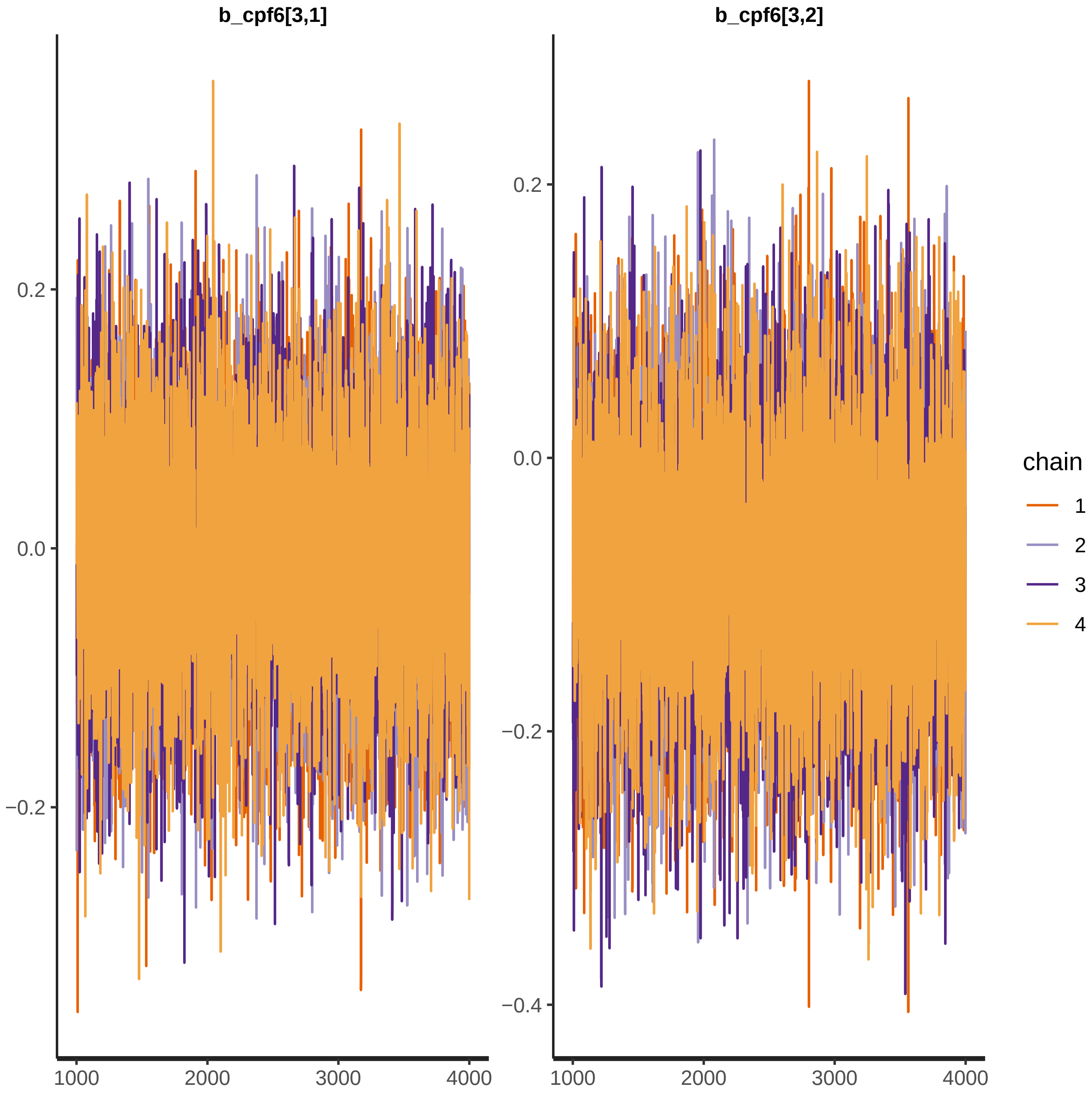

b_cpf6[3,1]
b_cpf6[3,2]
0.2
0.0
−0.2
−0.4
1000
2000
3000
4000
chain
1
2
3
4

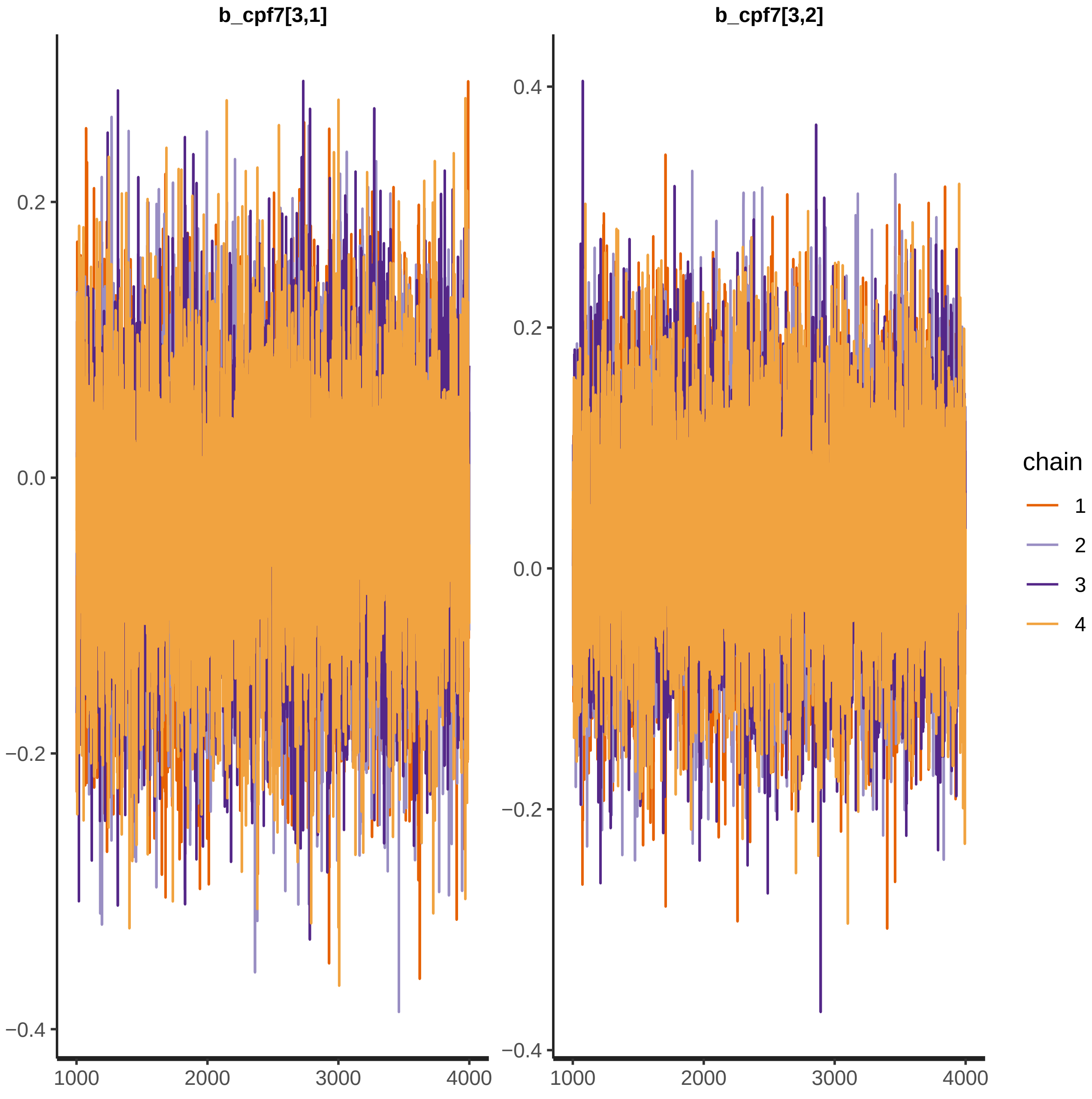
b_cpf7[3,1]
b_cpf7[3,2]
0.2
0.0
−0.2
−0.4
0.4
1000
2000
3000
4000
chain
1
2
3
4

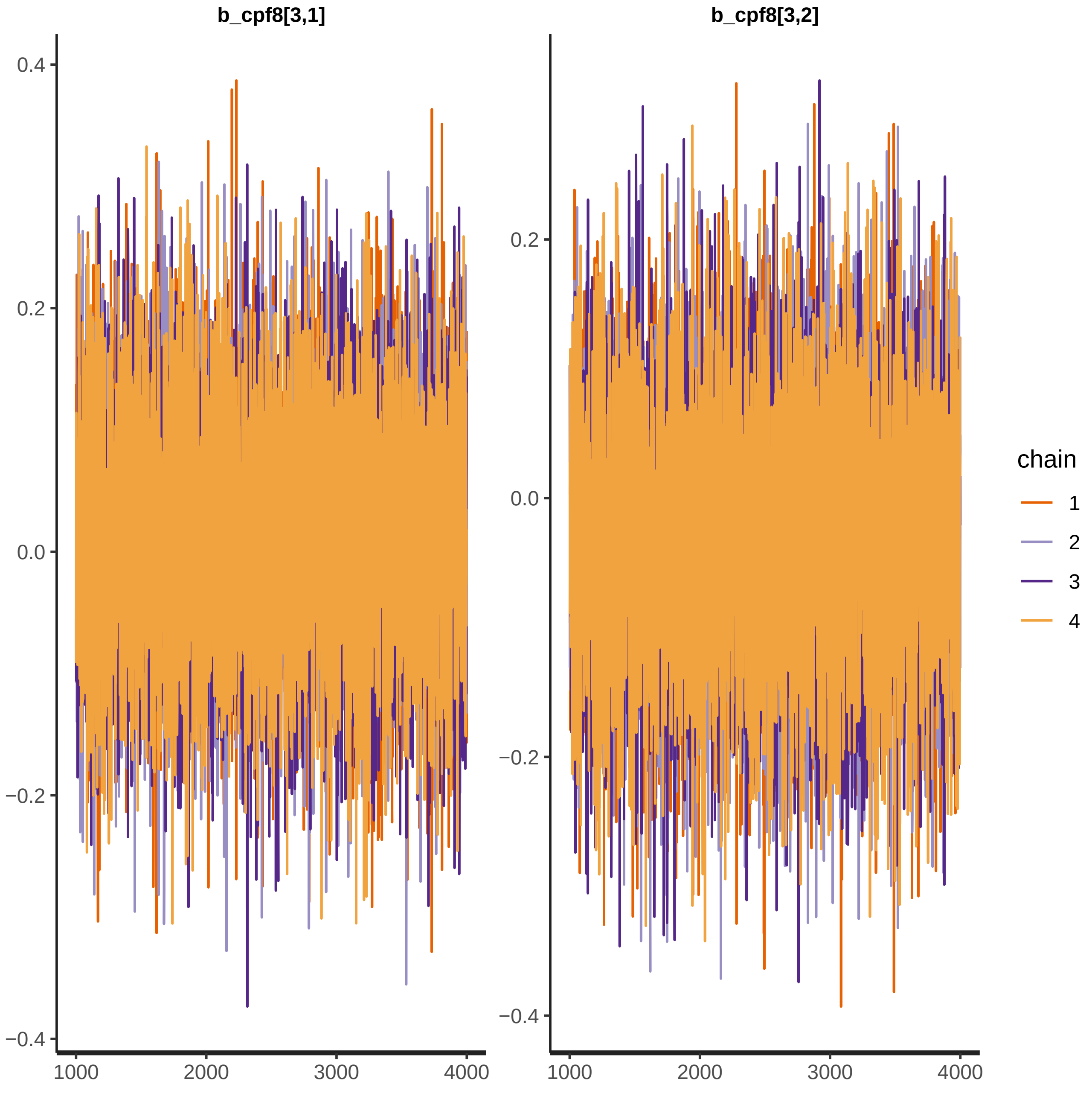
b_cpf8[3,1]
b_cpf8[3,2]
0.4
0.2
0.0
−0.2
−0.4
1000
2000
3000
4000
chain
1
2
3
4

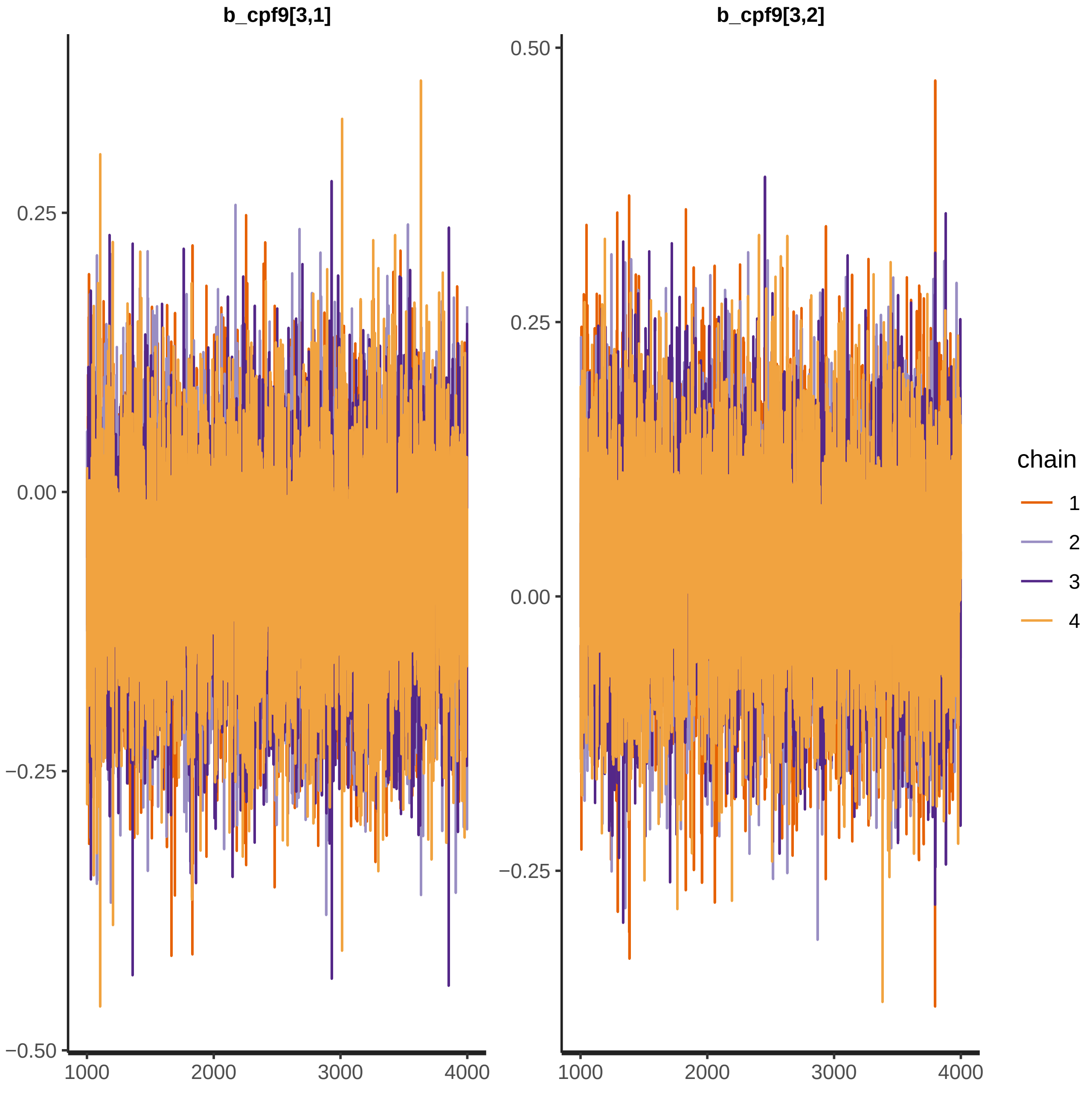

b_cpf9[3,1]
b_cpf9[3,2]
0.50
0.25
0.00
−0.25
−0.50
1000
2000
3000
4000
chain
1
2
3
4

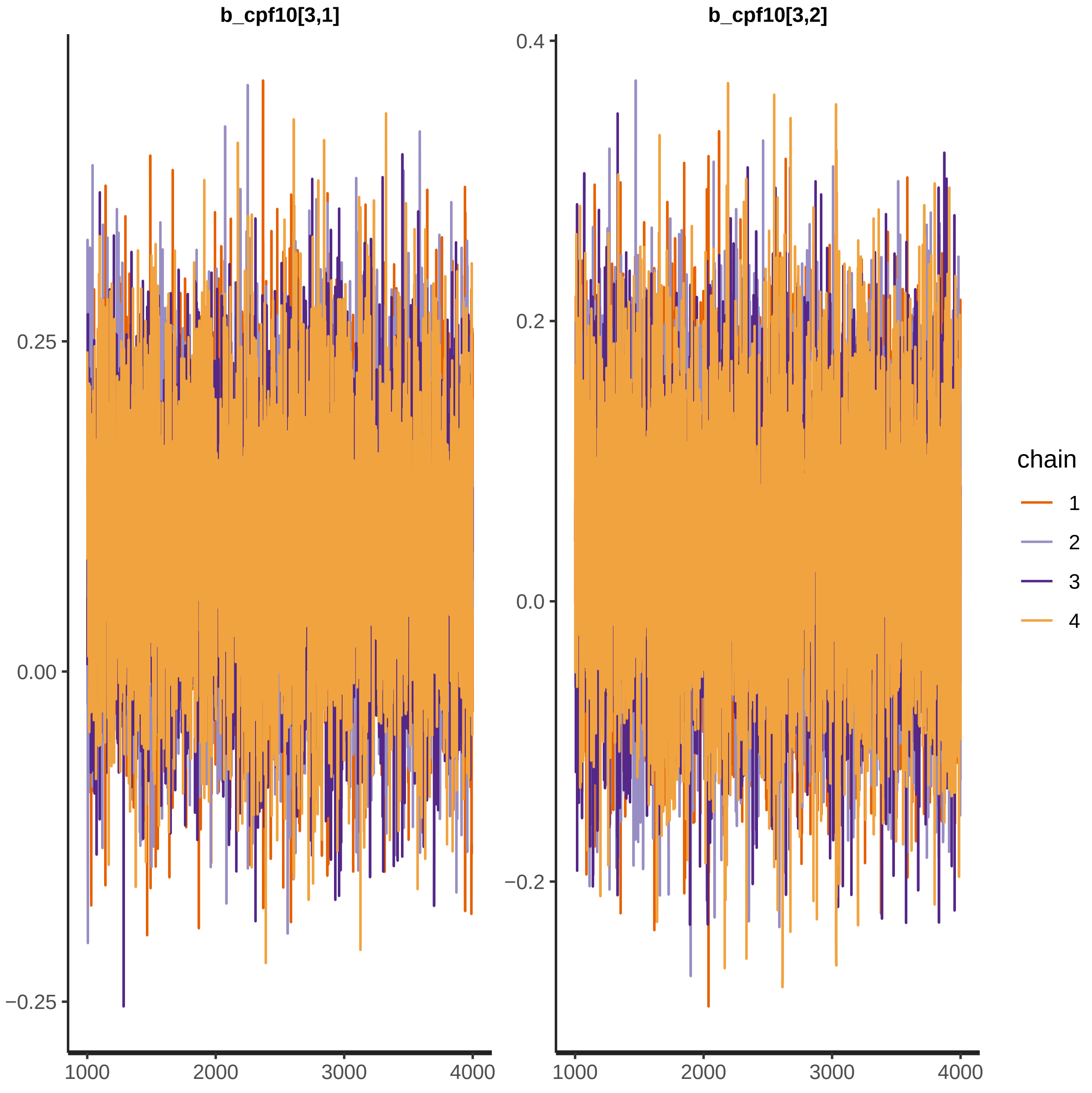

b_cpf10[3,1]
b_cpf10[3,2]
0.25
0.00
−0.25
0.4
0.2
0.0
−0.2
1000
2000
3000
4000
chain
1
2
3
4

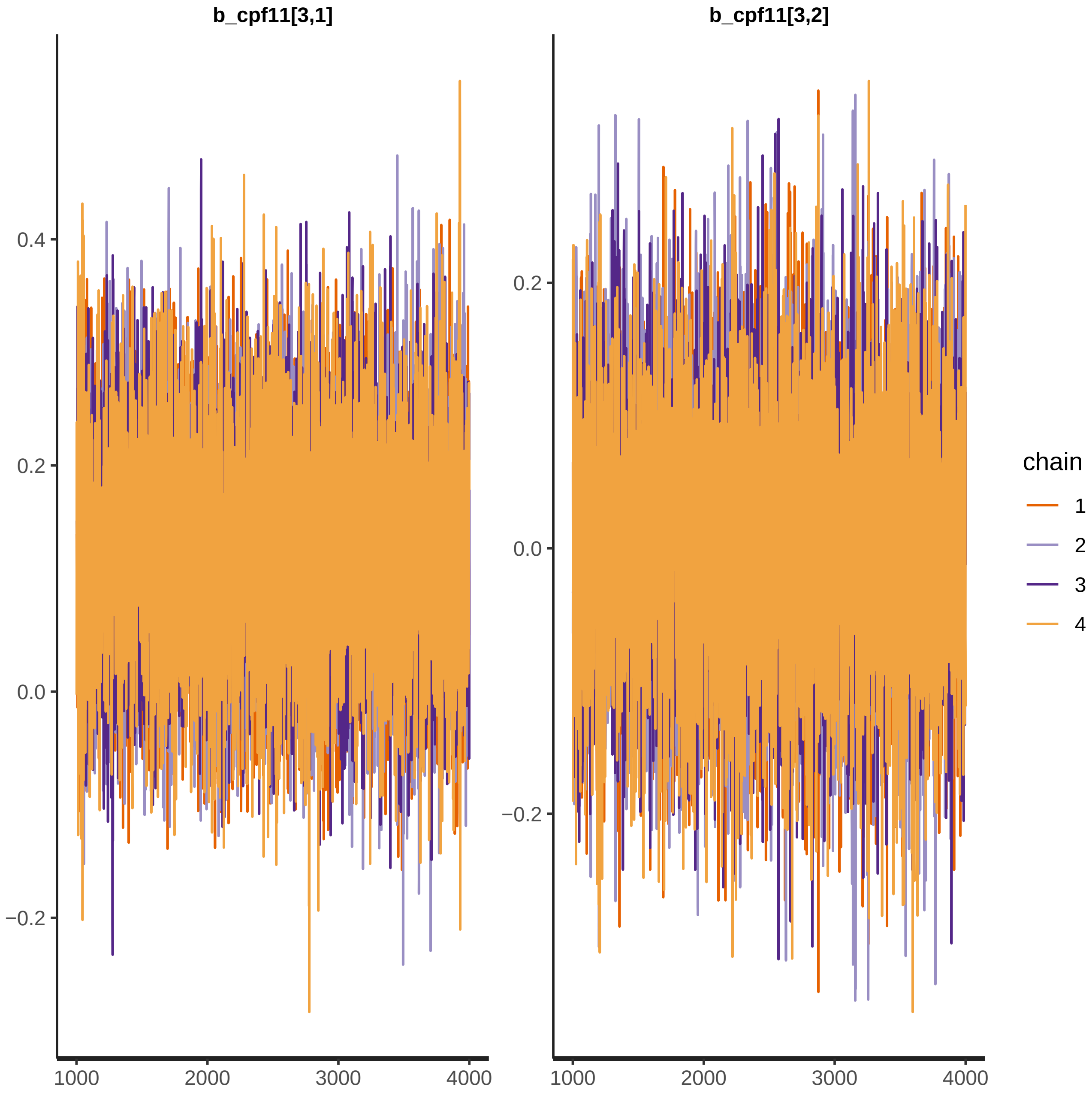

b_cpf11[3,1]
b_cpf11[3,2]
0.4
0.2
0.0
−0.2
0.2
0.0
−0.2
1000
2000
3000
4000
1000
2000
3000
4000
chain
1
2
3
4

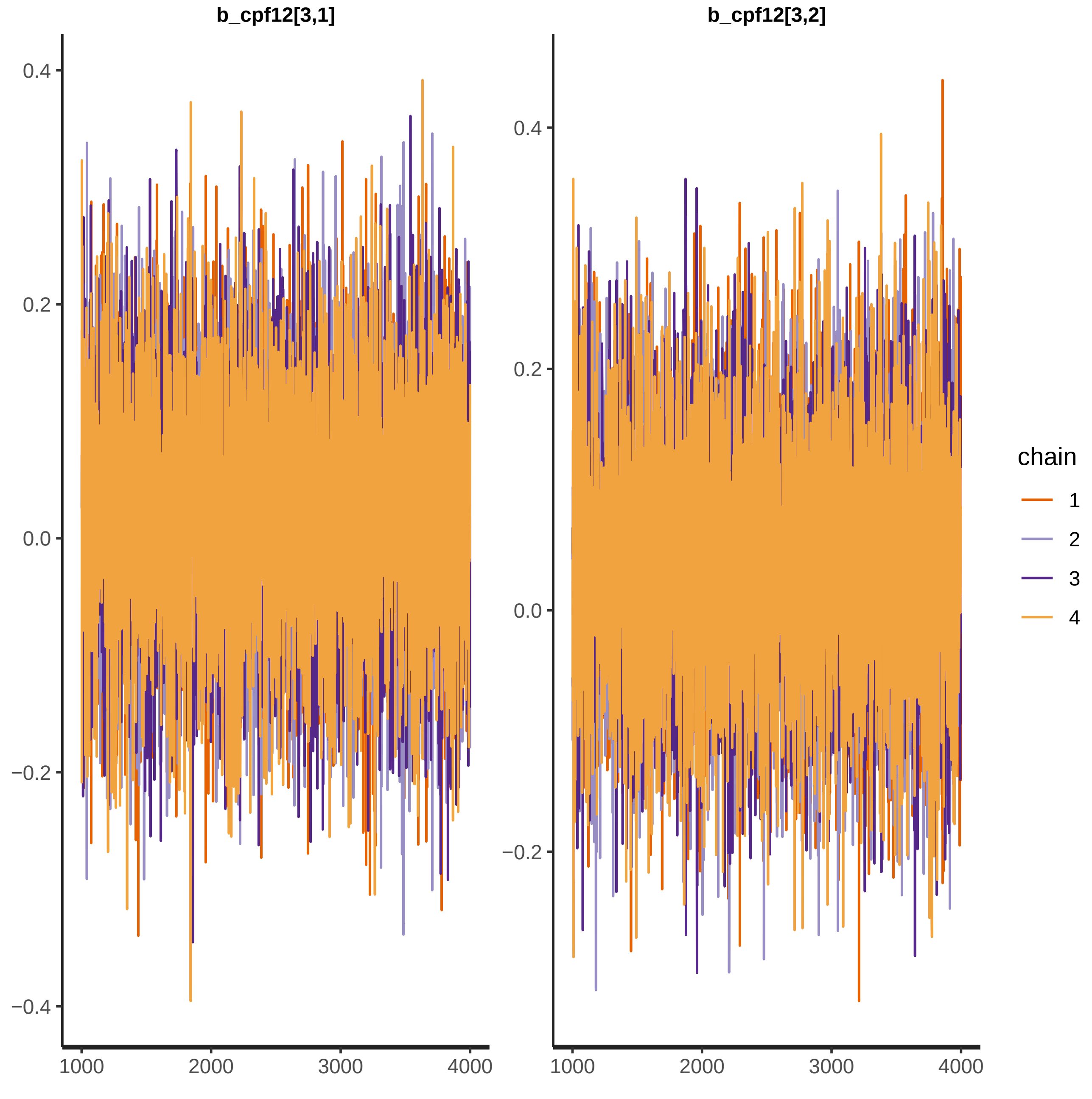

b_cpf12[3,1]
b_cpf12[3,2]
0.4
0.2
0.0
−0.2
−0.4
1000
2000
3000
4000
chain
1
2
3
4

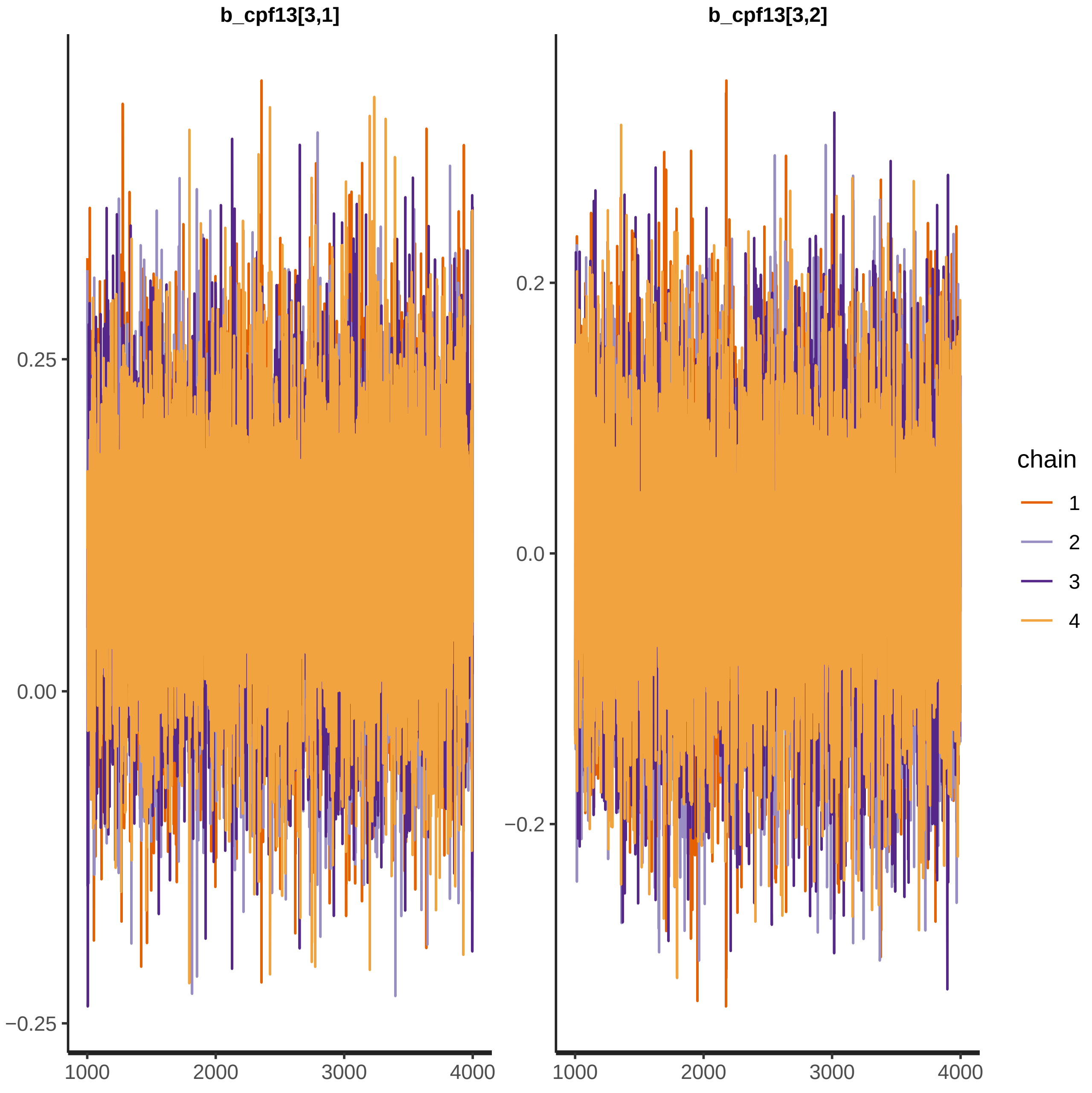
b_cpf13[3,1]
b_cpf13[3,2]
0.25
0.00
−0.25
0.2
0.0
−0.2
1000
2000
3000
4000
chain
1
2
3
4

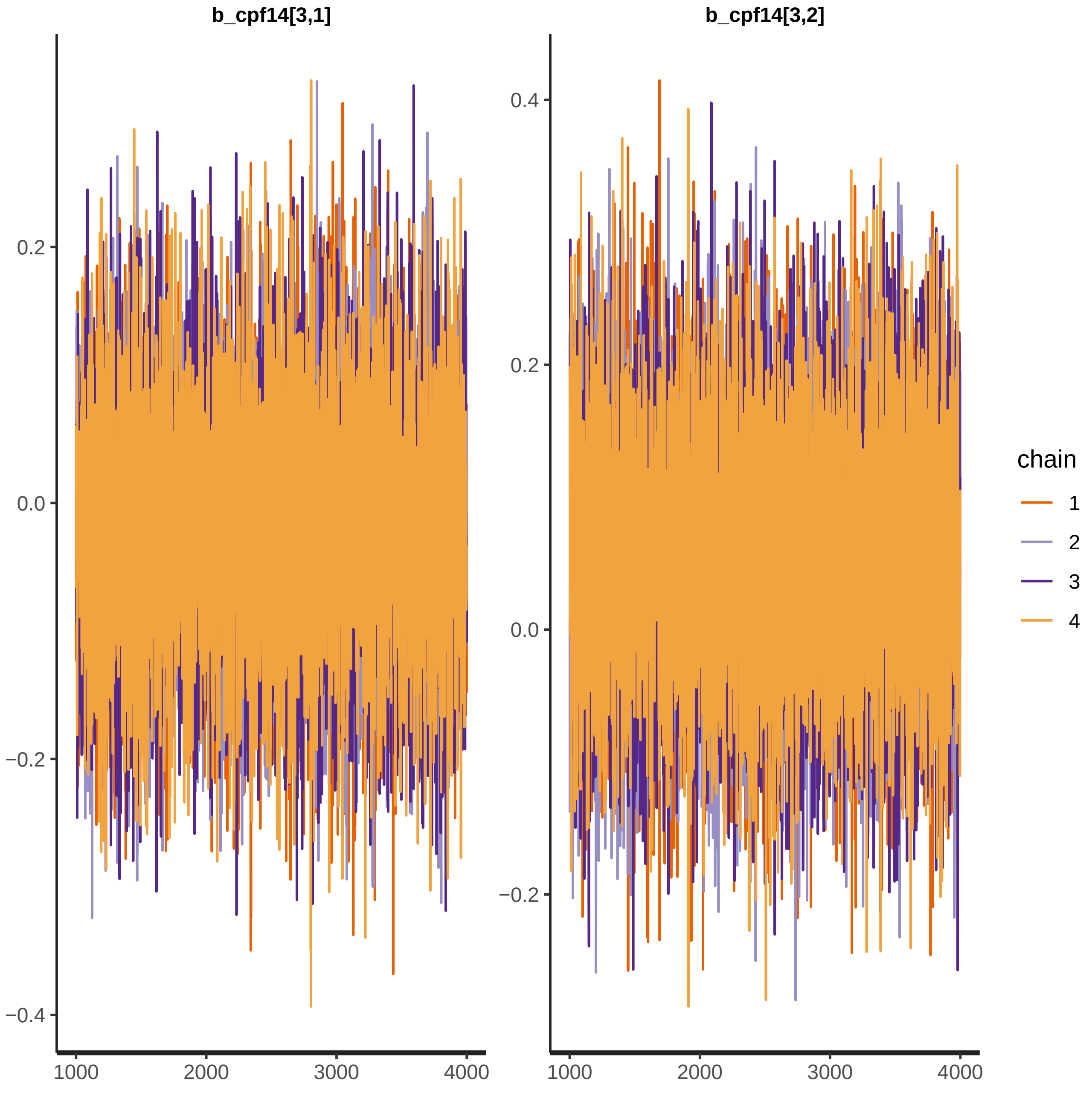
b_cpf14[3,1]
b_cpf14[3,2]
0.2
0.0
−0.2
−0.4
0.4
0.2
0.0
−0.2
1000
2000
3000
4000
1000
2000
3000
4000
chain
1
2
3
4

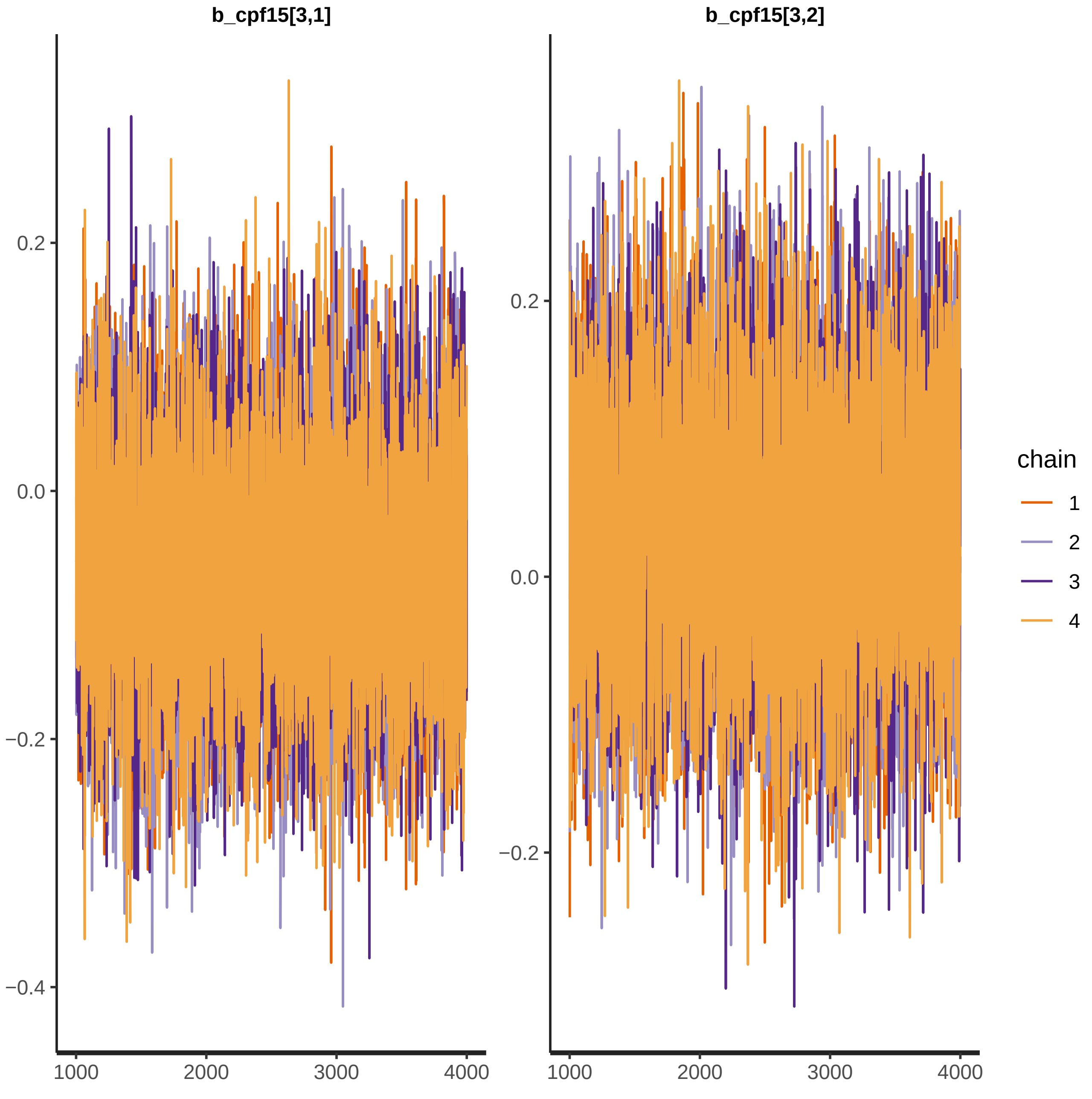
b_cpf15[3,1]
b_cpf15[3,2]
0.2
0.0
−0.2
−0.4
1000
2000
3000
4000
chain
1
2
3
4

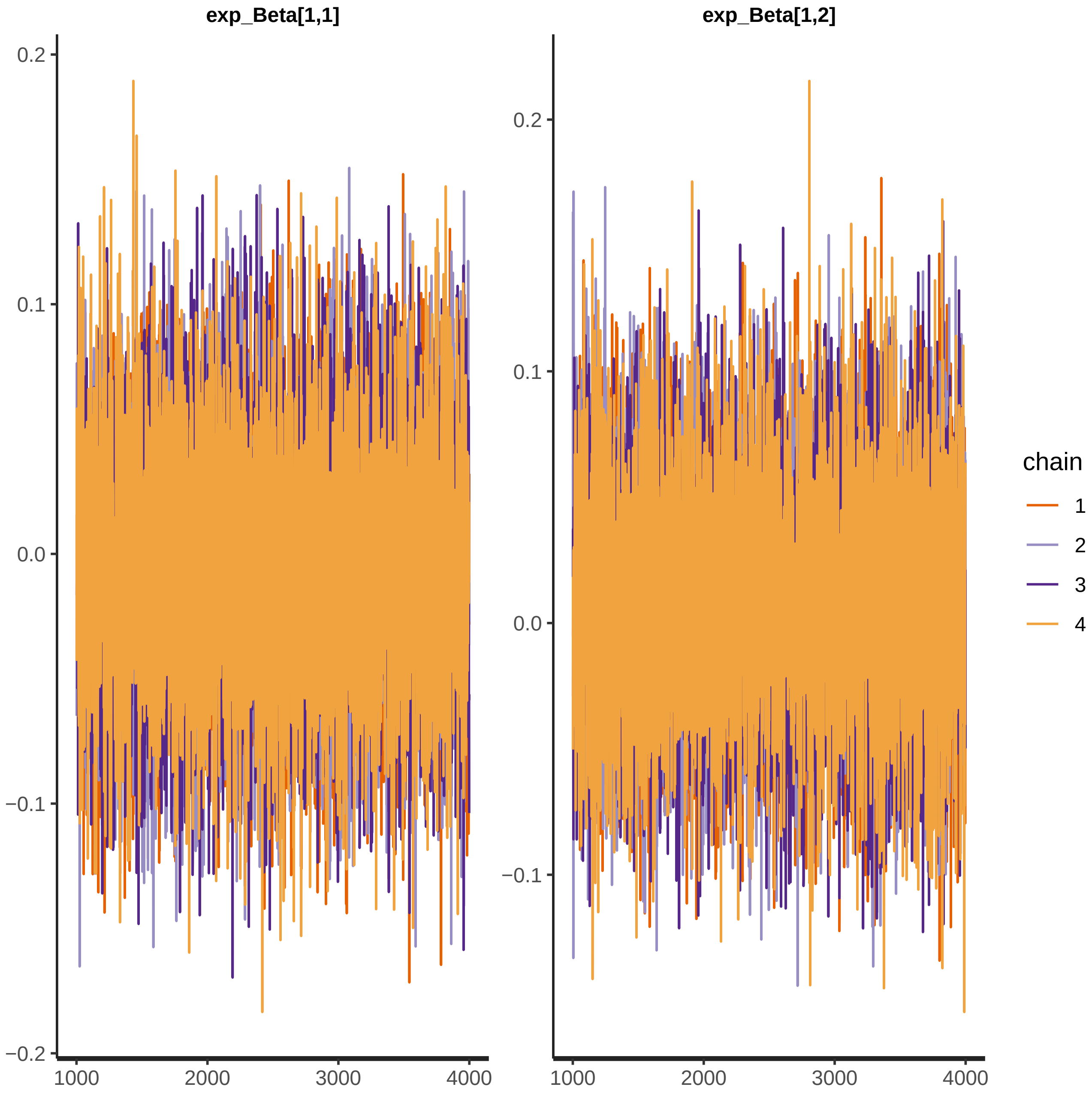

exp_Beta[1,1]
exp_Beta[1,2]
0.2
0.1
0.0
−0.1
−0.2
1000
2000
3000
4000
chain
1
2
3
4

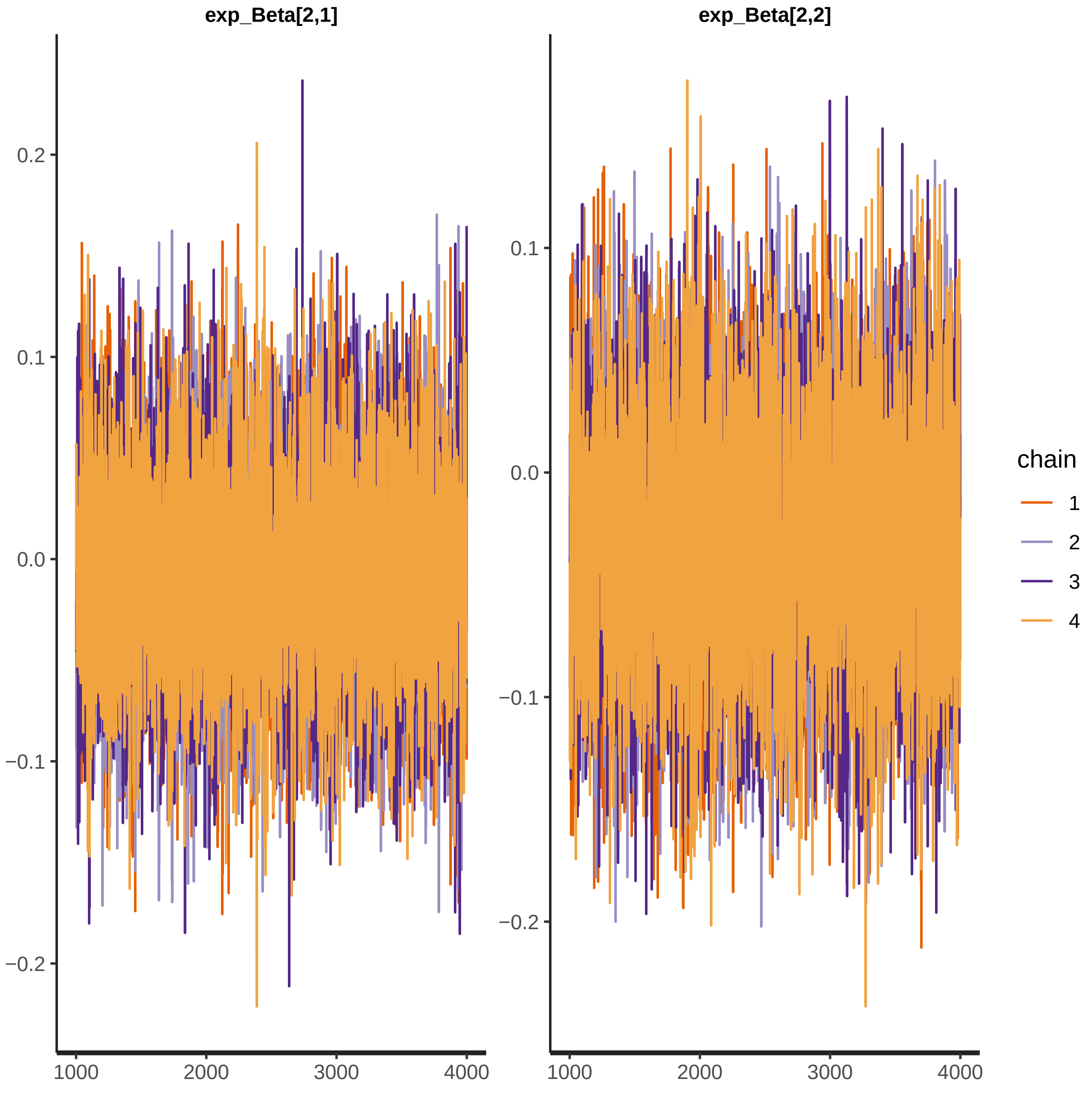

exp_Beta[2,1]
exp_Beta[2,2]
0.2
0.1
0.0
−0.1
−0.2
1000
2000
3000
4000
chain
1
2
3
4

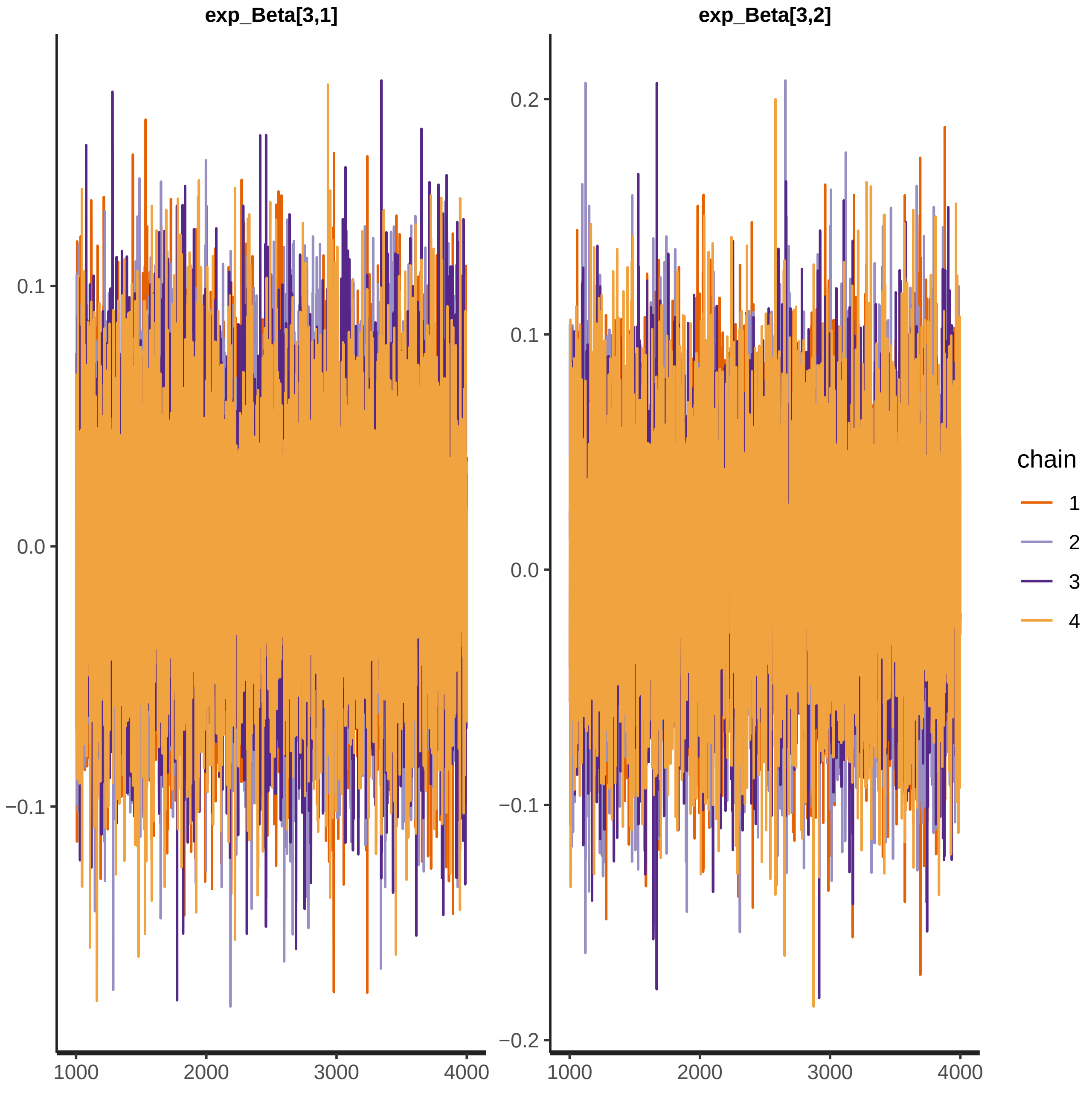
exp_Beta[3,1]
exp_Beta[3,2]
0.1
0.0
−0.1
0.2
−0.2
1000
2000
3000
4000
chain
1
2
3
4

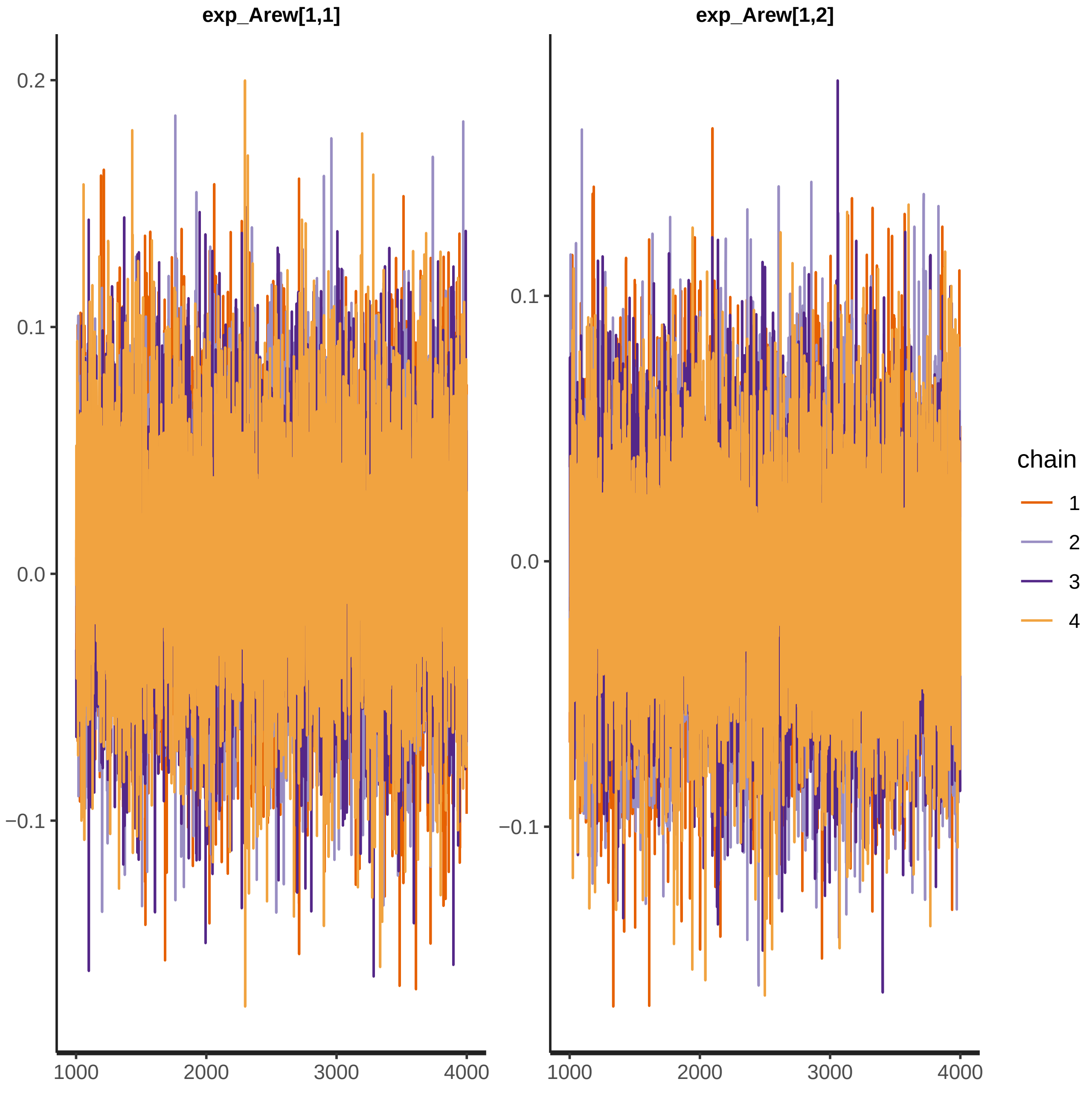

exp_Arew[1,1]
exp_Arew[1,2]
0.2
0.1
0.0
−0.1
1000
2000
3000
4000
chain
1
2
3
4

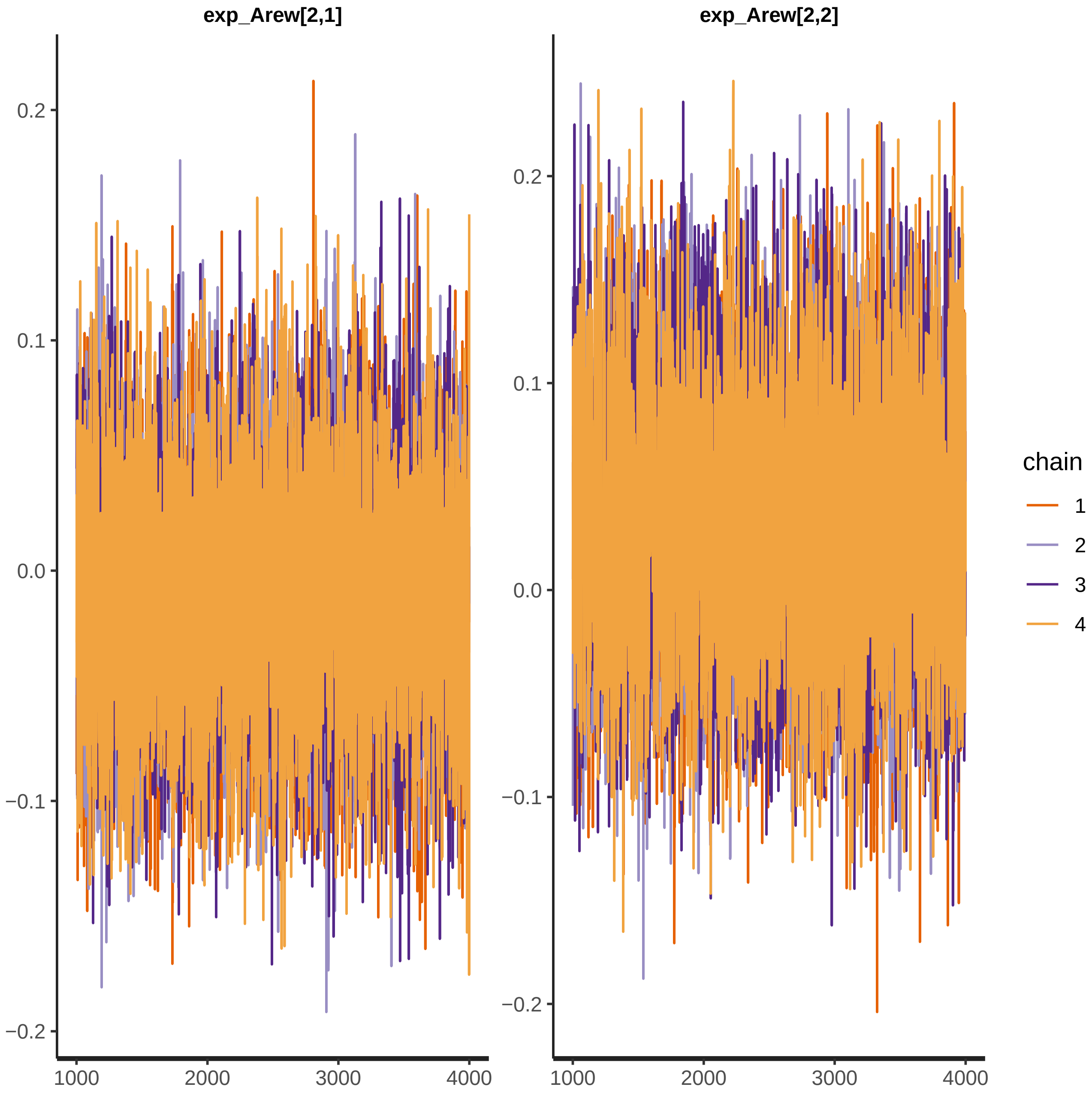

exp_Arew[2,1]
exp_Arew[2,2]
0.2
0.1
0.0
−0.1
−0.2
1000
2000
3000
4000
chain
1
2
3
4

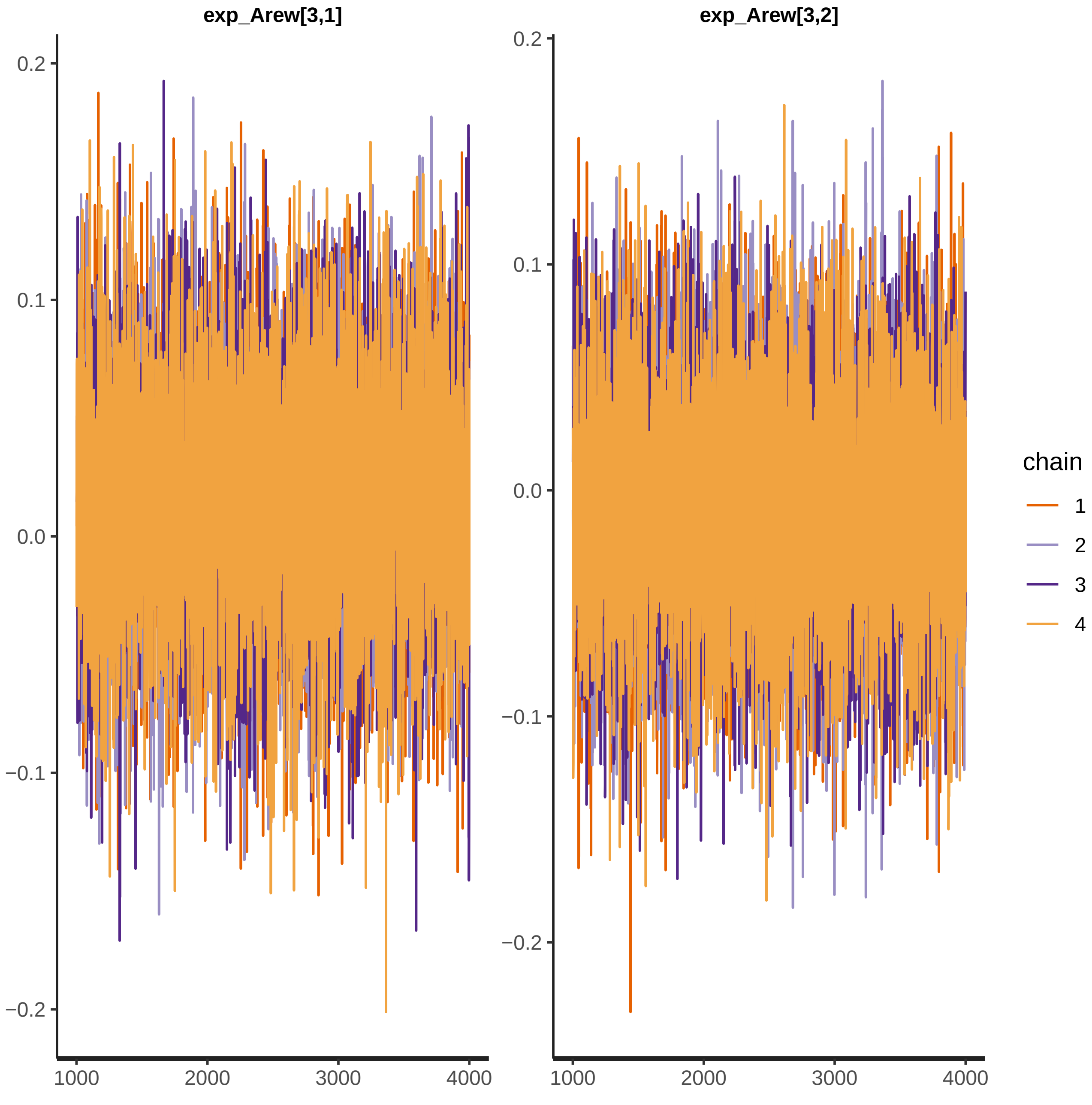
exp_Arew[3,1]
exp_Arew[3,2]
0.2
0.1
0.0
−0.1
−0.2
1000
2000
3000
4000
chain
1
2
3
4
