## Supplementary figures and images for "Contingency-based flexibility mechanisms through a reinforcement learning model in adults with Attention-Deficit/Hyperactivity Disorder and Obsessive-Compulsive Disorder"

### RL_betas_group_graphs.pdf

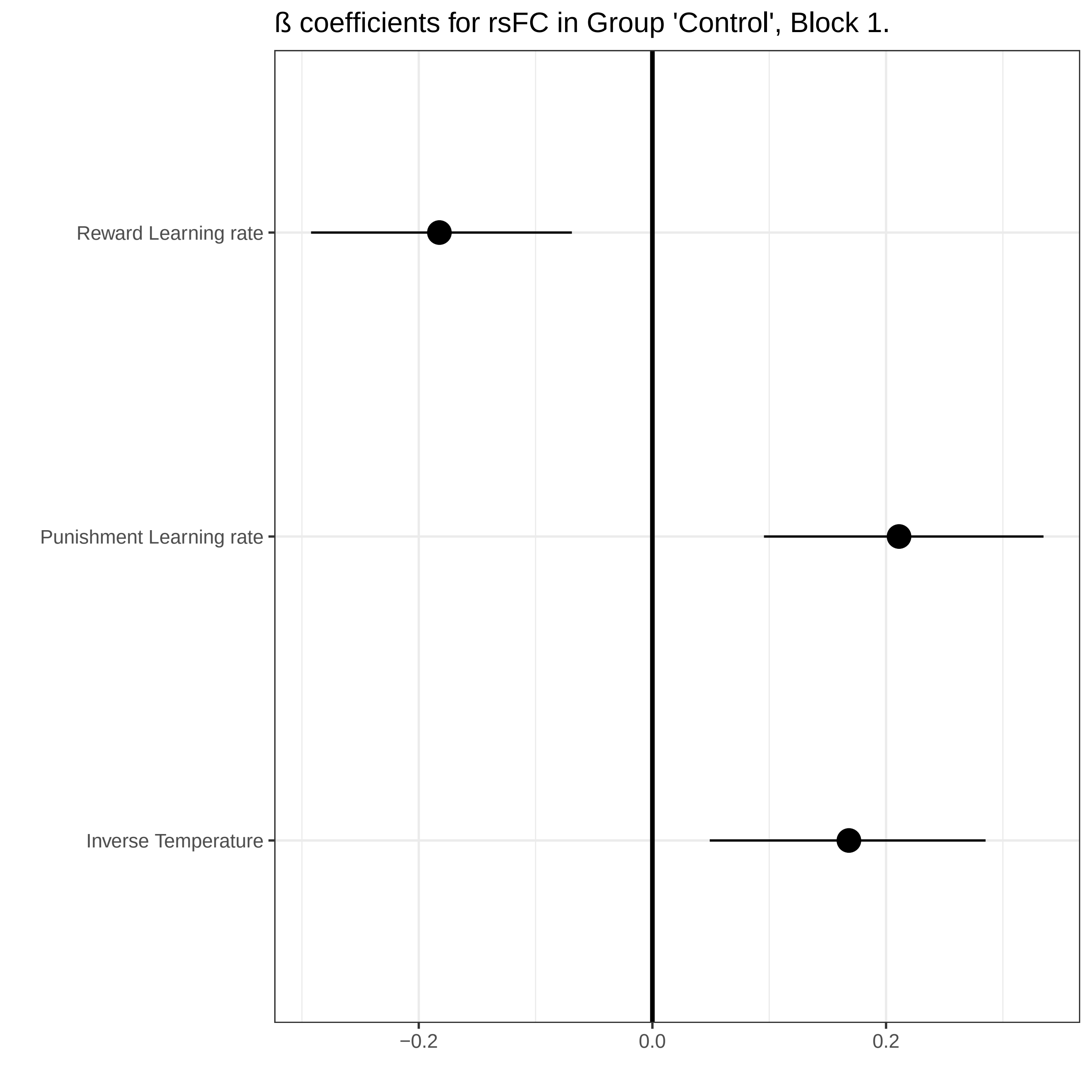

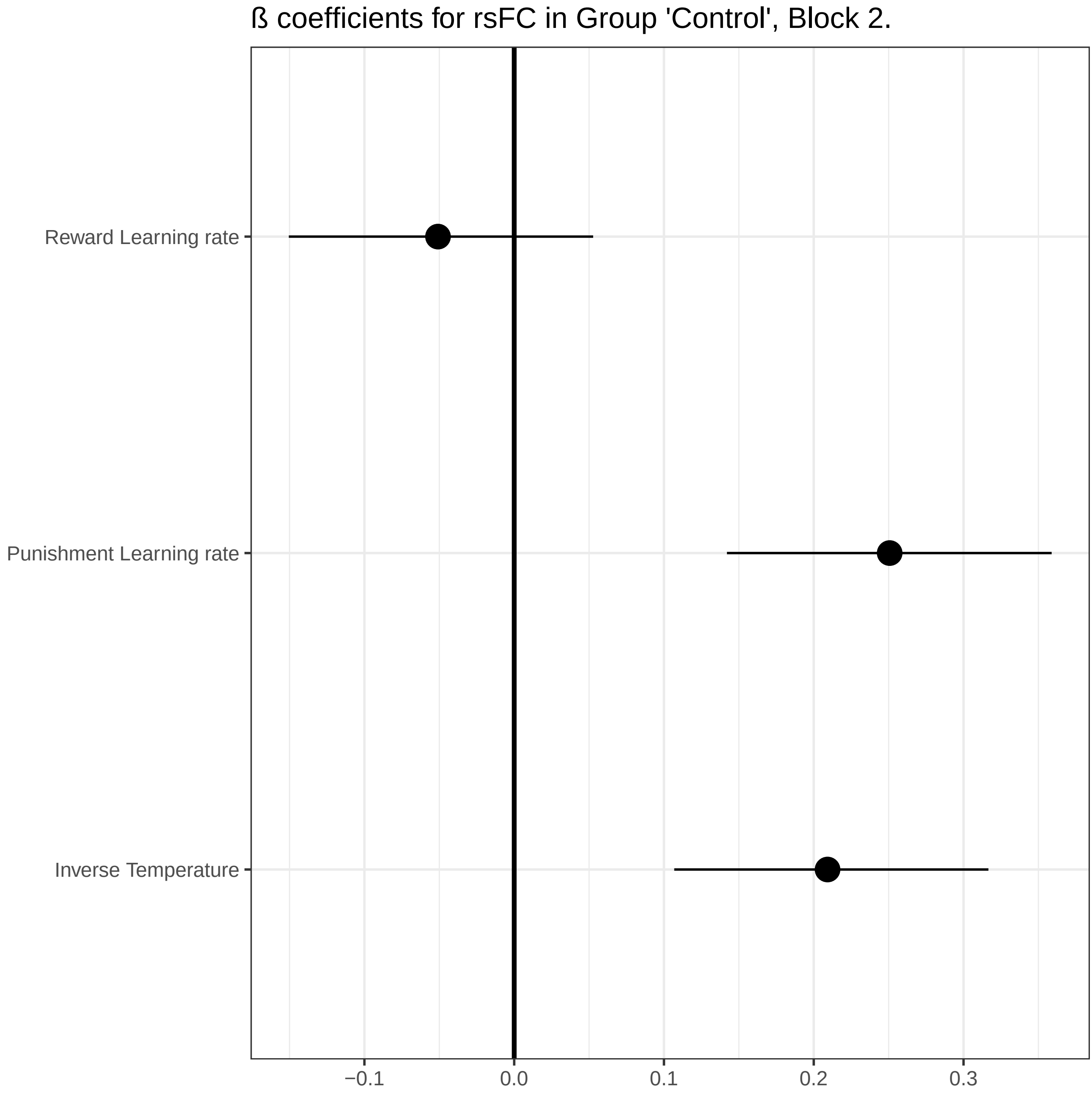

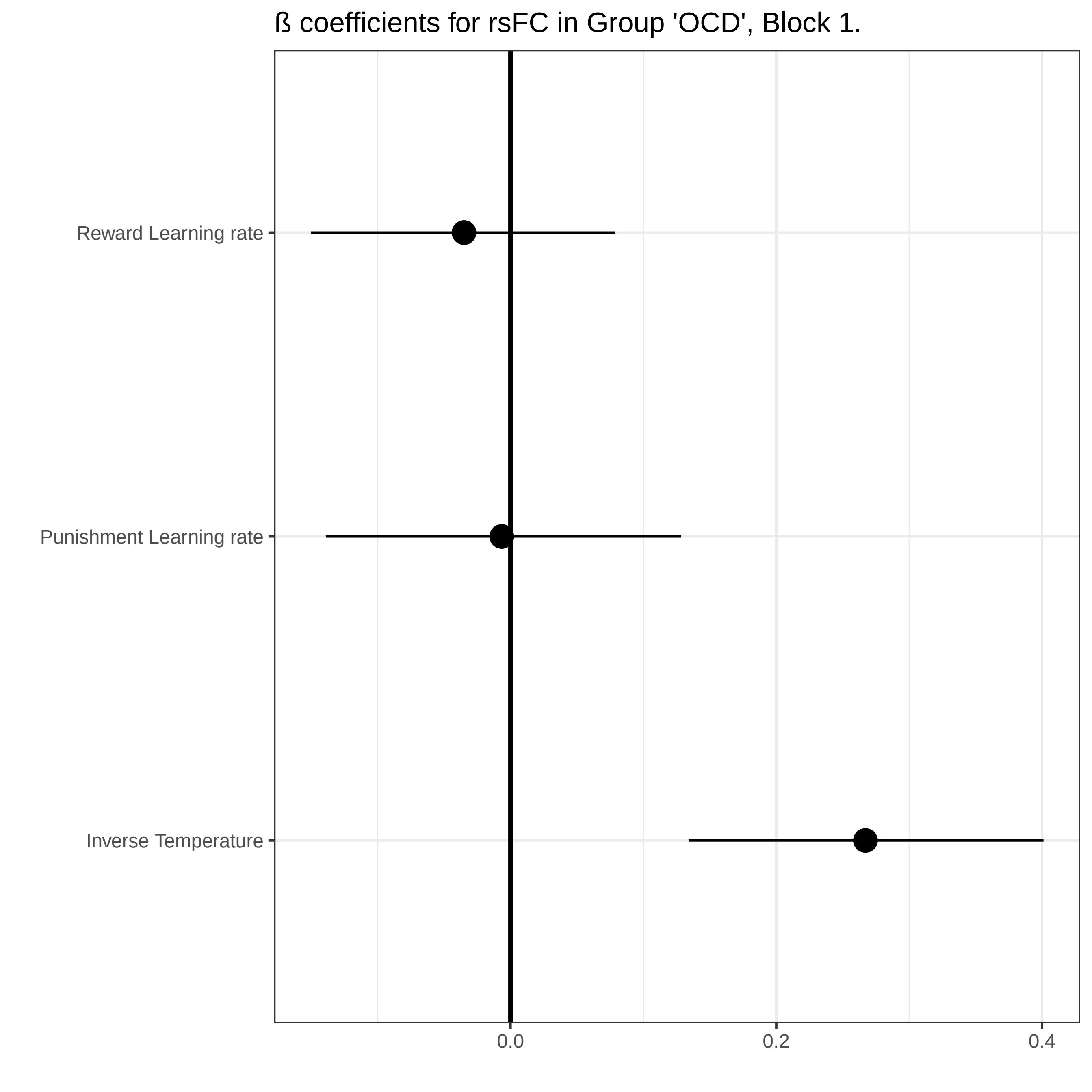

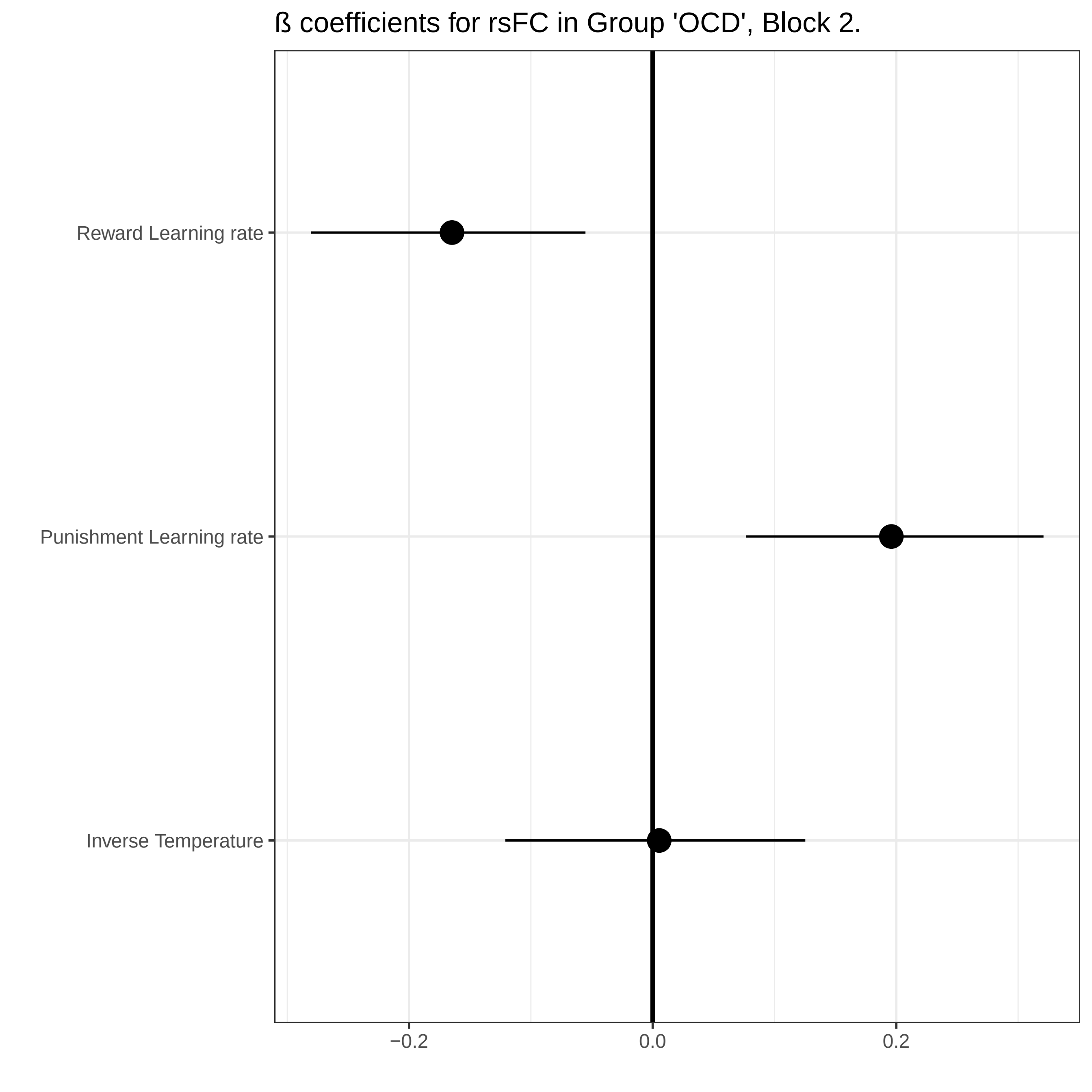

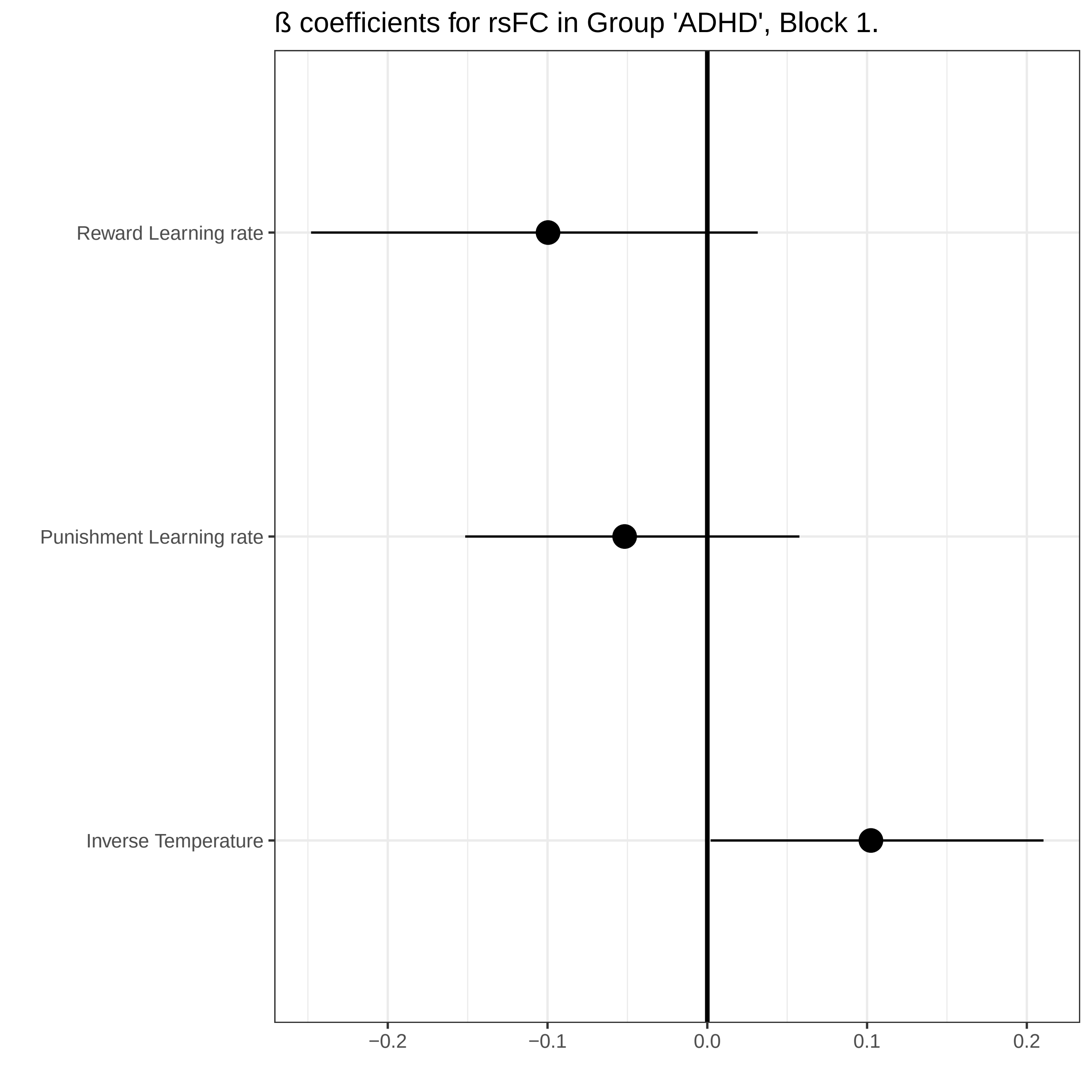

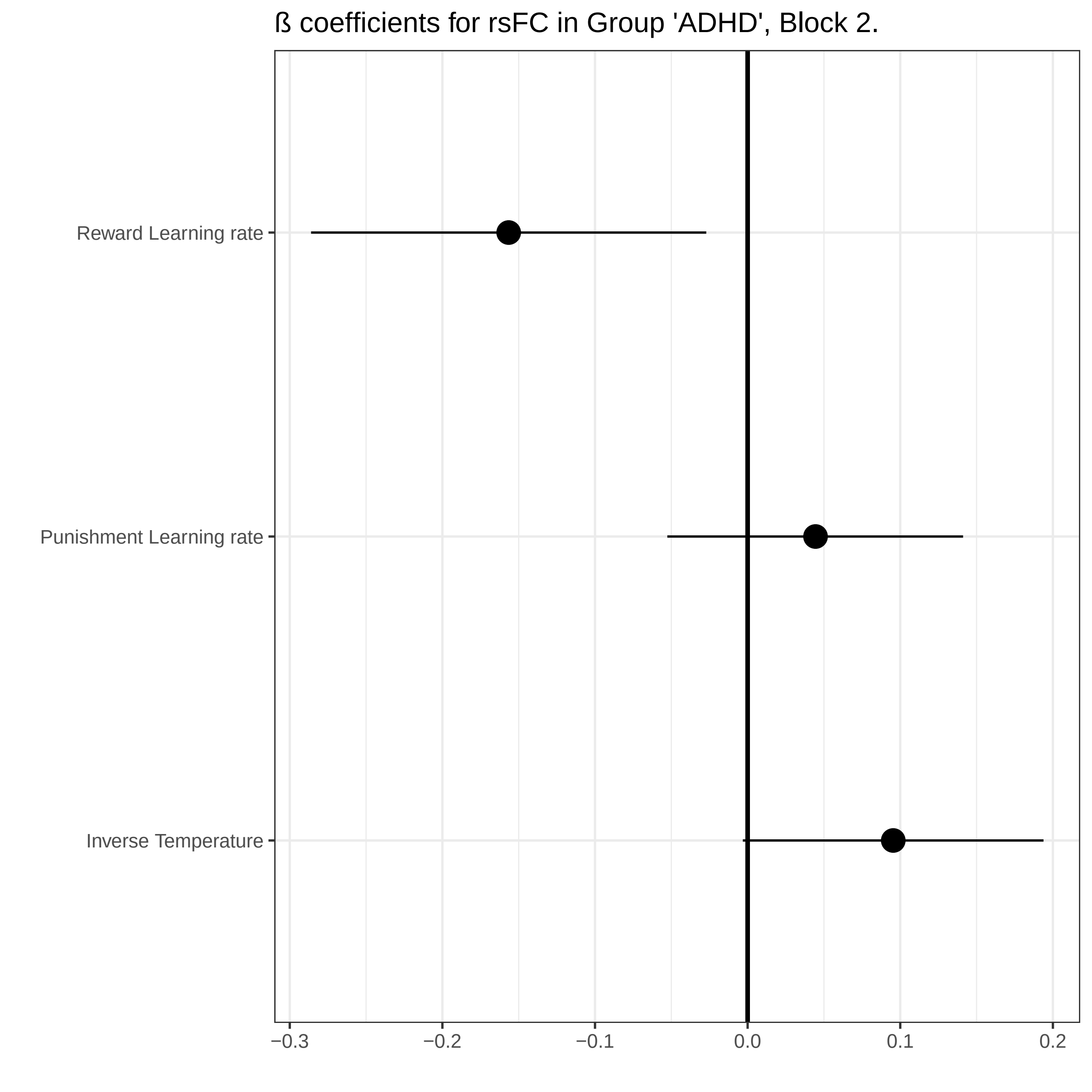

### rsFC_betas_group_graphs.pdf

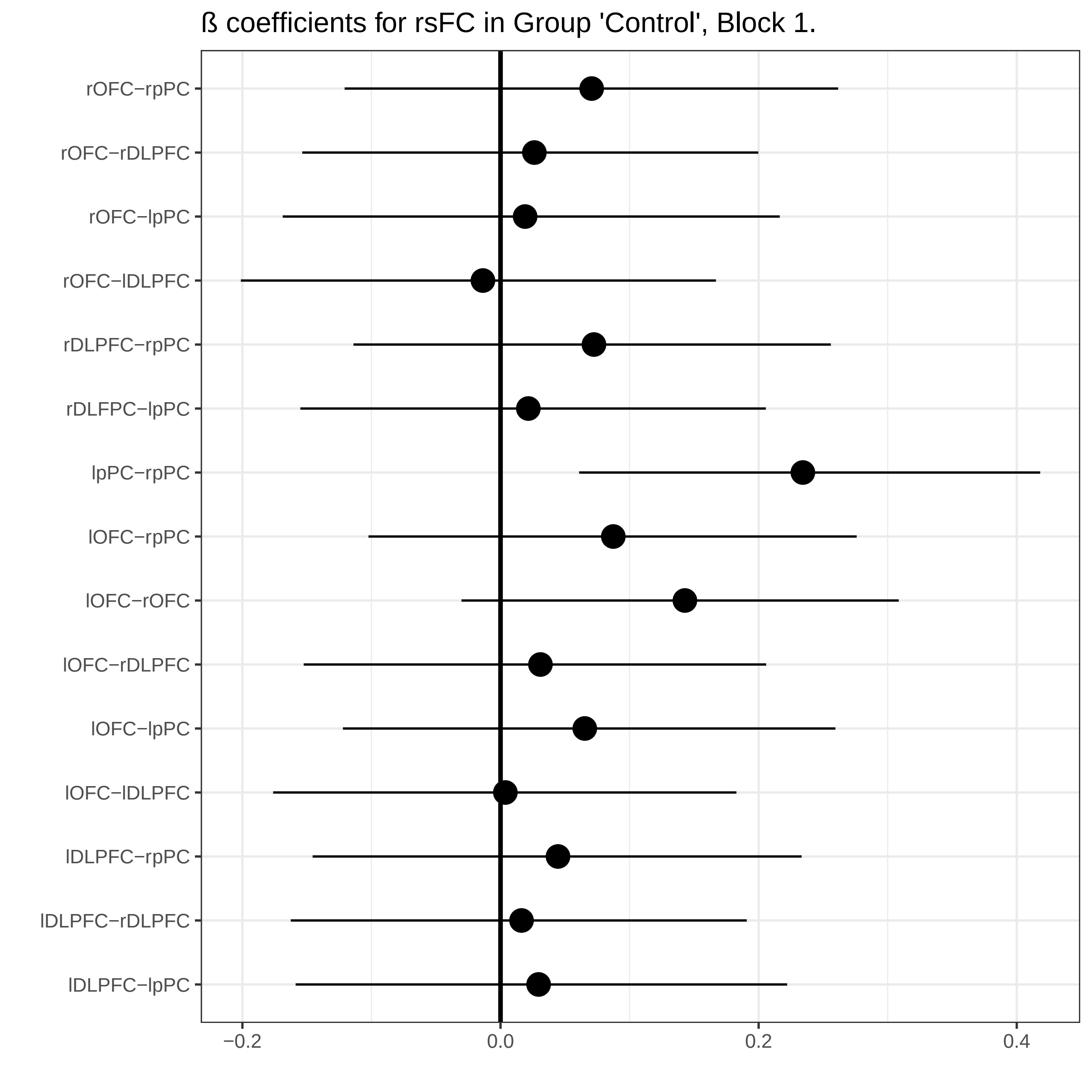

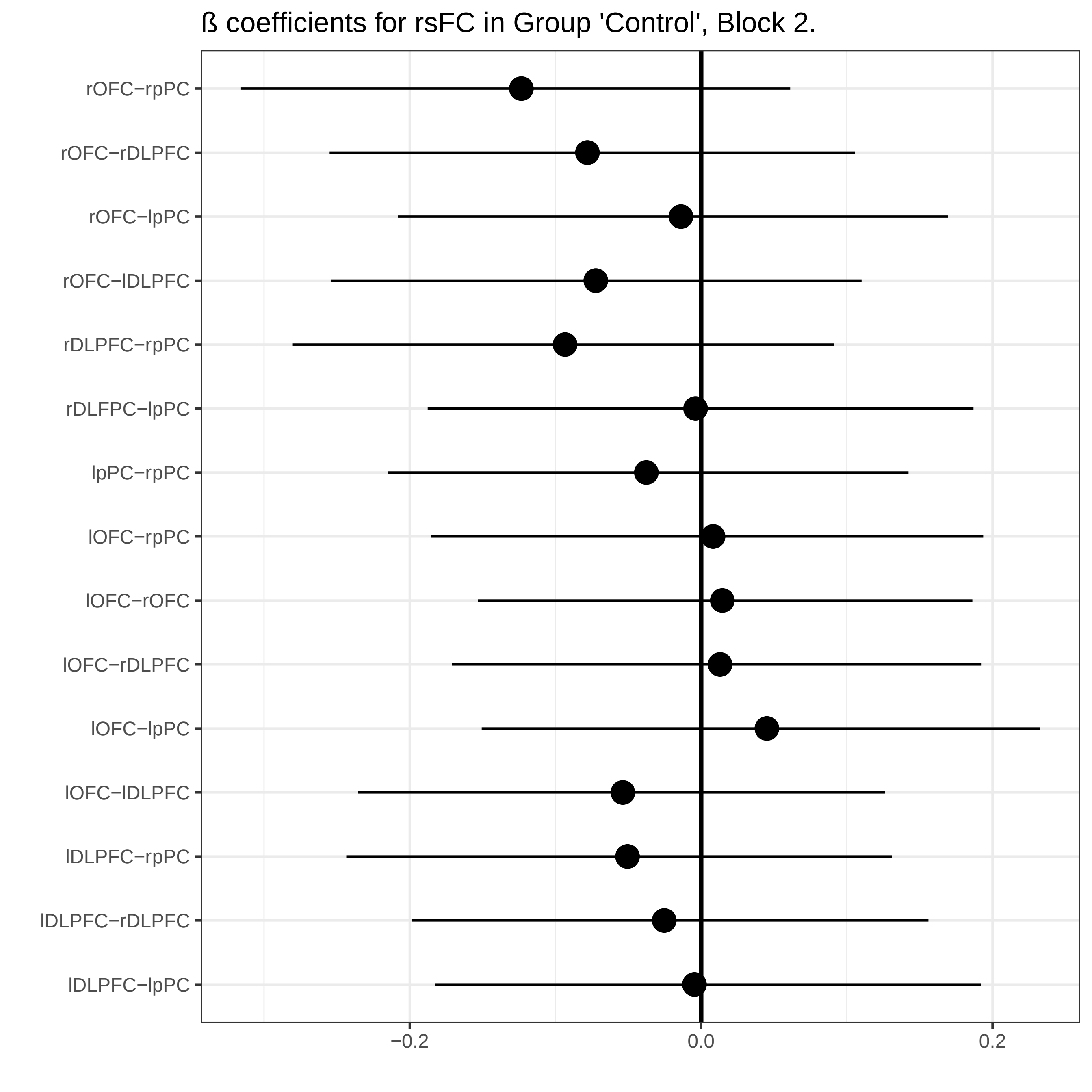

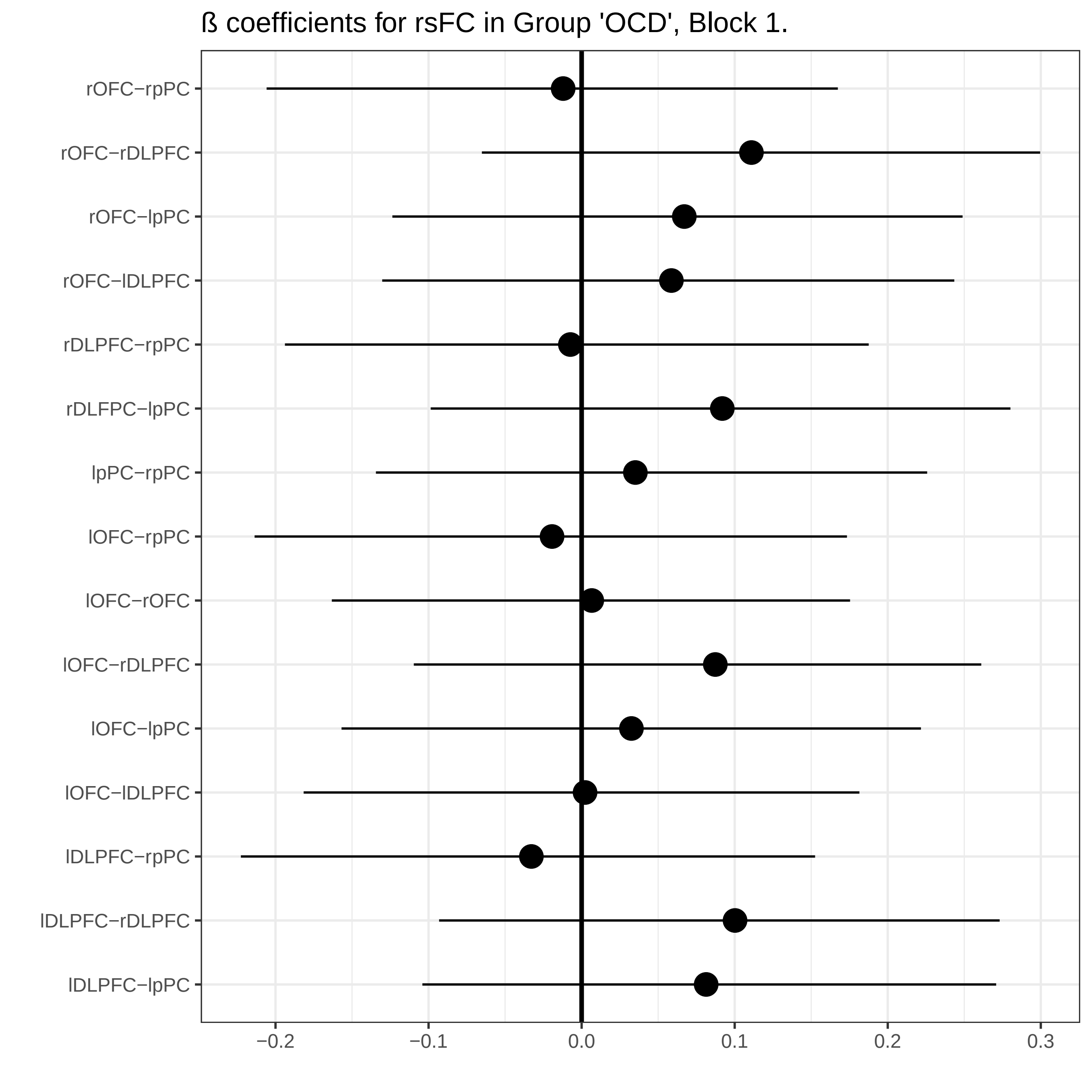

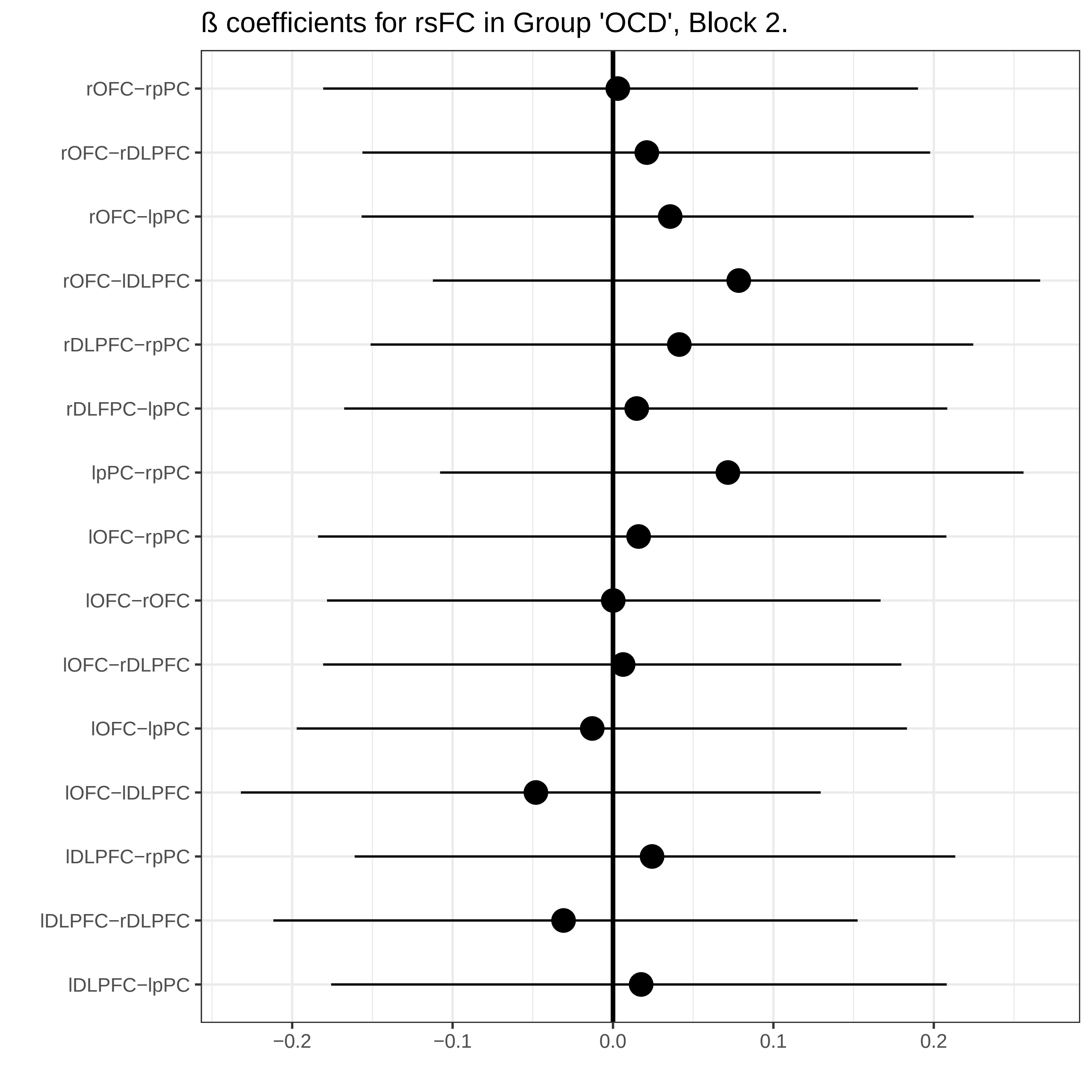

ß coefficients for rsFC in Group 'ADHD', Block 1.

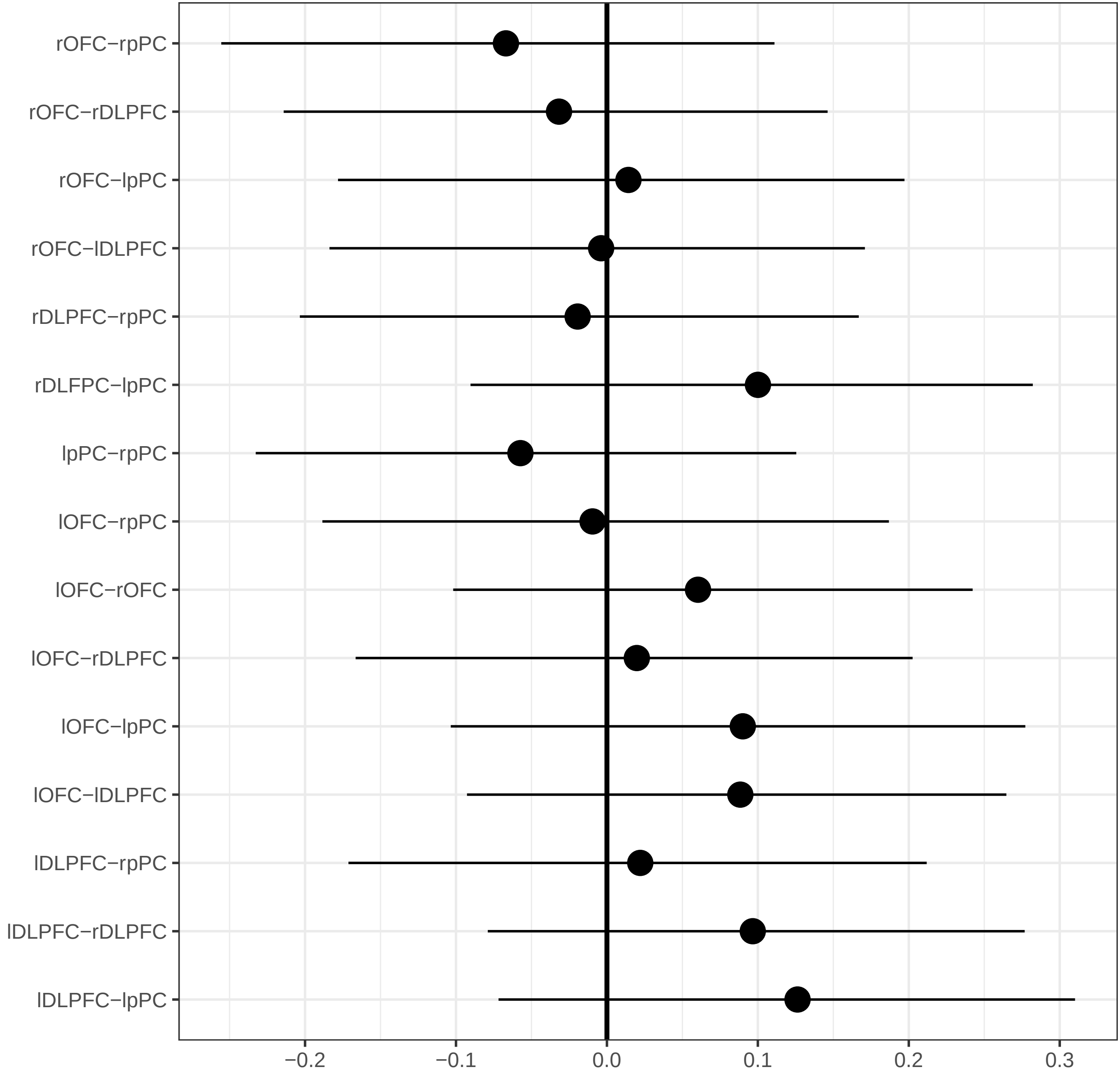

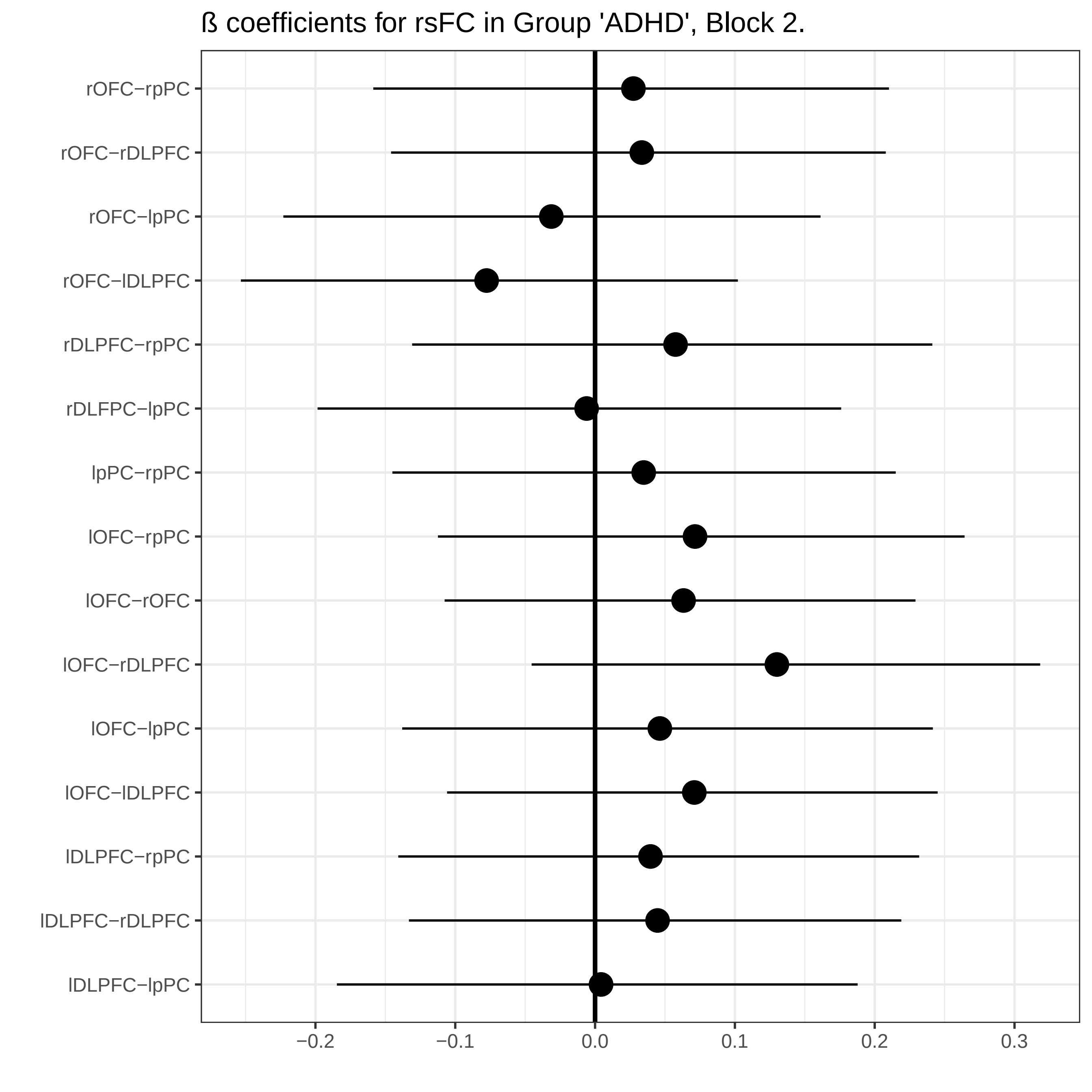
